# Predicting the Evolution of COVID-19 Mortality Risk: a Recurrent Neural Network Approach

**DOI:** 10.1101/2020.12.22.20244061

**Authors:** Marta Villegas, Aitor Gonzalez-Agirre, Asier Gutiérrez-Fandiño, Jordi Armengol-Estapé, Casimiro Pio Carrino, David Pérez Fernández, Felipe Soares, Pablo Serrano, Miguel Pedrera, Noelia García, Alfonso Valencia

## Abstract

**Background:** The propagation of COVID-19 in Spain prompted the declaration of the state of alarm on March 14, 2020. On 2 December 2020, the infection had been confirmed in 1,665,775 patients and caused 45,784 deaths. This unprecedented health crisis challenged the ingenuity of all professionals involved. Decision support systems in clinical care and health services management were identified as crucial in the fight against the pandemic.

**Methods:** This study applies Deep Learning techniques for mortality prediction of COVID-19 patients. Two datasets with clinical information (medication, laboratory tests, vital signs etc.) were used. They are comprised of 2,307 and 3,870 COVID-19 infected patients admitted to two Spanish hospital chains. Firstly, we built a sequence of temporal events gathering all the clinical information for each patient. Next, we used the temporal sequences to train a Recurrent Neural Network (RNN) model with an attention mechanism exploring interpretability. We conducted extensive experiments and trained the RNNs in different settings, performing hyperparameter search and cross-validation. We ensembled resulting RNNs to reduce variability and enhance sensitivity.

**Results:** We assessed the performance of our models using global metrics, by averaging the performance across all the days in the sequences. We also measured day-by-day metrics starting from the day of hospital admission and the outcome day and evaluated the daily predictions. Regarding sensitivity, when compared to more traditional models, our best two RNN ensemble models outperform a Support Vector Classifier in 6 and 16 percentage points, and Random Forest in 23 and 18 points. For the day-by-day predictions from the outcome date, the models also achieved better results than baselines showing its ability towards early predictions.

**Conclusions:** We have shown the feasibility of our approach to predict the clinical outcome of patients infected with SARS-CoV-2. The result is a time series model that can support decision-making in healthcare systems and aims at interpretability. The system is robust enough to deal with real world data and it is able to overcome the problems derived from the sparsity and heterogeneity of the data. In addition, the approach was validated using two datasets showing substantial differences. This not only validates the robustness of the proposal but also meets the requirements of a real scenario where the interoperability between hospitals’ datasets is difficult to achieve.

## 1 Introduction

According to the daily report of the Coordination Center for Health Alerts and Emergencies^1^ of the Spanish Ministry of Health, as of 2 December 2020, a total of 1,665,775 confirmed cases of COVID-19 and 45,784 deaths had been reported in Spain. To effectively respond to a challenge of this magnitude and to optimize the hospitalization process of this emerging disease, decision support systems for clinical care and health services management are crucial. Artificial Intelligence and Deep Learning tools offer a range of possibilities for obtaining models that, trained with historical data, can anticipate future scenarios.

In this work, we apply Deep Learning techniques for predicting the clinical outcome of patients with COVID-19. The result is a model that predicts mortality risk to support decision making. This study leverages two datasets with clinical information on 2,307 and 3,870 patients from HM Hospitales (HM) and the Hospital Universitario 12 de Octubre (H120) in Spain.

We propose a time series analysis system using a Recurrent Neural Networks (RNN) based ensemble model that responds to the reality of hospitals during the pandemic. We developed a model that generates daily predictions in a hospital environment to be used as an early warning system. The system predicts the mortality risk for each inpatient on a daily basis considering all previous available data.

A relevant characteristic of our proposal is the effort devoted to the interpretability of the model, which is based on an attention mechanism. This mechanism considers the RNN’s output with respect to the previous ones to relevantly weight the past information for the daily prediction, aiming at providing explanatory capabilities in the temporal dimension.

Time series prediction problems are considered complex in predictive modeling. RNNs are well suited to capture sequential information from temporal data, since they learn long-term dependencies by incorporating a memory effect that is able to preserve state over time. RNNs have been applied in different proposals addressing Intensive Care Unit (ICU) mortality prediction based on Electronic Health Records (EHR), as in [1, 2, 3, 4].

Lipton et al. [5] produced the first study using a Long Short Term Memory RNN [6] to classify diagnoses given multivariate time series. In this experiment, the diagnostic labels had no timestamps and they were used only as predictive labels. Similarly to our work, in the training phase, the authors used target replication (an output is generated at every sequence step). However, at prediction time, the system considers only the output during the final step so that, although the system considers the whole series of events, it does not give a prediction until the end.

Choi et al. [7] developed Doctor AI, a predictive model that uses diagnosis, medication or procedure codes as input for the RNN to predict the diagnosis and medication categories for a subsequent visit. Contrary to us, they had enough data to use Skip-gram [8] to capture the latent representation of medical codes from the EHR.

Choi et al., in [9], suggest a Reverse Time Attention Mechanism with a RNN to create an interpretable predictive model and to cope with long sequences of visits. They used a two-level neural attention model that detects influential past visits and relevant clinical variables within those visits. We also use the attention mechanism to focus on the time sequence trying to identify influential episodes but we do not reverse sequences as, in our case, these are much shorter.

Fenglong et al. [10] use RNNs with a simple attention mechanism to interpret the results and to forecast health information of patients from the historical sequences of visits over time. They employ bidirectional RNNs to remember all the information of both past and future visits, and they introduce three attention mechanisms to measure the relationships of different visits for the prediction. We also experiment with attention mechanism but we use unidirectional RNN as, in our setting, we want daily predictions and thus we unroll the RNN.

Xia et al. [11] used an ensemble algorithm of LSTMs to deal with heterogeneous ICU data for mortality prediction. The eventual ensemble models make their predictions by merging the results of multiple parallel LSTM classifiers. We also explore the use of ensemble methods to improve sensitivity. However, our approach is different as we did not use bootstrapping neither random feature subspace on training data but we rather designed a voting system and defined an algorithm that, starting with the best model in terms of cross-validated F1 score and sensitivity, heuristically chooses the most different models (i.e., the ones that make the most different predictions, to encourage model diversity in the ensemble).

More recently, a COVID-19 mortality risk study that used HM data among others was published by Bertsimas et al. [12]. This is the first multi-center COVID-19 mortality risk study that uses data from 3,062 patients across four different countries. They used the XGBoost algorithm to predict mortality using 22 features. Unlike us the experiment does not consider the sequence of events but rather uses the data from the first event in its analysis.

Our work constitutes one of the first effective predictive models of time series data with COVID-19 patient records. Unlike [13] and [12], we apply full time series inference and make daily predictions, instead of using static data or collapsing the dynamic data into fixed-size vectors. To achieve this goal, we trained different RNN models, and assembled them prioritising a given metric. By using this technique, we can substantially increase the system’s sensitivity, while producing considerably more stable predictions.

## 2 Results

For **HM**, Table 1 shows the global evaluation results on cross-validation, comparing the best RNN models for each “data representation method” as described in Section 4.2 and the best ensemble models. All models have two versions depending on whether accuracy or F1 are used for early stopping and model selection. We also experimented with feature selection using three different thresholds (100, 150 and 300) and, for each representation method, we give the best results. The best ‘non-ensemble’ model gets 90.72% in accuracy, 67.75% in sensitivity and 83.74% in F1. This winner model only uses imputation method, does not need feature selection and it is better in all metrics but specificity. In general, feature selection does not have a significant impact in the results. Ensemble models show a remarkable gain in sensitivity, reaching an impressive 84.50% with virtually no negative impact on F1. Ensemble models that use sensitivity as discard criteria, show much better results in sensitivity (84.50 and 84.25 vs. 74.94 and 76.56) and almost unnoticeable fall in F1. The fact that the ensemble mechanism applies on the top 100 models guarantees good results.

**Table 1:** HM, best RNN models for each “data representation method” as described in Section 4.2.

| *Imp* | *Mis* | *Ref* | *FS* | *Met* | Accuracy | AUC | Sensitivity | Specificity | F1 |
| --- | --- | --- | --- | --- | --- | --- | --- | --- | --- |
| × | × | × | × | Acc | 0.8933±0.0062 | 0.9166±0.0118 | 0.5319±0.0890 | **0.9760±0.0153** | 0.7917±0.0261 |
| ✓ | × | × | × | Acc | 0.8982±0.0056 | 0.9385±0.0063 | 0.6194±0.0806 | 0.9620±0.0129 | 0.8152±0.0203 |
| ✓ | ✓ | × | × | Acc | 0.8917±0.0092 | 0.9271±0.0082 | 0.6094±0.0844 | 0.9563±0.0183 | 0.8049±0.0206 |
| ✓ | ✓ | ✓ | × | Acc | 0.8892±0.0107 | 0.9317±0.0062 | 0.5569±0.0931 | 0.9653±0.0100 | 0.7912±0.0330 |
| ✓ | × | ✓ | × | Acc | 0.8916±0.0050 | 0.9216±0.0100 | 0.5569±0.0479 | 0.9681±0.0080 | 0.7956±0.0152 |
| × | ✓ | × | × | Acc | 0.8911±0.0102 | 0.9169±0.0065 | 0.5431±0.0888 | 0.9707±0.0183 | 0.7912±0.0284 |
| × | ✓ | ✓ | × | Acc | 0.8937±0.0087 | 0.9222±0.0112 | 0.5506±0.0467 | 0.9721±0.0105 | 0.7974±0.0193 |
| ✓ | × | × | × | Acc | 0.8962±0.0062 | 0.9225±0.0114 | 0.5731±0.0758 | 0.9701±0.0187 | 0.8046±0.0172 |
| × | × | × | × | F1 | 0.8982±0.0036 | 0.9247±0.0088 | 0.5931±0.0594 | 0.9680±0.0103 | 0.8112±0.0147 |
| ✓ | × | × | × | F1 | **0.9072±0.0034** | **0.9408±0.0062** | 0.6775±0.0228 | 0.9597±0.0041 | 0.8374±0.0073 |
| ✓ | ✓ | × | × | F1 | 0.8949±0.0059 | 0.9269±0.0147 | 0.6738±0.0783 | 0.9455±0.0131 | 0.8195±0.0196 |
| ✓ | ✓ | ✓ | × | F1 | 0.8824±0.0091 | 0.9259±0.0090 | 0.5713±0.0876 | 0.9535±0.0134 | 0.7853±0.0276 |
| ✓ | × | ✓ | × | F1 | 0.8938±0.0056 | 0.9324±0.0071 | 0.5994±0.0737 | 0.9611±0.0114 | 0.8060±0.0194 |
| × | ✓ | × | × | F1 | 0.8888±0.0048 | 0.9184±0.0138 | 0.5794±0.0758 | 0.9595±0.0143 | 0.7955±0.0199 |
| × | ✓ | ✓ | × | F1 | 0.8967±0.0031 | 0.9287±0.0107 | 0.6150±0.0513 | 0.9611±0.0123 | 0.8132±0.0099 |
| ✓ | × | × | × | F1 | 0.8925±0.0049 | 0.9196±0.0149 | 0.6031±0.0368 | 0.9587±0.0049 | 0.8057±0.0124 |
| × | × | × | 100 | Acc | 0.8909±0.0080 | 0.9135±0.0200 | 0.5556±0.0712 | 0.9675±0.0089 | 0.7938±0.0255 |
| ✓ | × | × | 300 | Acc | 0.8990±0.0044 | 0.9395±0.0089 | 0.6563±0.0423 | 0.9545±0.0075 | 0.8230±0.0114 |
| ✓ | ✓ | × | 100 | Acc | 0.8942±0.0149 | 0.9413±0.0123 | 0.6288±0.0726 | 0.9550±0.0078 | 0.8118±0.0323 |
| ✓ | ✓ | ✓ | 150 | Acc | 0.8839±0.0056 | 0.9236±0.0090 | 0.5238±0.1068 | 0.9663±0.0202 | 0.7765±0.0293 |
| ✓ | × | ✓ | 150 | Acc | 0.8911±0.0046 | 0.9282±0.0055 | 0.6200±0.0773 | 0.9531±0.0185 | 0.8060±0.0150 |
| × | ✓ | × | 100 | Acc | 0.8933±0.0090 | 0.9063±0.0181 | 0.5481±0.0630 | 0.9723±0.0121 | 0.7960±0.0225 |
| × | ✓ | ✓ | × | Acc | 0.8937±0.0087 | 0.9222±0.0112 | 0.5506±0.0467 | 0.9721±0.0105 | 0.7974±0.0193 |
| ✓ | × | × | 100 | Acc | 0.8894±0.0046 | 0.9106±0.0111 | 0.5419±0.0204 | 0.9688±0.0071 | 0.7901±0.0076 |
| × | × | × | × | F1 | 0.8941±0.0076 | 0.9164±0.0118 | 0.5875±0.0492 | 0.9643±0.0140 | 0.8050±0.0143 |
| ✓ | × | × | × | F1 | **0.9072±0.0034** | **0.9408±0.0062** | 0.6775±0.0228 | 0.9597±0.0041 | 0.8374±0.0073 |
| ✓ | ✓ | × | × | F1 | 0.8949±0.0059 | 0.9269±0.0147 | 0.6738±0.0783 | 0.9455±0.0131 | 0.8195±0.0196 |
| ✓ | ✓ | ✓ | 150 | F1 | 0.8917±0.0076 | 0.9281±0.0016 | 0.6169±0.0679 | 0.9545±0.0144 | 0.8064±0.0191 |
| ✓ | × | ✓ | 100 | F1 | 0.8881±0.0034 | 0.9263±0.0055 | 0.5906±0.0612 | 0.9561±0.0177 | 0.7972±0.0085 |
| × | ✓ | × | 100 | F1 | 0.8867±0.0078 | 0.9093±0.0158 | 0.5756±0.0649 | 0.9578±0.0080 | 0.7924±0.0214 |
| × | ✓ | ✓ | 100 | F1 | 0.8949±0.0111 | 0.9062±0.0134 | 0.6250±0.0311 | 0.9567±0.0076 | 0.8129±0.0196 |
| ✓ | × | × | 100 | F1 | 0.8951±0.0064 | 0.9186±0.0121 | 0.6256±0.0515 | 0.9567±0.0120 | 0.8128±0.0137 |
| RNN-Ensemble-F1 |  |  |  | Acc | 0.9054±0.0045 | - | 0.7494±0.0244 | 0.9411±0.0106 | **0.8443±0.0041** |
| RNN-Ensemble-F1 |  |  |  | F1 | 0.8990±0.0075 | - | 0.7656±0.0189 | 0.9295±0.0129 | 0.8380±0.0081 |
| RNN-Ensemble-SEN |  |  |  | Acc | 0.8896±0.0054 | - | **0.8450±0.0075** | 0.8998±0.0073 | 0.8351±0.0063 |
| RNN-Ensemble-SEN |  |  |  | F1 | 0.8821±0.0036 | - | 0.8425±0.0157 | 0.8912±0.0067 | 0.8258±0.0038 |

Table 2 shows the global evaluation results for **HM** on cross-validation, comparing the best SVC, RF, and the top RNN models (all of them selected as per results in validation set).

**Table 2:** HM, final evaluation results on cross-validations.

| *Mod* | *Imp* | *Mis* | *Ref* | *FS* | *Met* | Accuracy | AUC | Sensitivity | Specificity | F1 |
| --- | --- | --- | --- | --- | --- | --- | --- | --- | --- | --- |
| RNN | ✓ | × | × | × | F1 | **0.9072±0.0034** | 0.9408±0.0062 | 0.6775±0.0228 | 0.9597±0.0041 | 0.8374±0.0073 |
| RF | × | × | × | - | Acc | 0.9032±0.0032 | 0.9361±0.0009 | 0.6169±0.0109 | **0.9687±0.0018** | 0.8228±0.0061 |
| SVC | ✓ | ✓ | × | - | F1 | 0.8889±0.0035 | **0.9458±0.0035** | 0.7806±0.0121 | 0.9137±0.0028 | 0.8269±0.0055 |
| RNN | Ensemble-F1 |  |  |  | Acc | 0.9054±0.0045 | - | 0.7494±0.0244 | 0.9411±0.0106 | **0.8443±0.0041** |
| RNN | Ensemble-SEN |  |  |  | Acc | 0.8896±0.0054 | - | **0.8450±0.0075** | 0.8998±0.0073 | 0.8351±0.0063 |

We ran one-tailed Student’s t-test and Wilcoxon signed-rank tests on sensitivity scores by folds, Student’s t-test shows that there is a significant statistical difference in the 5-fold cross-validation sensitivity scores for RNN (mean 0.8442, SD 0.0157) and RF (mean 0.6168, SD 0.0122), with *t*(4) = 27.3674, *p* = 0.008. Also for the RNN with respect to the SVC (mean 0.7806, SD 0.0135) there is a significant difference, with *t*(4) = 5.9812, *p* = 0.01. In the Wilcoxon signed-rank, test results are positive and the same as previous Wilcoxon test on the accuracy, in both RNN over RF and RNN over SVC (with *z* = *-*2.0226, *p* < 0.05).

For **H12O**, Table 3 shows the global evaluation results on cross-validation, comparing the best RNN models for different “data representation methods” and the best ensemble models. Since *imputation & missing* models showed better performance, we also experimented with feature selection using three different thresholds (100, 150 and 300). As in the case of HM, all models have two versions depending on whether accuracy or F1 are used for early stopping and model selection. Models using feature selection show slightly better results than those not using it and there is no significant difference between accuracy and F1 models. Again, ensemble models are remarkably better in sensitivity, reaching 80.82% and still keeping satisfactory F1 scores. Ensemble models using sensitivity as discard criteria are better than those using F1.

**Table 3:** H12O, best RNN models for each “data representation method”.

| Imp | Mis | Ref | FS | Met | Accuracy | AUC | Sensitivity | Specificity | F1 |
| --- | --- | --- | --- | --- | --- | --- | --- | --- | --- |
| × | × | × | × | Acc | 0.9023±0.0061 | 0.9174±0.0026 | 0.6073±0.0799 | 0.9468±0.0185 | 0.7811±0.0088 |
| ✓ | × | × | × | Acc | 0.9186±0.0062 | 0.9338±0.0055 | 0.6462±0.0357 | 0.9597±0.0103 | 0.8145±0.0100 |
| ✓ | ✓ | × | × | Acc | 0.9195±0.0028 | 0.9411±0.0040 | 0.6458±0.0357 | 0.9608±0.0075 | 0.8157±0.0056 |
| × | ✓ | × | × | Acc | 0.9140±0.0061 | 0.9316±0.0055 | 0.6683±0.0880 | 0.9511±0.0180 | 0.8099±0.0128 |
| × | × | ✓ | × | Acc | 0.9029±0.0051 | 0.9161±0.0055 | 0.6056±0.0512 | 0.9477±0.0115 | 0.7821±0.0099 |
| × | × | × | × | F1 | 0.9029±0.0041 | 0.9138±0.0114 | 0.6013±0.0767 | 0.9485±0.0151 | 0.7809±0.0117 |
| ✓ | × | × | × | F1 | 0.9164±0.0061 | 0.9349±0.0058 | 0.6765±0.0704 | 0.9526±0.0169 | 0.8154±0.0069 |
| ✓ | ✓ | × | × | F1 | 0.9228±0.0057 | 0.9396±0.0064 | 0.6765±0.0528 | 0.9599±0.0138 | 0.8261±0.0061 |
| × | ✓ | × | × | F1 | 0.9202±0.0025 | 0.9356±0.0033 | 0.6739±0.0433 | 0.9574±0.0071 | 0.8213±0.0080 |
| × | × | ✓ | × | F1 | 0.9036±0.0042 | 0.9166±0.0046 | 0.5853±0.0423 | 0.9516±0.0082 | 0.7793±0.0105 |
| ✓ | ✓ | × | 100 | Acc | 0.9236±0.0033 | 0.9392±0.0019 | 0.6112±0.0316 | 0.9707±0.0062 | 0.8167±0.0078 |
| ✓ | ✓ | × | 150 | Acc | 0.9246±0.0031 | 0.9466±0.0025 | 0.6700±0.0497 | 0.9630±0.0101 | 0.8281±0.0063 |
| ✓ | ✓ | × | 300 | Acc | 0.9197±0.0048 | 0.9387±0.0038 | 0.6596±0.0532 | 0.9589±0.0107 | 0.8182±0.0103 |
| ✓ | ✓ | × | 100 | F1 | 0.9238±0.0017 | 0.9444±0.0021 | 0.6505±0.0244 | 0.9650±0.0041 | 0.8238±0.0049 |
| ✓ | ✓ | × | 150 | F1 | **0.9264±0.0039** | 0.9474±0.0034 | 0.6609±0.0448 | 0.9664±0.0071 | 0.8297±0.0111 |
| ✓ | ✓ | × | 300 | F1 | 0.9178±0.0018 | 0.9389±0.0020 | 0.6488±0.0511 | 0.9584±0.0084 | 0.8133±0.0081 |
| RNN-Ensemble-F1 |  |  |  | Acc | 0.9201±0.0047 | - | 0.7434±0.0294 | 0.9468±0.0092 | **0.8315±0.0052** |
| RNN-Ensemble-F1 |  |  |  | F1 | 0.9198±0.0058 | - | 0.7356±0.0332 | 0.9475±0.0111 | 0.8299±0.0060 |
| RNN-Ensemble-SEN |  |  |  | Acc | 0.9097±0.0099 | - | 0.7879±0.0228 | 0.9281±0.0144 | 0.8217±0.0117 |
| RNN-Ensemble-SEN |  |  |  | F1 | 0.9011±0.0106 | - | **0.8082±0.0327** | 0.9152±0.0163 | 0.8119±0.0115 |

Table 4 shows the global evaluation results for **H12O** on cross-validation, comparing the best SVC, RF, and the top RNN models (all of them selected as per results in validation set).

**Table 4:** H12O, final evaluation results on cross-validations.

| Mod | Imp | Mis | Ref | FS | Met | Accuracy | AUC | Sensitivity | Specificity | F1 |
| --- | --- | --- | --- | --- | --- | --- | --- | --- | --- | --- |
| RNN | ✓ | ✓ | × | 150 | Acc | 0.9246±0.0031 | 0.9466±0.0025 | 0.6700±0.0497 | 0.9630±0.0101 | 0.8281±0.0063 |
| RF | ✓ | ✓ | × | - | Acc | **0.9341±0.0010** | **0.9504±0.0012** | 0.6255±0.0102 | **0.9806±0.0015** | **0.8380±0.0030** |
| SVC | ✓ | ✓ | ✓ | - | Acc | 0.9203±0.0005 | 0.9413±0.0024 | 0.6397±0.0087 | 0.9626±0.0011 | 0.8161±0.0021 |
| RNN | Ensemble-F1 |  |  |  | Acc | 0.9201±0.0047 | - | 0.7434±0.0294 | 0.9468±0.0092 | 0.8315±0.0052 |
| RNN | Ensemble-SEN |  |  |  | F1 | 0.9011±0.0106 | - | **0.8082±0.0327** | 0.9152±0.0163 | 0.8119±0.0115 |

We run the same significance tests as in HM for H12O for sensitivity by folds, one-tailed Student’s t-test shows that there is a significant statistical difference in the 5-fold cross-validation sensitivity scores for RNN (mean 0.8082, SD 0.0326) and RF (mean 0.6255, SD 0.0114), with *t*(4) = 10.9826, *p* = 0.017 and also for the RNN with respect to SVC (mean 0.6397, SD 0.0097) there is a significant difference, with *t*(4) = 9.2303, *p* = 0.018. In the Wilcoxon signed-rank, test results are positive in both RNN over RF and RNN over SVC (with *z* = *-*2.0226, *p* < 0.05).

Complete results by fold in sensitivity in both datasets are provided in the Additional Material. There, we also include the tables with the daily results for all the models. Here, Figures 1 and 2 summarize the results.

**Figure 1:**
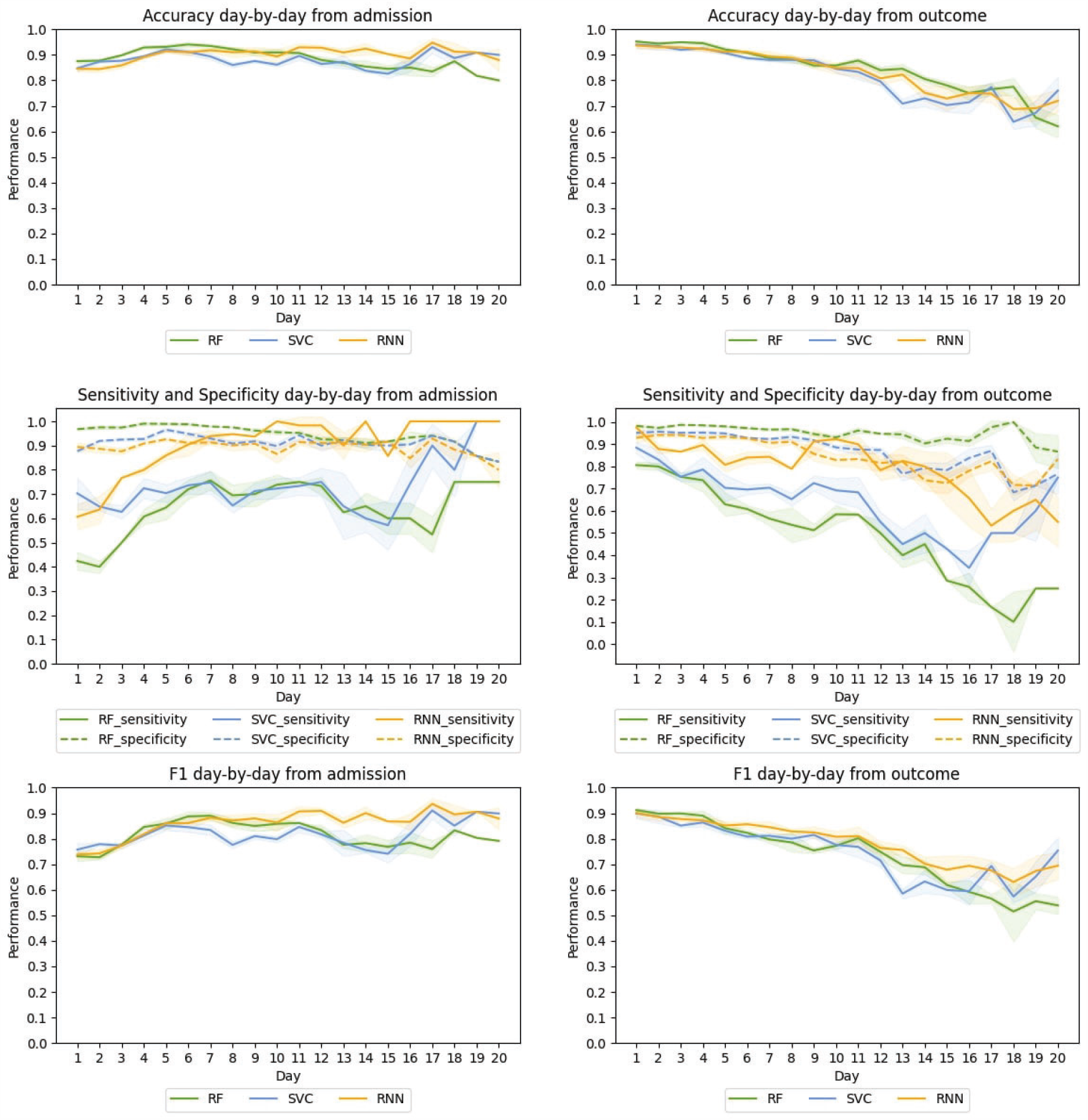
HM, Average day by day test performance from the admission (left) and from the outcome (right) in the cross-validation for the best RF, SVC, and RNN-based models.

**Figure 2:**
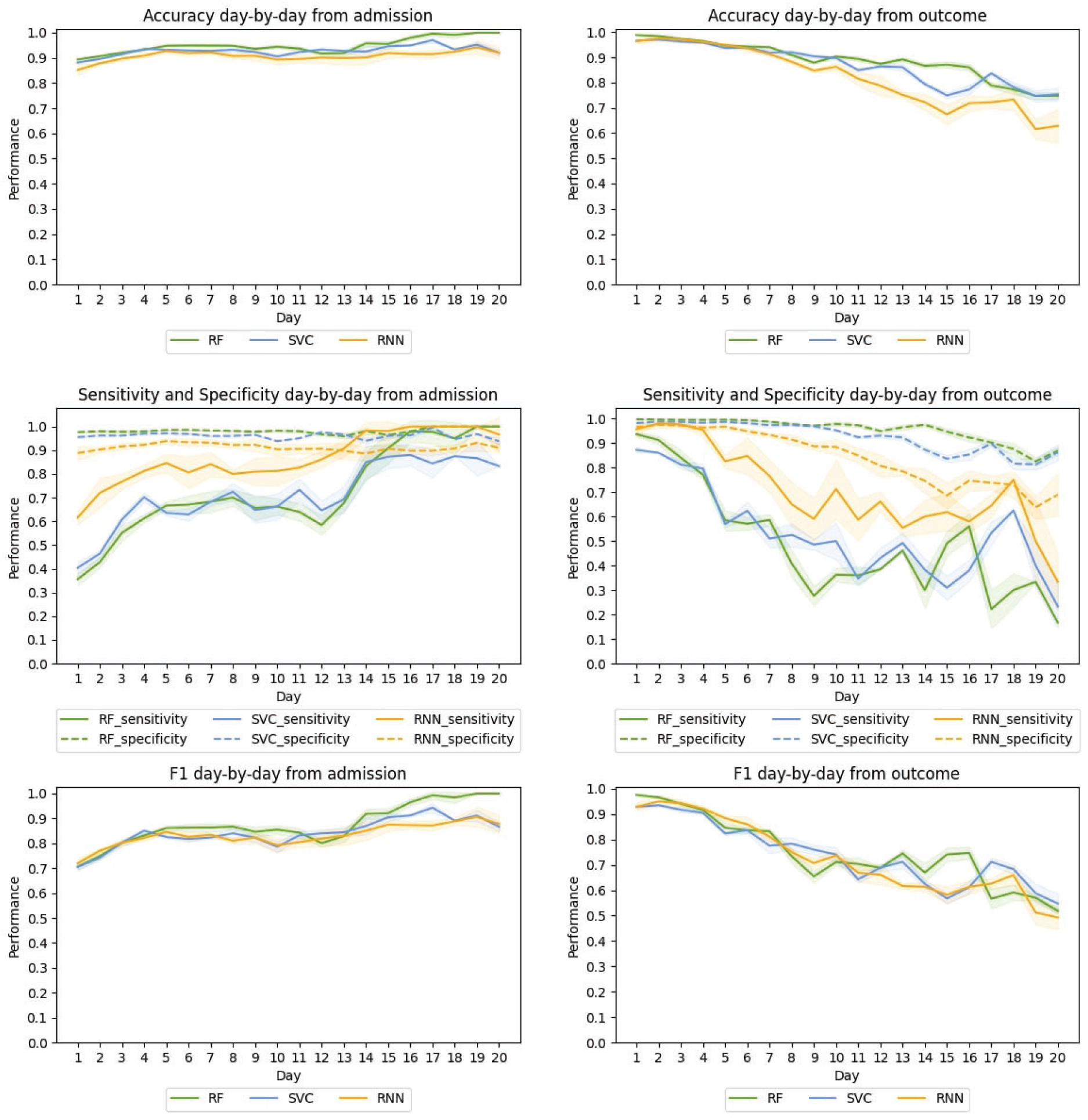
H12O, Average day by day test performance from the admission (left) and from the outcome (right) in the cross-validation for the best RF, SVC, and RNN-based models.

### 2.1 Global performance

For **HM**, the best ensemble RNN model is significantly better than the other models when considering sensitivity, reaching 0.84 in sensitivity with no penalty in F1 and a small impact in accuracy. These interesting results are achieved in all the folds, demonstrating its consistency and robustness (see sensitivity global results by folds in the Additional Material). All RNN models show slightly better scores in accuracy and F1 when compared to baselines, which is a positive result taking into account that F1 is specially useful when having an uneven class distribution. SVC results are surprisingly performant in sensitivity, clearly surpassing those of the RF, the non ensemble RNN and the Ensemble-F1 models. All HM models (60) use **attention** and, as described in the attention peak analysis in the Additional Material, the results show that 56.59% of the patients had relevant peak days.

For **H12O**, the RNN ensemble model outperforms all models in sensitivity. In particular, the Ensemble-SEN model outperforms RF in 18 points and SVC in 16 points reaching 0.80 in sensitivity. Again, for sensitivity, the results are the same in all folds. Although RF is slightly better in accuracy and F1, the difference is minimal. As for **attention**, it is worth mentioning that 53 out of the top 60 RNN models use **attention** and the best non-ensemble RNN model had 66.36% patients with peak days.

### 2.2 Day-by-day performance

When evaluating day-by-day **from the admission**, we have longer sequences as we go on. Thus, at each new step the model uses more information and gets closer to the outcome. As Figure 1 shows, in **HM**, this has small positive effect for RF and SVC: accuracy, specificity and F1 curves are high and rather flat with no noticeable improvement. For sensitivity, the RF shows a moderate improvement until day 7, a slight fall from day 12 onward and a sudden peak on day 18. SVC shows a long and strong performance curve, starting at 0.70 with a sudden rise on day 17.

The RNN-model shows a similar behaviour, with high, rather flat curves for accuracy, specificity and F1. However, the model performs better with respect to sensitivity, with a curve starting at 0.60, a peak of 1.00 on day 10, and then stabilizing between 0.85 and 1.00 until the last day.

When evaluating **from the outcome**, we have shorter sequences as we evaluate backwards. In this case, at each new step, the model has less information, and it is farther away from the outcome. Notice also that, the farther we are from the outcome, the fewer examples we have. As shown in Table 1 from Additional Material, this reduces the number of patients labeled with *death*, which may cause fluctuations, especially in sensitivity.

For **HM**, all models show similar descending curves for accuracy, specificity and F1. For sensitivity, SVC has a sharper decrease. The RNN model shows a higher and flatter curve in sensitivity, starting at 0.97, ending at 0.55 and maintaining a performance of 0.79 or better untill day 14. This shows that the system is efficient at making early predictions, since it provides reliable mortality predictions up to 14 days before the outcome.

For **H12O**, the day-bay-day performance **from the admission** is similar to that of HM. Thus, in terms of sensitivity, the RNN shows a substantial improvement with respect to the baselines. RNN mainly leads the sensitivity curve and it is very performant on both F1 and accuracy which confirms the enhancement.

When evaluating **H12O from the outcome**, the RF and the SVC show a similar behaviour: the specificity curve keeps high and flat, accuracy shows a moderate decrease, F1 suffers a sharper drop and sensitivity plummeted. Again RNN show a higher curve and a better performance in sensitivity, maintaining a score of 0.64 or better until 7 days before the outcome,

As in the case of HM, sensitivity curves from the outcome have greater fluctuations. As mentioned before, the gradual reduction of data and death labels (in absolute terms) as we move forward makes the sensitivity curves more unstable.

## 3 Discussion

This work is a first approximation of a RNN based model to predict the outcome of patients with COVID-19. The models were trained with two relatively small COVID-19 datasets and will be the basis of a future system in a larger setting. The models consume dynamic information to generate daily predictions. Unlike standard training schemes for recurrent models, we do not feed the entire sequence of dynamic data into the RNN to output a single prediction but, we feed each daily record of temporal and static data day-by-day and output a prediction for each day.

We defined an ensemble method as a voting system and developed an algorithm that, starting with the best model in terms of cross-validated accuracy or F1 score, heuristically chooses the most different models (i.e. the ones that make the most different predictions) to encourage model diversity in the ensemble but without penalizing the results. Results obtained with this ensemble method are not only superior, but they also reduce the standard deviation, producing less variation in all metrics among folds.

The system is robust enough to deal with real world data and it is able to overcome the problems derived from the sparsity and heterogeneity of the data. In addition, the approach was validated using two datasets showing substantial differences. Although both datasets contain information from COVID patients, they have different characteristics which made it impossible to join the datasets in a single one. The disparity in coding conventions and the differences in the type of information contained precluded aggregating data, as this would produce a reduced dataset due to the low overlap. Instead, we generated a model for each dataset, following the same method and using the same architecture. This not only validates the robustness of the proposal but also meets the requirements of a real scenario where the interoperability between hospitals’ datasets is difficult to achieve. This ended up in an exhaustive experimentation maximizing the potential of real world data. We explored and evaluated different data representation methods and this information is reported in detail in Section 4.2.

Most importantly, we were extremely conservative when filtering out patients or features. For instance, in [11], which is a study substantially similar to ours, they only considered patients with a time window of 10 days. On the contrary, we do not apply such filters and, consequently, the lengths of the dynamic data exhibit high variance. Similarly, we are really permissive with features. Whereas [12] omit features whose values are more than 40% missing, we do not filter features based on the percentage of missing values, and deal with sparse data representations. Our system worked with real world data and it is more realistic in medical terms, so that it can be run out of the box.

Regarding interpretability, when using the attention mechanism on the dynamic data vector, we show that there are relevant days that the model focuses on. In other words, the model keeps track of relevant days to enhance its predictions. Visualizing attention weights is useful for understanding the evolution of the patient over time. In addition, for both HM and H12O, we analyzed the attention on dynamic data in the TEST set to see whether the attention is evenly distributed or, on the contrary, it has peak days. The results show that 56.59% and 66.36% of the patients have peak days. This corroborates our hypothesis that the attention mechanism targets relevant days to perform correctly. Ensembling models can reduce interpretability if some of the models selected do not use attention. One should notice, however, that the vast majority of the top 60 models in both datasets use attention (all 60 for HM and 53 for H12O). When all the models selected use attention, it is recommended to group them by their predicted outcome and then analyze the aggregated attention to check why some of the models predict one outcome or another. We are planning new experiments to get more insights from attention data.

Regarding the global results, for **HM**, the best ensemble RNN model is significantly better than the other models when considering sensitivity, reaching an important result of 0.84 in sensitivity with no penalty in F1 and a small impact in accuracy.

Similarly, for **H12O**, the RNN ensemble model outperforms all models in sensitivity. In particular, the Ensemble-SEN model outperforms RF in 18 percentage points and SVC in 16 percentage points reaching an **0**.**80 in sensitivity**.

When evaluating day-by-day performance from the outcome, the **HM** RNN model shows a higher and flatter curve in sensitivity compared to the baselines, starting at 0.97, ending at 0.55 and maintaining a performance of 0.79 or better until day 14. This shows that the system is capable of making early predictions. Similarly, the **H12O** RNN model shows a better performance in sensitivity, maintaining a score of 0.64 or better until 7 days before the outcome. Even in our data constrained scenario, the models outperform the strong baselines of tuned RF and SVC in terms of sensitivity. We hypothesize that the model will further improve its performance when more data are available.

## 4 Methods

### 4.1 Datasets

We used two different COVID datasets^2^. One is the dataset made available by HM from April 25th 2020, in their “COVID Data Save Lives” initiative and contains information of patients diagnosed with COVID until that date. The other one comes from H12O and the cohort extends to October 14th. In both cases, the data include two different types of variables: **static variables** that are constant throughout admission, such as sex or age, and **dynamic variables** that are measured at different times during the hospital stay, such as medication, laboratory analysis and vital signs.

Although both datasets contain information from COVID patients, they have different characteristics which prevented to join the datasets in a single one. The disparity in coding (e.g. ICD-9 vs ICD-10) and the differences in the type of information contained made it difficult to aggregate data and would produce a reduced dataset due to the low overlap. Note, however, that having two different datasets allowed us to validate the robustness of the proposal. Further details about the datasets and data cleaning are given in the Additional Material.

### 4.2 Feature Representation

Information in EHR needs to be transformed into appropriate data representations to be used on clinical ML. Learning better representations is critical for performance improvement. The feature generation is a data processing used to encode variables into numerical values. The encoding operation depends on the values of the variables and our dataset includes

- Categorical variables: a variable that has multiple values with no order, each one associated to a given case (e.g. medications).
- Ordinal variables: a variable taking discrete values with a given order. The discrete values are defined by comparing the original values of the variables with a given interval of reference (e.g. vitals signs and some lab results).
- Continuous variables: a sub-type of ordinal variable that takes numerical values ordered along a continuous scale (e.g temperature).

In general, categorical variables are *one-hot encoded*, where each value of the variable becomes a binary variable itself, indicating absence or presence of the original variable’s value. Ordinal variables can be either encoded using binary values or 0, 0.5, 1 values depending on the reference interval or normalized in the [0,1] interval. Continuous variables are normalized in the [0, 1] interval.

For many variables, we found a considerable amount of **missing values** that can be classified into ‘zero values’ and ‘missing measurements’. Medications and diagnosis do typically have ‘zero values’ and, in these cases, they indicate that the patient was not prescribed a certain medication or was not diagnosed with a certain feature. On the other hand, laboratory results and vital signs do typically have ‘missing measurements’. Thus, a missing value in blood pressure does not mean that the patient has no blood pressure but, simply, that it was not measured or recorded. To handle the missing values, we use the following criteria: (i) ‘zero values’ are assigned 0 whereas (ii) ‘missing measurements’ are replaced with their previous values for the same patient, if they exist, otherwise, they are filled with the median of the original value over the whole sample.

For **static variables**, the gender variable was binarized and the age variable was normalized between [0, 1]. Diagnosis (HM and H12O) and procedures on admission (H12O) were included as one-hot-encoded values, taking the three first digits of the ICD-10 and ICD-9 for HM and H12O respectively.

For **dynamic variables**, we applied similar methods for both datasets, but with slight variations. For **HM** the information about **ICU stay** was used to create a binary variable indicating, whether the patient is in the ICU or not each day of the hospital stay; **in the case of H12O**, information about **ICU stay** could not be extracted and was not included. **Diagnostics and procedure** variables, encoded as ICD-10 (HM) or ICD-9 (H12O) codes, were one-hot-encoded taking only the first three digits. For **missing measurements**, we experimented with two different ways of imputation: (i) we used the previous day values of the same patient, when available, and the median value of of the dataset (excluding the test set) when not available and (ii) just used median value for all cases. Additionally, we also used the extra binary missing values indicators and also generated extra datasets without this feature. For the **laboratory** determinations, we tried two methods: applying reference values and normalizing between [0,1].

We experimented and generated different dataset representations identified as follows:

- *base*: when using mean for missing measurement imputation.
- *imputation*: when using previous value of a feature for that patient (if available), and mean of the sample (if not), for missing measurement imputation.
- *missing*: a new binary feature is added to indicate if the original value is missing following Che et al. [14].
- *reference*: when using reference values in determination results.

Finally, for a given patient, we joined all static features together to form the final static feature vector **x**_*s*_. Similarly, for a given patient and a given day, we joined all the dynamic features together to create a sequence of dynamic feature vectors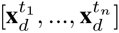 where *t*_*i*_ are the hospital days. Note that, as mentioned above, while creating the dynamic feature vectors, a few extra features may be generated to identify ‘missing measurements’.

### 4.3 Architecture

Motivated by the temporal nature of our problem and the effectiveness of the RNN models in the medical domain [15, 16, 17], we propose a RNN-based model to monitor the mortality risk of the patient by producing a daily prediction during the patient’s hospital stay. To this purpose, we feed each daily records of temporal and static data day-by-day to output a prediction.

As Figure 3 shows, we designed an Artificial Neural Network with four modules, namely, the embedding module, the recurrent module, the classifier module and, optionally, the attention modules.

**Figure 3:**
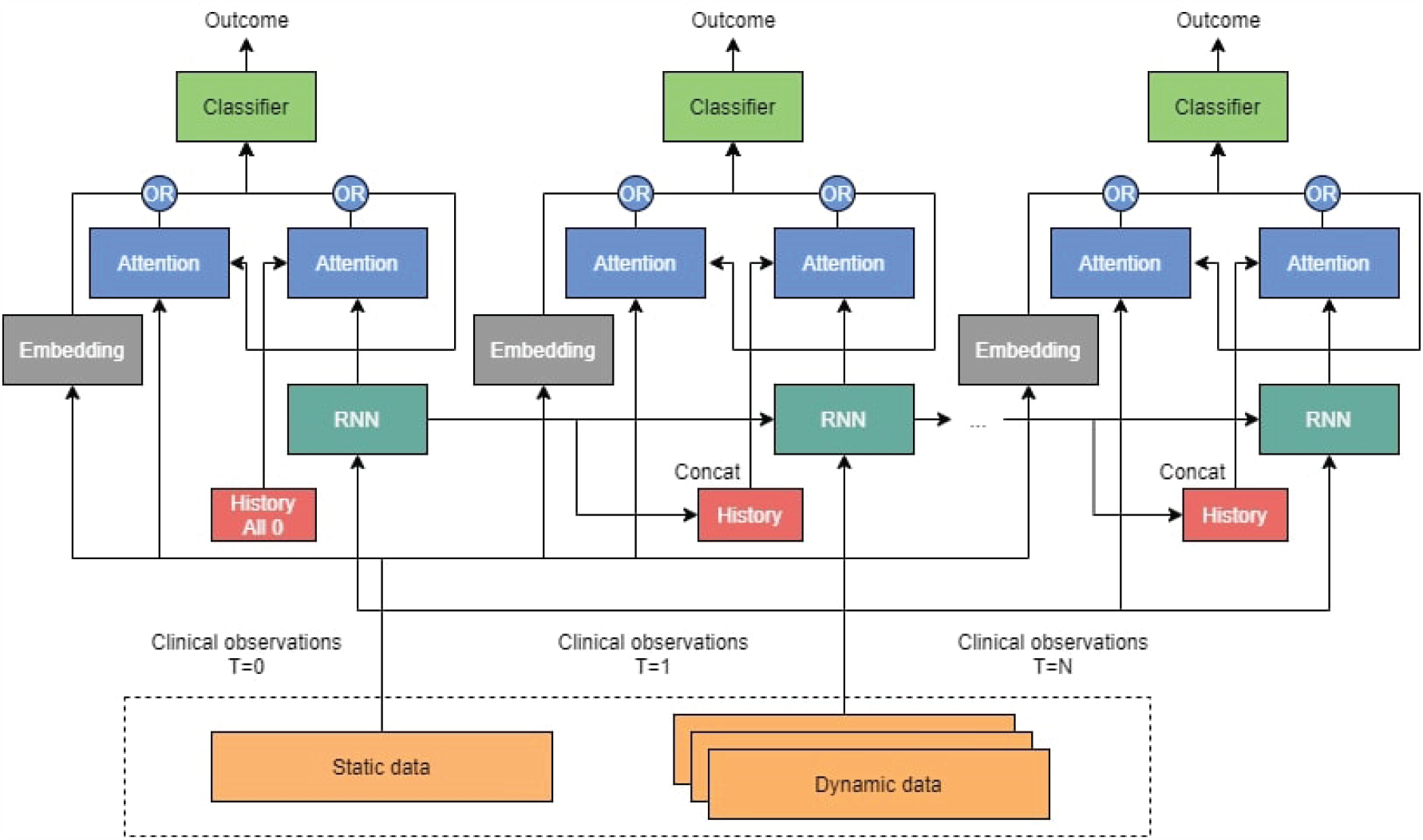
Architecture of the RNN described in Section 4.3.

The embedding module only acts on the static vector through a fully-connected layer that encodes the high-dimensional sparse feature vector into a lower dimensional dense vector.

The recurrent module accounts for both the encoding and the memorization of the temporal information provided by the dynamic vectors (red boxes). It consists of an unidirectional^3^ RNN, built with either an LSTM or a Gated Recurrent Unit (GRU) [18]. At each day, the RNN cells process the relevant information of the patient dynamic data and produce an output vector that stores the relevant clinical information until that day to perform the predictions.

The attention module of dynamic field finds the correlations of all previous RNN’s outputs and merges all the global relevant information of the sequence until a given day. First, the attention module creates a context vector as a linear combination of the RNN’s outputs using the dot-product of the attention scores as in [19]. Then, at every day, the context vector and the current RNN output are concatenated and fed to a fully-connected layer to which it was applied a hyperbolic tangent function to get the final attention vector^4^. The attention module on the static field finds correlations of unembedded static data with respect to the dynamic hidden layer. As we will discuss in the next sections, in our work, the attention module is the component responsible for the interpretability of the model.

Finally, the classifier module consists of two fully-connected layers followed by a sigmoid activation function to produce the binary mortality predictions. We placed the classifier module on top of the RNN cells and the static embeddings to predict the mortality risk at each day.

### 4.4 Training and Inference

Unlike standard training scheme for recurrent models, we designed an ad-hoc training scheme to produce daily predictions. Specifically, we do *not* feed the entire sequence of dynamic data into the RNN to output a single prediction. But, we feed each daily records of temporal and static data day-by-day and output a prediction for each day.

To mitigate the impact of the class-imbalanced data on the learning process, we employed the Focal Loss introduced in [20] besides the classical Vanilla Binary Cross-Entropy. The Focal Loss was originally developed for computer vision as a strategy for countering class imbalance. Based on the assumption that the most frequent label will generate more confident predictions, the Focal Loss reshapes the Vanilla Cross-Entropy through two parameters (*α* and *γ*) such that well-classified samples have less weight than the incorrectly classified ones. During training, we fixed *α* to 1 and used *γ* as hyperparameter.

In addition, as detailed in the Additional Material, we experimented with other model’s hyperparameters configurations. All the layers are trained end-to-end with stochastic gradient descent.

At inference, the model is fed with the static vector and then, at each time step, we input the corresponding dynamic vector and the model outputs the probability of patient mortality. To obtain the mortality predictions, the probability is discretized into a binary label using a threshold, as in logistic regression. We set the threshold to the default value of 0.5. Notice that, after training, other values of the model’s threshold can be used, depending on how much we want to be confident in the prediction. For the metrics used for evaluation, see Section 4.7.

### 4.5 Interpretability

The attention layer is the building block of the interpretability in our model. The attention module of dynamic field relates the RNN’s outputs to the RNN’s output history vectors to retrieve meaningful past dynamic information for the daily prediction. The idea behind is to identify which days in the previous sequence are influencing most the prediction of the current day. The attention can be useful as a starting point to learn which clinical variables could have a critical role. See the animated figure^5^ to visualize how the attention scores dynamically change as sequence evolves and focuses on the most relevant days for a given patient. See the Additional Material to learn about attention analysis.

### 4.6 Experimental Framework

We conducted extensive experiments within an experimental setting that includes: dataset splits, a feature selection mechanism, a proper model’s hyperparameter tuning and model selection, and two competitive baselines.

In particular, we want to point out the role of model tuning as a necessary step considering our case. Since we designed our network from scratch and applied it on a dataset not yet studied, a suitable assessment of the model’s capability through a functional exploration of hyperparameters was required. Analogously, a thorough evaluation helped in testing the robustness of the model. See the Additional Material for detailed descriptions.

### 4.7 Metrics and Evaluation

To evaluate the performance of the models and the consistency of the model’s predictions, we considered the following metrics:

- Accuracy: measures the number of correct predictions over the total number of cases.
- F1 score: harmonic mean of the model’s precision and recall. For this project, we use a macro F1 score, computing the metric for each label, and then finding the unweighted mean. Macro F1 does not take label imbalance into account, and therefore the score for each class counts up to half of the total score.
- Sensitivity: measures the capacity to correctly predict mortality, also known as recall.
- Specificity: proportion of the capacity to correctly predict survivals.
- Area Under the Curve (AUC): probability of a random example with true label of fatality to get a higher score than a random example with true label of survival.

When computing the **global performances**, for each of the metrics above, we used the average of metric computed at every day in the patient’s sequence for all patients.

In addition, and more interestingly, we designed specific evaluation metrics to better test the model and reflect our intended use case providing daily predictions. In this case, we get the prediction vectors already returned by the system, and use them to calculate the daily performances as follows:

- **Daily performances from the admission** computes the performance every day from the admission day.
- **Daily performances from the outcome** computes the performance every day from the outcome day.

Notice that, in daily performance calculations, the system ignores whether the patients are long-term patients or not and there is no leakage of other past or future events; we just evaluate on the predictions already done by the system.

Figure 4 shows an example of how day by day predictions are calculated. To compute the daily performances from admission, we use the predictions of each patient, starting from the left. To compute the daily performances from the outcome, first we align all the predictions to the right, and then, we use the predictions of each patient, starting from the right. In this case, Day 1 is the outcome day.

**Figure 4:**
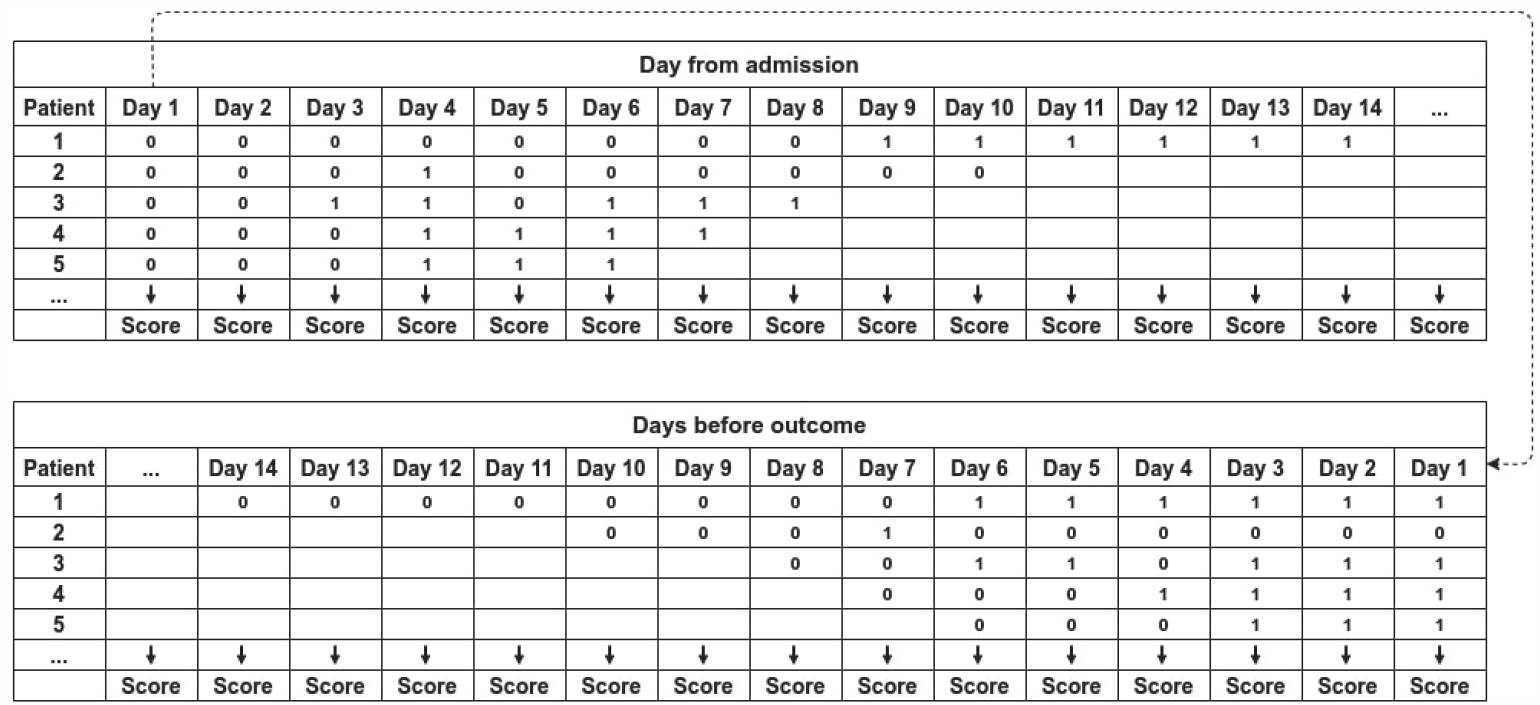
Example of system’s predictions from admission and from outcome.

## Data Availability

HM is available in the provided link.
H12O data is subject to legal restrictions and cannot be currently distributed.
Code is available under MIT license.

https://www.hmhospitales.com/coronavirus/covid-data-save-lives

https://github.com/PlanTL-SANIDAD/covid-predictive-model

## 5 Data availability

HM raw data can be accessed prior application^6^ for access.

The data sharing agreement with H12O does not permit us to make the raw data available to third parties.

## 6 Code availability

The code is available^7^ under MIT licence.

## 7 Acknowledgements

We want to thank,

José Antonio López-Martin from Hospital Universitario 12 de Octubre; Xavier Pastor and M. Jesús Betran from the Hospital Clínic de Barcelona; and Carlos Luis Parra and Sara González from Hospital Virgen del Rocio for their interest and support to the project.

HM Hospitales for sharing the data to the scientific community.

And, finally, our colleague Eulàlia Farré i Maduell, a medical doctor and researcher at the Text Mining Unit at BSC for her support on medical issues.

Hospital 12 de Octubre is supported by PI18/00981 “*Arquitectura normalizada de datos clínicos para la generación de infobancos y su uso secundario en investigación: caso de uso cáncer de mama, cérvix y útero, y evaluación*” funded by the Carlos III Health Institute from the Spanish National plan for Scientific and Technical Research and Innovation 2017-2020 and the European Regional Development Funds (FEDER).

This work was funded by the State Secretariat for Digitalization and Artificial Intelligence to carry out support activities in supercomputing within the framework of the Plan TL^8^ signed on 14 December 2018.

## 9 Author Contribution

Marta Villegas contributed to study design, organized the authors, synthesized the writing, and led the abstract, introduction, results and discussion sections. Aitor Gonzalez-Agirre and Asier Gutiérrez-Fandiño contributed to study design, data analysis and preparation, manuscript preparation and led the experimental framework and evaluation. Jordi Armengol-Estapé contributed to study design, manuscript preparation and led the experimental framework and evaluation Casimiro Pio Carrino contributed to study design, data analysis and preparation, experimental framework, models evaluation and manuscript preparation. David Pérez Fernández contritributed to study design and data analysis and preparation. Felipe Soares contributed to the experimental framework and evaluation. Miguel Pedrera and Noelia García contributed to data extraction, data analysis and preparation. Pablo Serrano and Alfonso Valencia oversaw and advised the work. All authors contributed to significant amendments to the final manuscript.

## Footnotes

1 https://www.mscbs.gob.es/profesionales/saludPublica/ccayes/alertasActual/nCov/documentos/Actualizacion_263_COVID-19.pdf

2 The involved hospitals obtained informed consent from all participants.

3 Given that the order of the causal relationship between the clinical event is strictly temporal, bidirectional RNN layers are not suitable for our use case.

4 The implementation is available in PyTorch: https://pytorchnlp.readthedocs.io/en/latest/_modules/torchnlp/nn/attention.html

5 Attention mechanism animation: https://drive.google.com/file/d/14ivLc1IRql6V9dT-cdddbdpjTbgn0WCN/view?usp=sharing

6 https://www.hmhospitales.com/coronavirus/covid-data-save-lives

7 https://github.com/PlanTL-SANIDAD/covid-predictive-model

8 https://www.plantl.gob.es/

