## Additional Material for "Predicting the Evolution of COVID-19 Mortality Risk: a Recurrent Neural Network Approach"

January 7, 2021

**2 Dataset Splits** **6**

**3 Feature Selection** **6**

**4 Hyperparameter Tuning and Model Selection** **7**

**5 Baseline Models** **7**

**6 Attention Analysis** **8**

**7 Ensemble** **8**

**8 Results** **9**

### 1 Datasets

#### 1.1 HM Hospitals Dataset

The HM dataset contains the anonymized records of 2,307 patients admitted with a diagnosis of COVID POSITIVE or COVID PENDING, since the beginning of the epidemic to April 24th 2020. HM Hospitales opened this anonymous dataset freely available to the medical and scientific community with clinical information on patients treated in their hospital centers. The dataset includes demographic data and information about medication, vital signs, laboratory, diagnostics, procedures and ICU stay.

Of the 2,307 patients, 1,377 are males and 930 are females, around 60% and 40% respectively. The minimum and maximum ages are 0 and 106 years, with a mean of 67.6 (see Figures 1 and 2). The mortality rate is 14.43% (16% for males and 12% for females). Of the total of 333 deaths, 218 are males and 115 are females, being the 65% and the 35% of the death population and the 9% and 6% of the whole patients.

The observed mean duration from admission to outcome (death, discharge, or other reason) is 8.08 days with a standard deviation (SD) of 6.18 days and a median of 7 days.

160 patients (7% of the records) were admitted to the ICU (9% for males and 4% for females) with the shortest stay being one day and the longest being 35 days, with a mean of 8.97 days, SD of 8.66 and median of 6 days. Of the total of 160 patients admitted to the ICU, 120 are male and 40 are female, being the 75% and the 25% of the ICU population and the 5% and the 2% of all patients.

Figures 1 and 2 show the age and sex distribution of patients in relation to mortality and ICU stay. Table 1 shows the number of occurrences by days for each label in the TEST set. This table evidences the differences in death ratios in the two datasets

Information about **medication** administered during admission includes the dates for the first and last administration of each drug. Medications are identified by their brand name and Anatomical Therapeutic Chemical (ATC) 5 and 7 classification codes. The data includes 60,460 rows with 275 and 464 ATC5 and ATC7 unique codes. During the hospital stay, the minimum number of medications per patient is 1 and the maximum is 183, with a mean of 26 and a median of 22. Table 2 summarizes the medication distributions.

The **vital signs** include heart rate, maximum and minimum blood pressure and temperature collected during admission, together with their date and time of registration. The minimum number of registrations per patient is 1 and the maximum is 190, with a mean of 24.5 registrations and a median of 21 as summarised in Table 2. There are 37,861 records other than 0s for heart frequency; 21,174 and 21,166 records for maximum and minimum blood pressure and 54,431 records for temperature.

The information about **laboratory** contains all the results of the requests made to each patient during admission and in the previous emergency, if any. This includes 398,884 results for 401 different determinations unevenly distributed. As shown in Figure 3, the 82% of determinations have less than 500 reported results and there is a long tail of really infrequent determinations. 2,080 patients have at least one lab result with a maximum of 4,395 and a mean of 190 and a median of 111 as summarized in Table 2.

Finally, the dataset also contains **diagnoses** present on admission using the International Classification of Diseases (ICD-10)[1]. Though the dataset also contains diagnoses for episodes of hospital admission, these were not considered in the experiment as they had no time information associated. Only the diagnoses marked as being present on admission were included, as part of the static variables.[2].

[1]Does not include COVID.

[2]Note that including diagnoses that lack temporal information would not be fair since we could be introducing information from future events.

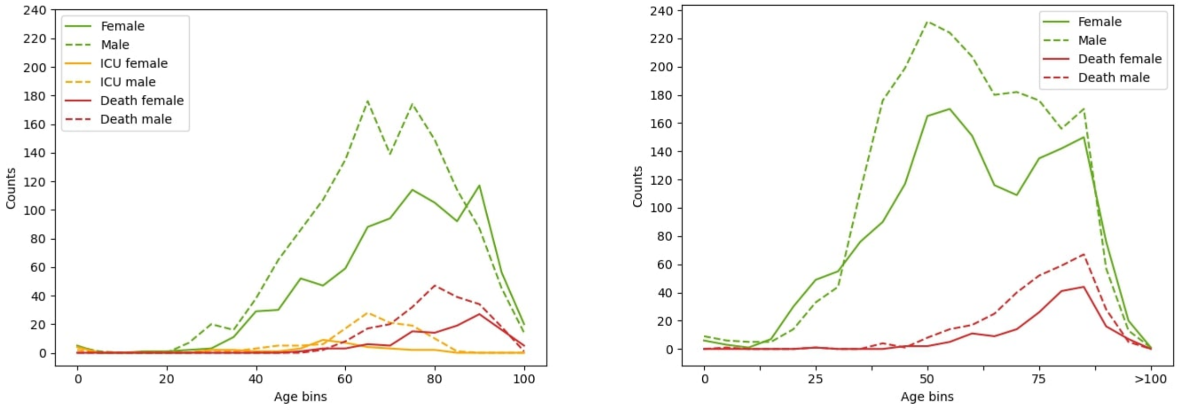

(a) HM (b) H12O

Figure 1: Age and sex distribution of patients in HM and H12O datasets. Solid lines are for females and dotted lines for males. Green lines show number of patients, read lines show the number of dead patients. For HM, orange lines show the number of 'in ICU' patients.

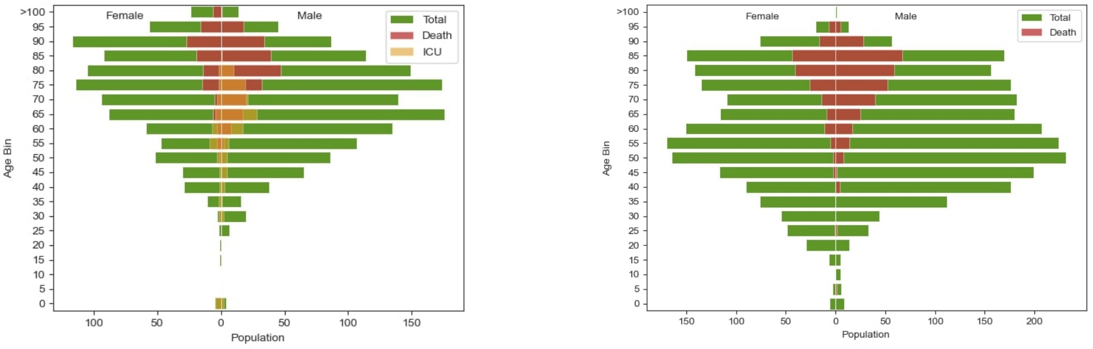

(a) HM (b) H12O

Figure 2: Age and sex distribution of patients in HM and H12O dataset. Left females, right males. Bar fills are the number of male/female (green) and dead male/female patients (red). For HM, also 'in ICU' male/female patients (orange).

| HM | 1 | 2 | 3 | 4 | 5 | 6 | 7 | 8 | 9 | 10 | 11 | 12 | 13 | 14 | 15 | 16 | 17 | 18 | 19 | 20 |
|---|---|---|---|---|---|---|---|---|---|---|---|---|---|---|---|---|---|---|---|---|
| Alive | 161 | 161 | 157 | 151 | 133 | 117 | 94 | 81 | 63 | 49 | 42 | 38 | 36 | 29 | 24 | 21 | 17 | 12 | 7 | 6 |
| Death | 33 | 33 | 30 | 29 | 27 | 25 | 23 | 19 | 16 | 13 | 12 | 12 | 8 | 8 | 7 | 7 | 6 | 4 | 4 | 4 |
| H12O | 1 | 2 | 3 | 4 | 5 | 6 | 7 | 8 | 9 | 10 | 11 | 12 | 13 | 14 | 15 | 16 | 17 | 18 | 19 | 20 |
| Alive | 324 | 324 | 324 | 317 | 293 | 257 | 221 | 177 | 138 | 117 | 102 | 86 | 78 | 63 | 56 | 49 | 45 | 37 | 32 | 29 |
| Death | 50 | 50 | 50 | 47 | 39 | 34 | 29 | 24 | 21 | 16 | 15 | 13 | 13 | 12 | 11 | 10 | 9 | 8 | 6 | 6 |

Table 1: Number of occurrences by days for each label.

| HM | | | | | |
|---|---|---|---|---|---|
| Metric | Medications | Vital sign records | Determinations | Diagnoses | Procedures |
| Mean | 26.28 | 24.51 | 189.66 | 7.44 | 6.69 |
| Standard Deviation | 0.40 | 0.37 | 6.56 | 4.28 | 2.69 |
| Median | 22 | 21 | 111 | 7 | 6 |
| Range | 182 | 189 | 4,394 | 18 | 19 |
| Minimum | 1 | 1 | 1 | 1 | 1 |
| Maximum | 183 | 190 | 4,395 | 19 | 20 |
| Sum | 60,460 | 55,515 | 394,490 | 13,213 | 11,878 |
| Count (patients) | 2,301 | 2,265 | 2,080 | 1,775 | 1,775 |
| Distinct | 275/464 | 4 | 401 | 1,363 | 251 |
| **H12O** | | | | | |
| Metric | Medications | Vital sign records | Determinations | Diagnoses | Procedures |
| Mean | 24.68 | 113.51 | 174.10 | 4.37 | 2.17 |
| Standard Deviation | 26.03 | 96.74 | 258.15 | 5.98 | 3.07 |
| Median | 16 | 87 | 108 | 2 | 1 |
| Range | 341 | 1,538 | 3,568 | 76 | 23 |
| Minimum | 1 | 1 | 1 | 1 | 1 |
| Maximum | 342 | 1,539 | 3,622 | 77 | 24 |
| Sum | 95,439 | 433,743 | 652,038 | 7,031 | 383 |
| Count (patients) | 3,867 | 3,821 | 3,745 | 1,607 | 176 |
| Distinct | 798 | 9 | 80 | 397 | 18 |

Table 2: Number of medications, vital sign records, determinations, diagnoses and procedures per patient in HM and H12O. Medications from HM are in ATC5 codes whereas in H12O they are in ATC7 (for the sake of comparison we show the number of distinct ATC7 codes in HM). Diagnoses and Procedures in HM include only those Present On Admission whereas in H12O their timestamp goes from 1st 2019 to discharge date. Diagnoses and Procedures codes from HM are in ICD-10 whereas in H12O they are ICD-9 encoded.

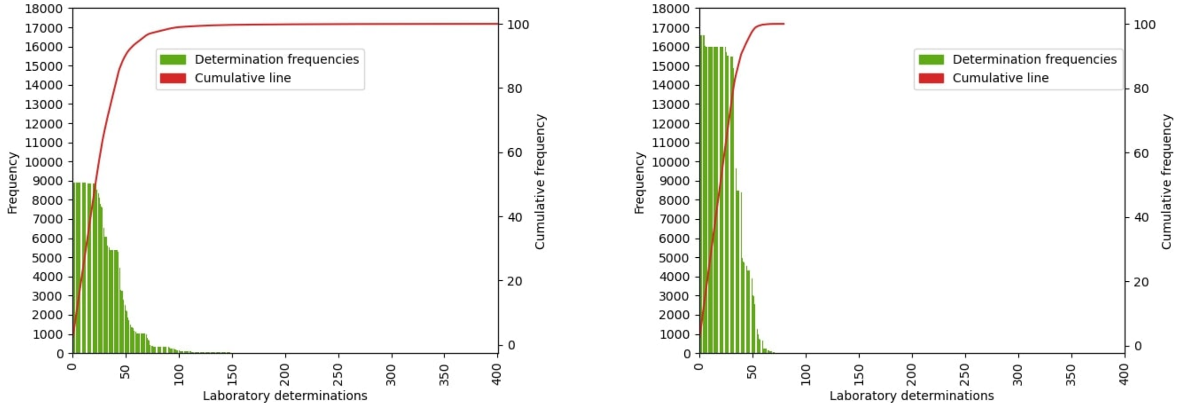

(a) HM

(b) H12O

Figure 3: Distribution of laboratory determinations in HM and H12O datasets in descending order with a cumulative line on the left axis as a percentage of the total.

##### 1.2 Hospital 12 de Octubre Dataset

The H12O dataset contains the anonymized records of 3,870 COVID patients since the beginning of the epidemic to October 14th. The dataset includes demographic data and information about medication, vital signs, laboratory, diagnostics and procedures.

Of the 3,870 patients, 2,201 are males and 1,669 are females, around 57% and 43% respectively. The minimum and maximum ages are 0 and 103 years, with a mean of 61.4 (see Table 2 and Figure 1). The mortality rate is 13% (14.63% for males and 10.67% for females). Of the total of 500 deaths, 322 are males and 178 are females, being the 64% and the 36% of the death population and the 8% and 5% of the whole patients.

The observed mean duration from admission to outcome (death, discharge, or other reason) is 9.51 days with SD 4.99 days and a median of 8 days, which is slightly longer than HM.

Figures 1 and 2 show the age and sex distribution of patients in relation to mortality and ICU stay. Table 1 shows the number of occurrences by days for each label in the TEST set. This table evidences the differences in death ratios in the two datasets

**Medications** in H12O dataset are represented in ATC7 coding, with 798 unique medications. The minimum number of medications per patient is 1 and the maximum is 342, with a mean of 24,68 per patient and SD of 26.03 and MD of 16.

**Vital signs** used in the H12O dataset include heart rate, breathing frequency, O2 saturation, temperature, mean blood pressure, diastolic tension, systolic tension, O2 saturation previous to intubation and minimum O2 saturation during intubation. There is a mean of 113.51 records per patient with an SD of 96.74 and a MD of 87. The minimum vital signs record for a patient is 1 and the maximum is 190. The total vital signs recorded are 433,743 which is much higher than in the case of HM because H12O has more vital signs (9 vs. 4) and more patients (3,821 vs. 2,265).

Records regarding **laboratory determinations** count 80 unique determinations, with a mean of 174.1 measurements per patient, a SD of 258.15 and a MD of 108. The minimum laboratory determinations for patients is 1 and the maximum is 3,622. As Table 2 and Figure 3 show, H12O has less unique determinations but higher amount of records than HM. HM has more distinct laboratory determinations but some of them have negligible appearances and, therefore, it shows a longer cumulative line.

Finally, for both **diagnoses** and **procedures**, we take the (ICD-9) codes from January 1st of 2019. There are 397 and 18 unique codes respectively distributed among patients with a mean of 4.37 diagnoses and 2.17 procedures, SD 5.98 and 3.07 and median of 2 and 1.

Table 2 summarizes the medications, vital signs and determination distributions.

##### 1.3 Data Cleaning

To guarantee the quality of the data we performed some data cleaning processes as follows:

- Temporal sequences of hospital stays are very varied in length. In order to reduce the length of the sequences, without removing the samples (i.e patients), sequences were cut to 20 events from outcome (last 20 events).
- Only patients that were sent home or resulted in death are taken into account. Other discharge reasons, like transfers to other hospitals, do not provide clear and explicit information about the outcome.
- For all variables we perform an IQR for each column; we define Q1 as a quantile of 0.01 and 0.03, and Q3 as a quantile of 0.99 and 0.97, respectively for HM and H12O, having $IQR = Q3 - Q1$, we set to missing values those values that were not between $[Q1 - 1.5 * IQR, Q3 + 1.5 * IQR]$.
- Binary columns, with values showing a frequency lower than 1% are removed, once the dataset is generated. Notice that this can include missing indicator columns.
- In HM the **medication** table, "XXXXX"[3] codes and codes starting with "D"[4] were removed, since they represent nested medications and dermatological medications, respectively, that do not provide additional meaningful information as stated by the medical doctor.

As a result of this process, the final dataset includes samples from 1,939 and 3,735 patients for HM and H12O respectively. For HM and H12O, the resulting vectors have different sizes as we generated different datasets combining different strategies (see Section 4), the default datasets have 54 and 55 features in $\mathbf{x}_s$, 177 and 246 in $\mathbf{x}_d$, label vector $\mathbf{y}$ contains 1 column. In the eventual datasets, the mortality rate increased x to 16.92% in HM (328 out of 1,939) and was unchanged 13.38% in H12O (500 out of 3,735).

[3]"XXXXX" code had 1,289 appearances.

[4]Codes starting with "D" had 6,904 appearances.

#### 2 Dataset Splits

We created two dataset splits that were applied in different experimental settings, as follows:

- Initial split: We splitted the dataset in train (TRAIN-GRID), validation (VALID-GRID) and TEST sets, with proportions of 80% for training, 10% for validation, and 10% for test. The dataset is split randomly[5]. The split is stratified as in [1], that is, respecting the proportion of the labels in the different subsets [2], which is crucial in the case of class imbalanced datasets. We use the TRAIN-GRID and VALID-GRID sets to perform a grid search of the hyperparameters. The TEST set is reserved and only used for the final evaluation of the system.
- Cross-validation split: Using the train and validation sets of the initial split, we apply a 5-fold cross-validation, with the same proportions as before, and with stratification as well (the size of each train and valid sets is the same to the ones used in the grid search). To avoid repeating the split used in the grid search, we used a different seed this time, obtaining 5 different splits for train (TRAIN-CV) and validation (VALID-CV) sets. We train each fold on the TRAIN-CV set, stopping based on the score of the appropriate metric on the VALID-CV set. The validation average score is used for selecting the best model, and then the TEST set (unused until now) is used for evaluating it.

All the models in our work are trained and evaluated with the same splits. The schema of the dataset split is shown Figure 4.

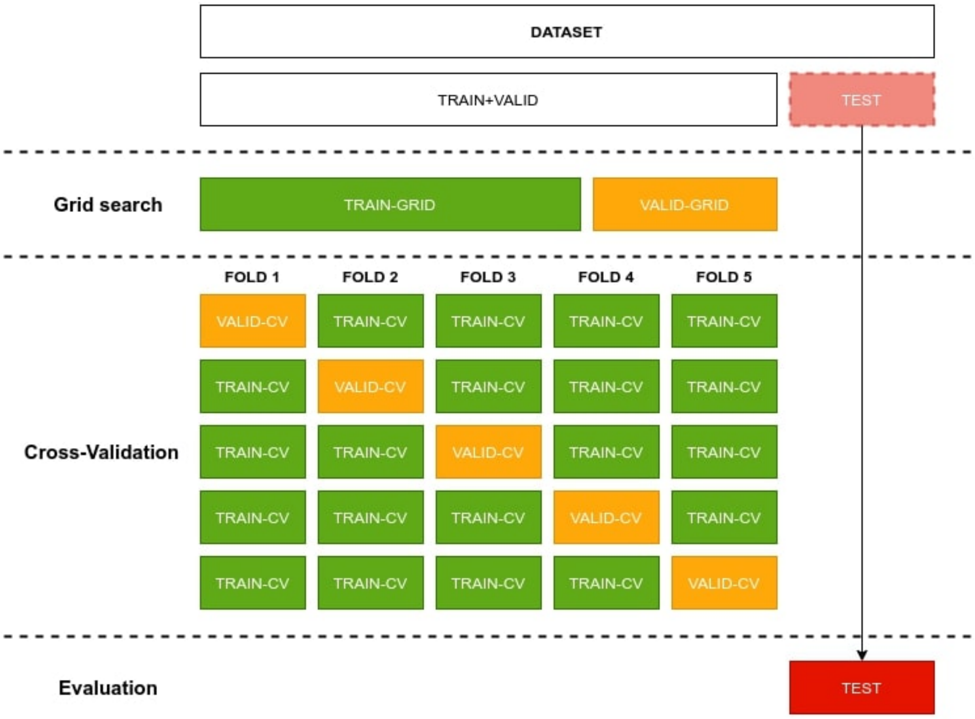

Figure 4: Dataset split of our experimental framework. The size of the train and validation sets is the same for each fold and for the grid search. None of the validation sets used in the CV is the one used in the grid search.

#### 3 Feature Selection

Given the small number of examples and the resulting high-dimensionality of the feature vectors, we decided to use features selection techniques to reduce the dimensionality by removing irrelevant features and, thus, avoid the problems related to the curse of dimensionality and overfitting.

[5]Making sure to use a fixed random seed, for the sake of reproducibility.

We started with the removal of binary columns with values less than $threshold = 1\%$ as explained in Section 1.3 for both static and dynamic vectors. This removes variables with insufficient variance.

Following [3], for dynamic variables, we used metrics of information theory, namely, entropy, information gain, Gini index and chi-square to rank features according to their deemed importance. Next, we select the final features by joining together the first top-k features for each metric. The result is a list of N features that we expect to be relevant for the classification problem. The number of the first top-k is a threshold that determines how many features are selected and we considered it as another RNN model's hyperparameter. For computing the metrics we did not use the test set as it should be maintained unseen until evaluation.

#### 4 Hyperparameter Tuning and Model Selection

To test the capabilities of our model, we organized the experiments in two phases. First, we used the model's hyperparameter tuning to select the optimal values that improve the learning process. Then, we used the model selection to find out which are the best models based on the evaluation metrics and statistical tests.

We ran all the experiments in the CTE-Power[6] High-Performance Computing resource equipped with Nvidia V100 Graphical Processing Units and IBM Power9 8335-GTH Central Processing Units.

| RNN's params | SVC's params | RF's params |
|---|---|---|
| Batch size | Number of estimators | C value |
| Dropout | Criterion | Kernel |
| Static embedding size | Minimum sample split | Tolerance |
| Dynamic embedding size | Class weighting | Gamma |
| RNN hidden state size | | Degree |
| RNN architecture (either LSTM or GRU) | | Class weighting |
| Number of RNN layers | | |
| Loss function (and $\gamma$ value for the Focal Loss) | | |
| Early stopping metric | | |
| Attention mechanism | | |
| Learning rate | | |
| Number of features (top-k) | | |

Table 3: Hyperparameters used in the grid search.

**Model Selection and Tuning:** We employed a grid search technique consisting of an exhaustive search over a manually specified configuration of hyperparameters representing a subset of the hyperparameter space and trained a model for each configuration. Table 3 shows the complete list of hyperparameters for RNN models.

For the RNN, we performed the grid search by training for 200 epochs with early stopping on validation accuracy and F1. We set the the early stop patience to 10 steps. For the baseline models, we executed a grid search with the TRAIN+VALID split (see Sec. 2). Next, we selected the top-30 best models (for both baselines and RNN) with the highest scores on the VALID-GRID set. In the case of H12O, we selected the top-30 best models for each combination of the Feature Representation strategies explained in the main article.

Finally, we employed cross-validation using the cross-validation split explained in 2. We used the average validation accuracy or F1 (depending on the early stopping metric) of the selected models to find the best one (i.e. the one that shows the higher generalization capabilities).

#### 5 Baseline Models

We trained two classical ML models: Random Forest[7] (RF) and Support Vector Machine for classification[8] (SVC). For each model, we designed two experiment types, EXP1 and EXP2, depending on how the data are fed to the model. In

[6] `https://www.bsc.es/user-support/power.php`

[7] RF: `https://scikit-learn.org/stable/modules/generated/sklearn.ensemble.RandomForestClassifier.html`

[8] SVC: `https://scikit-learn.org/stable/modules/generated/sklearn.svm.SVC.html`

EXP1, for each day, we concatenated the static and the dynamic vector. As a consequence, we have independent data representations for each day, where each event of a patient generates a sample (i.e. a patient with a stay of 17 days generates 17 samples).

In EXP2, for each day, we merged the information from all the previous days into a fixed-size vector. We obtained the vector by performing min, max and average operation between the different feature vectors and concatenating them to the static and dynamic vectors[9]. Concretely, given a static vector $\mathbf{x}_s$ and the sequence of dynamic vectors $[\mathbf{x}_d^{t_1}, ..., \mathbf{x}_d^{t_n}]$, the merged vectors for each day $k \in [1, N]$ are computed as follows:

$$\begin{aligned}
\mathbf{x'}^{t_1} &= \mathbf{x}_s \oplus \mathbf{x}_d^{t_1} \oplus min(\mathbf{x''}_d^{t_1}) \oplus max(\mathbf{x''}_d^{t_1}) \oplus avg(\mathbf{x''}_d^{t_1}) \\
&... \\
\mathbf{x'}^{t_k} &= \mathbf{x}_s \oplus \mathbf{x}_d^{t_k} \oplus min(\mathbf{x''}_d^{t_1}, ..., \mathbf{x''}_d^{t_k}) \oplus max(\mathbf{x''}_d^{t_1}, ..., \mathbf{x''}_d^{t_k}) \oplus avg(\mathbf{x''}_d^{t_1}, ..., \mathbf{x''}_d^{t_k}) \\
&... \\
\mathbf{x'}^{t_N} &= \mathbf{x}_s \oplus \mathbf{x}_d^{t_N} \oplus min(\mathbf{x''}_d^{t_1}, ..., \mathbf{x''}_d^{t_N}) \oplus max(\mathbf{x''}_d^{t_1}, ..., \mathbf{x''}_d^{t_N}) \oplus avg(\mathbf{x''}_d^{t_1}, ..., \mathbf{x''}_d^{t_N})
\end{aligned} \tag{1}$$

where the $min$, $max$ and $avg$ functions are applied on the same vector's components in the sequence, and **x''** represents the dynamic vectors after removing the 'missing' features.

In this way, the resulting sequence of merged vectors $[\mathbf{x'}^{t_1}, ..., \mathbf{x'}^{t_n}]$ is supposed to propagate information across all the days of the patient's hospital stay. The purpose of this experiment was to train a model that was at some extent equivalent to temporal RNN-based model and to make a comparison with our proposed recurrent model.

Notice however, that since the **results with EXP2 were systematically worse we decided not to include them**. In Section the main article, we explain how EXP1 is used in the baseline models. The results of the baselines are included in the paper for comparison with our model. The function included in *Scikit-learn* to perform the cross validation[10] does not return the predictions, only the scores; therefore, some day-by-day results may show minor inconsistencies.

#### 6 Attention Analysis

In order to understand how attention mechanism works for dynamic data in our model, we identified and characterized attention peaks. We hypothesize that the attention mechanism should target relevant days to perform correctly. If, for a given patient, there is a relevant day for the prediction, the attention mechanism should always focus on that day (during the whole sequence). Since the RNN runs with daily data, for each day, the model considers the attention of the dynamic embedding of the current day with respect to the previous days. This results in a 2-dimensional matrix.

We first start by finding peaks in the second dimension (i.e. grouping by the target day). Then, we group the days and we count the number of times they are peaks. Since first days appear more times than last days, a normalization is performed. For instance, the first day appears as many times as the length of the sequence; on the contrary, the second to last day only appears once. We normalize by doing $day\_count\_norm = day\_count/(patient\_length - day)$.

For this analysis, we use the best RNN with attention model and the test set of the TRAIN+VALID split.

#### 7 Ensemble

With the motivation of increasing the performance, stability, and robustness to noise, we built two ensembles of RNNs. The first aims at maximizing F1 score (Ensemble-F1), and the second focuses on sensitivity (Ensemble-SEN). The latter had some constraints to avoid the trivial solution of declaring all patients as dead.

In order to build the ensembles, we aggregated the predictions of the models following a voting schema. Specifically, for a given patient and the corresponding votes of the models, the final prediction of the ensemble is defined as follows:

$$\texttt{daily\_outcome\_patient(votes)} = \begin{cases} \texttt{'Dead'}, & \text{if } \frac{\texttt{count(votes = 'Dead')}}{\texttt{count(votes)}} \geq \lambda \\ \texttt{'Alive'}, & \text{otherwise} \end{cases}$$

where $\lambda$ is the threshold of the percentage of models required to predict whether a given patient will die. It follows that for a given patient and ensemble, the lower the $\lambda$, the better (or at least equal) the sensitivity.

[9]In order to reduce the number of features, we removed the 'missing' features before computing the min, max, and average vectors ('missing' features are still in the dynamic vector that represents the current day).

[10]Cross validation: `https://scikit-learn.org/stable/modules/generated/sklearn.model_selection.cross_validate.html`

To build the ensembles, we proceed with the following steps:

1. First, we pre-select the top 100 models, as per cross-validated accuracy or F1 score. This cut-off guarantees that the models integrating the ensemble will meet a reasonable performance requirement.
2. Then, we select 20 of them using an algorithm. Instead of just taking the top 20 best models as per cross-validated F1 or accuracy, we define an algorithm that heuristically maximizes the differences in predictions in the cross-validation folds of the different models. Otherwise, it could be the case that all models made too similar predictions, making the ensemble unnecessarily grow in number of models.
3. Finally, we optimize the number of models and the aggregation threshold, $\lambda$. Specifically, we apply a grid search over the following values:
   - `Number of models` $= \{2..20\}$: We need at least 2 models to build an ensemble, but having a large number of models might be infeasible in real-life settings, so, as we said, we restricted the ensemble to a maximum of 20 models. We experiment with adding the models in order from the algorithm used, and discard the ones that do not improve.
   - $\lambda = \{0.1, 0.2, 0.3, 0.4, 0.5\}$: One of the main goals was to improve the sensitivity, therefore it made no sense to try values larger than 0.5.

   Thus, the final configuration maximizes the F1 score, in case of Ensemble-F1, or sensitivity, in case of Ensemble-SEN.

**Algorithm 1:** Selection of $n$ candidate models for ensemble, with a cut-off of $top$ models

```
Result: candidate_models
Input: n: integer
Input: top: integer
Input: metric: {'f1', 'accuracy'}
candidate_models ← List();
candidate_models_indices ← List();
all_predictions ← get_flattened_all_predictions_in_cv_sorted_by_model_score_in_cv(metric)[:top] ;
prediction_similarity_matrix ← distance_matrix(all_predictions, all_predictions) ;
all_models ← get_all_models_sorted_by_cv_score(metric)[:top];
best_model ← all_models[0];
candidate_models.insert(best_model);
candidate_models_indices.insert(0);
while candidate_models.length() < n do
    max_distance ← 0;
    max_index ← null;
    for i = 0; i++; i < all_models.length() do
        new_candidate ← all_models[i];
        if new_candidate in candidate_models then
            break;
        end
        for j = 0; j++; j < candidate_models.length() do
            distance ← distance + prediction_similarity_matrix[i, candidate_models_indices[j]];
        end
        if max_index is null or distance > max_distance then
            max_distance ← distance;
            max_index ← i;
        end
    end
    candidate_models_indices.insert(max_index);
    candidate_models.insert(all_models[max_index]);
end
```

### 8 Results

#### 8.1 Best hyperparameters

Table 4 shows the best hyperparameters for RF and SVC baselines.

| RF Parameter | HM | H12O | SVC Parameter | HM | H12O |
|---|---|---|---|---|---|
| Class weighting | Balanced | None | C | 3 | 1.2 |
| Criterion | Gini | Gini | Class weighting | None | None |
| Minimum sample splits | 10 | 9 | Gamma | Scale | Scale |
| Nº estimators | 125 | 175 | Kernel | RBF | Poly |
| | | | Tolerance | 0.01 | 0.01 |

Table 4: HM and H12O hyperparameters for RF and SVC baselines.

In the case of the RNN-based models, in the top 60 configurations (top 30 by accuracy plus top 30 by F1 combined, in the validation set) for each dataset, we observed that:

- Most configurations used attention: 60 models in HM, 53 in H12O.
- Most configurations used LSTM instead of GRU in H12O (56 vs. 4). In HM, most configuration used GRU (36 vs 24), but is more equitably distributed.
- Most configurations used the focal loss instead of the vanilla binary cross-entropy (45 vs. 15 in HM, 46 vs. 14 in H12O), so the former seems a better choice for handling the class imbalance found in our data.
- Most configurations used a batch size of 16 instead of 32 (58 vs. 2 in HM, 38 vs. 22 in H12O).
- Most configurations used a learning rate of 0.001 (46 in HM and 53 in H12O).
- Dropout values are significantly different for both dataset. In HM, a dropout of 0.1 (37) was more frequent than a dropout of 0.2 (23 in both cases); a dropout of 0.4 was never used. In H12O, the dropouts of 0.1 and 0.2 practically tied (29 vs. 28, respectively), and a dropout of 0.4 was the less frequent (3 configurations).
- In the case of HM, the static embedding is considerably more effective when using an embedding size of 16 instead of 32 (37 vs. 23). As in the case of the dynamic embedding size, there are no big differences in the case of H12O (29 used an embedding size of 16, and 31 and embedding size of 32).
- For both HM and H12O, slightly greater number of models used dynamic embedding size of 64 instead of 32 (32 vs. 28 in HM and 35 vs. 25 in H12O).
- Finally, regarding feature selection, in HM, using the top 100 features proved to be considerably more effective (29 models) than 150 features (6 models) or 300 (10 models). In H120, using the top 100 features was the most used configuration (20 models), followed by 150 features (16 models) and 300 features (8 models). Discarding too many features induced a loss of task-relevant information, while keeping too many of them made more difficult the learning process. In the case of HM, 15 models did not use feature selection, and in the case of H120, 16 models did not use feature selection.

##### 8.2 Best RNN Configuration

For **HM**, best RNN-based model in terms of F1 is an ensemble that maximizes F1. This ensemble chooses among the 100 models with the highest accuracy in the VALID-CV split, combining 10 models with a $\lambda$ of 0.3. The ensemble that maximizes sensitivity is obtained analogously, but using sensitivity instead of F1 for discarding combinations that lower the score of the ensemble. This model combines 10 models, less than the maximum of 20 models, using a $\lambda$ of 0.1.

For **H12O**, the best RNN-based model in terms of F1 is an ensemble that maximizes F1. It combines 20 models with a $\lambda$ of 0.2. The ensemble that maximizes sensitivity combines 10 models, also using a $\lambda$ of 0.1.

These results demonstrate that we can build RNN ensemble models with a very high performance combining only 10 configurations.

##### 8.3 Sensitivity Global Results by Folds

| Mod | Imp | Mis | Ref | FS | Met | Fold 1 | Fold 2 | Fold 3 | Fold 4 | Fold 5 |
|---|---|---|---|---|---|---|---|---|---|---|
| RNN | ✔ | × | × | × | F1 | 0.6938 | 0.6406 | 0.6969 | 0.6844 | 0.6719 |
| RF | × | × | × | - | Acc | 0.6312 | 0.6094 | 0.6281 | 0.6031 | 0.6125 |
| SVC | ✔ | ✔ | × | - | F1 | 0.7781 | 0.7844 | 0.8000 | 0.7781 | 0.7625 |
| RNN | Ensemble-F1 | | | | Acc | 0.7813 | 0.7188 | 0.7438 | 0.7375 | 0.7656 |
| RNN | Ensemble-SEN | | | | F1 | **0.8500** | **0.8156** | **0.8438** | **0.8563** | **0.8469** |

Table 5: HM, sensitivity by folds of RNN, RNN & attention, SVC and RF.

| Mod | Imp | Mis | Ref | FS | Met | Fold 1 | Fold 2 | Fold 3 | Fold 4 | Fold 5 |
|---|---|---|---|---|---|---|---|---|---|---|
| RNN | ✔ | ✔ | × | 150 | Acc | 0.6241 | 0.6976 | 0.6220 | 0.7386 | 0.6673 |
| RF | ✔ | ✔ | × | - | Acc | 0.6242 | 0.6199 | 0.6307 | 0.6112 | 0.6415 |
| SVC | ✔ | ✔ | ✔ | - | Acc | 0.6523 | 0.6350 | 0.6415 | 0.6263 | 0.6436 |
| RNN | Ensemble-F1 | | | | Acc | 0.7192 | 0.7257 | 0.7322 | 0.7927 | 0.7473 |
| RNN | Ensemble-SEN | | | | F1 | **0.7711** | **0.8035** | **0.7862** | **0.8531** | **0.8272** |

Table 6: H12O, sensitivity by folds of RNN, RNN & attention, SVC and RF.

##### 8.4 Comparing Global Results Between Datasets

Figure 5 compares the results in the two datasets. We can observe that for accuracy, specificity and F1 the models show similar results in both datasets. However, for sensitivity they show appreciable differences: RF shows a similar low performance in both datasets; RNN is better in HM but still above 0.80 in H12O; but SVC shows a dramatic drop in H12O, with a performance that is practically on par with the RF. We have to be cautious when comparing two different datasets, but it seems that with more features and more examples, the SVC fails to generalize.

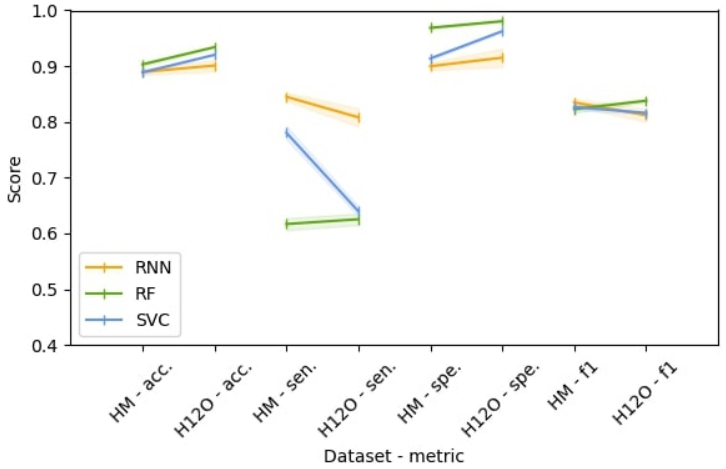

Figure 5: HM-H12O comparison in accuracy, sensitivity, specificity, and F1.

#### 8.5 Daily Results for HM

##### 8.5.1 Random Forest

| Day | Corrects | Totals | Accuracy | AUC | Sensitivity | Specificity | F1 |
|---|---|---|---|---|---|---|---|
| 1 | 169.8±1.3038 | 194 | 0.8753±0.0067 | 0.6960±0.0186 | 0.4242±0.0371 | 0.9677±0.0028 | 0.7319±0.0188 |
| 2 | 170.2±0.8367 | 194 | 0.8773±0.0043 | 0.6876±0.0102 | 0.4000±0.0254 | 0.9752±0.0076 | 0.7276±0.0092 |
| 3 | 168.0±0.7071 | 187 | 0.8984±0.0038 | 0.7373±0.0023 | 0.5000±0.0000 | 0.9745±0.0045 | 0.7769±0.0056 |
| 4 | 167.2±0.8367 | 180 | 0.9289±0.0046 | 0.7988±0.0140 | 0.6069±0.0308 | 0.9907±0.0059 | 0.8460±0.0108 |
| 5 | 149.0±1.4142 | 160 | 0.9313±0.0088 | 0.8170±0.0248 | 0.6444±0.0497 | 0.9895±0.0041 | 0.8595±0.0210 |
| 6 | 133.6±1.3416 | 142 | 0.9408±0.0094 | 0.8540±0.0212 | 0.7200±0.0400 | 0.9880±0.0047 | 0.8877±0.0190 |
| 7 | 109.4±0.8944 | 117 | 0.9350±0.0076 | 0.8676±0.0194 | 0.7565±0.0389 | 0.9787±0.0000 | 0.8903±0.0149 |
| 8 | 92.2±0.8367 | 100 | 0.9220±0.0084 | 0.8350±0.0220 | 0.6947±0.0440 | 0.9753±0.0000 | 0.8622±0.0176 |
| 9 | 71.8±0.8367 | 79 | 0.9089±0.0106 | 0.8310±0.0251 | 0.7000±0.0523 | 0.9619±0.0087 | 0.8500±0.0198 |
| 10 | 56.4±0.8944 | 62 | 0.9097±0.0144 | 0.8468±0.0241 | 0.7385±0.0421 | 0.9551±0.0091 | 0.8588±0.0230 |
| 11 | 49.0±0.0000 | 54 | 0.9074±0.0000 | 0.8512±0.0000 | 0.7500±0.0000 | 0.9524±0.0000 | 0.8619±0.0000 |
| 12 | 44.0±0.7071 | 50 | 0.8800±0.0141 | 0.8298±0.0209 | 0.7333±0.0373 | 0.9263±0.0118 | 0.8336±0.0201 |
| 13 | 38.2±0.4472 | 44 | 0.8682±0.0102 | 0.7736±0.0062 | 0.6250±0.0000 | 0.9222±0.0124 | 0.7765±0.0126 |
| 14 | 31.6±0.8944 | 37 | 0.8541±0.0242 | 0.7802±0.0346 | 0.6500±0.0559 | 0.9103±0.0189 | 0.7828±0.0348 |
| 15 | 26.2±0.4472 | 31 | 0.8452±0.0144 | 0.7583±0.0319 | 0.6000±0.0639 | 0.9167±0.0000 | 0.7684±0.0263 |
| 16 | 23.8±0.8367 | 28 | 0.8500±0.0299 | 0.7667±0.0391 | 0.6000±0.0639 | 0.9333±0.0261 | 0.7849±0.0410 |
| 17 | 19.2±0.4472 | 23 | 0.8348±0.0194 | 0.7373±0.0373 | 0.5333±0.0745 | 0.9412±0.0000 | 0.7597±0.0341 |
| 18 | 14.0±0.0000 | 16 | 0.8750±0.0000 | 0.8333±0.0000 | 0.7500±0.0000 | 0.9167±0.0000 | 0.8333±0.0000 |
| 19 | 9.0±0.0000 | 11 | 0.8182±0.0000 | 0.8036±0.0000 | 0.7500±0.0000 | 0.8571±0.0000 | 0.8036±0.0000 |
| 20 | 8.0±0.0000 | 10 | 0.8000±0.0000 | 0.7917±0.0000 | 0.7500±0.0000 | 0.8333±0.0000 | 0.7917±0.0000 |

Table 7: HM Random Forest baseline results of CV using base features, taking best model of grid using accuracy. Predictions from admission.

| Day | Corrects | Totals | Accuracy | AUC | Sensitivity | Specificity | F1 |
|---|---|---|---|---|---|---|---|
| 1 | 184.8±0.8367 | 194 | 0.9526±0.0043 | 0.8943±0.0090 | 0.8061±0.0166 | 0.9826±0.0028 | 0.9121±0.0082 |
| 2 | 183.2±0.4472 | 194 | 0.9443±0.0023 | 0.8870±0.0062 | 0.8000±0.0166 | 0.9739±0.0052 | 0.8985±0.0034 |
| 3 | 177.6±0.5477 | 187 | 0.9497±0.0029 | 0.8703±0.0091 | 0.7533±0.0183 | 0.9873±0.0000 | 0.8992±0.0067 |
| 4 | 170.2±1.4832 | 180 | 0.9456±0.0082 | 0.8617±0.0259 | 0.7379±0.0523 | 0.9854±0.0030 | 0.8905±0.0194 |
| 5 | 147.4±1.3416 | 160 | 0.9213±0.0084 | 0.8050±0.0256 | 0.6296±0.0524 | 0.9805±0.0041 | 0.8413±0.0210 |
| 6 | 129.0±0.7071 | 142 | 0.9085±0.0050 | 0.7903±0.0096 | 0.6080±0.0179 | 0.9726±0.0038 | 0.8232±0.0097 |
| 7 | 103.8±0.8367 | 117 | 0.8872±0.0072 | 0.7656±0.0156 | 0.5652±0.0307 | 0.9660±0.0048 | 0.7976±0.0146 |
| 8 | 88.6±1.5166 | 100 | 0.8860±0.0152 | 0.7524±0.0388 | 0.5368±0.0781 | 0.9679±0.0068 | 0.7858±0.0363 |
| 9 | 67.8±0.4472 | 79 | 0.8582±0.0057 | 0.7293±0.0118 | 0.5125±0.0280 | 0.9460±0.0087 | 0.7540±0.0100 |
| 10 | 53.2±0.4472 | 62 | 0.8581±0.0072 | 0.7576±0.0178 | 0.5846±0.0421 | 0.9306±0.0112 | 0.7724±0.0138 |
| 11 | 47.4±0.5477 | 54 | 0.8778±0.0101 | 0.7726±0.0065 | 0.5833±0.0000 | 0.9619±0.0130 | 0.8022±0.0125 |
| 12 | 42.0±0.7071 | 50 | 0.8400±0.0141 | 0.7237±0.0295 | 0.5000±0.0589 | 0.9474±0.0000 | 0.7493±0.0288 |
| 13 | 37.2±0.8367 | 44 | 0.8455±0.0190 | 0.6722±0.0296 | 0.4000±0.0559 | 0.9444±0.0196 | 0.6968±0.0324 |
| 14 | 29.8±0.4472 | 37 | 0.8054±0.0121 | 0.6767±0.0301 | 0.4500±0.0685 | 0.9034±0.0154 | 0.6886±0.0272 |
| 15 | 24.2±0.4472 | 31 | 0.7806±0.0144 | 0.6054±0.0093 | 0.2857±0.0000 | 0.9250±0.0186 | 0.6190±0.0130 |
| 16 | 21.0±0.7071 | 28 | 0.7500±0.0253 | 0.5857±0.0361 | 0.2571±0.0639 | 0.9143±0.0213 | 0.5920±0.0461 |
| 17 | 17.6±0.5477 | 23 | 0.7652±0.0238 | 0.5716±0.0161 | 0.1667±0.0000 | 0.9765±0.0322 | 0.5657±0.0179 |
| 18 | 12.4±0.5477 | 16 | 0.7750±0.0342 | 0.5500±0.0685 | 0.1000±0.1369 | 1.0000±0.0000 | 0.5149±0.1182 |
| 19 | 7.2±0.4472 | 11 | 0.6545±0.0407 | 0.5679±0.0319 | 0.2500±0.0000 | 0.8857±0.0639 | 0.5557±0.0313 |
| 20 | 6.2±0.4472 | 10 | 0.6200±0.0447 | 0.5583±0.0373 | 0.2500±0.0000 | 0.8667±0.0745 | 0.5390±0.0341 |

Table 8: HM Random Forest baseline results of CV using base features, taking best model of grid using accuracy. Predictions from outcome.

| Day | Corrects | Totals | Accuracy | AUC | Sensitivity | Specificity | F1 |
|---|---|---|---|---|---|---|---|
| 1 | 169.2±1.0954 | 194 | 0.8722±0.0056 | 0.6917±0.0207 | 0.4182±0.0449 | 0.9652±0.0056 | 0.7259±0.0191 |
| 2 | 170.8±0.8367 | 194 | 0.8804±0.0043 | 0.6943±0.0090 | 0.4121±0.0166 | 0.9764±0.0028 | 0.7354±0.0104 |
| 3 | 168.4±1.6733 | 187 | 0.9005±0.0089 | 0.7385±0.0127 | 0.5000±0.0236 | 0.9771±0.0097 | 0.7802±0.0165 |
| 4 | 168.2±1.3038 | 180 | 0.9344±0.0072 | 0.8161±0.0166 | 0.6414±0.0308 | 0.9907±0.0036 | 0.8605±0.0162 |
| 5 | 150.2±0.8367 | 160 | 0.9387±0.0052 | 0.8333±0.0155 | 0.6741±0.0310 | 0.9925±0.0000 | 0.8759±0.0125 |
| 6 | 133.4±0.8944 | 142 | 0.9394±0.0063 | 0.8532±0.0143 | 0.7200±0.0283 | 0.9863±0.0047 | 0.8856±0.0125 |
| 7 | 109.2±0.8367 | 117 | 0.9333±0.0072 | 0.8633±0.0182 | 0.7478±0.0364 | 0.9787±0.0000 | 0.8871±0.0139 |
| 8 | 92.2±1.0954 | 100 | 0.9220±0.0110 | 0.8431±0.0241 | 0.7158±0.0471 | 0.9704±0.0068 | 0.8647±0.0202 |
| 9 | 71.6±0.5477 | 79 | 0.9063±0.0069 | 0.8387±0.0171 | 0.7250±0.0342 | 0.9524±0.0000 | 0.8499±0.0132 |
| 10 | 56.2±0.8367 | 62 | 0.9065±0.0135 | 0.8391±0.0233 | 0.7231±0.0421 | 0.9551±0.0091 | 0.8528±0.0217 |
| 11 | 49.0±0.0000 | 54 | 0.9074±0.0000 | 0.8512±0.0000 | 0.7500±0.0000 | 0.9524±0.0000 | 0.8619±0.0000 |
| 12 | 44.2±0.8367 | 50 | 0.8840±0.0167 | 0.8268±0.0350 | 0.7167±0.0745 | 0.9368±0.0144 | 0.8356±0.0285 |
| 13 | 37.8±0.4472 | 44 | 0.8591±0.0102 | 0.7583±0.0280 | 0.6000±0.0559 | 0.9167±0.0000 | 0.7604±0.0233 |
| 14 | 31.4±0.5477 | 37 | 0.8486±0.0148 | 0.7948±0.0319 | 0.7000±0.0685 | 0.8897±0.0154 | 0.7839±0.0243 |
| 15 | 26.0±0.0000 | 31 | 0.8387±0.0000 | 0.7440±0.0000 | 0.5714±0.0000 | 0.9167±0.0000 | 0.7567±0.0000 |
| 16 | 24.0±0.0000 | 28 | 0.8571±0.0000 | 0.7619±0.0000 | 0.5714±0.0000 | 0.9524±0.0000 | 0.7879±0.0000 |
| 17 | 18.8±0.4472 | 23 | 0.8174±0.0194 | 0.7147±0.0132 | 0.5000±0.0000 | 0.9294±0.0263 | 0.7358±0.0193 |
| 18 | 13.8±0.4472 | 16 | 0.8625±0.0280 | 0.8250±0.0186 | 0.7500±0.0000 | 0.9000±0.0373 | 0.8203±0.0292 |
| 19 | 9.0±0.0000 | 11 | 0.8182±0.0000 | 0.8036±0.0000 | 0.7500±0.0000 | 0.8571±0.0000 | 0.8036±0.0000 |
| 20 | 8.0±0.0000 | 10 | 0.8000±0.0000 | 0.7917±0.0000 | 0.7500±0.0000 | 0.8333±0.0000 | 0.7917±0.0000 |

Table 9: HM Random Forest baseline results of CV using base features, taking best model of grid using F1. Predictions from admission.

| Day | Corrects | Totals | Accuracy | AUC | Sensitivity | Specificity | F1 |
|---|---|---|---|---|---|---|---|
| 1 | 185.2±0.4472 | 194 | 0.9546±0.0023 | 0.9004±0.0014 | 0.8182±0.0000 | 0.9826±0.0028 | 0.9164±0.0038 |
| 2 | 183.0±1.0000 | 194 | 0.9433±0.0052 | 0.8863±0.0095 | 0.8000±0.0166 | 0.9727±0.0034 | 0.8968±0.0094 |
| 3 | 178.0±0.7071 | 187 | 0.9519±0.0038 | 0.8824±0.0175 | 0.7800±0.0380 | 0.9847±0.0035 | 0.9050±0.0095 |
| 4 | 170.6±1.5166 | 180 | 0.9478±0.0084 | 0.8658±0.0261 | 0.7448±0.0523 | 0.9868±0.0000 | 0.8950±0.0199 |
| 5 | 147.6±0.8944 | 160 | 0.9225±0.0056 | 0.8087±0.0142 | 0.6370±0.0310 | 0.9805±0.0067 | 0.8447±0.0117 |
| 6 | 128.2±0.4472 | 142 | 0.9028±0.0031 | 0.7775±0.0096 | 0.5840±0.0219 | 0.9709±0.0047 | 0.8108±0.0072 |
| 7 | 103.8±0.4472 | 117 | 0.8872±0.0038 | 0.7656±0.0024 | 0.5652±0.0000 | 0.9660±0.0048 | 0.7978±0.0050 |
| 8 | 89.0±2.0000 | 100 | 0.8900±0.0200 | 0.7589±0.0486 | 0.5474±0.0956 | 0.9704±0.0068 | 0.7927±0.0464 |
| 9 | 68.6±0.8944 | 79 | 0.8684±0.0113 | 0.7449±0.0180 | 0.5375±0.0342 | 0.9524±0.0112 | 0.7717±0.0193 |
| 10 | 53.2±0.4472 | 62 | 0.8581±0.0072 | 0.7746±0.0172 | 0.6308±0.0344 | 0.9184±0.0000 | 0.7807±0.0138 |
| 11 | 47.2±0.4472 | 54 | 0.8741±0.0083 | 0.7583±0.0191 | 0.5500±0.0456 | 0.9667±0.0130 | 0.7910±0.0162 |
| 12 | 42.4±0.5477 | 50 | 0.8480±0.0110 | 0.7289±0.0254 | 0.5000±0.0589 | 0.9579±0.0144 | 0.7581±0.0232 |
| 13 | 36.8±1.3038 | 44 | 0.8364±0.0296 | 0.6667±0.0393 | 0.4000±0.0559 | 0.9333±0.0248 | 0.6878±0.0495 |
| 14 | 29.6±0.5477 | 37 | 0.8000±0.0148 | 0.6642±0.0319 | 0.4250±0.0685 | 0.9034±0.0154 | 0.6767±0.0299 |
| 15 | 24.2±0.4472 | 31 | 0.7806±0.0144 | 0.6054±0.0093 | 0.2857±0.0000 | 0.9250±0.0186 | 0.6190±0.0130 |
| 16 | 20.8±0.4472 | 28 | 0.7429±0.0160 | 0.5810±0.0319 | 0.2571±0.0639 | 0.9048±0.0000 | 0.5858±0.0407 |
| 17 | 17.8±0.4472 | 23 | 0.7739±0.0194 | 0.5775±0.0132 | 0.1667±0.0000 | 0.9882±0.0263 | 0.5722±0.0146 |
| 18 | 12.6±0.5477 | 16 | 0.7875±0.0342 | 0.5750±0.0685 | 0.1500±0.1369 | 1.0000±0.0000 | 0.5581±0.1182 |
| 19 | 7.0±0.0000 | 11 | 0.6364±0.0000 | 0.5536±0.0000 | 0.2500±0.0000 | 0.8571±0.0000 | 0.5417±0.0000 |
| 20 | 5.8±0.8367 | 10 | 0.5800±0.0837 | 0.5250±0.0697 | 0.2500±0.0000 | 0.8000±0.1394 | 0.5097±0.0623 |

Table 10: HM Random Forest baseline results of CV using base features, taking best model of grid using F1. Predictions from outcome.

| Day | Corrects | Totals | Accuracy | AUC | Sensitivity | Specificity | F1 |
|---|---|---|---|---|---|---|---|
| 1 | 170.0±1.4142 | 194 | 0.8763±0.0073 | 0.6918±0.0109 | 0.4121±0.0166 | 0.9714±0.0056 | 0.7301±0.0143 |
| 2 | 170.8±0.4472 | 194 | 0.8804±0.0023 | 0.6846±0.0109 | 0.3879±0.0254 | 0.9814±0.0044 | 0.7279±0.0099 |
| 3 | 167.8±0.8367 | 187 | 0.8973±0.0045 | 0.7097±0.0027 | 0.4333±0.0000 | 0.9860±0.0053 | 0.7585±0.0067 |
| 4 | 163.8±0.8367 | 180 | 0.9100±0.0046 | 0.7597±0.0075 | 0.5379±0.0189 | 0.9815±0.0073 | 0.8032±0.0074 |
| 5 | 146.8±0.8367 | 160 | 0.9175±0.0052 | 0.7792±0.0093 | 0.5704±0.0203 | 0.9880±0.0067 | 0.8261±0.0097 |
| 6 | 128.4±1.1402 | 142 | 0.9042±0.0080 | 0.7752±0.0221 | 0.5760±0.0456 | 0.9744±0.0060 | 0.8111±0.0188 |
| 7 | 106.2±0.8367 | 117 | 0.9077±0.0072 | 0.7981±0.0182 | 0.6174±0.0364 | 0.9787±0.0000 | 0.8343±0.0157 |
| 8 | 91.8±0.4472 | 100 | 0.9180±0.0045 | 0.8245±0.0118 | 0.6737±0.0235 | 0.9753±0.0000 | 0.8539±0.0096 |
| 9 | 73.4±0.8944 | 79 | 0.9291±0.0113 | 0.8810±0.0170 | 0.8000±0.0280 | 0.9619±0.0087 | 0.8882±0.0177 |
| 10 | 56.8±1.0954 | 62 | 0.9161±0.0177 | 0.8622±0.0309 | 0.7692±0.0544 | 0.9551±0.0091 | 0.8703±0.0282 |
| 11 | 50.2±0.4472 | 54 | 0.9296±0.0083 | 0.8714±0.0186 | 0.7667±0.0373 | 0.9762±0.0000 | 0.8921±0.0140 |
| 12 | 46.4±1.1402 | 50 | 0.9280±0.0228 | 0.8785±0.0475 | 0.7833±0.0950 | 0.9737±0.0000 | 0.8953±0.0363 |
| 13 | 41.0±1.0000 | 44 | 0.9318±0.0227 | 0.8611±0.0625 | 0.7500±0.1250 | 0.9722±0.0000 | 0.8774±0.0468 |
| 14 | 33.4±0.5477 | 37 | 0.9027±0.0148 | 0.8655±0.0342 | 0.8000±0.0685 | 0.9310±0.0000 | 0.8585±0.0246 |
| 15 | 26.4±0.5477 | 31 | 0.8516±0.0177 | 0.7726±0.0391 | 0.6286±0.0782 | 0.9167±0.0000 | 0.7802±0.0322 |
| 16 | 24.8±0.4472 | 28 | 0.8857±0.0160 | 0.8476±0.0319 | 0.7714±0.0782 | 0.9238±0.0261 | 0.8472±0.0221 |
| 17 | 20.0±0.0000 | 23 | 0.8696±0.0000 | 0.8039±0.0000 | 0.6667±0.0000 | 0.9412±0.0000 | 0.8208±0.0000 |
| 18 | 14.0±0.0000 | 16 | 0.8750±0.0000 | 0.8333±0.0000 | 0.7500±0.0000 | 0.9167±0.0000 | 0.8333±0.0000 |
| 19 | 9.0±0.0000 | 11 | 0.8182±0.0000 | 0.8036±0.0000 | 0.7500±0.0000 | 0.8571±0.0000 | 0.8036±0.0000 |
| 20 | 8.0±0.0000 | 10 | 0.8000±0.0000 | 0.7917±0.0000 | 0.7500±0.0000 | 0.8333±0.0000 | 0.7917±0.0000 |

Table 11: HM Random Forest with imputation, results of CV, taking best model of grid using accuracy. Predictions from admission.

| Day | Corrects | Totals | Accuracy | AUC | Sensitivity | Specificity | F1 |
|---|---|---|---|---|---|---|---|
| 1 | 183.8±1.4832 | 194 | 0.9474±0.0076 | 0.8768±0.0182 | 0.7697±0.0346 | 0.9839±0.0034 | 0.9007±0.0153 |
| 2 | 181.6±0.5477 | 194 | 0.9361±0.0028 | 0.8483±0.0083 | 0.7152±0.0166 | 0.9814±0.0000 | 0.8771±0.0064 |
| 3 | 173.6±0.8944 | 187 | 0.9283±0.0048 | 0.8198±0.0138 | 0.6600±0.0279 | 0.9796±0.0028 | 0.8526±0.0112 |
| 4 | 167.4±0.5477 | 180 | 0.9300±0.0030 | 0.8218±0.0133 | 0.6621±0.0289 | 0.9815±0.0030 | 0.8559±0.0086 |
| 5 | 146.8±0.8367 | 160 | 0.9175±0.0052 | 0.7910±0.0149 | 0.6000±0.0310 | 0.9820±0.0041 | 0.8311±0.0127 |
| 6 | 127.4±0.8944 | 142 | 0.8972±0.0063 | 0.7520±0.0092 | 0.5280±0.0179 | 0.9761±0.0072 | 0.7919±0.0108 |
| 7 | 103.6±1.1402 | 117 | 0.8855±0.0097 | 0.7415±0.0248 | 0.5043±0.0496 | 0.9787±0.0000 | 0.7825±0.0242 |
| 8 | 89.2±0.8367 | 100 | 0.8920±0.0084 | 0.7561±0.0220 | 0.5368±0.0440 | 0.9753±0.0000 | 0.7946±0.0205 |
| 9 | 70.4±0.5477 | 79 | 0.8911±0.0069 | 0.7825±0.0160 | 0.6000±0.0342 | 0.9651±0.0071 | 0.8121±0.0138 |
| 10 | 54.0±1.0000 | 62 | 0.8710±0.0161 | 0.7601±0.0251 | 0.5692±0.0421 | 0.9510±0.0112 | 0.7850±0.0269 |
| 11 | 48.0±0.7071 | 54 | 0.8889±0.0131 | 0.7917±0.0227 | 0.6167±0.0456 | 0.9667±0.0130 | 0.8212±0.0216 |
| 12 | 43.4±1.3416 | 50 | 0.8680±0.0268 | 0.7820±0.0418 | 0.6167±0.0745 | 0.9474±0.0186 | 0.8033±0.0425 |
| 13 | 39.0±0.0000 | 44 | 0.8864±0.0000 | 0.7847±0.0000 | 0.6250±0.0000 | 0.9444±0.0000 | 0.7991±0.0000 |
| 14 | 30.2±0.4472 | 37 | 0.8162±0.0121 | 0.7017±0.0077 | 0.5000±0.0000 | 0.9034±0.0154 | 0.7130±0.0127 |
| 15 | 24.6±0.5477 | 31 | 0.7935±0.0177 | 0.6339±0.0363 | 0.3429±0.0782 | 0.9250±0.0186 | 0.6497±0.0386 |
| 16 | 21.8±0.4472 | 28 | 0.7786±0.0160 | 0.6143±0.0106 | 0.2857±0.0000 | 0.9429±0.0213 | 0.6286±0.0137 |
| 17 | 18.0±0.0000 | 23 | 0.7826±0.0000 | 0.5833±0.0000 | 0.1667±0.0000 | 1.0000±0.0000 | 0.5788±0.0000 |
| 18 | 12.8±0.4472 | 16 | 0.8000±0.0280 | 0.6167±0.0186 | 0.2500±0.0000 | 0.9833±0.0373 | 0.6335±0.0245 |
| 19 | 7.0±0.0000 | 11 | 0.6364±0.0000 | 0.5536±0.0000 | 0.2500±0.0000 | 0.8571±0.0000 | 0.5417±0.0000 |
| 20 | 6.4±0.5477 | 10 | 0.6400±0.0548 | 0.5750±0.0456 | 0.2500±0.0000 | 0.9000±0.0913 | 0.5543±0.0417 |

Table 12: HM Random Forest with imputation, results of CV, taking best model of grid using accuracy. Predictions from outcome.

| Day | Corrects | Totals | Accuracy | AUC | Sensitivity | Specificity | F1 |
|---|---|---|---|---|---|---|---|
| 1 | 170.6±1.5166 | 194 | 0.8794±0.0078 | 0.6936±0.0153 | 0.4121±0.0271 | 0.9752±0.0044 | 0.7340±0.0184 |
| 2 | 170.8±0.8367 | 194 | 0.8804±0.0043 | 0.6919±0.0087 | 0.4061±0.0166 | 0.9776±0.0034 | 0.7336±0.0101 |
| 3 | 167.6±1.8166 | 187 | 0.8963±0.0097 | 0.7090±0.0209 | 0.4333±0.0408 | 0.9847±0.0085 | 0.7566±0.0233 |
| 4 | 164.0±1.0000 | 180 | 0.9111±0.0056 | 0.7576±0.0096 | 0.5310±0.0189 | 0.9841±0.0059 | 0.8035±0.0110 |
| 5 | 146.0±1.2247 | 160 | 0.9125±0.0077 | 0.7703±0.0214 | 0.5556±0.0454 | 0.9850±0.0075 | 0.8152±0.0192 |
| 6 | 129.0±1.5811 | 142 | 0.9085±0.0111 | 0.7809±0.0351 | 0.5840±0.0727 | 0.9778±0.0047 | 0.8181±0.0286 |
| 7 | 106.4±0.5477 | 117 | 0.9094±0.0047 | 0.7958±0.0137 | 0.6087±0.0307 | 0.9830±0.0058 | 0.8354±0.0104 |
| 8 | 91.8±0.4472 | 100 | 0.9180±0.0045 | 0.8245±0.0185 | 0.6737±0.0440 | 0.9753±0.0087 | 0.8538±0.0107 |
| 9 | 72.8±1.3038 | 79 | 0.9215±0.0165 | 0.8622±0.0280 | 0.7625±0.0523 | 0.9619±0.0142 | 0.8743±0.0267 |
| 10 | 56.4±0.8944 | 62 | 0.9097±0.0144 | 0.8524±0.0211 | 0.7538±0.0344 | 0.9510±0.0112 | 0.8606±0.0220 |
| 11 | 50.2±0.4472 | 54 | 0.9296±0.0083 | 0.8774±0.0200 | 0.7833±0.0456 | 0.9714±0.0106 | 0.8935±0.0136 |
| 12 | 45.8±1.0954 | 50 | 0.9160±0.0219 | 0.8592±0.0343 | 0.7500±0.0589 | 0.9684±0.0118 | 0.8782±0.0327 |
| 13 | 40.8±0.4472 | 44 | 0.9273±0.0102 | 0.8583±0.0395 | 0.7500±0.0884 | 0.9667±0.0124 | 0.8718±0.0238 |
| 14 | 33.4±0.8944 | 37 | 0.9027±0.0242 | 0.8655±0.0559 | 0.8000±0.1118 | 0.9310±0.0000 | 0.8576±0.0420 |
| 15 | 26.8±0.4472 | 31 | 0.8645±0.0144 | 0.8012±0.0319 | 0.6857±0.0639 | 0.9167±0.0000 | 0.8037±0.0263 |
| 16 | 24.4±0.8944 | 28 | 0.8714±0.0319 | 0.8286±0.0593 | 0.7429±0.1195 | 0.9143±0.0213 | 0.8271±0.0487 |
| 17 | 20.4±0.5477 | 23 | 0.8870±0.0238 | 0.8373±0.0456 | 0.7333±0.0913 | 0.9412±0.0000 | 0.8474±0.0364 |
| 18 | 14.0±0.0000 | 16 | 0.8750±0.0000 | 0.8333±0.0000 | 0.7500±0.0000 | 0.9167±0.0000 | 0.8333±0.0000 |
| 19 | 9.0±0.0000 | 11 | 0.8182±0.0000 | 0.8036±0.0000 | 0.7500±0.0000 | 0.8571±0.0000 | 0.8036±0.0000 |
| 20 | 8.0±0.0000 | 10 | 0.8000±0.0000 | 0.7917±0.0000 | 0.7500±0.0000 | 0.8333±0.0000 | 0.7917±0.0000 |

Table 13: HM Random Forest with imputation, results of CV, taking best model of grid using F1. Predictions from admission.

| Day | Corrects | Totals | Accuracy | AUC | Sensitivity | Specificity | F1 |
|---|---|---|---|---|---|---|---|
| 1 | 183.0±1.2247 | 194 | 0.9433±0.0063 | 0.8671±0.0138 | 0.7515±0.0254 | 0.9826±0.0028 | 0.8924±0.0126 |
| 2 | 181.4±1.5166 | 194 | 0.9351±0.0078 | 0.8428±0.0203 | 0.7030±0.0395 | 0.9826±0.0028 | 0.8739±0.0169 |
| 3 | 174.2±1.3038 | 187 | 0.9316±0.0070 | 0.8244±0.0205 | 0.6667±0.0408 | 0.9822±0.0028 | 0.8586±0.0170 |
| 4 | 167.8±1.3038 | 180 | 0.9322±0.0072 | 0.8259±0.0266 | 0.6690±0.0577 | 0.9828±0.0076 | 0.8602±0.0184 |
| 5 | 146.8±1.6432 | 160 | 0.9175±0.0103 | 0.7910±0.0247 | 0.6000±0.0483 | 0.9820±0.0067 | 0.8309±0.0231 |
| 6 | 127.6±2.0736 | 142 | 0.8986±0.0146 | 0.7560±0.0280 | 0.5360±0.0537 | 0.9761±0.0127 | 0.7953±0.0307 |
| 7 | 104.0±1.2247 | 117 | 0.8889±0.0105 | 0.7502±0.0221 | 0.5217±0.0435 | 0.9787±0.0075 | 0.7910±0.0223 |
| 8 | 89.0±1.4142 | 100 | 0.8900±0.0141 | 0.7589±0.0255 | 0.5474±0.0471 | 0.9704±0.0110 | 0.7942±0.0276 |
| 9 | 70.4±0.8944 | 79 | 0.8911±0.0113 | 0.7872±0.0140 | 0.6125±0.0280 | 0.9619±0.0142 | 0.8145±0.0170 |
| 10 | 54.2±0.8367 | 62 | 0.8742±0.0135 | 0.7678±0.0227 | 0.5846±0.0421 | 0.9510±0.0112 | 0.7917±0.0229 |
| 11 | 48.0±0.7071 | 54 | 0.8889±0.0131 | 0.7857±0.0323 | 0.6000±0.0697 | 0.9714±0.0106 | 0.8179±0.0275 |
| 12 | 43.0±1.2247 | 50 | 0.8600±0.0245 | 0.7596±0.0347 | 0.5667±0.0697 | 0.9526±0.0288 | 0.7857±0.0358 |
| 13 | 38.2±0.4472 | 44 | 0.8682±0.0102 | 0.7542±0.0297 | 0.5750±0.0685 | 0.9333±0.0152 | 0.7662±0.0224 |
| 14 | 30.4±0.5477 | 37 | 0.8216±0.0148 | 0.7052±0.0094 | 0.5000±0.0000 | 0.9103±0.0189 | 0.7187±0.0155 |
| 15 | 23.8±1.7889 | 31 | 0.7677±0.0577 | 0.6274±0.0637 | 0.3714±0.0782 | 0.8833±0.0543 | 0.6394±0.0742 |
| 16 | 21.6±1.1402 | 28 | 0.7714±0.0407 | 0.6095±0.0271 | 0.2857±0.0000 | 0.9333±0.0543 | 0.6237±0.0353 |
| 17 | 17.8±0.4472 | 23 | 0.7739±0.0194 | 0.5775±0.0132 | 0.1667±0.0000 | 0.9882±0.0263 | 0.5722±0.0146 |
| 18 | 12.8±0.4472 | 16 | 0.8000±0.0280 | 0.6167±0.0186 | 0.2500±0.0000 | 0.9833±0.0373 | 0.6335±0.0245 |
| 19 | 7.6±0.5477 | 11 | 0.6909±0.0498 | 0.5964±0.0391 | 0.2500±0.0000 | 0.9429±0.0782 | 0.5837±0.0384 |
| 20 | 6.6±0.5477 | 10 | 0.6600±0.0548 | 0.5917±0.0456 | 0.2500±0.0000 | 0.9333±0.0913 | 0.5695±0.0417 |

Table 14: HM Random Forest with imputation, results of CV, taking best model of grid using F1. Predictions from outcome.

| Day | Corrects | Totals | Accuracy | AUC | Sensitivity | Specificity | F1 |
|---|---|---|---|---|---|---|---|
| 1 | 169.0±2.2361 | 194 | 0.8711±0.0115 | 0.6766±0.0214 | 0.3818±0.0407 | 0.9714±0.0104 | 0.7138±0.0253 |
| 2 | 170.8±1.6432 | 194 | 0.8804±0.0085 | 0.6798±0.0192 | 0.3758±0.0407 | 0.9839±0.0094 | 0.7238±0.0216 |
| 3 | 166.8±1.6432 | 187 | 0.8920±0.0088 | 0.7092±0.0142 | 0.4400±0.0279 | 0.9783±0.0097 | 0.7524±0.0174 |
| 4 | 164.0±0.7071 | 180 | 0.9111±0.0039 | 0.7576±0.0080 | 0.5310±0.0189 | 0.9841±0.0059 | 0.8035±0.0073 |
| 5 | 145.6±1.3416 | 160 | 0.9100±0.0084 | 0.7629±0.0111 | 0.5407±0.0203 | 0.9850±0.0092 | 0.8089±0.0150 |
| 6 | 127.8±0.8367 | 142 | 0.9000±0.0059 | 0.7600±0.0205 | 0.5440±0.0456 | 0.9761±0.0072 | 0.7988±0.0165 |
| 7 | 105.8±0.8367 | 117 | 0.9043±0.0072 | 0.7894±0.0182 | 0.6000±0.0364 | 0.9787±0.0000 | 0.8268±0.0158 |
| 8 | 90.6±0.8944 | 100 | 0.9060±0.0089 | 0.8010±0.0250 | 0.6316±0.0526 | 0.9704±0.0068 | 0.8306±0.0198 |
| 9 | 72.2±1.0954 | 79 | 0.9139±0.0139 | 0.8621±0.0197 | 0.7750±0.0342 | 0.9492±0.0133 | 0.8656±0.0208 |
| 10 | 56.2±1.0954 | 62 | 0.9065±0.0177 | 0.8447±0.0354 | 0.7385±0.0688 | 0.9510±0.0112 | 0.8543±0.0291 |
| 11 | 49.6±1.1402 | 54 | 0.9185±0.0211 | 0.8643±0.0375 | 0.7667±0.0697 | 0.9619±0.0130 | 0.8773±0.0335 |
| 12 | 45.4±1.5166 | 50 | 0.9080±0.0303 | 0.8539±0.0541 | 0.7500±0.1021 | 0.9579±0.0144 | 0.8674±0.0482 |
| 13 | 40.6±1.1402 | 44 | 0.9227±0.0259 | 0.8653±0.0553 | 0.7750±0.1046 | 0.9556±0.0152 | 0.8680±0.0472 |
| 14 | 33.2±0.8367 | 37 | 0.8973±0.0226 | 0.8440±0.0449 | 0.7500±0.0884 | 0.9379±0.0154 | 0.8464±0.0370 |
| 15 | 26.6±0.8944 | 31 | 0.8581±0.0289 | 0.7768±0.0454 | 0.6286±0.0782 | 0.9250±0.0186 | 0.7879±0.0449 |
| 16 | 24.2±0.8367 | 28 | 0.8643±0.0299 | 0.8143±0.0516 | 0.7143±0.1010 | 0.9143±0.0213 | 0.8163±0.0444 |
| 17 | 20.2±0.4472 | 23 | 0.8783±0.0194 | 0.8206±0.0373 | 0.7000±0.0745 | 0.9412±0.0000 | 0.8341±0.0297 |
| 18 | 14.2±0.4472 | 16 | 0.8875±0.0280 | 0.8583±0.0559 | 0.8000±0.1118 | 0.9167±0.0000 | 0.8512±0.0400 |
| 19 | 9.2±0.4472 | 11 | 0.8364±0.0407 | 0.8286±0.0559 | 0.8000±0.1118 | 0.8571±0.0000 | 0.8241±0.0458 |
| 20 | 8.2±0.4472 | 10 | 0.8200±0.0447 | 0.8167±0.0559 | 0.8000±0.1118 | 0.8333±0.0000 | 0.8131±0.0480 |

Table 15: HM Random Forest with imputation, with missing flags, results of CV, taking best model of grid using accuracy. Predictions from admission.

| Day | Corrects | Totals | Accuracy | AUC | Sensitivity | Specificity | F1 |
|---|---|---|---|---|---|---|---|
| 1 | 183.4±1.5166 | 194 | 0.9454±0.0078 | 0.8731±0.0203 | 0.7636±0.0395 | 0.9826±0.0028 | 0.8967±0.0164 |
| 2 | 182.4±2.1909 | 194 | 0.9402±0.0113 | 0.8580±0.0299 | 0.7333±0.0583 | 0.9826±0.0028 | 0.8852±0.0247 |
| 3 | 174.0±1.2247 | 187 | 0.9305±0.0065 | 0.8238±0.0220 | 0.6667±0.0471 | 0.9809±0.0064 | 0.8568±0.0157 |
| 4 | 166.0±1.5811 | 180 | 0.9222±0.0088 | 0.8060±0.0166 | 0.6345±0.0308 | 0.9775±0.0076 | 0.8396±0.0173 |
| 5 | 145.8±1.9235 | 160 | 0.9113±0.0120 | 0.7813±0.0311 | 0.5852±0.0609 | 0.9774±0.0053 | 0.8185±0.0283 |
| 6 | 126.0±1.0000 | 142 | 0.8873±0.0070 | 0.7335±0.0168 | 0.4960±0.0358 | 0.9709±0.0076 | 0.7708±0.0162 |
| 7 | 103.0±1.2247 | 117 | 0.8803±0.0105 | 0.7318±0.0201 | 0.4870±0.0364 | 0.9766±0.0048 | 0.7721±0.0221 |
| 8 | 88.6±2.0736 | 100 | 0.8860±0.0207 | 0.7483±0.0474 | 0.5263±0.0912 | 0.9704±0.0068 | 0.7833±0.0447 |
| 9 | 69.6±1.3416 | 79 | 0.8810±0.0170 | 0.7762±0.0334 | 0.6000±0.0713 | 0.9524±0.0194 | 0.7988±0.0299 |
| 10 | 53.8±0.4472 | 62 | 0.8677±0.0072 | 0.7581±0.0178 | 0.5692±0.0421 | 0.9469±0.0112 | 0.7809±0.0138 |
| 11 | 47.0±0.7071 | 54 | 0.8704±0.0131 | 0.7500±0.0227 | 0.5333±0.0456 | 0.9667±0.0130 | 0.7833±0.0228 |
| 12 | 42.6±0.5477 | 50 | 0.8520±0.0110 | 0.7487±0.0305 | 0.5500±0.0745 | 0.9474±0.0186 | 0.7728±0.0233 |
| 13 | 37.6±1.8166 | 44 | 0.8545±0.0413 | 0.7167±0.0732 | 0.5000±0.1250 | 0.9333±0.0248 | 0.7337±0.0784 |
| 14 | 30.4±0.8944 | 37 | 0.8216±0.0242 | 0.6961±0.0346 | 0.4750±0.0559 | 0.9172±0.0189 | 0.7124±0.0383 |
| 15 | 24.6±0.5477 | 31 | 0.7935±0.0177 | 0.6137±0.0114 | 0.2857±0.0000 | 0.9417±0.0228 | 0.6307±0.0159 |
| 16 | 21.8±0.8367 | 28 | 0.7786±0.0299 | 0.5952±0.0445 | 0.2286±0.0782 | 0.9619±0.0213 | 0.6024±0.0616 |
| 17 | 17.4±0.5477 | 23 | 0.7565±0.0238 | 0.5657±0.0161 | 0.1667±0.0000 | 0.9647±0.0322 | 0.5591±0.0179 |
| 18 | 12.4±0.5477 | 16 | 0.7750±0.0342 | 0.6000±0.0228 | 0.2500±0.0000 | 0.9500±0.0456 | 0.6116±0.0300 |
| 19 | 7.2±0.8367 | 11 | 0.6545±0.0761 | 0.5893±0.0866 | 0.3500±0.1369 | 0.8286±0.0639 | 0.5862±0.0947 |
| 20 | 6.6±0.5477 | 10 | 0.6600±0.0548 | 0.6000±0.0559 | 0.3000±0.1118 | 0.9000±0.0913 | 0.5836±0.0617 |

Table 16: HM Random Forest with imputation, with missing flags, results of CV, taking best model of grid using accuracy. Predictions from outcome.

| Day | Corrects | Totals | Accuracy | AUC | Sensitivity | Specificity | F1 |
|---|---|---|---|---|---|---|---|
| 1 | 169.0±2.2361 | 194 | 0.8711±0.0115 | 0.6766±0.0214 | 0.3818±0.0407 | 0.9714±0.0104 | 0.7138±0.0253 |
| 2 | 170.8±1.6432 | 194 | 0.8804±0.0085 | 0.6798±0.0192 | 0.3758±0.0407 | 0.9839±0.0094 | 0.7238±0.0216 |
| 3 | 166.8±1.6432 | 187 | 0.8920±0.0088 | 0.7092±0.0142 | 0.4400±0.0279 | 0.9783±0.0097 | 0.7524±0.0174 |
| 4 | 164.0±0.7071 | 180 | 0.9111±0.0039 | 0.7576±0.0080 | 0.5310±0.0189 | 0.9841±0.0059 | 0.8035±0.0073 |
| 5 | 145.6±1.3416 | 160 | 0.9100±0.0084 | 0.7629±0.0111 | 0.5407±0.0203 | 0.9850±0.0092 | 0.8089±0.0150 |
| 6 | 127.8±0.8367 | 142 | 0.9000±0.0059 | 0.7600±0.0205 | 0.5440±0.0456 | 0.9761±0.0072 | 0.7988±0.0165 |
| 7 | 105.8±0.8367 | 117 | 0.9043±0.0072 | 0.7894±0.0182 | 0.6000±0.0364 | 0.9787±0.0000 | 0.8268±0.0158 |
| 8 | 90.6±0.8944 | 100 | 0.9060±0.0089 | 0.8010±0.0250 | 0.6316±0.0526 | 0.9704±0.0068 | 0.8306±0.0198 |
| 9 | 72.2±1.0954 | 79 | 0.9139±0.0139 | 0.8621±0.0197 | 0.7750±0.0342 | 0.9492±0.0133 | 0.8656±0.0208 |
| 10 | 56.2±1.0954 | 62 | 0.9065±0.0177 | 0.8447±0.0354 | 0.7385±0.0688 | 0.9510±0.0112 | 0.8543±0.0291 |
| 11 | 49.6±1.1402 | 54 | 0.9185±0.0211 | 0.8643±0.0375 | 0.7667±0.0697 | 0.9619±0.0130 | 0.8773±0.0335 |
| 12 | 45.4±1.5166 | 50 | 0.9080±0.0303 | 0.8539±0.0541 | 0.7500±0.1021 | 0.9579±0.0144 | 0.8674±0.0482 |
| 13 | 40.6±1.1402 | 44 | 0.9227±0.0259 | 0.8653±0.0553 | 0.7750±0.1046 | 0.9556±0.0152 | 0.8680±0.0472 |
| 14 | 33.2±0.8367 | 37 | 0.8973±0.0226 | 0.8440±0.0449 | 0.7500±0.0884 | 0.9379±0.0154 | 0.8464±0.0370 |
| 15 | 26.6±0.8944 | 31 | 0.8581±0.0289 | 0.7768±0.0454 | 0.6286±0.0782 | 0.9250±0.0186 | 0.7879±0.0449 |
| 16 | 24.2±0.8367 | 28 | 0.8643±0.0299 | 0.8143±0.0516 | 0.7143±0.1010 | 0.9143±0.0213 | 0.8163±0.0444 |
| 17 | 20.2±0.4472 | 23 | 0.8783±0.0194 | 0.8206±0.0373 | 0.7000±0.0745 | 0.9412±0.0000 | 0.8341±0.0297 |
| 18 | 14.2±0.4472 | 16 | 0.8875±0.0280 | 0.8583±0.0559 | 0.8000±0.1118 | 0.9167±0.0000 | 0.8512±0.0400 |
| 19 | 9.2±0.4472 | 11 | 0.8364±0.0407 | 0.8286±0.0559 | 0.8000±0.1118 | 0.8571±0.0000 | 0.8241±0.0458 |
| 20 | 8.2±0.4472 | 10 | 0.8200±0.0447 | 0.8167±0.0559 | 0.8000±0.1118 | 0.8333±0.0000 | 0.8131±0.0480 |

Table 17: HM Random Forest with imputation, with missing flags, results of CV, taking best model of grid using F1. Predictions from admission.

| Day | Corrects | Totals | Accuracy | AUC | Sensitivity | Specificity | F1 |
|---|---|---|---|---|---|---|---|
| 1 | 183.4±1.5166 | 194 | 0.9454±0.0078 | 0.8731±0.0203 | 0.7636±0.0395 | 0.9826±0.0028 | 0.8967±0.0164 |
| 2 | 182.4±2.1909 | 194 | 0.9402±0.0113 | 0.8580±0.0299 | 0.7333±0.0583 | 0.9826±0.0028 | 0.8852±0.0247 |
| 3 | 174.0±1.2247 | 187 | 0.9305±0.0065 | 0.8238±0.0220 | 0.6667±0.0471 | 0.9809±0.0064 | 0.8568±0.0157 |
| 4 | 166.0±1.5811 | 180 | 0.9222±0.0088 | 0.8060±0.0166 | 0.6345±0.0308 | 0.9775±0.0076 | 0.8396±0.0173 |
| 5 | 145.8±1.9235 | 160 | 0.9113±0.0120 | 0.7813±0.0311 | 0.5852±0.0609 | 0.9774±0.0053 | 0.8185±0.0283 |
| 6 | 126.0±1.0000 | 142 | 0.8873±0.0070 | 0.7335±0.0168 | 0.4960±0.0358 | 0.9709±0.0076 | 0.7708±0.0162 |
| 7 | 103.0±1.2247 | 117 | 0.8803±0.0105 | 0.7318±0.0201 | 0.4870±0.0364 | 0.9766±0.0048 | 0.7721±0.0221 |
| 8 | 88.6±2.0736 | 100 | 0.8860±0.0207 | 0.7483±0.0474 | 0.5263±0.0912 | 0.9704±0.0068 | 0.7833±0.0447 |
| 9 | 69.6±1.3416 | 79 | 0.8810±0.0170 | 0.7762±0.0334 | 0.6000±0.0713 | 0.9524±0.0194 | 0.7988±0.0299 |
| 10 | 53.8±0.4472 | 62 | 0.8677±0.0072 | 0.7581±0.0178 | 0.5692±0.0421 | 0.9469±0.0112 | 0.7809±0.0138 |
| 11 | 47.0±0.7071 | 54 | 0.8704±0.0131 | 0.7500±0.0227 | 0.5333±0.0456 | 0.9667±0.0130 | 0.7833±0.0228 |
| 12 | 42.6±0.5477 | 50 | 0.8520±0.0110 | 0.7487±0.0305 | 0.5500±0.0745 | 0.9474±0.0186 | 0.7728±0.0233 |
| 13 | 37.6±1.8166 | 44 | 0.8545±0.0413 | 0.7167±0.0732 | 0.5000±0.1250 | 0.9333±0.0248 | 0.7337±0.0784 |
| 14 | 30.4±0.8944 | 37 | 0.8216±0.0242 | 0.6961±0.0346 | 0.4750±0.0559 | 0.9172±0.0189 | 0.7124±0.0383 |
| 15 | 24.6±0.5477 | 31 | 0.7935±0.0177 | 0.6137±0.0114 | 0.2857±0.0000 | 0.9417±0.0228 | 0.6307±0.0159 |
| 16 | 21.8±0.8367 | 28 | 0.7786±0.0299 | 0.5952±0.0445 | 0.2286±0.0782 | 0.9619±0.0213 | 0.6024±0.0616 |
| 17 | 17.4±0.5477 | 23 | 0.7565±0.0238 | 0.5657±0.0161 | 0.1667±0.0000 | 0.9647±0.0322 | 0.5591±0.0179 |
| 18 | 12.4±0.5477 | 16 | 0.7750±0.0342 | 0.6000±0.0228 | 0.2500±0.0000 | 0.9500±0.0456 | 0.6116±0.0300 |
| 19 | 7.2±0.8367 | 11 | 0.6545±0.0761 | 0.5893±0.0866 | 0.3500±0.1369 | 0.8286±0.0639 | 0.5862±0.0947 |
| 20 | 6.6±0.5477 | 10 | 0.6600±0.0548 | 0.6000±0.0559 | 0.3000±0.1118 | 0.9000±0.0913 | 0.5836±0.0617 |

Table 18: HM Random Forest with imputation, with missing flags, results of CV, taking best model of grid using F1. Predictions from outcome.

| Day | Corrects | Totals | Accuracy | AUC | Sensitivity | Specificity | F1 |
|---|---|---|---|---|---|---|---|
| 1 | 171.4±1.6733 | 194 | 0.8835±0.0086 | 0.6961±0.0153 | 0.4121±0.0271 | 0.9801±0.0068 | 0.7397±0.0186 |
| 2 | 170.4±2.3022 | 194 | 0.8784±0.0119 | 0.6906±0.0168 | 0.4061±0.0271 | 0.9752±0.0108 | 0.7311±0.0230 |
| 3 | 167.8±0.4472 | 187 | 0.8973±0.0024 | 0.7258±0.0065 | 0.4733±0.0149 | 0.9783±0.0035 | 0.7689±0.0056 |
| 4 | 163.4±1.1402 | 180 | 0.9078±0.0063 | 0.7528±0.0095 | 0.5241±0.0154 | 0.9815±0.0055 | 0.7969±0.0125 |
| 5 | 145.0±1.8708 | 160 | 0.9062±0.0117 | 0.7695±0.0192 | 0.5630±0.0310 | 0.9759±0.0082 | 0.8076±0.0230 |
| 6 | 130.4±1.6733 | 142 | 0.9183±0.0118 | 0.8026±0.0363 | 0.6240±0.0780 | 0.9812±0.0111 | 0.8395±0.0277 |
| 7 | 108.6±1.5166 | 117 | 0.9282±0.0130 | 0.8568±0.0294 | 0.7391±0.0615 | 0.9745±0.0121 | 0.8787±0.0238 |
| 8 | 92.6±0.8944 | 100 | 0.9260±0.0089 | 0.8536±0.0189 | 0.7368±0.0372 | 0.9704±0.0068 | 0.8729±0.0161 |
| 9 | 73.4±0.5477 | 79 | 0.9291±0.0069 | 0.8716±0.0171 | 0.7750±0.0342 | 0.9683±0.0000 | 0.8858±0.0127 |
| 10 | 55.6±0.5477 | 62 | 0.8968±0.0088 | 0.8386±0.0211 | 0.7385±0.0421 | 0.9388±0.0000 | 0.8423±0.0161 |
| 11 | 49.2±0.8367 | 54 | 0.9111±0.0155 | 0.8595±0.0349 | 0.7667±0.0697 | 0.9524±0.0000 | 0.8677±0.0267 |
| 12 | 45.0±0.0000 | 50 | 0.9000±0.0000 | 0.8487±0.0000 | 0.7500±0.0000 | 0.9474±0.0000 | 0.8588±0.0000 |
| 13 | 39.0±0.0000 | 44 | 0.8864±0.0000 | 0.7847±0.0000 | 0.6250±0.0000 | 0.9444±0.0000 | 0.7991±0.0000 |
| 14 | 32.8±0.8367 | 37 | 0.8865±0.0226 | 0.8009±0.0330 | 0.6500±0.0559 | 0.9517±0.0189 | 0.8208±0.0342 |
| 15 | 26.2±0.4472 | 31 | 0.8452±0.0144 | 0.7583±0.0319 | 0.6000±0.0639 | 0.9167±0.0000 | 0.7684±0.0263 |
| 16 | 24.6±0.5477 | 28 | 0.8786±0.0196 | 0.8048±0.0391 | 0.6571±0.0782 | 0.9524±0.0000 | 0.8250±0.0339 |
| 17 | 19.2±0.4472 | 23 | 0.8348±0.0194 | 0.7373±0.0373 | 0.5333±0.0745 | 0.9412±0.0000 | 0.7597±0.0341 |
| 18 | 13.8±0.4472 | 16 | 0.8625±0.0280 | 0.8083±0.0559 | 0.7000±0.1118 | 0.9167±0.0000 | 0.8118±0.0481 |
| 19 | 9.0±0.0000 | 11 | 0.8182±0.0000 | 0.8036±0.0000 | 0.7500±0.0000 | 0.8571±0.0000 | 0.8036±0.0000 |
| 20 | 8.0±0.0000 | 10 | 0.8000±0.0000 | 0.7917±0.0000 | 0.7500±0.0000 | 0.8333±0.0000 | 0.7917±0.0000 |

Table 19: HM Random Forest with imputation, with missing flags, applying reference values, results of CV, taking best model of grid using accuracy. Predictions from admission.

| Day | Corrects | Totals | Accuracy | AUC | Sensitivity | Specificity | F1 |
|---|---|---|---|---|---|---|---|
| 1 | 186.8±1.0954 | 194 | 0.9629±0.0056 | 0.9150±0.0166 | 0.8424±0.0332 | 0.9876±0.0000 | 0.9315±0.0115 |
| 2 | 184.0±1.4142 | 194 | 0.9485±0.0073 | 0.8822±0.0175 | 0.7818±0.0332 | 0.9826±0.0028 | 0.9034±0.0147 |
| 3 | 177.0±1.5811 | 187 | 0.9465±0.0085 | 0.8576±0.0193 | 0.7267±0.0365 | 0.9885±0.0053 | 0.8910±0.0180 |
| 4 | 166.6±1.3416 | 180 | 0.9256±0.0075 | 0.8108±0.0165 | 0.6414±0.0308 | 0.9801±0.0047 | 0.8459±0.0160 |
| 5 | 146.8±1.3038 | 160 | 0.9175±0.0081 | 0.7910±0.0238 | 0.6000±0.0483 | 0.9820±0.0041 | 0.8308±0.0201 |
| 6 | 126.2±0.4472 | 142 | 0.8887±0.0031 | 0.7438±0.0019 | 0.5200±0.0000 | 0.9675±0.0038 | 0.7784±0.0044 |
| 7 | 102.2±1.3038 | 117 | 0.8735±0.0111 | 0.7472±0.0216 | 0.5391±0.0389 | 0.9553±0.0048 | 0.7749±0.0221 |
| 8 | 87.2±1.7889 | 100 | 0.8720±0.0179 | 0.7558±0.0270 | 0.5684±0.0440 | 0.9432±0.0141 | 0.7754±0.0299 |
| 9 | 68.6±1.8166 | 79 | 0.8684±0.0230 | 0.7683±0.0335 | 0.6000±0.0559 | 0.9365±0.0194 | 0.7839±0.0355 |
| 10 | 54.4±1.8166 | 62 | 0.8774±0.0293 | 0.7755±0.0540 | 0.6000±0.1003 | 0.9510±0.0183 | 0.7974±0.0523 |
| 11 | 47.2±0.4472 | 54 | 0.8741±0.0083 | 0.7583±0.0314 | 0.5500±0.0745 | 0.9667±0.0130 | 0.7902±0.0227 |
| 12 | 40.6±0.8944 | 50 | 0.8120±0.0179 | 0.6939±0.0246 | 0.4667±0.0456 | 0.9211±0.0186 | 0.7125±0.0271 |
| 13 | 37.2±0.4472 | 44 | 0.8455±0.0102 | 0.6625±0.0062 | 0.3750±0.0000 | 0.9500±0.0124 | 0.6894±0.0118 |
| 14 | 30.2±0.4472 | 37 | 0.8162±0.0121 | 0.6655±0.0280 | 0.4000±0.0559 | 0.9310±0.0000 | 0.6859±0.0279 |
| 15 | 24.4±0.5477 | 31 | 0.7871±0.0177 | 0.6095±0.0114 | 0.2857±0.0000 | 0.9333±0.0228 | 0.6248±0.0159 |
| 16 | 21.4±0.5477 | 28 | 0.7643±0.0196 | 0.5762±0.0391 | 0.2000±0.0782 | 0.9524±0.0000 | 0.5759±0.0538 |
| 17 | 17.8±0.4472 | 23 | 0.7739±0.0194 | 0.5667±0.0373 | 0.1333±0.0745 | 1.0000±0.0000 | 0.5480±0.0688 |
| 18 | 12.8±0.4472 | 16 | 0.8000±0.0280 | 0.6167±0.0186 | 0.2500±0.0000 | 0.9833±0.0373 | 0.6335±0.0245 |
| 19 | 7.6±0.5477 | 11 | 0.6909±0.0498 | 0.5964±0.0391 | 0.2500±0.0000 | 0.9429±0.0782 | 0.5837±0.0384 |
| 20 | 6.4±0.5477 | 10 | 0.6400±0.0548 | 0.5750±0.0456 | 0.2500±0.0000 | 0.9000±0.0913 | 0.5543±0.0417 |

Table 20: HM Random Forest with imputation, with missing flags, applying reference values, results of CV, taking best model of grid using accuracy. Predictions from outcome.

| Day | Corrects | Totals | Accuracy | AUC | Sensitivity | Specificity | F1 |
|---|---|---|---|---|---|---|---|
| 1 | 170.8±1.4832 | 194 | 0.8804±0.0076 | 0.6919±0.0171 | 0.4061±0.0346 | 0.9776±0.0071 | 0.7334±0.0186 |
| 2 | 171.2±1.7889 | 194 | 0.8825±0.0092 | 0.7003±0.0164 | 0.4242±0.0303 | 0.9764±0.0081 | 0.7417±0.0195 |
| 3 | 167.6±0.5477 | 187 | 0.8963±0.0029 | 0.7252±0.0191 | 0.4733±0.0435 | 0.9771±0.0057 | 0.7669±0.0145 |
| 4 | 163.4±1.6733 | 180 | 0.9078±0.0093 | 0.7528±0.0114 | 0.5241±0.0154 | 0.9815±0.0086 | 0.7971±0.0174 |
| 5 | 145.2±1.7889 | 160 | 0.9075±0.0112 | 0.7761±0.0231 | 0.5778±0.0422 | 0.9744±0.0067 | 0.8120±0.0240 |
| 6 | 130.8±1.0954 | 142 | 0.9211±0.0077 | 0.8137±0.0272 | 0.6480±0.0593 | 0.9795±0.0076 | 0.8477±0.0193 |
| 7 | 109.6±1.1402 | 117 | 0.9368±0.0097 | 0.8720±0.0198 | 0.7652±0.0389 | 0.9787±0.0075 | 0.8937±0.0174 |
| 8 | 92.4±1.3416 | 100 | 0.9240±0.0134 | 0.8564±0.0240 | 0.7474±0.0440 | 0.9654±0.0103 | 0.8712±0.0230 |
| 9 | 73.0±0.7071 | 79 | 0.9241±0.0090 | 0.8685±0.0169 | 0.7750±0.0342 | 0.9619±0.0087 | 0.8790±0.0144 |
| 10 | 56.2±0.4472 | 62 | 0.9065±0.0072 | 0.8617±0.0172 | 0.7846±0.0344 | 0.9388±0.0000 | 0.8595±0.0124 |
| 11 | 49.8±0.4472 | 54 | 0.9222±0.0083 | 0.8845±0.0186 | 0.8167±0.0373 | 0.9524±0.0000 | 0.8867±0.0138 |
| 12 | 45.0±0.0000 | 50 | 0.9000±0.0000 | 0.8487±0.0000 | 0.7500±0.0000 | 0.9474±0.0000 | 0.8588±0.0000 |
| 13 | 39.0±0.0000 | 44 | 0.8864±0.0000 | 0.7847±0.0000 | 0.6250±0.0000 | 0.9444±0.0000 | 0.7991±0.0000 |
| 14 | 32.2±0.4472 | 37 | 0.8703±0.0121 | 0.7815±0.0077 | 0.6250±0.0000 | 0.9379±0.0154 | 0.7975±0.0147 |
| 15 | 26.2±0.4472 | 31 | 0.8452±0.0144 | 0.7583±0.0319 | 0.6000±0.0639 | 0.9167±0.0000 | 0.7684±0.0263 |
| 16 | 24.4±0.5477 | 28 | 0.8714±0.0196 | 0.7905±0.0391 | 0.6286±0.0782 | 0.9524±0.0000 | 0.8126±0.0339 |
| 17 | 19.4±0.5477 | 23 | 0.8435±0.0238 | 0.7539±0.0456 | 0.5667±0.0913 | 0.9412±0.0000 | 0.7750±0.0418 |
| 18 | 14.0±0.0000 | 16 | 0.8750±0.0000 | 0.8333±0.0000 | 0.7500±0.0000 | 0.9167±0.0000 | 0.8333±0.0000 |
| 19 | 9.0±0.0000 | 11 | 0.8182±0.0000 | 0.8036±0.0000 | 0.7500±0.0000 | 0.8571±0.0000 | 0.8036±0.0000 |
| 20 | 8.0±0.0000 | 10 | 0.8000±0.0000 | 0.7917±0.0000 | 0.7500±0.0000 | 0.8333±0.0000 | 0.7917±0.0000 |

Table 21: HM Random Forest with imputation, with missing flags, applying reference values, results of CV, taking best model of grid using F1. Predictions from admission.

| Day | Corrects | Totals | Accuracy | AUC | Sensitivity | Specificity | F1 |
|---|---|---|---|---|---|---|---|
| 1 | 188.4±0.8944 | 194 | 0.9711±0.0046 | 0.9392±0.0136 | 0.8909±0.0271 | 0.9876±0.0000 | 0.9478±0.0090 |
| 2 | 184.0±0.7071 | 194 | 0.9485±0.0036 | 0.8870±0.0058 | 0.7939±0.0136 | 0.9801±0.0052 | 0.9045±0.0060 |
| 3 | 176.4±1.3416 | 187 | 0.9433±0.0072 | 0.8611±0.0142 | 0.7400±0.0279 | 0.9822±0.0070 | 0.8870±0.0140 |
| 4 | 167.6±1.5166 | 180 | 0.9311±0.0084 | 0.8252±0.0208 | 0.6690±0.0393 | 0.9815±0.0030 | 0.8586±0.0189 |
| 5 | 146.0±1.2247 | 160 | 0.9125±0.0077 | 0.7762±0.0210 | 0.5704±0.0422 | 0.9820±0.0041 | 0.8180±0.0189 |
| 6 | 125.8±0.8367 | 142 | 0.8859±0.0059 | 0.7389±0.0094 | 0.5120±0.0179 | 0.9658±0.0060 | 0.7728±0.0108 |
| 7 | 102.6±0.8944 | 117 | 0.8769±0.0076 | 0.7526±0.0125 | 0.5478±0.0238 | 0.9574±0.0075 | 0.7811±0.0131 |
| 8 | 87.4±1.5166 | 100 | 0.8740±0.0152 | 0.7571±0.0160 | 0.5684±0.0235 | 0.9457±0.0166 | 0.7781±0.0219 |
| 9 | 67.8±0.4472 | 79 | 0.8582±0.0057 | 0.7526±0.0118 | 0.5750±0.0280 | 0.9302±0.0087 | 0.7671±0.0093 |
| 10 | 54.8±0.8367 | 62 | 0.8839±0.0135 | 0.7965±0.0227 | 0.6462±0.0421 | 0.9469±0.0112 | 0.8139±0.0220 |
| 11 | 48.0±1.2247 | 54 | 0.8889±0.0227 | 0.7917±0.0404 | 0.6167±0.0745 | 0.9667±0.0130 | 0.8207±0.0402 |
| 12 | 41.2±1.3038 | 50 | 0.8240±0.0261 | 0.7018±0.0486 | 0.4667±0.0950 | 0.9368±0.0144 | 0.7233±0.0505 |
| 13 | 37.0±0.0000 | 44 | 0.8409±0.0000 | 0.6597±0.0000 | 0.3750±0.0000 | 0.9444±0.0000 | 0.6841±0.0000 |
| 14 | 30.2±0.4472 | 37 | 0.8162±0.0121 | 0.6655±0.0280 | 0.4000±0.0559 | 0.9310±0.0000 | 0.6859±0.0279 |
| 15 | 24.4±0.5477 | 31 | 0.7871±0.0177 | 0.6095±0.0114 | 0.2857±0.0000 | 0.9333±0.0228 | 0.6248±0.0159 |
| 16 | 21.8±0.4472 | 28 | 0.7786±0.0160 | 0.6048±0.0319 | 0.2571±0.0639 | 0.9524±0.0000 | 0.6152±0.0439 |
| 17 | 17.4±0.5477 | 23 | 0.7565±0.0238 | 0.5333±0.0456 | 0.0667±0.0913 | 1.0000±0.0000 | 0.4865±0.0842 |
| 18 | 12.8±0.4472 | 16 | 0.8000±0.0280 | 0.6167±0.0186 | 0.2500±0.0000 | 0.9833±0.0373 | 0.6335±0.0245 |
| 19 | 7.0±0.7071 | 11 | 0.6364±0.0643 | 0.5536±0.0505 | 0.2500±0.0000 | 0.8571±0.1010 | 0.5426±0.0479 |
| 20 | 6.6±0.5477 | 10 | 0.6600±0.0548 | 0.5917±0.0456 | 0.2500±0.0000 | 0.9333±0.0913 | 0.5695±0.0417 |

Table 22: HM Random Forest with imputation, with missing flags, applying reference values, results of CV, taking best model of grid using F1. Predictions from outcome.

| Day | Corrects | Totals | Accuracy | AUC | Sensitivity | Specificity | F1 |
|---|---|---|---|---|---|---|---|
| 1 | 168.2±1.3038 | 194 | 0.8670±0.0067 | 0.6814±0.0137 | 0.4000±0.0254 | 0.9627±0.0044 | 0.7144±0.0156 |
| 2 | 170.0±0.0000 | 194 | 0.8763±0.0000 | 0.6797±0.0066 | 0.3818±0.0166 | 0.9776±0.0034 | 0.7206±0.0054 |
| 3 | 168.4±2.6077 | 187 | 0.9005±0.0139 | 0.7331±0.0212 | 0.4867±0.0380 | 0.9796±0.0138 | 0.7772±0.0274 |
| 4 | 168.0±0.7071 | 180 | 0.9333±0.0039 | 0.8154±0.0080 | 0.6414±0.0189 | 0.9894±0.0059 | 0.8588±0.0071 |
| 5 | 148.8±1.4832 | 160 | 0.9300±0.0093 | 0.8133±0.0266 | 0.6370±0.0549 | 0.9895±0.0067 | 0.8563±0.0222 |
| 6 | 133.6±1.1402 | 142 | 0.9408±0.0080 | 0.8477±0.0228 | 0.7040±0.0456 | 0.9915±0.0000 | 0.8859±0.0180 |
| 7 | 108.4±0.5477 | 117 | 0.9265±0.0047 | 0.8492±0.0111 | 0.7217±0.0238 | 0.9766±0.0048 | 0.8747±0.0087 |
| 8 | 91.0±1.2247 | 100 | 0.9100±0.0122 | 0.8196±0.0243 | 0.6737±0.0440 | 0.9654±0.0055 | 0.8425±0.0229 |
| 9 | 72.2±1.3038 | 79 | 0.9139±0.0165 | 0.8435±0.0222 | 0.7250±0.0342 | 0.9619±0.0142 | 0.8603±0.0258 |
| 10 | 56.4±0.8944 | 62 | 0.9097±0.0144 | 0.8524±0.0211 | 0.7538±0.0344 | 0.9510±0.0112 | 0.8606±0.0220 |
| 11 | 49.2±0.4472 | 54 | 0.9111±0.0083 | 0.8536±0.0053 | 0.7500±0.0000 | 0.9571±0.0106 | 0.8667±0.0107 |
| 12 | 44.8±0.8367 | 50 | 0.8960±0.0167 | 0.8461±0.0110 | 0.7500±0.0000 | 0.9421±0.0220 | 0.8544±0.0200 |
| 13 | 38.6±0.5477 | 44 | 0.8773±0.0124 | 0.7792±0.0076 | 0.6250±0.0000 | 0.9333±0.0152 | 0.7878±0.0155 |
| 14 | 32.6±0.5477 | 37 | 0.8811±0.0148 | 0.8246±0.0271 | 0.7250±0.0559 | 0.9241±0.0154 | 0.8243±0.0231 |
| 15 | 26.0±0.0000 | 31 | 0.8387±0.0000 | 0.7440±0.0000 | 0.5714±0.0000 | 0.9167±0.0000 | 0.7567±0.0000 |
| 16 | 23.4±0.5477 | 28 | 0.8357±0.0196 | 0.7571±0.0310 | 0.6000±0.0639 | 0.9143±0.0213 | 0.7692±0.0280 |
| 17 | 19.0±0.0000 | 23 | 0.8261±0.0000 | 0.7206±0.0000 | 0.5000±0.0000 | 0.9412±0.0000 | 0.7444±0.0000 |
| 18 | 14.0±0.0000 | 16 | 0.8750±0.0000 | 0.8333±0.0000 | 0.7500±0.0000 | 0.9167±0.0000 | 0.8333±0.0000 |
| 19 | 9.0±0.0000 | 11 | 0.8182±0.0000 | 0.8036±0.0000 | 0.7500±0.0000 | 0.8571±0.0000 | 0.8036±0.0000 |
| 20 | 8.0±0.0000 | 10 | 0.8000±0.0000 | 0.7917±0.0000 | 0.7500±0.0000 | 0.8333±0.0000 | 0.7917±0.0000 |

Table 23: HM Random Forest with missing flags, results of CV, taking best model of grid using accuracy. Predictions from admission.

| Day | Corrects | Totals | Accuracy | AUC | Sensitivity | Specificity | F1 |
|---|---|---|---|---|---|---|---|
| 1 | 184.8±1.4832 | 194 | 0.9526±0.0076 | 0.8943±0.0097 | 0.8061±0.0166 | 0.9826±0.0081 | 0.9123±0.0132 |
| 2 | 183.8±0.8367 | 194 | 0.9474±0.0043 | 0.8888±0.0155 | 0.8000±0.0346 | 0.9776±0.0056 | 0.9032±0.0089 |
| 3 | 177.4±1.1402 | 187 | 0.9487±0.0061 | 0.8724±0.0146 | 0.7600±0.0279 | 0.9847±0.0035 | 0.8979±0.0127 |
| 4 | 169.6±0.5477 | 180 | 0.9422±0.0030 | 0.8541±0.0100 | 0.7241±0.0244 | 0.9841±0.0059 | 0.8838±0.0058 |
| 5 | 148.0±1.2247 | 160 | 0.9250±0.0077 | 0.8073±0.0221 | 0.6296±0.0454 | 0.9850±0.0053 | 0.8474±0.0178 |
| 6 | 128.0±1.2247 | 142 | 0.9014±0.0086 | 0.7798±0.0088 | 0.5920±0.0179 | 0.9675±0.0111 | 0.8105±0.0125 |
| 7 | 104.0±1.0000 | 117 | 0.8889±0.0085 | 0.7667±0.0158 | 0.5652±0.0307 | 0.9681±0.0075 | 0.7999±0.0159 |
| 8 | 89.4±1.3416 | 100 | 0.8940±0.0134 | 0.7694±0.0340 | 0.5684±0.0686 | 0.9704±0.0068 | 0.8030±0.0308 |
| 9 | 68.0±0.7071 | 79 | 0.8608±0.0090 | 0.7262±0.0056 | 0.5000±0.0000 | 0.9524±0.0112 | 0.7545±0.0107 |
| 10 | 53.2±0.8367 | 62 | 0.8581±0.0135 | 0.7633±0.0201 | 0.6000±0.0344 | 0.9265±0.0112 | 0.7755±0.0211 |
| 11 | 47.2±1.3038 | 54 | 0.8741±0.0241 | 0.7583±0.0416 | 0.5500±0.0745 | 0.9667±0.0130 | 0.7907±0.0448 |
| 12 | 41.8±0.8367 | 50 | 0.8360±0.0167 | 0.7096±0.0355 | 0.4667±0.0745 | 0.9526±0.0118 | 0.7366±0.0342 |
| 13 | 37.8±0.8367 | 44 | 0.8591±0.0190 | 0.6806±0.0318 | 0.4000±0.0559 | 0.9611±0.0152 | 0.7127±0.0369 |
| 14 | 29.4±0.5477 | 37 | 0.7946±0.0148 | 0.6517±0.0271 | 0.4000±0.0559 | 0.9034±0.0154 | 0.6647±0.0263 |
| 15 | 24.0±0.0000 | 31 | 0.7742±0.0000 | 0.6012±0.0000 | 0.2857±0.0000 | 0.9167±0.0000 | 0.6132±0.0000 |
| 16 | 20.6±0.5477 | 28 | 0.7357±0.0196 | 0.5667±0.0391 | 0.2286±0.0782 | 0.9048±0.0000 | 0.5676±0.0498 |
| 17 | 17.8±0.4472 | 23 | 0.7739±0.0194 | 0.5775±0.0132 | 0.1667±0.0000 | 0.9882±0.0263 | 0.5722±0.0146 |
| 18 | 11.6±0.5477 | 16 | 0.7250±0.0342 | 0.5000±0.0510 | 0.0500±0.1118 | 0.9500±0.0456 | 0.4523±0.0775 |
| 19 | 7.0±0.0000 | 11 | 0.6364±0.0000 | 0.5536±0.0000 | 0.2500±0.0000 | 0.8571±0.0000 | 0.5417±0.0000 |
| 20 | 6.2±0.4472 | 10 | 0.6200±0.0447 | 0.5583±0.0373 | 0.2500±0.0000 | 0.8667±0.0745 | 0.5390±0.0341 |

Table 24: HM Random Forest with missing flags, results of CV, taking best model of grid using accuracy. Predictions from outcome.

| Day | Corrects | Totals | Accuracy | AUC | Sensitivity | Specificity | F1 |
|---|---|---|---|---|---|---|---|
| 1 | 168.0±1.2247 | 194 | 0.8660±0.0063 | 0.6783±0.0120 | 0.3939±0.0214 | 0.9627±0.0044 | 0.7113±0.0139 |
| 2 | 169.4±0.8944 | 194 | 0.8732±0.0046 | 0.6682±0.0124 | 0.3576±0.0254 | 0.9789±0.0034 | 0.7084±0.0139 |
| 3 | 168.2±2.1679 | 187 | 0.8995±0.0116 | 0.7298±0.0268 | 0.4800±0.0558 | 0.9796±0.0123 | 0.7733±0.0271 |
| 4 | 166.8±1.4832 | 180 | 0.9267±0.0082 | 0.7975±0.0201 | 0.6069±0.0393 | 0.9881±0.0055 | 0.8423±0.0192 |
| 5 | 148.4±2.0736 | 160 | 0.9275±0.0130 | 0.8088±0.0385 | 0.6296±0.0786 | 0.9880±0.0067 | 0.8506±0.0322 |
| 6 | 132.4±1.5166 | 142 | 0.9324±0.0107 | 0.8300±0.0235 | 0.6720±0.0438 | 0.9880±0.0047 | 0.8687±0.0221 |
| 7 | 108.8±0.8367 | 117 | 0.9299±0.0072 | 0.8546±0.0182 | 0.7304±0.0364 | 0.9787±0.0000 | 0.8804±0.0140 |
| 8 | 90.4±1.3416 | 100 | 0.9040±0.0134 | 0.8159±0.0250 | 0.6737±0.0440 | 0.9580±0.0068 | 0.8344±0.0242 |
| 9 | 71.6±1.1402 | 79 | 0.9063±0.0144 | 0.8294±0.0279 | 0.7000±0.0523 | 0.9587±0.0087 | 0.8467±0.0253 |
| 10 | 56.0±0.7071 | 62 | 0.9032±0.0114 | 0.8427±0.0209 | 0.7385±0.0421 | 0.9469±0.0112 | 0.8505±0.0181 |
| 11 | 49.2±1.0954 | 54 | 0.9111±0.0203 | 0.8536±0.0338 | 0.7500±0.0589 | 0.9571±0.0106 | 0.8663±0.0318 |
| 12 | 44.8±0.8367 | 50 | 0.8960±0.0167 | 0.8461±0.0110 | 0.7500±0.0000 | 0.9421±0.0220 | 0.8544±0.0200 |
| 13 | 38.6±0.8944 | 44 | 0.8773±0.0203 | 0.7792±0.0124 | 0.6250±0.0000 | 0.9333±0.0248 | 0.7883±0.0264 |
| 14 | 32.4±0.5477 | 37 | 0.8757±0.0148 | 0.8211±0.0256 | 0.7250±0.0559 | 0.9172±0.0189 | 0.8180±0.0218 |
| 15 | 25.8±0.4472 | 31 | 0.8323±0.0144 | 0.7399±0.0093 | 0.5714±0.0000 | 0.9083±0.0186 | 0.7500±0.0150 |
| 16 | 23.6±0.5477 | 28 | 0.8429±0.0196 | 0.7619±0.0292 | 0.6000±0.0639 | 0.9238±0.0261 | 0.7769±0.0265 |
| 17 | 19.2±0.4472 | 23 | 0.8348±0.0194 | 0.7373±0.0373 | 0.5333±0.0745 | 0.9412±0.0000 | 0.7597±0.0341 |
| 18 | 14.0±0.0000 | 16 | 0.8750±0.0000 | 0.8333±0.0000 | 0.7500±0.0000 | 0.9167±0.0000 | 0.8333±0.0000 |
| 19 | 9.0±0.0000 | 11 | 0.8182±0.0000 | 0.8036±0.0000 | 0.7500±0.0000 | 0.8571±0.0000 | 0.8036±0.0000 |
| 20 | 8.0±0.0000 | 10 | 0.8000±0.0000 | 0.7917±0.0000 | 0.7500±0.0000 | 0.8333±0.0000 | 0.7917±0.0000 |

Table 25: HM Random Forest with missing flags, results of CV, taking best model of grid using F1. Predictions from admission.

| Day | Corrects | Totals | Accuracy | AUC | Sensitivity | Specificity | F1 |
|---|---|---|---|---|---|---|---|
| 1 | 184.4±1.5166 | 194 | 0.9505±0.0078 | 0.8883±0.0100 | 0.7939±0.0136 | 0.9826±0.0068 | 0.9079±0.0139 |
| 2 | 183.4±0.8944 | 194 | 0.9454±0.0046 | 0.8828±0.0099 | 0.7879±0.0214 | 0.9776±0.0056 | 0.8990±0.0082 |
| 3 | 177.0±0.7071 | 187 | 0.9465±0.0038 | 0.8657±0.0139 | 0.7467±0.0298 | 0.9847±0.0035 | 0.8930±0.0090 |
| 4 | 169.6±0.5477 | 180 | 0.9422±0.0030 | 0.8541±0.0100 | 0.7241±0.0244 | 0.9841±0.0059 | 0.8838±0.0058 |
| 5 | 147.4±1.1402 | 160 | 0.9213±0.0071 | 0.7962±0.0120 | 0.6074±0.0203 | 0.9850±0.0053 | 0.8383±0.0142 |
| 6 | 128.0±0.7071 | 142 | 0.9014±0.0050 | 0.7829±0.0030 | 0.6000±0.0000 | 0.9658±0.0060 | 0.8118±0.0070 |
| 7 | 102.6±0.5477 | 117 | 0.8769±0.0047 | 0.7461±0.0089 | 0.5304±0.0194 | 0.9617±0.0058 | 0.7775±0.0084 |
| 8 | 88.8±0.8367 | 100 | 0.8880±0.0084 | 0.7576±0.0224 | 0.5474±0.0471 | 0.9679±0.0068 | 0.7913±0.0188 |
| 9 | 67.8±1.3038 | 79 | 0.8582±0.0165 | 0.7199±0.0290 | 0.4875±0.0523 | 0.9524±0.0112 | 0.7481±0.0308 |
| 10 | 53.0±1.0000 | 62 | 0.8548±0.0161 | 0.7556±0.0251 | 0.5846±0.0421 | 0.9265±0.0112 | 0.7689±0.0260 |
| 11 | 46.8±0.4472 | 54 | 0.8667±0.0083 | 0.7476±0.0191 | 0.5333±0.0456 | 0.9619±0.0130 | 0.7787±0.0159 |
| 12 | 42.2±0.8367 | 50 | 0.8440±0.0167 | 0.7206±0.0332 | 0.4833±0.0697 | 0.9579±0.0144 | 0.7496±0.0327 |
| 13 | 37.2±0.8367 | 44 | 0.8455±0.0190 | 0.6625±0.0116 | 0.3750±0.0000 | 0.9500±0.0232 | 0.6899±0.0212 |
| 14 | 29.4±0.5477 | 37 | 0.7946±0.0148 | 0.6517±0.0271 | 0.4000±0.0559 | 0.9034±0.0154 | 0.6647±0.0263 |
| 15 | 24.0±0.0000 | 31 | 0.7742±0.0000 | 0.6012±0.0000 | 0.2857±0.0000 | 0.9167±0.0000 | 0.6132±0.0000 |
| 16 | 20.6±0.5477 | 28 | 0.7357±0.0196 | 0.5667±0.0391 | 0.2286±0.0782 | 0.9048±0.0000 | 0.5676±0.0498 |
| 17 | 17.8±0.4472 | 23 | 0.7739±0.0194 | 0.5775±0.0132 | 0.1667±0.0000 | 0.9882±0.0263 | 0.5722±0.0146 |
| 18 | 11.4±0.5477 | 16 | 0.7125±0.0342 | 0.4917±0.0543 | 0.0500±0.1118 | 0.9333±0.0373 | 0.4481±0.0797 |
| 19 | 7.0±0.0000 | 11 | 0.6364±0.0000 | 0.5536±0.0000 | 0.2500±0.0000 | 0.8571±0.0000 | 0.5417±0.0000 |
| 20 | 6.2±0.4472 | 10 | 0.6200±0.0447 | 0.5583±0.0373 | 0.2500±0.0000 | 0.8667±0.0745 | 0.5390±0.0341 |

Table 26: HM Random Forest with missing flags, results of CV, taking best model of grid using F1. Predictions from outcome.

| Day | Corrects | Totals | Accuracy | AUC | Sensitivity | Specificity | F1 |
|---|---|---|---|---|---|---|---|
| 1 | 168.8±1.7889 | 194 | 0.8701±0.0092 | 0.6880±0.0237 | 0.4121±0.0460 | 0.9640±0.0028 | 0.7216±0.0253 |
| 2 | 171.4±1.5166 | 194 | 0.8835±0.0078 | 0.7034±0.0145 | 0.4303±0.0254 | 0.9764±0.0052 | 0.7449±0.0174 |
| 3 | 168.4±1.3416 | 187 | 0.9005±0.0072 | 0.7412±0.0082 | 0.5067±0.0149 | 0.9758±0.0083 | 0.7817±0.0119 |
| 4 | 168.4±1.1402 | 180 | 0.9356±0.0063 | 0.8167±0.0107 | 0.6414±0.0189 | 0.9921±0.0055 | 0.8625±0.0129 |
| 5 | 149.2±1.4832 | 160 | 0.9325±0.0093 | 0.8295±0.0275 | 0.6741±0.0549 | 0.9850±0.0000 | 0.8653±0.0222 |
| 6 | 133.4±1.3416 | 142 | 0.9394±0.0094 | 0.8532±0.0239 | 0.7200±0.0490 | 0.9863±0.0076 | 0.8854±0.0195 |
| 7 | 109.8±0.4472 | 117 | 0.9385±0.0038 | 0.8829±0.0157 | 0.7913±0.0364 | 0.9745±0.0058 | 0.8984±0.0083 |
| 8 | 91.6±1.1402 | 100 | 0.9160±0.0114 | 0.8313±0.0148 | 0.6947±0.0235 | 0.9679±0.0110 | 0.8540±0.0180 |
| 9 | 71.4±0.5477 | 79 | 0.9038±0.0069 | 0.8138±0.0249 | 0.6625±0.0559 | 0.9651±0.0071 | 0.8382±0.0164 |
| 10 | 56.2±1.3038 | 62 | 0.9065±0.0210 | 0.8447±0.0276 | 0.7385±0.0421 | 0.9510±0.0183 | 0.8549±0.0306 |
| 11 | 48.8±0.4472 | 54 | 0.9037±0.0083 | 0.8369±0.0319 | 0.7167±0.0745 | 0.9571±0.0106 | 0.8527±0.0206 |
| 12 | 43.4±0.5477 | 50 | 0.8680±0.0110 | 0.7991±0.0072 | 0.6667±0.0000 | 0.9316±0.0144 | 0.8115±0.0125 |
| 13 | 38.4±0.5477 | 44 | 0.8727±0.0124 | 0.7764±0.0076 | 0.6250±0.0000 | 0.9278±0.0152 | 0.7821±0.0155 |
| 14 | 32.2±0.8367 | 37 | 0.8703±0.0226 | 0.7996±0.0380 | 0.6750±0.0685 | 0.9241±0.0154 | 0.8047±0.0349 |
| 15 | 26.0±0.0000 | 31 | 0.8387±0.0000 | 0.7440±0.0000 | 0.5714±0.0000 | 0.9167±0.0000 | 0.7567±0.0000 |
| 16 | 23.6±0.5477 | 28 | 0.8429±0.0196 | 0.7524±0.0130 | 0.5714±0.0000 | 0.9333±0.0261 | 0.7725±0.0210 |
| 17 | 18.8±0.4472 | 23 | 0.8174±0.0194 | 0.7147±0.0132 | 0.5000±0.0000 | 0.9294±0.0263 | 0.7358±0.0193 |
| 18 | 14.0±0.0000 | 16 | 0.8750±0.0000 | 0.8333±0.0000 | 0.7500±0.0000 | 0.9167±0.0000 | 0.8333±0.0000 |
| 19 | 9.0±0.0000 | 11 | 0.8182±0.0000 | 0.8036±0.0000 | 0.7500±0.0000 | 0.8571±0.0000 | 0.8036±0.0000 |
| 20 | 8.0±0.0000 | 10 | 0.8000±0.0000 | 0.7917±0.0000 | 0.7500±0.0000 | 0.8333±0.0000 | 0.7917±0.0000 |

Table 27: HM Random Forest applying laboratory reference values, results of CV, taking best model of grid using accuracy. Predictions from admission.

| Day | Corrects | Totals | Accuracy | AUC | Sensitivity | Specificity | F1 |
|---|---|---|---|---|---|---|---|
| 1 | 185.0±1.5811 | 194 | 0.9536±0.0082 | 0.8950±0.0111 | 0.8061±0.0166 | 0.9839±0.0071 | 0.9139±0.0145 |
| 2 | 184.0±0.7071 | 194 | 0.9485±0.0036 | 0.9039±0.0082 | 0.8364±0.0166 | 0.9714±0.0034 | 0.9078±0.0066 |
| 3 | 177.8±0.8367 | 187 | 0.9508±0.0045 | 0.8844±0.0098 | 0.7867±0.0183 | 0.9822±0.0028 | 0.9039±0.0090 |
| 4 | 169.0±0.7071 | 180 | 0.9389±0.0039 | 0.8493±0.0137 | 0.7172±0.0289 | 0.9815±0.0030 | 0.8774±0.0093 |
| 5 | 148.2±0.4472 | 160 | 0.9263±0.0028 | 0.8140±0.0092 | 0.6444±0.0203 | 0.9835±0.0034 | 0.8518±0.0066 |
| 6 | 128.0±0.7071 | 142 | 0.9014±0.0050 | 0.7672±0.0030 | 0.5600±0.0000 | 0.9744±0.0060 | 0.8045±0.0072 |
| 7 | 103.0±1.4142 | 117 | 0.8803±0.0121 | 0.7613±0.0269 | 0.5652±0.0532 | 0.9574±0.0075 | 0.7885±0.0243 |
| 8 | 89.6±1.5166 | 100 | 0.8960±0.0152 | 0.7827±0.0322 | 0.6000±0.0600 | 0.9654±0.0055 | 0.8117±0.0307 |
| 9 | 68.2±1.0954 | 79 | 0.8633±0.0139 | 0.7371±0.0290 | 0.5250±0.0559 | 0.9492±0.0071 | 0.7625±0.0273 |
| 10 | 53.6±0.8944 | 62 | 0.8645±0.0144 | 0.7730±0.0278 | 0.6154±0.0544 | 0.9306±0.0112 | 0.7854±0.0250 |
| 11 | 47.8±0.4472 | 54 | 0.8852±0.0083 | 0.7774±0.0053 | 0.5833±0.0000 | 0.9714±0.0106 | 0.8113±0.0102 |
| 12 | 41.4±0.5477 | 50 | 0.8280±0.0110 | 0.6987±0.0228 | 0.4500±0.0456 | 0.9474±0.0000 | 0.7246±0.0232 |
| 13 | 36.6±1.1402 | 44 | 0.8318±0.0259 | 0.6542±0.0158 | 0.3750±0.0000 | 0.9333±0.0317 | 0.6755±0.0273 |
| 14 | 29.4±0.5477 | 37 | 0.7946±0.0148 | 0.6427±0.0094 | 0.3750±0.0000 | 0.9103±0.0189 | 0.6579±0.0141 |
| 15 | 24.2±0.4472 | 31 | 0.7806±0.0144 | 0.6054±0.0093 | 0.2857±0.0000 | 0.9250±0.0186 | 0.6190±0.0130 |
| 16 | 22.0±0.0000 | 28 | 0.7857±0.0000 | 0.6190±0.0000 | 0.2857±0.0000 | 0.9524±0.0000 | 0.6348±0.0000 |
| 17 | 17.6±0.8944 | 23 | 0.7652±0.0389 | 0.5608±0.0504 | 0.1333±0.0745 | 0.9882±0.0263 | 0.5451±0.0754 |
| 18 | 12.4±0.5477 | 16 | 0.7750±0.0342 | 0.5500±0.0685 | 0.1000±0.1369 | 1.0000±0.0000 | 0.5149±0.1182 |
| 19 | 7.4±0.5477 | 11 | 0.6727±0.0498 | 0.5821±0.0391 | 0.2500±0.0000 | 0.9143±0.0782 | 0.5697±0.0384 |
| 20 | 5.6±0.5477 | 10 | 0.5600±0.0548 | 0.5083±0.0456 | 0.2500±0.0000 | 0.7667±0.0913 | 0.4945±0.0401 |

Table 28: HM Random Forest applying laboratory reference values, results of CV, taking best model of grid using accuracy. Predictions from outcome.

| Day | Corrects | Totals | Accuracy | AUC | Sensitivity | Specificity | F1 |
|---|---|---|---|---|---|---|---|
| 1 | 168.4±1.5166 | 194 | 0.8680±0.0078 | 0.6868±0.0265 | 0.4121±0.0550 | 0.9615±0.0028 | 0.7186±0.0262 |
| 2 | 170.8±1.6432 | 194 | 0.8804±0.0085 | 0.7015±0.0150 | 0.4303±0.0254 | 0.9727±0.0056 | 0.7407±0.0181 |
| 3 | 168.4±0.8944 | 187 | 0.9005±0.0048 | 0.7466±0.0131 | 0.5200±0.0298 | 0.9732±0.0070 | 0.7844±0.0106 |
| 4 | 168.0±0.7071 | 180 | 0.9333±0.0039 | 0.8098±0.0128 | 0.6276±0.0289 | 0.9921±0.0055 | 0.8567±0.0092 |
| 5 | 149.4±0.8944 | 160 | 0.9337±0.0056 | 0.8332±0.0166 | 0.6815±0.0331 | 0.9850±0.0000 | 0.8686±0.0133 |
| 6 | 133.2±1.6432 | 142 | 0.9380±0.0116 | 0.8555±0.0196 | 0.7280±0.0335 | 0.9829±0.0085 | 0.8843±0.0213 |
| 7 | 109.4±0.8944 | 117 | 0.9350±0.0076 | 0.8709±0.0223 | 0.7652±0.0496 | 0.9766±0.0089 | 0.8911±0.0147 |
| 8 | 91.2±1.0954 | 100 | 0.9120±0.0110 | 0.8288±0.0155 | 0.6947±0.0235 | 0.9630±0.0087 | 0.8484±0.0180 |
| 9 | 71.4±0.5477 | 79 | 0.9038±0.0069 | 0.8138±0.0249 | 0.6625±0.0559 | 0.9651±0.0071 | 0.8382±0.0164 |
| 10 | 56.6±0.8944 | 62 | 0.9129±0.0144 | 0.8545±0.0218 | 0.7538±0.0344 | 0.9551±0.0091 | 0.8647±0.0224 |
| 11 | 49.0±0.7071 | 54 | 0.9074±0.0131 | 0.8512±0.0295 | 0.7500±0.0589 | 0.9524±0.0000 | 0.8615±0.0227 |
| 12 | 43.4±0.5477 | 50 | 0.8680±0.0110 | 0.7991±0.0072 | 0.6667±0.0000 | 0.9316±0.0144 | 0.8115±0.0125 |
| 13 | 38.6±0.5477 | 44 | 0.8773±0.0124 | 0.7792±0.0076 | 0.6250±0.0000 | 0.9333±0.0152 | 0.7878±0.0155 |
| 14 | 32.0±0.7071 | 37 | 0.8649±0.0191 | 0.7961±0.0340 | 0.6750±0.0685 | 0.9172±0.0189 | 0.7985±0.0292 |
| 15 | 26.0±0.0000 | 31 | 0.8387±0.0000 | 0.7440±0.0000 | 0.5714±0.0000 | 0.9167±0.0000 | 0.7567±0.0000 |
| 16 | 23.4±0.5477 | 28 | 0.8357±0.0196 | 0.7476±0.0130 | 0.5714±0.0000 | 0.9238±0.0261 | 0.7649±0.0210 |
| 17 | 19.0±0.0000 | 23 | 0.8261±0.0000 | 0.7206±0.0000 | 0.5000±0.0000 | 0.9412±0.0000 | 0.7444±0.0000 |
| 18 | 14.0±0.0000 | 16 | 0.8750±0.0000 | 0.8333±0.0000 | 0.7500±0.0000 | 0.9167±0.0000 | 0.8333±0.0000 |
| 19 | 9.0±0.0000 | 11 | 0.8182±0.0000 | 0.8036±0.0000 | 0.7500±0.0000 | 0.8571±0.0000 | 0.8036±0.0000 |
| 20 | 8.0±0.0000 | 10 | 0.8000±0.0000 | 0.7917±0.0000 | 0.7500±0.0000 | 0.8333±0.0000 | 0.7917±0.0000 |

Table 29: HM Random Forest applying laboratory reference values, results of CV, taking best model of grid using F1. Predictions from admission.

| Day | Corrects | Totals | Accuracy | AUC | Sensitivity | Specificity | F1 |
|---|---|---|---|---|---|---|---|
| 1 | 184.6±1.1402 | 194 | 0.9515±0.0059 | 0.8961±0.0085 | 0.8121±0.0136 | 0.9801±0.0052 | 0.9110±0.0104 |
| 2 | 184.2±0.8367 | 194 | 0.9495±0.0043 | 0.9021±0.0124 | 0.8303±0.0271 | 0.9739±0.0052 | 0.9090±0.0081 |
| 3 | 178.0±1.0000 | 187 | 0.9519±0.0053 | 0.8877±0.0082 | 0.7933±0.0149 | 0.9822±0.0053 | 0.9064±0.0098 |
| 4 | 169.0±0.0000 | 180 | 0.9389±0.0000 | 0.8493±0.0062 | 0.7172±0.0154 | 0.9815±0.0030 | 0.8775±0.0018 |
| 5 | 148.2±1.3038 | 160 | 0.9263±0.0081 | 0.8199±0.0207 | 0.6593±0.0406 | 0.9805±0.0041 | 0.8537±0.0180 |
| 6 | 127.6±0.5477 | 142 | 0.8986±0.0039 | 0.7623±0.0087 | 0.5520±0.0179 | 0.9726±0.0038 | 0.7988±0.0082 |
| 7 | 103.2±1.4832 | 117 | 0.8821±0.0127 | 0.7690±0.0318 | 0.5826±0.0659 | 0.9553±0.0089 | 0.7937±0.0270 |
| 8 | 89.4±1.5166 | 100 | 0.8940±0.0152 | 0.7775±0.0314 | 0.5895±0.0577 | 0.9654±0.0055 | 0.8072±0.0303 |
| 9 | 67.8±1.0954 | 79 | 0.8582±0.0139 | 0.7339±0.0271 | 0.5250±0.0559 | 0.9429±0.0142 | 0.7565±0.0245 |
| 10 | 53.8±1.3038 | 62 | 0.8677±0.0210 | 0.7750±0.0318 | 0.6154±0.0544 | 0.9347±0.0171 | 0.7894±0.0333 |
| 11 | 47.8±0.4472 | 54 | 0.8852±0.0083 | 0.7774±0.0053 | 0.5833±0.0000 | 0.9714±0.0106 | 0.8113±0.0102 |
| 12 | 41.2±0.4472 | 50 | 0.8240±0.0089 | 0.6904±0.0186 | 0.4333±0.0373 | 0.9474±0.0000 | 0.7161±0.0190 |
| 13 | 36.4±1.1402 | 44 | 0.8273±0.0259 | 0.6514±0.0158 | 0.3750±0.0000 | 0.9278±0.0317 | 0.6707±0.0275 |
| 14 | 29.2±0.4472 | 37 | 0.7892±0.0121 | 0.6392±0.0077 | 0.3750±0.0000 | 0.9034±0.0154 | 0.6528±0.0115 |
| 15 | 24.2±0.4472 | 31 | 0.7806±0.0144 | 0.6054±0.0093 | 0.2857±0.0000 | 0.9250±0.0186 | 0.6190±0.0130 |
| 16 | 22.0±0.0000 | 28 | 0.7857±0.0000 | 0.6190±0.0000 | 0.2857±0.0000 | 0.9524±0.0000 | 0.6348±0.0000 |
| 17 | 17.4±0.5477 | 23 | 0.7565±0.0238 | 0.5549±0.0340 | 0.1333±0.0745 | 0.9765±0.0322 | 0.5349±0.0636 |
| 18 | 12.4±0.5477 | 16 | 0.7750±0.0342 | 0.5500±0.0685 | 0.1000±0.1369 | 1.0000±0.0000 | 0.5149±0.1182 |
| 19 | 7.2±0.4472 | 11 | 0.6545±0.0407 | 0.5679±0.0319 | 0.2500±0.0000 | 0.8857±0.0639 | 0.5557±0.0313 |
| 20 | 5.6±0.5477 | 10 | 0.5600±0.0548 | 0.5083±0.0456 | 0.2500±0.0000 | 0.7667±0.0913 | 0.4945±0.0401 |

Table 30: HM Random Forest applying laboratory reference values, results of CV, taking best model of grid using F1. Predictions from outcome.

###### 8.5.2 SVC

| Day | Corrects | Totals | Accuracy | AUC | Sensitivity | Specificity | F1 |
|---|---|---|---|---|---|---|---|
| 1 | 162.8±1.3038 | 194 | 0.8392±0.0067 | 0.7610±0.0034 | 0.6424±0.0136 | 0.8795±0.0104 | 0.7385±0.0057 |
| 2 | 168.8±1.6432 | 194 | 0.8701±0.0085 | 0.7386±0.0165 | 0.5394±0.0332 | 0.9379±0.0088 | 0.7542±0.0158 |
| 3 | 166.2±1.6432 | 187 | 0.8888±0.0088 | 0.7639±0.0339 | 0.5800±0.0730 | 0.9478±0.0083 | 0.7794±0.0243 |
| 4 | 161.2±2.2804 | 180 | 0.8956±0.0127 | 0.7957±0.0156 | 0.6483±0.0289 | 0.9430±0.0145 | 0.8026±0.0199 |
| 5 | 148.2±1.3038 | 160 | 0.9263±0.0081 | 0.8228±0.0147 | 0.6667±0.0262 | 0.9789±0.0063 | 0.8549±0.0158 |
| 6 | 129.2±1.7889 | 142 | 0.9099±0.0126 | 0.8163±0.0268 | 0.6720±0.0522 | 0.9607±0.0097 | 0.8349±0.0244 |
| 7 | 106.2±0.8367 | 117 | 0.9077±0.0072 | 0.8440±0.0045 | 0.7391±0.0000 | 0.9489±0.0089 | 0.8510±0.0094 |
| 8 | 87.6±1.5166 | 100 | 0.8760±0.0152 | 0.7623±0.0094 | 0.5789±0.0000 | 0.9457±0.0187 | 0.7828±0.0195 |
| 9 | 69.6±0.5477 | 79 | 0.8810±0.0069 | 0.7855±0.0043 | 0.6250±0.0000 | 0.9460±0.0087 | 0.8037±0.0086 |
| 10 | 53.4±0.8944 | 62 | 0.8613±0.0144 | 0.7766±0.0319 | 0.6308±0.0644 | 0.9224±0.0091 | 0.7840±0.0268 |
| 11 | 48.0±0.0000 | 54 | 0.8889±0.0000 | 0.8095±0.0000 | 0.6667±0.0000 | 0.9524±0.0000 | 0.8288±0.0000 |
| 12 | 42.6±0.5477 | 50 | 0.8520±0.0110 | 0.7487±0.0228 | 0.5500±0.0456 | 0.9474±0.0000 | 0.7734±0.0214 |
| 13 | 37.0±0.7071 | 44 | 0.8409±0.0161 | 0.6694±0.0301 | 0.4000±0.0559 | 0.9389±0.0124 | 0.6915±0.0317 |
| 14 | 30.8±0.4472 | 37 | 0.8324±0.0121 | 0.7121±0.0077 | 0.5000±0.0000 | 0.9241±0.0154 | 0.7300±0.0127 |
| 15 | 24.2±0.4472 | 31 | 0.7806±0.0144 | 0.6155±0.0319 | 0.3143±0.0639 | 0.9167±0.0000 | 0.6286±0.0344 |
| 16 | 22.6±0.8944 | 28 | 0.8071±0.0319 | 0.6905±0.0412 | 0.4571±0.0639 | 0.9238±0.0261 | 0.7101±0.0461 |
| 17 | 20.2±0.8367 | 23 | 0.8783±0.0364 | 0.8206±0.0697 | 0.7000±0.1394 | 0.9412±0.0000 | 0.8321±0.0592 |
| 18 | 14.8±0.4472 | 16 | 0.9250±0.0280 | 0.8667±0.0186 | 0.7500±0.0000 | 0.9833±0.0373 | 0.8935±0.0336 |
| 19 | 8.8±0.8367 | 11 | 0.8000±0.0761 | 0.8000±0.0803 | 0.8000±0.1118 | 0.8000±0.0782 | 0.7898±0.0778 |
| 20 | 8.0±0.0000 | 10 | 0.8000±0.0000 | 0.7917±0.0000 | 0.7500±0.0000 | 0.8333±0.0000 | 0.7917±0.0000 |

Table 31: HM SVC baseline results of CV using base features, taking best model of grid using accuracy. Predictions from admission.

| Day | Corrects | Totals | Accuracy | AUC | Sensitivity | Specificity | F1 |
|---|---|---|---|---|---|---|---|
| 1 | 175.6±1.6733 | 194 | 0.9052±0.0086 | 0.8465±0.0132 | 0.7576±0.0214 | 0.9354±0.0071 | 0.8368±0.0140 |
| 2 | 178.6±0.8944 | 194 | 0.9206±0.0046 | 0.8293±0.0091 | 0.6909±0.0254 | 0.9677±0.0092 | 0.8502±0.0062 |
| 3 | 173.0±1.0000 | 187 | 0.9251±0.0053 | 0.8422±0.0093 | 0.7200±0.0183 | 0.9643±0.0057 | 0.8555±0.0095 |
| 4 | 165.2±0.8367 | 180 | 0.9178±0.0046 | 0.8284±0.0229 | 0.6966±0.0511 | 0.9603±0.0066 | 0.8413±0.0134 |
| 5 | 145.4±1.5166 | 160 | 0.9087±0.0095 | 0.8034±0.0221 | 0.6444±0.0422 | 0.9624±0.0053 | 0.8251±0.0198 |
| 6 | 124.8±1.3038 | 142 | 0.8789±0.0092 | 0.7692±0.0056 | 0.6000±0.0000 | 0.9385±0.0111 | 0.7817±0.0117 |
| 7 | 103.6±0.8944 | 117 | 0.8855±0.0076 | 0.7809±0.0206 | 0.6087±0.0435 | 0.9532±0.0058 | 0.8031±0.0166 |
| 8 | 86.6±1.6733 | 100 | 0.8660±0.0167 | 0.7360±0.0193 | 0.5263±0.0372 | 0.9457±0.0207 | 0.7594±0.0230 |
| 9 | 68.8±1.3038 | 79 | 0.8709±0.0165 | 0.7745±0.0196 | 0.6125±0.0280 | 0.9365±0.0159 | 0.7893±0.0239 |
| 10 | 51.6±0.8944 | 62 | 0.8323±0.0144 | 0.7413±0.0223 | 0.5846±0.0421 | 0.8980±0.0144 | 0.7439±0.0217 |
| 11 | 46.0±0.0000 | 54 | 0.8519±0.0000 | 0.7619±0.0133 | 0.6000±0.0373 | 0.9238±0.0106 | 0.7745±0.0063 |
| 12 | 41.4±0.5477 | 50 | 0.8280±0.0110 | 0.7272±0.0183 | 0.5333±0.0456 | 0.9211±0.0186 | 0.7441±0.0160 |
| 13 | 36.0±1.0000 | 44 | 0.8182±0.0227 | 0.6458±0.0139 | 0.3750±0.0000 | 0.9167±0.0278 | 0.6610±0.0229 |
| 14 | 27.8±0.8367 | 37 | 0.7514±0.0226 | 0.6151±0.0144 | 0.3750±0.0000 | 0.8552±0.0289 | 0.6195±0.0193 |
| 15 | 23.0±0.7071 | 31 | 0.7419±0.0228 | 0.6411±0.0270 | 0.4571±0.0639 | 0.8250±0.0349 | 0.6377±0.0241 |
| 16 | 19.8±0.4472 | 28 | 0.7071±0.0160 | 0.5667±0.0106 | 0.2857±0.0000 | 0.8476±0.0213 | 0.5704±0.0119 |
| 17 | 18.0±0.7071 | 23 | 0.7826±0.0307 | 0.6480±0.0425 | 0.3667±0.0745 | 0.9294±0.0263 | 0.6650±0.0474 |
| 18 | 9.8±1.3038 | 16 | 0.6125±0.0815 | 0.4917±0.1079 | 0.2500±0.1768 | 0.7333±0.0697 | 0.4889±0.1120 |
| 19 | 7.6±0.5477 | 11 | 0.6909±0.0498 | 0.6821±0.0685 | 0.6500±0.1369 | 0.7143±0.0000 | 0.6736±0.0607 |
| 20 | 7.6±0.5477 | 10 | 0.7600±0.0548 | 0.7500±0.0589 | 0.7000±0.1118 | 0.8000±0.0745 | 0.7485±0.0599 |

Table 32: HM SVC baseline results of CV using base features, taking best model of grid using accuracy. Predictions from outcome.

| Day | Corrects | Totals | Accuracy | AUC | Sensitivity | Specificity | F1 |
|---|---|---|---|---|---|---|---|
| 1 | 159.0±1.4142 | 194 | 0.8196±0.0073 | 0.7612±0.0194 | 0.6727±0.0395 | 0.8497±0.0052 | 0.7228±0.0136 |
| 2 | 169.2±1.9235 | 194 | 0.8722±0.0099 | 0.7592±0.0073 | 0.5879±0.0166 | 0.9304±0.0135 | 0.7670±0.0119 |
| 3 | 165.0±1.5811 | 187 | 0.8824±0.0085 | 0.8005±0.0153 | 0.6800±0.0298 | 0.9210±0.0085 | 0.7895±0.0142 |
| 4 | 161.6±1.1402 | 180 | 0.8978±0.0063 | 0.8276±0.0038 | 0.7241±0.0000 | 0.9311±0.0076 | 0.8171±0.0086 |
| 5 | 145.0±0.7071 | 160 | 0.9062±0.0044 | 0.8196±0.0085 | 0.6889±0.0203 | 0.9504±0.0067 | 0.8283±0.0069 |
| 6 | 129.8±0.8367 | 142 | 0.9141±0.0059 | 0.8535±0.0143 | 0.7600±0.0283 | 0.9470±0.0038 | 0.8523±0.0111 |
| 7 | 106.2±0.8367 | 117 | 0.9077±0.0072 | 0.8605±0.0205 | 0.7826±0.0435 | 0.9383±0.0048 | 0.8556±0.0135 |
| 8 | 84.8±1.6432 | 100 | 0.8480±0.0164 | 0.7531±0.0158 | 0.6000±0.0288 | 0.9062±0.0207 | 0.7533±0.0201 |
| 9 | 67.4±0.8944 | 79 | 0.8532±0.0113 | 0.7681±0.0071 | 0.6250±0.0000 | 0.9111±0.0142 | 0.7707±0.0127 |
| 10 | 52.6±0.8944 | 62 | 0.8484±0.0144 | 0.7741±0.0241 | 0.6462±0.0421 | 0.9020±0.0091 | 0.7725±0.0226 |
| 11 | 47.2±0.4472 | 54 | 0.8741±0.0083 | 0.8000±0.0053 | 0.6667±0.0000 | 0.9333±0.0106 | 0.8111±0.0099 |
| 12 | 42.8±1.4832 | 50 | 0.8560±0.0297 | 0.7798±0.0435 | 0.6333±0.0745 | 0.9263±0.0220 | 0.7926±0.0428 |
| 13 | 37.4±1.5166 | 44 | 0.8500±0.0345 | 0.7236±0.0604 | 0.5250±0.1046 | 0.9222±0.0232 | 0.7343±0.0627 |
| 14 | 30.8±0.8367 | 37 | 0.8324±0.0226 | 0.7211±0.0330 | 0.5250±0.0559 | 0.9172±0.0189 | 0.7354±0.0341 |
| 15 | 25.2±0.8367 | 31 | 0.8129±0.0270 | 0.6869±0.0598 | 0.4571±0.1195 | 0.9167±0.0000 | 0.7013±0.0595 |
| 16 | 21.8±0.4472 | 28 | 0.7786±0.0160 | 0.7000±0.0319 | 0.5429±0.0639 | 0.8571±0.0000 | 0.7013±0.0290 |
| 17 | 20.6±1.6733 | 23 | 0.8957±0.0728 | 0.8755±0.1240 | 0.8333±0.2357 | 0.9176±0.0322 | 0.8624±0.1037 |
| 18 | 14.2±0.4472 | 16 | 0.8875±0.0280 | 0.8417±0.0186 | 0.7500±0.0000 | 0.9333±0.0373 | 0.8484±0.0336 |
| 19 | 9.8±0.8367 | 11 | 0.8909±0.0761 | 0.9143±0.0598 | 1.0000±0.0000 | 0.8286±0.1195 | 0.8891±0.0764 |
| 20 | 8.0±0.0000 | 10 | 0.8000±0.0000 | 0.7917±0.0000 | 0.7500±0.0000 | 0.8333±0.0000 | 0.7917±0.0000 |

Table 33: HM SVC baseline results of CV using base features, taking best model of grid using F1. Predictions from admission.

| Day | Corrects | Totals | Accuracy | AUC | Sensitivity | Specificity | F1 |
|---|---|---|---|---|---|---|---|
| 1 | 174.6±1.8166 | 194 | 0.9000±0.0094 | 0.8651±0.0176 | 0.8121±0.0332 | 0.9180±0.0081 | 0.8363±0.0152 |
| 2 | 177.8±0.8367 | 194 | 0.9165±0.0043 | 0.8533±0.0099 | 0.7576±0.0214 | 0.9491±0.0052 | 0.8525±0.0075 |
| 3 | 170.8±0.4472 | 187 | 0.9134±0.0024 | 0.8432±0.0065 | 0.7400±0.0149 | 0.9465±0.0035 | 0.8405±0.0043 |
| 4 | 164.6±2.0736 | 180 | 0.9144±0.0115 | 0.8571±0.0240 | 0.7724±0.0463 | 0.9417±0.0098 | 0.8463±0.0211 |
| 5 | 146.2±1.7889 | 160 | 0.9138±0.0112 | 0.8419±0.0219 | 0.7333±0.0406 | 0.9504±0.0086 | 0.8449±0.0204 |
| 6 | 125.8±1.6432 | 142 | 0.8859±0.0116 | 0.8081±0.0273 | 0.6880±0.0522 | 0.9282±0.0047 | 0.8049±0.0223 |
| 7 | 104.2±1.9235 | 117 | 0.8906±0.0164 | 0.8137±0.0173 | 0.6870±0.0194 | 0.9404±0.0161 | 0.8224±0.0231 |
| 8 | 87.2±1.9235 | 100 | 0.8720±0.0192 | 0.7719±0.0373 | 0.6105±0.0706 | 0.9333±0.0141 | 0.7828±0.0346 |
| 9 | 67.8±1.0954 | 79 | 0.8582±0.0139 | 0.7759±0.0172 | 0.6375±0.0280 | 0.9143±0.0142 | 0.7786±0.0188 |
| 10 | 49.6±0.8944 | 62 | 0.8000±0.0144 | 0.7378±0.0314 | 0.6308±0.0644 | 0.8449±0.0112 | 0.7192±0.0245 |
| 11 | 46.0±0.7071 | 54 | 0.8519±0.0131 | 0.7917±0.0157 | 0.6833±0.0373 | 0.9000±0.0199 | 0.7882±0.0153 |
| 12 | 39.4±0.8944 | 50 | 0.7880±0.0179 | 0.6895±0.0118 | 0.5000±0.0000 | 0.8789±0.0235 | 0.6973±0.0176 |
| 13 | 33.2±0.8367 | 44 | 0.7545±0.0190 | 0.5778±0.0266 | 0.3000±0.0685 | 0.8556±0.0304 | 0.5781±0.0246 |
| 14 | 26.4±1.1402 | 37 | 0.7135±0.0308 | 0.6091±0.0431 | 0.4250±0.0685 | 0.7931±0.0244 | 0.6018±0.0409 |
| 15 | 22.8±0.8367 | 31 | 0.7355±0.0270 | 0.6369±0.0304 | 0.4571±0.0639 | 0.8167±0.0373 | 0.6324±0.0285 |
| 16 | 19.2±1.3038 | 28 | 0.6857±0.0466 | 0.5524±0.0616 | 0.2857±0.1010 | 0.8190±0.0398 | 0.5538±0.0637 |
| 17 | 18.0±1.0000 | 23 | 0.7826±0.0435 | 0.7020±0.0473 | 0.5333±0.0745 | 0.8706±0.0492 | 0.7088±0.0502 |
| 18 | 10.0±1.0000 | 16 | 0.6250±0.0625 | 0.5833±0.1250 | 0.5000±0.2500 | 0.6667±0.0000 | 0.5578±0.0974 |
| 19 | 7.0±0.7071 | 11 | 0.6364±0.0643 | 0.6071±0.0884 | 0.5000±0.1768 | 0.7143±0.0000 | 0.6031±0.0857 |
| 20 | 7.8±0.4472 | 10 | 0.7800±0.0447 | 0.7667±0.0559 | 0.7000±0.1118 | 0.8333±0.0000 | 0.7674±0.0543 |

Table 34: HM SVC baseline results of CV using base features, taking best model of grid using F1. Predictions from outcome.

| Day | Corrects | Totals | Accuracy | AUC | Sensitivity | Specificity | F1 |
|---|---|---|---|---|---|---|---|
| 1 | 165.8±2.0494 | 194 | 0.8546±0.0106 | 0.7871±0.0150 | 0.6848±0.0271 | 0.8894±0.0111 | 0.7631±0.0149 |
| 2 | 168.2±0.8367 | 194 | 0.8670±0.0043 | 0.7609±0.0076 | 0.6000±0.0136 | 0.9217±0.0034 | 0.7628±0.0075 |
| 3 | 165.4±1.5166 | 187 | 0.8845±0.0081 | 0.7802±0.0150 | 0.6267±0.0279 | 0.9338±0.0073 | 0.7833±0.0146 |
| 4 | 164.6±1.5166 | 180 | 0.9144±0.0084 | 0.8292±0.0220 | 0.7034±0.0463 | 0.9550±0.0086 | 0.8374±0.0174 |
| 5 | 149.4±1.5166 | 160 | 0.9337±0.0095 | 0.8362±0.0225 | 0.6889±0.0422 | 0.9835±0.0034 | 0.8694±0.0203 |
| 6 | 132.0±1.2247 | 142 | 0.9296±0.0086 | 0.8566±0.0116 | 0.7440±0.0219 | 0.9692±0.0097 | 0.8731±0.0141 |
| 7 | 107.6±1.1402 | 117 | 0.9197±0.0097 | 0.8548±0.0182 | 0.7478±0.0364 | 0.9617±0.0095 | 0.8679±0.0160 |
| 8 | 88.4±1.3416 | 100 | 0.8840±0.0134 | 0.8035±0.0106 | 0.6737±0.0235 | 0.9333±0.0187 | 0.8087±0.0158 |
| 9 | 69.0±0.7071 | 79 | 0.8734±0.0090 | 0.7994±0.0118 | 0.6750±0.0280 | 0.9238±0.0133 | 0.8022±0.0115 |
| 10 | 53.2±0.8367 | 62 | 0.8581±0.0135 | 0.7915±0.0162 | 0.6769±0.0344 | 0.9061±0.0183 | 0.7883±0.0168 |
| 11 | 49.2±0.4472 | 54 | 0.9111±0.0083 | 0.8476±0.0155 | 0.7333±0.0373 | 0.9619±0.0130 | 0.8647±0.0125 |
| 12 | 45.6±0.8944 | 50 | 0.9120±0.0179 | 0.8680±0.0336 | 0.7833±0.0745 | 0.9526±0.0220 | 0.8761±0.0267 |
| 13 | 39.6±1.6733 | 44 | 0.9000±0.0380 | 0.8125±0.0769 | 0.6750±0.1425 | 0.9500±0.0232 | 0.8236±0.0694 |
| 14 | 31.8±0.4472 | 37 | 0.8595±0.0121 | 0.7836±0.0234 | 0.6500±0.0559 | 0.9172±0.0189 | 0.7885±0.0174 |
| 15 | 25.8±0.8367 | 31 | 0.8323±0.0270 | 0.7399±0.0514 | 0.5714±0.1010 | 0.9083±0.0186 | 0.7484±0.0466 |
| 16 | 24.0±0.0000 | 28 | 0.8571±0.0000 | 0.8095±0.0000 | 0.7143±0.0000 | 0.9048±0.0000 | 0.8095±0.0000 |
| 17 | 21.4±0.5477 | 23 | 0.9304±0.0238 | 0.9206±0.0456 | 0.9000±0.0913 | 0.9412±0.0000 | 0.9109±0.0324 |
| 18 | 14.2±0.4472 | 16 | 0.8875±0.0280 | 0.8583±0.0559 | 0.8000±0.1118 | 0.9167±0.0000 | 0.8512±0.0400 |
| 19 | 9.8±0.4472 | 11 | 0.8909±0.0407 | 0.9036±0.0559 | 0.9500±0.1118 | 0.8571±0.0000 | 0.8855±0.0458 |
| 20 | 8.8±0.4472 | 10 | 0.8800±0.0447 | 0.8917±0.0559 | 0.9500±0.1118 | 0.8333±0.0000 | 0.8775±0.0480 |

Table 35: HM SVC with imputation, results of CV, taking best model of grid using accuracy. Predictions from admission.

| Day | Corrects | Totals | Accuracy | AUC | Sensitivity | Specificity | F1 |
|---|---|---|---|---|---|---|---|
| 1 | 183.2±2.1679 | 194 | 0.9443±0.0112 | 0.9135±0.0116 | 0.8667±0.0166 | 0.9602±0.0121 | 0.9040±0.0180 |
| 2 | 183.0±1.5811 | 194 | 0.9433±0.0082 | 0.8960±0.0148 | 0.8242±0.0254 | 0.9677±0.0052 | 0.8988±0.0146 |
| 3 | 172.8±1.0954 | 187 | 0.9241±0.0059 | 0.8523±0.0152 | 0.7467±0.0298 | 0.9580±0.0035 | 0.8570±0.0122 |
| 4 | 167.6±1.5166 | 180 | 0.9311±0.0084 | 0.8726±0.0269 | 0.7862±0.0567 | 0.9589±0.0073 | 0.8722±0.0182 |
| 5 | 146.0±1.4142 | 160 | 0.9125±0.0088 | 0.8352±0.0274 | 0.7185±0.0620 | 0.9519±0.0126 | 0.8409±0.0175 |
| 6 | 126.0±1.0000 | 142 | 0.8873±0.0070 | 0.7995±0.0096 | 0.6640±0.0219 | 0.9350±0.0097 | 0.8034±0.0098 |
| 7 | 105.4±0.8944 | 117 | 0.9009±0.0076 | 0.8332±0.0118 | 0.7217±0.0238 | 0.9447±0.0089 | 0.8399±0.0114 |
| 8 | 88.8±0.8367 | 100 | 0.8880±0.0084 | 0.7818±0.0226 | 0.6105±0.0471 | 0.9531±0.0055 | 0.8031±0.0180 |
| 9 | 70.4±1.1402 | 79 | 0.8911±0.0144 | 0.8292±0.0279 | 0.7250±0.0559 | 0.9333±0.0133 | 0.8305±0.0242 |
| 10 | 54.0±1.2247 | 62 | 0.8710±0.0198 | 0.8053±0.0290 | 0.6923±0.0544 | 0.9184±0.0204 | 0.8054±0.0284 |
| 11 | 46.2±1.3038 | 54 | 0.8556±0.0241 | 0.7821±0.0284 | 0.6500±0.0373 | 0.9143±0.0213 | 0.7876±0.0323 |
| 12 | 41.2±0.8367 | 50 | 0.8240±0.0167 | 0.7189±0.0208 | 0.5167±0.0373 | 0.9211±0.0186 | 0.7366±0.0224 |
| 13 | 34.4±1.1402 | 44 | 0.7818±0.0259 | 0.6722±0.0158 | 0.5000±0.0000 | 0.8444±0.0317 | 0.6598±0.0242 |
| 14 | 28.6±0.5477 | 37 | 0.7730±0.0148 | 0.6741±0.0094 | 0.5000±0.0000 | 0.8483±0.0189 | 0.6712±0.0135 |
| 15 | 24.0±0.0000 | 31 | 0.7742±0.0000 | 0.6619±0.0226 | 0.4571±0.0639 | 0.8667±0.0186 | 0.6659±0.0147 |
| 16 | 19.0±1.5811 | 28 | 0.6786±0.0565 | 0.5381±0.0868 | 0.2571±0.1565 | 0.8190±0.0398 | 0.5356±0.0918 |
| 17 | 17.8±0.8367 | 23 | 0.7739±0.0364 | 0.6529±0.0424 | 0.4000±0.0913 | 0.9059±0.0526 | 0.6667±0.0457 |
| 18 | 10.2±0.4472 | 16 | 0.6375±0.0280 | 0.5917±0.0186 | 0.5000±0.0000 | 0.6833±0.0373 | 0.5736±0.0223 |
| 19 | 7.2±0.4472 | 11 | 0.6545±0.0407 | 0.6321±0.0559 | 0.5500±0.1118 | 0.7143±0.0000 | 0.6293±0.0496 |
| 20 | 8.0±0.0000 | 10 | 0.8000±0.0000 | 0.7917±0.0000 | 0.7500±0.0000 | 0.8333±0.0000 | 0.7917±0.0000 |

Table 36: HM SVC with imputation, results of CV, taking best model of grid using accuracy. Predictions from outcome.

| Day | Corrects | Totals | Accuracy | AUC | Sensitivity | Specificity | F1 |
|---|---|---|---|---|---|---|---|
| 1 | 164.4±2.6077 | 194 | 0.8474±0.0134 | 0.7900±0.0337 | 0.7030±0.0657 | 0.8770±0.0068 | 0.7575±0.0251 |
| 2 | 169.4±0.5477 | 194 | 0.8732±0.0028 | 0.7839±0.0071 | 0.6485±0.0166 | 0.9193±0.0044 | 0.7791±0.0048 |
| 3 | 164.0±1.0000 | 187 | 0.8770±0.0053 | 0.7758±0.0134 | 0.6267±0.0279 | 0.9248±0.0053 | 0.7735±0.0106 |
| 4 | 161.0±1.4142 | 180 | 0.8944±0.0079 | 0.8256±0.0201 | 0.7241±0.0422 | 0.9272±0.0081 | 0.8124±0.0151 |
| 5 | 147.4±1.5166 | 160 | 0.9213±0.0095 | 0.8346±0.0197 | 0.7037±0.0370 | 0.9654±0.0067 | 0.8520±0.0182 |
| 6 | 129.4±1.5166 | 142 | 0.9113±0.0107 | 0.8424±0.0195 | 0.7360±0.0358 | 0.9487±0.0085 | 0.8456±0.0186 |
| 7 | 104.6±1.5166 | 117 | 0.8940±0.0130 | 0.8388±0.0241 | 0.7478±0.0476 | 0.9298±0.0121 | 0.8343±0.0208 |
| 8 | 86.0±1.0000 | 100 | 0.8600±0.0100 | 0.7806±0.0149 | 0.6526±0.0288 | 0.9086±0.0110 | 0.7762±0.0144 |
| 9 | 69.2±0.4472 | 79 | 0.8759±0.0057 | 0.8150±0.0152 | 0.7125±0.0342 | 0.9175±0.0071 | 0.8105±0.0103 |
| 10 | 53.4±0.5477 | 62 | 0.8613±0.0088 | 0.8105±0.0173 | 0.7231±0.0421 | 0.8980±0.0144 | 0.7984±0.0123 |
| 11 | 48.4±0.8944 | 54 | 0.8963±0.0166 | 0.8381±0.0190 | 0.7333±0.0373 | 0.9429±0.0213 | 0.8465±0.0215 |
| 12 | 43.2±0.4472 | 50 | 0.8640±0.0089 | 0.8250±0.0210 | 0.7500±0.0589 | 0.9000±0.0220 | 0.8174±0.0124 |
| 13 | 38.4±1.1402 | 44 | 0.8727±0.0259 | 0.7861±0.0681 | 0.6500±0.1369 | 0.9222±0.0124 | 0.7840±0.0516 |
| 14 | 31.0±0.0000 | 37 | 0.8378±0.0000 | 0.7517±0.0202 | 0.6000±0.0559 | 0.9034±0.0154 | 0.7558±0.0112 |
| 15 | 25.6±0.5477 | 31 | 0.8258±0.0177 | 0.7357±0.0440 | 0.5714±0.1010 | 0.9000±0.0228 | 0.7413±0.0351 |
| 16 | 24.2±0.8367 | 28 | 0.8643±0.0299 | 0.8238±0.0598 | 0.7429±0.1195 | 0.9048±0.0000 | 0.8191±0.0473 |
| 17 | 21.4±0.5477 | 23 | 0.9304±0.0238 | 0.9206±0.0456 | 0.9000±0.0913 | 0.9412±0.0000 | 0.9109±0.0324 |
| 18 | 14.2±0.4472 | 16 | 0.8875±0.0280 | 0.8583±0.0559 | 0.8000±0.1118 | 0.9167±0.0000 | 0.8512±0.0400 |
| 19 | 10.0±0.0000 | 11 | 0.9091±0.0000 | 0.9286±0.0000 | 1.0000±0.0000 | 0.8571±0.0000 | 0.9060±0.0000 |
| 20 | 9.0±0.0000 | 10 | 0.9000±0.0000 | 0.9167±0.0000 | 1.0000±0.0000 | 0.8333±0.0000 | 0.8990±0.0000 |

Table 37: HM SVC with imputation, results of CV, taking best model of grid using F1. Predictions from admission.

| Day | Corrects | Totals | Accuracy | AUC | Sensitivity | Specificity | F1 |
|---|---|---|---|---|---|---|---|
| 1 | 182.4±2.3022 | 194 | 0.9402±0.0119 | 0.9182±0.0155 | 0.8848±0.0254 | 0.9516±0.0119 | 0.8991±0.0189 |
| 2 | 181.4±2.9665 | 194 | 0.9351±0.0153 | 0.8934±0.0272 | 0.8303±0.0460 | 0.9565±0.0098 | 0.8869±0.0264 |
| 3 | 172.0±0.7071 | 187 | 0.9198±0.0038 | 0.8525±0.0077 | 0.7533±0.0183 | 0.9516±0.0057 | 0.8515±0.0060 |
| 4 | 166.6±1.5166 | 180 | 0.9256±0.0084 | 0.8693±0.0256 | 0.7862±0.0567 | 0.9523±0.0109 | 0.8639±0.0170 |
| 5 | 145.2±0.8367 | 160 | 0.9075±0.0052 | 0.8263±0.0263 | 0.7037±0.0586 | 0.9489±0.0063 | 0.8317±0.0153 |
| 6 | 126.0±0.7071 | 142 | 0.8873±0.0050 | 0.8121±0.0108 | 0.6960±0.0219 | 0.9282±0.0047 | 0.8082±0.0088 |
| 7 | 103.0±0.7071 | 117 | 0.8803±0.0060 | 0.8139±0.0106 | 0.7043±0.0194 | 0.9234±0.0048 | 0.8118±0.0096 |
| 8 | 88.0±1.2247 | 100 | 0.8800±0.0122 | 0.7930±0.0155 | 0.6526±0.0288 | 0.9333±0.0141 | 0.8003±0.0177 |
| 9 | 69.4±0.5477 | 79 | 0.8785±0.0069 | 0.8212±0.0160 | 0.7250±0.0342 | 0.9175±0.0071 | 0.8152±0.0119 |
| 10 | 52.4±0.8944 | 62 | 0.8452±0.0144 | 0.7890±0.0278 | 0.6923±0.0544 | 0.8857±0.0112 | 0.7761±0.0227 |
| 11 | 45.0±1.5811 | 54 | 0.8333±0.0293 | 0.7798±0.0431 | 0.6833±0.0697 | 0.8762±0.0199 | 0.7683±0.0411 |
| 12 | 39.8±0.8367 | 50 | 0.7960±0.0167 | 0.7118±0.0258 | 0.5500±0.0456 | 0.8737±0.0118 | 0.7153±0.0250 |
| 13 | 31.2±0.8367 | 44 | 0.7091±0.0190 | 0.6083±0.0253 | 0.4500±0.0685 | 0.7667±0.0317 | 0.5851±0.0183 |
| 14 | 27.0±1.2247 | 37 | 0.7297±0.0331 | 0.6466±0.0514 | 0.5000±0.0884 | 0.7931±0.0244 | 0.6326±0.0455 |
| 15 | 21.8±0.8367 | 31 | 0.7032±0.0270 | 0.6060±0.0174 | 0.4286±0.0000 | 0.7833±0.0349 | 0.5994±0.0217 |
| 16 | 20.0±1.2247 | 28 | 0.7143±0.0437 | 0.5905±0.0516 | 0.3429±0.0782 | 0.8381±0.0426 | 0.5948±0.0541 |
| 17 | 17.8±0.4472 | 23 | 0.7739±0.0194 | 0.6853±0.0132 | 0.5000±0.0000 | 0.8706±0.0263 | 0.6934±0.0177 |
| 18 | 10.2±0.4472 | 16 | 0.6375±0.0280 | 0.5917±0.0186 | 0.5000±0.0000 | 0.6833±0.0373 | 0.5736±0.0223 |
| 19 | 7.4±0.5477 | 11 | 0.6727±0.0498 | 0.6571±0.0685 | 0.6000±0.1369 | 0.7143±0.0000 | 0.6515±0.0607 |
| 20 | 7.6±0.5477 | 10 | 0.7600±0.0548 | 0.7583±0.0456 | 0.7500±0.0000 | 0.7667±0.0913 | 0.7538±0.0519 |

Table 38: HM SVC with imputation, results of CV, taking best model of grid using F1. Predictions from outcome.

| Day | Corrects | Totals | Accuracy | AUC | Sensitivity | Specificity | F1 |
|---|---|---|---|---|---|---|---|
| 1 | 162.6±0.8944 | 194 | 0.8381±0.0046 | 0.7700±0.0108 | 0.6667±0.0214 | 0.8733±0.0034 | 0.7415±0.0083 |
| 2 | 166.8±1.3038 | 194 | 0.8598±0.0067 | 0.7806±0.0123 | 0.6606±0.0254 | 0.9006±0.0076 | 0.7650±0.0104 |
| 3 | 167.0±1.5811 | 187 | 0.8930±0.0085 | 0.8204±0.0235 | 0.7133±0.0506 | 0.9274±0.0097 | 0.8084±0.0163 |
| 4 | 162.6±0.5477 | 180 | 0.9033±0.0030 | 0.8560±0.0057 | 0.7862±0.0154 | 0.9258±0.0055 | 0.8326±0.0038 |
| 5 | 146.0±1.0000 | 160 | 0.9125±0.0062 | 0.8441±0.0098 | 0.7407±0.0262 | 0.9474±0.0106 | 0.8441±0.0082 |
| 6 | 129.0±0.7071 | 142 | 0.9085±0.0050 | 0.8501±0.0030 | 0.7600±0.0000 | 0.9402±0.0060 | 0.8447±0.0068 |
| 7 | 104.2±1.3038 | 117 | 0.8906±0.0111 | 0.8432±0.0252 | 0.7652±0.0496 | 0.9213±0.0058 | 0.8320±0.0193 |
| 8 | 86.6±1.1402 | 100 | 0.8660±0.0114 | 0.8327±0.0223 | 0.7789±0.0440 | 0.8864±0.0103 | 0.8014±0.0174 |
| 9 | 69.4±0.5477 | 79 | 0.8785±0.0069 | 0.8306±0.0196 | 0.7500±0.0442 | 0.9111±0.0087 | 0.8184±0.0125 |
| 10 | 54.4±1.8166 | 62 | 0.8774±0.0293 | 0.8546±0.0552 | 0.8154±0.1032 | 0.8939±0.0171 | 0.8275±0.0437 |
| 11 | 48.8±1.0954 | 54 | 0.9037±0.0203 | 0.8488±0.0311 | 0.7500±0.0589 | 0.9476±0.0199 | 0.8572±0.0299 |
| 12 | 45.0±1.5811 | 50 | 0.9000±0.0316 | 0.8658±0.0583 | 0.8000±0.1118 | 0.9316±0.0144 | 0.8626±0.0469 |
| 13 | 38.4±0.8944 | 44 | 0.8727±0.0203 | 0.7861±0.0518 | 0.6500±0.1046 | 0.9222±0.0124 | 0.7848±0.0416 |
| 14 | 32.4±1.3416 | 37 | 0.8757±0.0363 | 0.8664±0.0607 | 0.8500±0.1046 | 0.8828±0.0189 | 0.8319±0.0504 |
| 15 | 27.6±0.5477 | 31 | 0.8903±0.0177 | 0.8583±0.0391 | 0.8000±0.0782 | 0.9167±0.0000 | 0.8470±0.0288 |
| 16 | 25.2±0.4472 | 28 | 0.9000±0.0160 | 0.8952±0.0319 | 0.8857±0.0639 | 0.9048±0.0000 | 0.8732±0.0220 |
| 17 | 21.8±0.4472 | 23 | 0.9478±0.0194 | 0.9539±0.0373 | 0.9667±0.0745 | 0.9412±0.0000 | 0.9346±0.0264 |
| 18 | 14.0±1.0000 | 16 | 0.8750±0.0625 | 0.8333±0.1250 | 0.7500±0.2500 | 0.9167±0.0000 | 0.8260±0.0986 |
| 19 | 10.4±0.5477 | 11 | 0.9455±0.0498 | 0.9571±0.0391 | 1.0000±0.0000 | 0.9143±0.0782 | 0.9436±0.0515 |
| 20 | 9.0±0.0000 | 10 | 0.9000±0.0000 | 0.9167±0.0000 | 1.0000±0.0000 | 0.8333±0.0000 | 0.8990±0.0000 |

Table 39: HM SVC with imputation, with missing flags, results of CV, taking best model of grid using accuracy. Predictions from admission.

| Day | Corrects | Totals | Accuracy | AUC | Sensitivity | Specificity | F1 |
|---|---|---|---|---|---|---|---|
| 1 | 182.6±1.8166 | 194 | 0.9412±0.0094 | 0.9333±0.0121 | 0.9212±0.0166 | 0.9453±0.0081 | 0.9031±0.0147 |
| 2 | 182.4±1.3416 | 194 | 0.9402±0.0069 | 0.9086±0.0091 | 0.8606±0.0166 | 0.9565±0.0076 | 0.8971±0.0109 |
| 3 | 174.2±0.4472 | 187 | 0.9316±0.0024 | 0.8783±0.0143 | 0.8000±0.0333 | 0.9567±0.0053 | 0.8742±0.0062 |
| 4 | 166.6±0.8944 | 180 | 0.9256±0.0050 | 0.8860±0.0030 | 0.8276±0.0000 | 0.9444±0.0059 | 0.8685±0.0073 |
| 5 | 144.0±2.1213 | 160 | 0.9000±0.0133 | 0.8483±0.0234 | 0.7704±0.0406 | 0.9263±0.0098 | 0.8306±0.0222 |
| 6 | 127.4±1.8166 | 142 | 0.8972±0.0128 | 0.8464±0.0304 | 0.7680±0.0593 | 0.9248±0.0072 | 0.8304±0.0235 |
| 7 | 103.6±1.8166 | 117 | 0.8855±0.0155 | 0.8171±0.0338 | 0.7043±0.0645 | 0.9298±0.0058 | 0.8177±0.0281 |
| 8 | 88.8±1.3038 | 100 | 0.8880±0.0130 | 0.8100±0.0213 | 0.6842±0.0372 | 0.9358±0.0103 | 0.8151±0.0212 |
| 9 | 70.2±2.2804 | 79 | 0.8886±0.0289 | 0.8462±0.0428 | 0.7750±0.0713 | 0.9175±0.0235 | 0.8338±0.0424 |
| 10 | 51.0±1.2247 | 62 | 0.8226±0.0198 | 0.7691±0.0428 | 0.6769±0.0843 | 0.8612±0.0091 | 0.7493±0.0340 |
| 11 | 45.8±1.6432 | 54 | 0.8481±0.0304 | 0.8131±0.0483 | 0.7500±0.0833 | 0.8762±0.0199 | 0.7931±0.0427 |
| 12 | 39.2±1.9235 | 50 | 0.7840±0.0385 | 0.7096±0.0447 | 0.5667±0.0697 | 0.8526±0.0399 | 0.7075±0.0466 |
| 13 | 33.2±0.8367 | 44 | 0.7545±0.0190 | 0.6653±0.0296 | 0.5250±0.0559 | 0.8056±0.0196 | 0.6401±0.0246 |
| 14 | 26.8±1.7889 | 37 | 0.7243±0.0483 | 0.6884±0.0704 | 0.6250±0.1250 | 0.7517±0.0450 | 0.6523±0.0590 |
| 15 | 22.2±0.8367 | 31 | 0.7161±0.0270 | 0.6345±0.0438 | 0.4857±0.0782 | 0.7833±0.0186 | 0.6227±0.0383 |
| 16 | 20.4±2.3022 | 28 | 0.7286±0.0822 | 0.6857±0.1134 | 0.6000±0.1863 | 0.7714±0.0621 | 0.6666±0.1010 |
| 17 | 17.4±0.5477 | 23 | 0.7565±0.0238 | 0.7059±0.0312 | 0.6000±0.0913 | 0.8118±0.0492 | 0.6958±0.0251 |
| 18 | 9.6±0.5477 | 16 | 0.6000±0.0342 | 0.5667±0.0228 | 0.5000±0.0000 | 0.6333±0.0456 | 0.5442±0.0266 |
| 19 | 7.8±0.4472 | 11 | 0.7091±0.0407 | 0.7071±0.0559 | 0.7000±0.1118 | 0.7143±0.0000 | 0.6958±0.0496 |
| 20 | 8.0±0.0000 | 10 | 0.8000±0.0000 | 0.7917±0.0000 | 0.7500±0.0000 | 0.8333±0.0000 | 0.7917±0.0000 |

Table 40: HM SVC with imputation, with missing flags, results of CV, taking best model of grid using accuracy. Predictions from outcome.

| Day | Corrects | Totals | Accuracy | AUC | Sensitivity | Specificity | F1 |
|---|---|---|---|---|---|---|---|
| 1 | 164.2±1.9235 | 194 | 0.8464±0.0099 | 0.7918±0.0148 | 0.7091±0.0271 | 0.8745±0.0102 | 0.7577±0.0139 |
| 2 | 167.2±0.4472 | 194 | 0.8619±0.0023 | 0.7867±0.0109 | 0.6727±0.0254 | 0.9006±0.0044 | 0.7694±0.0061 |
| 3 | 167.4±0.8944 | 187 | 0.8952±0.0048 | 0.8351±0.0094 | 0.7467±0.0183 | 0.9236±0.0045 | 0.8162±0.0081 |
| 4 | 163.0±0.7071 | 180 | 0.9056±0.0039 | 0.8601±0.0112 | 0.7931±0.0244 | 0.9272±0.0047 | 0.8364±0.0072 |
| 5 | 146.4±1.1402 | 160 | 0.9150±0.0071 | 0.8544±0.0208 | 0.7630±0.0422 | 0.9459±0.0034 | 0.8501±0.0148 |
| 6 | 128.0±1.0000 | 142 | 0.9014±0.0070 | 0.8458±0.0043 | 0.7600±0.0000 | 0.9316±0.0085 | 0.8353±0.0093 |
| 7 | 104.4±0.8944 | 117 | 0.8923±0.0076 | 0.8542±0.0180 | 0.7913±0.0364 | 0.9170±0.0048 | 0.8373±0.0131 |
| 8 | 87.0±1.5811 | 100 | 0.8700±0.0158 | 0.8432±0.0238 | 0.8000±0.0440 | 0.8864±0.0161 | 0.8088±0.0219 |
| 9 | 70.0±0.7071 | 79 | 0.8861±0.0090 | 0.8493±0.0255 | 0.7875±0.0559 | 0.9111±0.0087 | 0.8318±0.0158 |
| 10 | 55.6±1.1402 | 62 | 0.8968±0.0184 | 0.8951±0.0422 | 0.8923±0.0877 | 0.8980±0.0144 | 0.8574±0.0282 |
| 11 | 49.4±1.1402 | 54 | 0.9148±0.0211 | 0.8798±0.0247 | 0.8167±0.0373 | 0.9429±0.0213 | 0.8778±0.0291 |
| 12 | 45.4±1.5166 | 50 | 0.9080±0.0303 | 0.8882±0.0511 | 0.8500±0.0913 | 0.9263±0.0118 | 0.8767±0.0421 |
| 13 | 39.2±1.3038 | 44 | 0.8909±0.0296 | 0.8264±0.0583 | 0.7250±0.1046 | 0.9278±0.0152 | 0.8195±0.0514 |
| 14 | 32.8±0.8367 | 37 | 0.8865±0.0226 | 0.8823±0.0449 | 0.8750±0.0884 | 0.8897±0.0154 | 0.8464±0.0327 |
| 15 | 27.6±0.5477 | 31 | 0.8903±0.0177 | 0.8583±0.0391 | 0.8000±0.0782 | 0.9167±0.0000 | 0.8470±0.0288 |
| 16 | 25.0±0.7071 | 28 | 0.8929±0.0253 | 0.8810±0.0505 | 0.8571±0.1010 | 0.9048±0.0000 | 0.8625±0.0364 |
| 17 | 21.8±0.4472 | 23 | 0.9478±0.0194 | 0.9539±0.0373 | 0.9667±0.0745 | 0.9412±0.0000 | 0.9346±0.0264 |
| 18 | 14.4±0.5477 | 16 | 0.9000±0.0342 | 0.8833±0.0685 | 0.8500±0.1369 | 0.9167±0.0000 | 0.8691±0.0490 |
| 19 | 10.2±0.4472 | 11 | 0.9273±0.0407 | 0.9429±0.0319 | 1.0000±0.0000 | 0.8857±0.0639 | 0.9248±0.0420 |
| 20 | 9.0±0.0000 | 10 | 0.9000±0.0000 | 0.9167±0.0000 | 1.0000±0.0000 | 0.8333±0.0000 | 0.8990±0.0000 |

Table 41: HM SVC with imputation, with missing flags, results of CV, taking best model of grid using F1. Predictions from admission.

| Day | Corrects | Totals | Accuracy | AUC | Sensitivity | Specificity | F1 |
|---|---|---|---|---|---|---|---|
| 1 | 183.4±1.6733 | 194 | 0.9454±0.0086 | 0.9430±0.0132 | 0.9394±0.0214 | 0.9466±0.0071 | 0.9102±0.0137 |
| 2 | 182.2±0.8367 | 194 | 0.9392±0.0043 | 0.9079±0.0078 | 0.8606±0.0166 | 0.9553±0.0052 | 0.8955±0.0069 |
| 3 | 173.4±0.5477 | 187 | 0.9273±0.0029 | 0.8812±0.0079 | 0.8133±0.0183 | 0.9490±0.0045 | 0.8692±0.0050 |
| 4 | 166.4±1.1402 | 180 | 0.9244±0.0063 | 0.8881±0.0082 | 0.8345±0.0154 | 0.9417±0.0073 | 0.8676±0.0098 |
| 5 | 144.0±1.4142 | 160 | 0.9000±0.0088 | 0.8513±0.0196 | 0.7778±0.0370 | 0.9248±0.0053 | 0.8314±0.0159 |
| 6 | 128.2±0.4472 | 142 | 0.9028±0.0031 | 0.8655±0.0143 | 0.8080±0.0335 | 0.9231±0.0060 | 0.8426±0.0069 |
| 7 | 104.6±0.5477 | 117 | 0.8940±0.0047 | 0.8519±0.0029 | 0.7826±0.0000 | 0.9213±0.0058 | 0.8385±0.0058 |
| 8 | 89.0±1.7321 | 100 | 0.8900±0.0173 | 0.8274±0.0322 | 0.7263±0.0577 | 0.9284±0.0103 | 0.8233±0.0292 |
| 9 | 70.2±1.3038 | 79 | 0.8886±0.0165 | 0.8556±0.0296 | 0.8000±0.0523 | 0.9111±0.0087 | 0.8363±0.0253 |
| 10 | 51.6±1.3416 | 62 | 0.8323±0.0216 | 0.7978±0.0273 | 0.7385±0.0421 | 0.8571±0.0204 | 0.7694±0.0274 |
| 11 | 45.8±1.0954 | 54 | 0.8481±0.0203 | 0.8131±0.0420 | 0.7500±0.0833 | 0.8762±0.0106 | 0.7927±0.0318 |
| 12 | 39.6±1.6733 | 50 | 0.7920±0.0335 | 0.7320±0.0531 | 0.6167±0.0950 | 0.8474±0.0220 | 0.7235±0.0474 |
| 13 | 34.2±0.8367 | 44 | 0.7773±0.0190 | 0.7083±0.0503 | 0.6000±0.1046 | 0.8167±0.0152 | 0.6749±0.0352 |
| 14 | 27.2±1.6432 | 37 | 0.7351±0.0444 | 0.7134±0.0680 | 0.6750±0.1118 | 0.7517±0.0289 | 0.6702±0.0567 |
| 15 | 22.2±0.8367 | 31 | 0.7161±0.0270 | 0.6446±0.0425 | 0.5143±0.0782 | 0.7750±0.0228 | 0.6289±0.0370 |
| 16 | 22.0±1.0000 | 28 | 0.7857±0.0357 | 0.7429±0.0593 | 0.6571±0.1278 | 0.8286±0.0426 | 0.7272±0.0519 |
| 17 | 17.8±0.4472 | 23 | 0.7739±0.0194 | 0.7176±0.0270 | 0.6000±0.0913 | 0.8353±0.0492 | 0.7117±0.0189 |
| 18 | 10.4±0.5477 | 16 | 0.6500±0.0342 | 0.6167±0.0543 | 0.5500±0.1118 | 0.6833±0.0373 | 0.5916±0.0409 |
| 19 | 7.8±0.4472 | 11 | 0.7091±0.0407 | 0.7071±0.0559 | 0.7000±0.1118 | 0.7143±0.0000 | 0.6958±0.0496 |
| 20 | 8.0±0.0000 | 10 | 0.8000±0.0000 | 0.7917±0.0000 | 0.7500±0.0000 | 0.8333±0.0000 | 0.7917±0.0000 |

Table 42: HM SVC with imputation, with missing flags, results of CV, taking best model of grid using F1. Predictions from outcome.

| Day | Corrects | Totals | Accuracy | AUC | Sensitivity | Specificity | F1 |
|---|---|---|---|---|---|---|---|
| 1 | 163.8±3.2711 | 194 | 0.8443±0.0169 | 0.7978±0.0245 | 0.7273±0.0371 | 0.8683±0.0135 | 0.7583±0.0240 |
| 2 | 167.6±1.1402 | 194 | 0.8639±0.0059 | 0.7686±0.0130 | 0.6242±0.0271 | 0.9130±0.0062 | 0.7635±0.0105 |
| 3 | 163.8±0.4472 | 187 | 0.8759±0.0024 | 0.7724±0.0069 | 0.6200±0.0183 | 0.9248±0.0053 | 0.7709±0.0035 |
| 4 | 160.4±1.6733 | 180 | 0.8911±0.0093 | 0.8014±0.0161 | 0.6690±0.0308 | 0.9338±0.0094 | 0.7997±0.0158 |
| 5 | 147.0±1.2247 | 160 | 0.9187±0.0077 | 0.8508±0.0160 | 0.7481±0.0310 | 0.9534±0.0063 | 0.8539±0.0141 |
| 6 | 127.2±2.1679 | 142 | 0.8958±0.0153 | 0.8298±0.0289 | 0.7280±0.0522 | 0.9316±0.0105 | 0.8236±0.0261 |
| 7 | 102.2±1.9235 | 117 | 0.8735±0.0164 | 0.8195±0.0129 | 0.7304±0.0194 | 0.9085±0.0207 | 0.8076±0.0193 |
| 8 | 86.4±0.5477 | 100 | 0.8640±0.0055 | 0.8194±0.0114 | 0.7474±0.0235 | 0.8914±0.0055 | 0.7950±0.0086 |
| 9 | 67.4±0.5477 | 79 | 0.8532±0.0069 | 0.7774±0.0171 | 0.6500±0.0342 | 0.9048±0.0000 | 0.7747±0.0134 |
| 10 | 52.6±1.1402 | 62 | 0.8484±0.0184 | 0.7797±0.0263 | 0.6615±0.0421 | 0.8980±0.0144 | 0.7751±0.0264 |
| 11 | 49.2±0.4472 | 54 | 0.9111±0.0083 | 0.8595±0.0186 | 0.7667±0.0373 | 0.9524±0.0000 | 0.8681±0.0138 |
| 12 | 43.8±0.4472 | 50 | 0.8760±0.0089 | 0.8272±0.0126 | 0.7333±0.0373 | 0.9211±0.0186 | 0.8290±0.0098 |
| 13 | 39.0±0.7071 | 44 | 0.8864±0.0161 | 0.8042±0.0338 | 0.6750±0.0685 | 0.9333±0.0152 | 0.8068±0.0281 |
| 14 | 32.4±0.5477 | 37 | 0.8757±0.0148 | 0.7759±0.0256 | 0.6000±0.0559 | 0.9517±0.0189 | 0.7992±0.0245 |
| 15 | 26.4±0.5477 | 31 | 0.8516±0.0177 | 0.7827±0.0363 | 0.6571±0.0782 | 0.9083±0.0186 | 0.7849±0.0295 |
| 16 | 22.8±0.4472 | 28 | 0.8143±0.0160 | 0.7333±0.0106 | 0.5714±0.0000 | 0.8952±0.0213 | 0.7425±0.0158 |
| 17 | 20.8±0.4472 | 23 | 0.9043±0.0194 | 0.8167±0.0373 | 0.6333±0.0745 | 1.0000±0.0000 | 0.8563±0.0355 |
| 18 | 13.0±0.7071 | 16 | 0.8125±0.0442 | 0.7083±0.0884 | 0.5000±0.1768 | 0.9167±0.0000 | 0.7200±0.0865 |
| 19 | 10.2±0.8367 | 11 | 0.9273±0.0761 | 0.9321±0.0803 | 0.9500±0.1118 | 0.9143±0.0782 | 0.9231±0.0817 |
| 20 | 8.0±0.7071 | 10 | 0.8000±0.0707 | 0.7917±0.0884 | 0.7500±0.1768 | 0.8333±0.0000 | 0.7889±0.0809 |

Table 43: HM SVC with imputation, with missing flags, applying reference values, results of CV, taking best model of grid using accuracy. Predictions from admission.

| Day | Corrects | Totals | Accuracy | AUC | Sensitivity | Specificity | F1 |
|---|---|---|---|---|---|---|---|
| 1 | 181.8±1.6432 | 194 | 0.9371±0.0085 | 0.9163±0.0206 | 0.8848±0.0395 | 0.9478±0.0034 | 0.8943±0.0154 |
| 2 | 181.8±2.1679 | 194 | 0.9371±0.0112 | 0.8971±0.0199 | 0.8364±0.0346 | 0.9578±0.0081 | 0.8905±0.0193 |
| 3 | 170.2±0.4472 | 187 | 0.9102±0.0024 | 0.8413±0.0112 | 0.7400±0.0279 | 0.9427±0.0064 | 0.8358±0.0047 |
| 4 | 165.0±1.7321 | 180 | 0.9167±0.0096 | 0.8389±0.0185 | 0.7241±0.0345 | 0.9536±0.0081 | 0.8437±0.0179 |
| 5 | 143.6±2.0736 | 160 | 0.8975±0.0130 | 0.8262±0.0243 | 0.7185±0.0422 | 0.9338±0.0082 | 0.8204±0.0230 |
| 6 | 121.8±1.3038 | 142 | 0.8577±0.0092 | 0.7533±0.0182 | 0.5920±0.0335 | 0.9145±0.0060 | 0.7540±0.0167 |
| 7 | 102.6±1.5166 | 117 | 0.8769±0.0130 | 0.7756±0.0278 | 0.6087±0.0532 | 0.9426±0.0058 | 0.7923±0.0247 |
| 8 | 86.2±1.6432 | 100 | 0.8620±0.0164 | 0.7698±0.0421 | 0.6211±0.0865 | 0.9185±0.0110 | 0.7721±0.0331 |
| 9 | 67.2±1.6432 | 79 | 0.8506±0.0208 | 0.7805±0.0215 | 0.6625±0.0342 | 0.8984±0.0241 | 0.7744±0.0266 |
| 10 | 49.8±1.3038 | 62 | 0.8032±0.0210 | 0.7625±0.0412 | 0.6923±0.0769 | 0.8327±0.0091 | 0.7325±0.0323 |
| 11 | 44.6±0.8944 | 54 | 0.8259±0.0166 | 0.7393±0.0106 | 0.5833±0.0000 | 0.8952±0.0213 | 0.7439±0.0176 |
| 12 | 38.2±0.4472 | 50 | 0.7640±0.0089 | 0.6908±0.0188 | 0.5500±0.0456 | 0.8316±0.0144 | 0.6850±0.0148 |
| 13 | 35.2±1.3038 | 44 | 0.8000±0.0296 | 0.6931±0.0375 | 0.5250±0.0559 | 0.8611±0.0278 | 0.6826±0.0392 |
| 14 | 30.2±0.4472 | 37 | 0.8162±0.0121 | 0.7289±0.0431 | 0.5750±0.1118 | 0.8828±0.0308 | 0.7265±0.0321 |
| 15 | 23.2±0.8367 | 31 | 0.7484±0.0270 | 0.6351±0.0174 | 0.4286±0.0000 | 0.8417±0.0349 | 0.6370±0.0231 |
| 16 | 22.6±1.1402 | 28 | 0.8071±0.0407 | 0.7571±0.0391 | 0.6571±0.0782 | 0.8571±0.0583 | 0.7500±0.0430 |
| 17 | 16.4±0.5477 | 23 | 0.7130±0.0238 | 0.6441±0.0161 | 0.5000±0.0000 | 0.7882±0.0322 | 0.6395±0.0203 |
| 18 | 7.8±0.8367 | 16 | 0.4875±0.0523 | 0.4417±0.0757 | 0.3500±0.1369 | 0.5333±0.0456 | 0.4306±0.0601 |
| 19 | 7.6±0.5477 | 11 | 0.6909±0.0498 | 0.6821±0.0685 | 0.6500±0.1369 | 0.7143±0.0000 | 0.6736±0.0607 |
| 20 | 8.2±0.8367 | 10 | 0.8200±0.0837 | 0.8417±0.0854 | 0.9500±0.1118 | 0.7333±0.0913 | 0.8190±0.0843 |

Table 44: HM SVC with imputation, with missing flags, applying reference values, results of CV, taking best model of grid using accuracy. Predictions from outcome.

| Day | Corrects | Totals | Accuracy | AUC | Sensitivity | Specificity | F1 |
|---|---|---|---|---|---|---|---|
| 1 | 164.8±2.7749 | 194 | 0.8495±0.0143 | 0.8202±0.0143 | 0.7758±0.0166 | 0.8646±0.0148 | 0.7712±0.0177 |
| 2 | 166.6±2.0736 | 194 | 0.8588±0.0107 | 0.7631±0.0163 | 0.6182±0.0271 | 0.9081±0.0092 | 0.7563±0.0169 |
| 3 | 163.4±1.3416 | 187 | 0.8738±0.0072 | 0.7711±0.0142 | 0.6200±0.0298 | 0.9223±0.0083 | 0.7682±0.0121 |
| 4 | 160.4±0.5477 | 180 | 0.8911±0.0030 | 0.8097±0.0088 | 0.6897±0.0244 | 0.9298±0.0076 | 0.8029±0.0033 |
| 5 | 147.6±1.3416 | 160 | 0.9225±0.0084 | 0.8707±0.0273 | 0.7926±0.0562 | 0.9489±0.0034 | 0.8639±0.0173 |
| 6 | 127.0±1.8708 | 142 | 0.8944±0.0132 | 0.8321±0.0249 | 0.7360±0.0456 | 0.9282±0.0097 | 0.8228±0.0222 |
| 7 | 102.2±2.1679 | 117 | 0.8735±0.0185 | 0.8293±0.0244 | 0.7565±0.0389 | 0.9021±0.0175 | 0.8108±0.0251 |
| 8 | 87.0±1.2247 | 100 | 0.8700±0.0122 | 0.8311±0.0174 | 0.7684±0.0288 | 0.8938±0.0110 | 0.8048±0.0174 |
| 9 | 67.4±0.5477 | 79 | 0.8532±0.0069 | 0.7774±0.0171 | 0.6500±0.0342 | 0.9048±0.0000 | 0.7747±0.0134 |
| 10 | 53.0±0.7071 | 62 | 0.8548±0.0114 | 0.8008±0.0300 | 0.7077±0.0644 | 0.8939±0.0091 | 0.7887±0.0214 |
| 11 | 48.4±0.5477 | 54 | 0.8963±0.0101 | 0.8440±0.0257 | 0.7500±0.0589 | 0.9381±0.0130 | 0.8478±0.0173 |
| 12 | 43.6±0.5477 | 50 | 0.8720±0.0110 | 0.8246±0.0181 | 0.7333±0.0373 | 0.9158±0.0118 | 0.8245±0.0156 |
| 13 | 39.0±0.7071 | 44 | 0.8864±0.0161 | 0.8236±0.0301 | 0.7250±0.0559 | 0.9222±0.0124 | 0.8142±0.0274 |
| 14 | 32.6±0.5477 | 37 | 0.8811±0.0148 | 0.7884±0.0094 | 0.6250±0.0000 | 0.9517±0.0189 | 0.8107±0.0180 |
| 15 | 26.6±0.5477 | 31 | 0.8581±0.0177 | 0.7970±0.0310 | 0.6857±0.0639 | 0.9083±0.0186 | 0.7966±0.0271 |
| 16 | 22.8±0.4472 | 28 | 0.8143±0.0160 | 0.7333±0.0106 | 0.5714±0.0000 | 0.8952±0.0213 | 0.7425±0.0158 |
| 17 | 21.0±0.0000 | 23 | 0.9130±0.0000 | 0.8333±0.0000 | 0.6667±0.0000 | 1.0000±0.0000 | 0.8722±0.0000 |
| 18 | 13.0±0.7071 | 16 | 0.8125±0.0442 | 0.7083±0.0884 | 0.5000±0.1768 | 0.9167±0.0000 | 0.7200±0.0865 |
| 19 | 10.2±0.8367 | 11 | 0.9273±0.0761 | 0.9321±0.0803 | 0.9500±0.1118 | 0.9143±0.0782 | 0.9231±0.0817 |
| 20 | 8.0±0.7071 | 10 | 0.8000±0.0707 | 0.7917±0.0884 | 0.7500±0.1768 | 0.8333±0.0000 | 0.7889±0.0809 |

Table 45: HM SVC with imputation, with missing flags, applying reference values, results of CV, taking best model of grid using F1. Predictions from admission.

| Day | Corrects | Totals | Accuracy | AUC | Sensitivity | Specificity | F1 |
|---|---|---|---|---|---|---|---|
| 1 | 181.2±1.4832 | 194 | 0.9340±0.0076 | 0.9121±0.0192 | 0.8788±0.0371 | 0.9453±0.0028 | 0.8893±0.0140 |
| 2 | 181.4±1.3416 | 194 | 0.9351±0.0069 | 0.8982±0.0142 | 0.8424±0.0254 | 0.9540±0.0034 | 0.8879±0.0123 |
| 3 | 169.6±1.1402 | 187 | 0.9070±0.0061 | 0.8421±0.0148 | 0.7467±0.0298 | 0.9376±0.0053 | 0.8322±0.0116 |
| 4 | 165.6±1.5166 | 180 | 0.9200±0.0084 | 0.8520±0.0163 | 0.7517±0.0289 | 0.9523±0.0055 | 0.8520±0.0157 |
| 5 | 143.4±1.8166 | 160 | 0.8962±0.0114 | 0.8254±0.0142 | 0.7185±0.0203 | 0.9323±0.0106 | 0.8190±0.0175 |
| 6 | 122.6±1.5166 | 142 | 0.8634±0.0107 | 0.7787±0.0315 | 0.6480±0.0657 | 0.9094±0.0076 | 0.7705±0.0223 |
| 7 | 102.8±1.3038 | 117 | 0.8786±0.0111 | 0.7833±0.0252 | 0.6261±0.0496 | 0.9404±0.0058 | 0.7974±0.0216 |
| 8 | 86.8±1.3038 | 100 | 0.8680±0.0130 | 0.7977±0.0353 | 0.6842±0.0744 | 0.9111±0.0103 | 0.7899±0.0255 |
| 9 | 67.2±0.4472 | 79 | 0.8506±0.0057 | 0.7898±0.0169 | 0.6875±0.0442 | 0.8921±0.0133 | 0.7777±0.0094 |
| 10 | 49.6±1.3416 | 62 | 0.8000±0.0216 | 0.7661±0.0372 | 0.7077±0.0644 | 0.8245±0.0112 | 0.7320±0.0309 |
| 11 | 45.0±1.0000 | 54 | 0.8333±0.0185 | 0.7500±0.0179 | 0.6000±0.0373 | 0.9000±0.0261 | 0.7547±0.0207 |
| 12 | 38.8±1.0954 | 50 | 0.7760±0.0219 | 0.7158±0.0371 | 0.6000±0.0697 | 0.8316±0.0144 | 0.7055±0.0324 |
| 13 | 35.2±1.3038 | 44 | 0.8000±0.0296 | 0.6931±0.0375 | 0.5250±0.0559 | 0.8611±0.0278 | 0.6826±0.0392 |
| 14 | 30.6±0.8944 | 37 | 0.8270±0.0242 | 0.7539±0.0406 | 0.6250±0.0884 | 0.8828±0.0308 | 0.7486±0.0338 |
| 15 | 23.0±0.7071 | 31 | 0.7419±0.0228 | 0.6310±0.0147 | 0.4286±0.0000 | 0.8333±0.0295 | 0.6313±0.0195 |
| 16 | 22.2±0.8367 | 28 | 0.7929±0.0299 | 0.7571±0.0310 | 0.6857±0.0639 | 0.8286±0.0426 | 0.7402±0.0317 |
| 17 | 16.2±0.4472 | 23 | 0.7043±0.0194 | 0.6382±0.0132 | 0.5000±0.0000 | 0.7765±0.0263 | 0.6321±0.0166 |
| 18 | 7.8±0.8367 | 16 | 0.4875±0.0523 | 0.4417±0.0757 | 0.3500±0.1369 | 0.5333±0.0456 | 0.4306±0.0601 |
| 19 | 7.4±0.5477 | 11 | 0.6727±0.0498 | 0.6571±0.0685 | 0.6000±0.1369 | 0.7143±0.0000 | 0.6515±0.0607 |
| 20 | 8.2±0.8367 | 10 | 0.8200±0.0837 | 0.8417±0.0854 | 0.9500±0.1118 | 0.7333±0.0913 | 0.8190±0.0843 |

Table 46: HM SVC with imputation, with missing flags, applying reference values, results of CV, taking best model of grid using F1. Predictions from outcome.

| Day | Corrects | Totals | Accuracy | AUC | Sensitivity | Specificity | F1 |
|---|---|---|---|---|---|---|---|
| 1 | 165.6±2.7019 | 194 | 0.8536±0.0139 | 0.7769±0.0390 | 0.6606±0.0784 | 0.8932±0.0068 | 0.7572±0.0292 |
| 2 | 168.4±2.9665 | 194 | 0.8680±0.0153 | 0.7519±0.0152 | 0.5758±0.0214 | 0.9280±0.0168 | 0.7596±0.0211 |
| 3 | 165.0±1.5811 | 187 | 0.8824±0.0085 | 0.7466±0.0198 | 0.5467±0.0380 | 0.9465±0.0057 | 0.7647±0.0185 |
| 4 | 161.2±1.6432 | 180 | 0.8956±0.0091 | 0.7789±0.0129 | 0.6069±0.0189 | 0.9510±0.0076 | 0.7953±0.0160 |
| 5 | 147.0±1.0000 | 160 | 0.9187±0.0063 | 0.8183±0.0123 | 0.6667±0.0262 | 0.9699±0.0075 | 0.8434±0.0114 |
| 6 | 129.0±1.0000 | 142 | 0.9085±0.0070 | 0.8186±0.0191 | 0.6800±0.0400 | 0.9573±0.0060 | 0.8341±0.0147 |
| 7 | 104.6±1.6733 | 117 | 0.8940±0.0143 | 0.8093±0.0168 | 0.6696±0.0238 | 0.9489±0.0139 | 0.8242±0.0213 |
| 8 | 86.4±1.5166 | 100 | 0.8640±0.0152 | 0.7670±0.0196 | 0.6105±0.0288 | 0.9235±0.0135 | 0.7737±0.0224 |
| 9 | 68.4±0.5477 | 79 | 0.8658±0.0069 | 0.7620±0.0171 | 0.5875±0.0342 | 0.9365±0.0000 | 0.7783±0.0145 |
| 10 | 51.4±0.8944 | 62 | 0.8290±0.0144 | 0.7166±0.0211 | 0.5231±0.0344 | 0.9102±0.0112 | 0.7279±0.0225 |
| 11 | 48.6±1.1402 | 54 | 0.9000±0.0211 | 0.8286±0.0396 | 0.7000±0.0745 | 0.9571±0.0106 | 0.8463±0.0360 |
| 12 | 42.4±0.5477 | 50 | 0.8480±0.0110 | 0.7404±0.0228 | 0.5333±0.0456 | 0.9474±0.0000 | 0.7656±0.0214 |
| 13 | 36.6±1.1402 | 44 | 0.8318±0.0259 | 0.6736±0.0804 | 0.4250±0.1677 | 0.9222±0.0124 | 0.6832±0.0758 |
| 14 | 29.6±0.5477 | 37 | 0.8000±0.0148 | 0.6552±0.0256 | 0.4000±0.0559 | 0.9103±0.0189 | 0.6699±0.0246 |
| 15 | 24.8±0.8367 | 31 | 0.8000±0.0270 | 0.6786±0.0711 | 0.4571±0.1565 | 0.9000±0.0228 | 0.6859±0.0678 |
| 16 | 22.0±0.7071 | 28 | 0.7857±0.0253 | 0.6667±0.0505 | 0.4286±0.1010 | 0.9048±0.0000 | 0.6798±0.0515 |
| 17 | 21.0±0.7071 | 23 | 0.9130±0.0307 | 0.8441±0.0425 | 0.7000±0.0745 | 0.9882±0.0263 | 0.8755±0.0425 |
| 18 | 14.8±0.4472 | 16 | 0.9250±0.0280 | 0.8667±0.0186 | 0.7500±0.0000 | 0.9833±0.0373 | 0.8935±0.0336 |
| 19 | 9.4±0.5477 | 11 | 0.8545±0.0498 | 0.8536±0.0685 | 0.8500±0.1369 | 0.8571±0.0000 | 0.8445±0.0561 |
| 20 | 8.2±0.4472 | 10 | 0.8200±0.0447 | 0.8167±0.0559 | 0.8000±0.1118 | 0.8333±0.0000 | 0.8131±0.0480 |

Table 47: HM SVC with imputation, applying reference values, results of CV, taking best model of grid using accuracy. Predictions from admission.

| Day | Corrects | Totals | Accuracy | AUC | Sensitivity | Specificity | F1 |
|---|---|---|---|---|---|---|---|
| 1 | 182.8±1.6432 | 194 | 0.9423±0.0085 | 0.9170±0.0124 | 0.8788±0.0214 | 0.9553±0.0081 | 0.9016±0.0138 |
| 2 | 179.8±1.6432 | 194 | 0.9268±0.0085 | 0.8571±0.0205 | 0.7515±0.0395 | 0.9627±0.0044 | 0.8667±0.0168 |
| 3 | 172.0±0.7071 | 187 | 0.9198±0.0038 | 0.8390±0.0077 | 0.7200±0.0183 | 0.9580±0.0057 | 0.8474±0.0061 |
| 4 | 163.8±1.7889 | 180 | 0.9100±0.0099 | 0.8238±0.0172 | 0.6966±0.0378 | 0.9510±0.0129 | 0.8303±0.0161 |
| 5 | 143.4±0.5477 | 160 | 0.8962±0.0034 | 0.7871±0.0210 | 0.6222±0.0483 | 0.9519±0.0067 | 0.8035±0.0127 |
| 6 | 122.2±1.0954 | 142 | 0.8606±0.0077 | 0.7392±0.0106 | 0.5520±0.0179 | 0.9265±0.0076 | 0.7493±0.0120 |
| 7 | 102.4±0.5477 | 117 | 0.8752±0.0047 | 0.7778±0.0163 | 0.6174±0.0364 | 0.9383±0.0048 | 0.7918±0.0117 |
| 8 | 83.4±0.8944 | 100 | 0.8340±0.0089 | 0.6800±0.0272 | 0.4316±0.0577 | 0.9284±0.0055 | 0.6981±0.0246 |
| 9 | 66.2±0.8367 | 79 | 0.8380±0.0106 | 0.6979±0.0162 | 0.4625±0.0342 | 0.9333±0.0133 | 0.7189±0.0166 |
| 10 | 49.8±1.9235 | 62 | 0.8032±0.0310 | 0.7003±0.0535 | 0.5231±0.1003 | 0.8776±0.0250 | 0.7006±0.0480 |
| 11 | 45.8±0.4472 | 54 | 0.8481±0.0083 | 0.7536±0.0053 | 0.5833±0.0000 | 0.9238±0.0106 | 0.7676±0.0091 |
| 12 | 41.0±0.7071 | 50 | 0.8200±0.0141 | 0.6934±0.0228 | 0.4500±0.0456 | 0.9368±0.0144 | 0.7163±0.0238 |
| 13 | 37.0±1.0000 | 44 | 0.8409±0.0227 | 0.6986±0.0258 | 0.4750±0.0559 | 0.9222±0.0304 | 0.7127±0.0300 |
| 14 | 29.4±0.8944 | 37 | 0.7946±0.0242 | 0.6427±0.0154 | 0.3750±0.0000 | 0.9103±0.0308 | 0.6583±0.0223 |
| 15 | 24.8±0.8367 | 31 | 0.8000±0.0270 | 0.6583±0.0352 | 0.4000±0.0639 | 0.9167±0.0295 | 0.6752±0.0415 |
| 16 | 22.0±1.0000 | 28 | 0.7857±0.0357 | 0.6571±0.0573 | 0.4000±0.1195 | 0.9143±0.0398 | 0.6710±0.0603 |
| 17 | 16.2±1.3038 | 23 | 0.7043±0.0567 | 0.5520±0.0664 | 0.2333±0.0913 | 0.8706±0.0492 | 0.5531±0.0774 |
| 18 | 8.4±0.5477 | 16 | 0.5250±0.0342 | 0.4667±0.0685 | 0.3500±0.1369 | 0.5833±0.0000 | 0.4570±0.0531 |
| 19 | 6.2±0.4472 | 11 | 0.5636±0.0407 | 0.5071±0.0559 | 0.3000±0.1118 | 0.7143±0.0000 | 0.5024±0.0586 |
| 20 | 7.8±0.4472 | 10 | 0.7800±0.0447 | 0.7667±0.0559 | 0.7000±0.1118 | 0.8333±0.0000 | 0.7674±0.0543 |

Table 48: HM SVC with imputation, applying reference values, results of CV, taking best model of grid using accuracy. Predictions from outcome.

| Day | Corrects | Totals | Accuracy | AUC | Sensitivity | Specificity | F1 |
|---|---|---|---|---|---|---|---|
| 1 | 166.0±3.3912 | 194 | 0.8557±0.0175 | 0.7974±0.0365 | 0.7091±0.0664 | 0.8857±0.0094 | 0.7679±0.0299 |
| 2 | 168.2±2.8636 | 194 | 0.8670±0.0148 | 0.7512±0.0178 | 0.5758±0.0303 | 0.9267±0.0161 | 0.7583±0.0217 |
| 3 | 164.8±0.8367 | 187 | 0.8813±0.0045 | 0.7648±0.0169 | 0.5933±0.0365 | 0.9363±0.0045 | 0.7727±0.0120 |
| 4 | 161.2±1.6432 | 180 | 0.8956±0.0091 | 0.7817±0.0098 | 0.6138±0.0154 | 0.9497±0.0100 | 0.7966±0.0141 |
| 5 | 147.0±2.0000 | 160 | 0.9188±0.0125 | 0.8301±0.0232 | 0.6963±0.0406 | 0.9639±0.0082 | 0.8474±0.0237 |
| 6 | 129.6±1.3416 | 142 | 0.9127±0.0094 | 0.8369±0.0160 | 0.7200±0.0283 | 0.9538±0.0076 | 0.8456±0.0163 |
| 7 | 105.2±0.8367 | 117 | 0.8991±0.0072 | 0.8321±0.0127 | 0.7217±0.0238 | 0.9426±0.0058 | 0.8376±0.0117 |
| 8 | 86.6±1.1402 | 100 | 0.8660±0.0114 | 0.7884±0.0239 | 0.6632±0.0471 | 0.9136±0.0087 | 0.7847±0.0197 |
| 9 | 69.2±0.8367 | 79 | 0.8759±0.0106 | 0.7917±0.0188 | 0.6500±0.0342 | 0.9333±0.0071 | 0.8013±0.0176 |
| 10 | 51.2±1.0954 | 62 | 0.8258±0.0177 | 0.7146±0.0214 | 0.5231±0.0344 | 0.9061±0.0183 | 0.7246±0.0242 |
| 11 | 48.6±0.5477 | 54 | 0.9000±0.0101 | 0.8345±0.0228 | 0.7167±0.0456 | 0.9524±0.0000 | 0.8486±0.0182 |
| 12 | 43.0±0.7071 | 50 | 0.8600±0.0141 | 0.7768±0.0228 | 0.6167±0.0456 | 0.9368±0.0144 | 0.7945±0.0214 |
| 13 | 37.6±0.8944 | 44 | 0.8545±0.0203 | 0.7458±0.0559 | 0.5750±0.1118 | 0.9167±0.0000 | 0.7487±0.0495 |
| 14 | 30.2±0.8367 | 37 | 0.8162±0.0226 | 0.6836±0.0528 | 0.4500±0.1118 | 0.9172±0.0189 | 0.6982±0.0477 |
| 15 | 25.6±0.8944 | 31 | 0.8258±0.0289 | 0.7357±0.0670 | 0.5714±0.1429 | 0.9000±0.0228 | 0.7398±0.0561 |
| 16 | 22.2±0.8367 | 28 | 0.7929±0.0299 | 0.6810±0.0598 | 0.4571±0.1195 | 0.9048±0.0000 | 0.6934±0.0603 |
| 17 | 20.8±0.8367 | 23 | 0.9043±0.0364 | 0.8382±0.0462 | 0.7000±0.0745 | 0.9765±0.0322 | 0.8653±0.0492 |
| 18 | 14.4±0.5477 | 16 | 0.9000±0.0342 | 0.8500±0.0228 | 0.7500±0.0000 | 0.9500±0.0456 | 0.8634±0.0412 |
| 19 | 9.4±0.5477 | 11 | 0.8545±0.0498 | 0.8536±0.0685 | 0.8500±0.1369 | 0.8571±0.0000 | 0.8445±0.0561 |
| 20 | 8.4±0.5477 | 10 | 0.8400±0.0548 | 0.8417±0.0685 | 0.8500±0.1369 | 0.8333±0.0000 | 0.8346±0.0588 |

Table 49: HM SVC with imputation, applying reference values, results of CV, taking best model of grid using F1. Predictions from admission.

| Day | Corrects | Totals | Accuracy | AUC | Sensitivity | Specificity | F1 |
|---|---|---|---|---|---|---|---|
| 1 | 182.4±1.8166 | 194 | 0.9402±0.0094 | 0.9182±0.0200 | 0.8848±0.0395 | 0.9516±0.0081 | 0.8989±0.0159 |
| 2 | 181.8±1.3038 | 194 | 0.9371±0.0067 | 0.8898±0.0205 | 0.8182±0.0429 | 0.9615±0.0052 | 0.8887±0.0135 |
| 3 | 172.4±2.0736 | 187 | 0.9219±0.0111 | 0.8537±0.0183 | 0.7533±0.0298 | 0.9541±0.0083 | 0.8548±0.0197 |
| 4 | 164.8±0.4472 | 180 | 0.9156±0.0025 | 0.8494±0.0060 | 0.7517±0.0154 | 0.9470±0.0047 | 0.8455±0.0036 |
| 5 | 145.2±1.3038 | 160 | 0.9075±0.0081 | 0.8233±0.0100 | 0.6963±0.0166 | 0.9504±0.0086 | 0.8312±0.0130 |
| 6 | 122.6±1.5166 | 142 | 0.8634±0.0107 | 0.7567±0.0088 | 0.5920±0.0179 | 0.9214±0.0140 | 0.7609±0.0128 |
| 7 | 103.0±0.0000 | 117 | 0.8803±0.0000 | 0.7909±0.0073 | 0.6435±0.0194 | 0.9383±0.0048 | 0.8026±0.0032 |
| 8 | 83.2±1.0954 | 100 | 0.8320±0.0110 | 0.6868±0.0241 | 0.4526±0.0471 | 0.9210±0.0068 | 0.7020±0.0231 |
| 9 | 66.4±1.6733 | 79 | 0.8405±0.0212 | 0.7182±0.0425 | 0.5125±0.0815 | 0.9238±0.0133 | 0.7331±0.0401 |
| 10 | 50.4±2.4083 | 62 | 0.8129±0.0388 | 0.7403±0.0615 | 0.6154±0.1088 | 0.8653±0.0310 | 0.7292±0.0550 |
| 11 | 45.2±0.4472 | 54 | 0.8370±0.0083 | 0.7464±0.0053 | 0.5833±0.0000 | 0.9095±0.0106 | 0.7555±0.0091 |
| 12 | 40.4±1.1402 | 50 | 0.8080±0.0228 | 0.6855±0.0225 | 0.4500±0.0456 | 0.9211±0.0322 | 0.7044±0.0269 |
| 13 | 37.0±1.4142 | 44 | 0.8409±0.0321 | 0.7083±0.0196 | 0.5000±0.0000 | 0.9167±0.0393 | 0.7204±0.0349 |
| 14 | 28.6±0.8944 | 37 | 0.7730±0.0242 | 0.6289±0.0154 | 0.3750±0.0000 | 0.8828±0.0308 | 0.6384±0.0221 |
| 15 | 24.4±0.8944 | 31 | 0.7871±0.0289 | 0.6601±0.0186 | 0.4286±0.0000 | 0.8917±0.0373 | 0.6721±0.0265 |
| 16 | 22.8±1.3038 | 28 | 0.8143±0.0466 | 0.6952±0.0661 | 0.4571±0.1195 | 0.9333±0.0426 | 0.7154±0.0731 |
| 17 | 16.2±0.8367 | 23 | 0.7043±0.0364 | 0.5735±0.0462 | 0.3000±0.0745 | 0.8471±0.0322 | 0.5772±0.0523 |
| 18 | 8.4±0.5477 | 16 | 0.5250±0.0342 | 0.4667±0.0685 | 0.3500±0.1369 | 0.5833±0.0000 | 0.4570±0.0531 |
| 19 | 6.2±0.4472 | 11 | 0.5636±0.0407 | 0.5071±0.0559 | 0.3000±0.1118 | 0.7143±0.0000 | 0.5024±0.0586 |
| 20 | 7.8±0.4472 | 10 | 0.7800±0.0447 | 0.7667±0.0559 | 0.7000±0.1118 | 0.8333±0.0000 | 0.7674±0.0543 |

Table 50: HM SVC with imputation, applying reference values, results of CV, taking best model of grid using F1. Predictions from outcome.

| Day | Corrects | Totals | Accuracy | AUC | Sensitivity | Specificity | F1 |
|---|---|---|---|---|---|---|---|
| 1 | 157.2±2.3875 | 194 | 0.8103±0.0123 | 0.7556±0.0049 | 0.6727±0.0136 | 0.8385±0.0170 | 0.7136±0.0108 |
| 2 | 167.2±1.6432 | 194 | 0.8619±0.0085 | 0.7698±0.0088 | 0.6303±0.0136 | 0.9093±0.0094 | 0.7623±0.0115 |
| 3 | 167.8±0.8367 | 187 | 0.8973±0.0045 | 0.8283±0.0063 | 0.7267±0.0149 | 0.9299±0.0064 | 0.8163±0.0062 |
| 4 | 160.6±0.8944 | 180 | 0.8922±0.0050 | 0.8327±0.0070 | 0.7448±0.0189 | 0.9205±0.0081 | 0.8125±0.0060 |
| 5 | 146.4±1.1402 | 160 | 0.9150±0.0071 | 0.8456±0.0186 | 0.7407±0.0370 | 0.9504±0.0041 | 0.8475±0.0143 |
| 6 | 131.2±1.3038 | 142 | 0.9239±0.0092 | 0.8784±0.0182 | 0.8080±0.0335 | 0.9487±0.0060 | 0.8713±0.0160 |
| 7 | 106.2±1.3038 | 117 | 0.9077±0.0111 | 0.8572±0.0205 | 0.7739±0.0364 | 0.9404±0.0058 | 0.8548±0.0183 |
| 8 | 86.6±1.8166 | 100 | 0.8660±0.0182 | 0.8206±0.0273 | 0.7474±0.0440 | 0.8938±0.0141 | 0.7974±0.0266 |
| 9 | 69.2±1.3038 | 79 | 0.8759±0.0165 | 0.8150±0.0226 | 0.7125±0.0342 | 0.9175±0.0133 | 0.8107±0.0240 |
| 10 | 53.2±1.6432 | 62 | 0.8581±0.0265 | 0.8198±0.0531 | 0.7538±0.1003 | 0.8857±0.0112 | 0.7982±0.0424 |
| 11 | 47.8±1.4832 | 54 | 0.8852±0.0275 | 0.8131±0.0409 | 0.6833±0.0697 | 0.9429±0.0213 | 0.8264±0.0421 |
| 12 | 43.4±1.5166 | 50 | 0.8680±0.0303 | 0.8048±0.0428 | 0.6833±0.0697 | 0.9263±0.0220 | 0.8136±0.0429 |
| 13 | 39.8±1.0954 | 44 | 0.9045±0.0249 | 0.8056±0.0333 | 0.6500±0.0559 | 0.9611±0.0248 | 0.8281±0.0385 |
| 14 | 31.6±0.8944 | 37 | 0.8541±0.0242 | 0.7802±0.0346 | 0.6500±0.0559 | 0.9103±0.0189 | 0.7828±0.0348 |
| 15 | 27.8±1.0954 | 31 | 0.8968±0.0353 | 0.8321±0.0582 | 0.7143±0.1010 | 0.9500±0.0186 | 0.8453±0.0547 |
| 16 | 23.2±0.4472 | 28 | 0.8286±0.0160 | 0.8095±0.0337 | 0.7714±0.0782 | 0.8476±0.0213 | 0.7862±0.0228 |
| 17 | 20.6±1.5166 | 23 | 0.8957±0.0659 | 0.8647±0.0876 | 0.8000±0.1394 | 0.9294±0.0492 | 0.8643±0.0859 |
| 18 | 13.6±0.8944 | 16 | 0.8500±0.0559 | 0.7833±0.0745 | 0.6500±0.1369 | 0.9167±0.0589 | 0.7923±0.0785 |
| 19 | 10.4±0.5477 | 11 | 0.9455±0.0498 | 0.9571±0.0391 | 1.0000±0.0000 | 0.9143±0.0782 | 0.9436±0.0515 |
| 20 | 8.4±0.5477 | 10 | 0.8400±0.0548 | 0.8417±0.0685 | 0.8500±0.1369 | 0.8333±0.0000 | 0.8346±0.0588 |

Table 51: HM SVC with missing flags, results of CV, taking best model of grid using accuracy. Predictions from admission.

| Day | Corrects | Totals | Accuracy | AUC | Sensitivity | Specificity | F1 |
|---|---|---|---|---|---|---|---|
| 1 | 176.8±1.3038 | 194 | 0.9113±0.0067 | 0.8888±0.0082 | 0.8545±0.0136 | 0.9230±0.0071 | 0.8559±0.0097 |
| 2 | 181.8±0.8367 | 194 | 0.9371±0.0043 | 0.8898±0.0099 | 0.8182±0.0214 | 0.9615±0.0052 | 0.8889±0.0074 |
| 3 | 172.2±1.7889 | 187 | 0.9209±0.0096 | 0.8612±0.0104 | 0.7733±0.0149 | 0.9490±0.0101 | 0.8556±0.0151 |
| 4 | 167.4±1.3416 | 180 | 0.9300±0.0075 | 0.8858±0.0160 | 0.8207±0.0289 | 0.9510±0.0036 | 0.8743±0.0139 |
| 5 | 144.8±0.4472 | 160 | 0.9050±0.0028 | 0.8632±0.0092 | 0.8000±0.0203 | 0.9263±0.0034 | 0.8408±0.0055 |
| 6 | 125.6±1.5166 | 142 | 0.8845±0.0107 | 0.8230±0.0260 | 0.7280±0.0522 | 0.9179±0.0076 | 0.8090±0.0197 |
| 7 | 102.8±1.6432 | 117 | 0.8786±0.0140 | 0.8062±0.0357 | 0.6870±0.0714 | 0.9255±0.0000 | 0.8066±0.0285 |
| 8 | 89.0±1.0000 | 100 | 0.8900±0.0100 | 0.8193±0.0285 | 0.7053±0.0600 | 0.9333±0.0068 | 0.8201±0.0203 |
| 9 | 66.8±2.0494 | 79 | 0.8456±0.0259 | 0.7913±0.0336 | 0.7000±0.0523 | 0.8825±0.0241 | 0.7746±0.0352 |
| 10 | 50.8±0.8367 | 62 | 0.8194±0.0135 | 0.7614±0.0233 | 0.6615±0.0421 | 0.8612±0.0091 | 0.7441±0.0203 |
| 11 | 46.4±0.8944 | 54 | 0.8593±0.0166 | 0.7964±0.0346 | 0.6833±0.0697 | 0.9095±0.0106 | 0.7959±0.0281 |
| 12 | 40.2±1.9235 | 50 | 0.8040±0.0385 | 0.7342±0.0364 | 0.6000±0.0373 | 0.8684±0.0416 | 0.7338±0.0440 |
| 13 | 34.2±0.4472 | 44 | 0.7773±0.0102 | 0.6208±0.0349 | 0.3750±0.0884 | 0.8667±0.0232 | 0.6201±0.0287 |
| 14 | 27.0±1.0000 | 37 | 0.7297±0.0270 | 0.6375±0.0539 | 0.4750±0.1046 | 0.8000±0.0154 | 0.6261±0.0455 |
| 15 | 22.6±0.5477 | 31 | 0.7290±0.0177 | 0.6327±0.0516 | 0.4571±0.1195 | 0.8083±0.0228 | 0.6247±0.0428 |
| 16 | 20.2±2.7749 | 28 | 0.7214±0.0991 | 0.6714±0.1230 | 0.5714±0.1750 | 0.7714±0.0782 | 0.6572±0.1198 |
| 17 | 18.4±0.8944 | 23 | 0.8000±0.0389 | 0.7245±0.0502 | 0.5667±0.0913 | 0.8824±0.0416 | 0.7313±0.0491 |
| 18 | 10.0±0.7071 | 16 | 0.6250±0.0442 | 0.6000±0.0475 | 0.5500±0.1118 | 0.6500±0.0697 | 0.5712±0.0383 |
| 19 | 8.0±0.7071 | 11 | 0.7273±0.0643 | 0.7214±0.0710 | 0.7000±0.1118 | 0.7429±0.0639 | 0.7129±0.0698 |
| 20 | 7.2±0.4472 | 10 | 0.7200±0.0447 | 0.7250±0.0373 | 0.7500±0.0000 | 0.7000±0.0745 | 0.7159±0.0423 |

Table 52: HM SVC with missing flags, results of CV, taking best model of grid using accuracy. Predictions from outcome.

| Day | Corrects | Totals | Accuracy | AUC | Sensitivity | Specificity | F1 |
|---|---|---|---|---|---|---|---|
| 1 | 156.6±2.3022 | 194 | 0.8072±0.0119 | 0.7730±0.0155 | 0.7212±0.0254 | 0.8248±0.0119 | 0.7184±0.0147 |
| 2 | 167.0±1.5811 | 194 | 0.8608±0.0082 | 0.7740±0.0142 | 0.6424±0.0254 | 0.9056±0.0068 | 0.7631±0.0134 |
| 3 | 166.8±0.8367 | 187 | 0.8920±0.0045 | 0.8305±0.0118 | 0.7400±0.0279 | 0.9210±0.0073 | 0.8110±0.0073 |
| 4 | 159.6±0.8944 | 180 | 0.8867±0.0050 | 0.8294±0.0085 | 0.7448±0.0189 | 0.9139±0.0066 | 0.8052±0.0072 |
| 5 | 146.8±1.3038 | 160 | 0.9175±0.0081 | 0.8618±0.0195 | 0.7778±0.0370 | 0.9459±0.0034 | 0.8554±0.0156 |
| 6 | 130.8±1.4832 | 142 | 0.9211±0.0104 | 0.8766±0.0188 | 0.8080±0.0335 | 0.9453±0.0076 | 0.8674±0.0177 |
| 7 | 106.2±0.8367 | 117 | 0.9077±0.0072 | 0.8703±0.0086 | 0.8087±0.0238 | 0.9319±0.0121 | 0.8585±0.0086 |
| 8 | 87.2±2.0494 | 100 | 0.8720±0.0205 | 0.8404±0.0316 | 0.7895±0.0526 | 0.8914±0.0161 | 0.8098±0.0295 |
| 9 | 69.8±0.8367 | 79 | 0.8835±0.0106 | 0.8244±0.0188 | 0.7250±0.0342 | 0.9238±0.0071 | 0.8213±0.0170 |
| 10 | 54.2±1.3038 | 62 | 0.8742±0.0210 | 0.8526±0.0276 | 0.8154±0.0421 | 0.8898±0.0183 | 0.8246±0.0281 |
| 11 | 48.0±1.0000 | 54 | 0.8889±0.0185 | 0.8274±0.0276 | 0.7167±0.0456 | 0.9381±0.0130 | 0.8353±0.0277 |
| 12 | 45.2±1.6432 | 50 | 0.9040±0.0329 | 0.8684±0.0592 | 0.8000±0.1118 | 0.9368±0.0144 | 0.8674±0.0466 |
| 13 | 40.0±1.4142 | 44 | 0.9091±0.0321 | 0.8375±0.0582 | 0.7250±0.1046 | 0.9500±0.0232 | 0.8438±0.0546 |
| 14 | 32.2±1.0954 | 37 | 0.8703±0.0296 | 0.8358±0.0537 | 0.7750±0.1046 | 0.8966±0.0244 | 0.8175±0.0439 |
| 15 | 28.6±0.8944 | 31 | 0.9226±0.0289 | 0.8893±0.0413 | 0.8286±0.0639 | 0.9500±0.0186 | 0.8893±0.0413 |
| 16 | 23.0±1.0000 | 28 | 0.8214±0.0357 | 0.8048±0.0488 | 0.7714±0.0782 | 0.8381±0.0261 | 0.7795±0.0439 |
| 17 | 20.6±1.5166 | 23 | 0.8957±0.0659 | 0.8647±0.0876 | 0.8000±0.1394 | 0.9294±0.0492 | 0.8643±0.0859 |
| 18 | 13.8±0.8367 | 16 | 0.8625±0.0523 | 0.8083±0.0632 | 0.7000±0.1118 | 0.9167±0.0589 | 0.8138±0.0700 |
| 19 | 10.2±0.4472 | 11 | 0.9273±0.0407 | 0.9429±0.0319 | 1.0000±0.0000 | 0.8857±0.0639 | 0.9248±0.0420 |
| 20 | 8.6±0.5477 | 10 | 0.8600±0.0548 | 0.8667±0.0685 | 0.9000±0.1369 | 0.8333±0.0000 | 0.8561±0.0588 |

Table 53: HM SVC with missing flags, results of CV, taking best model of grid using F1. Predictions from admission.

| Day | Corrects | Totals | Accuracy | AUC | Sensitivity | Specificity | F1 |
|---|---|---|---|---|---|---|---|
| 1 | 176.6±1.3416 | 194 | 0.9103±0.0069 | 0.8881±0.0085 | 0.8545±0.0136 | 0.9217±0.0071 | 0.8545±0.0100 |
| 2 | 182.4±1.5166 | 194 | 0.9402±0.0078 | 0.8965±0.0094 | 0.8303±0.0166 | 0.9627±0.0088 | 0.8947±0.0125 |
| 3 | 171.8±0.8367 | 187 | 0.9187±0.0045 | 0.8707±0.0119 | 0.8000±0.0236 | 0.9414±0.0028 | 0.8552±0.0087 |
| 4 | 167.6±2.4083 | 180 | 0.9311±0.0134 | 0.8949±0.0266 | 0.8414±0.0463 | 0.9483±0.0073 | 0.8779±0.0241 |
| 5 | 144.0±1.2247 | 160 | 0.9000±0.0077 | 0.8602±0.0175 | 0.8000±0.0331 | 0.9203±0.0041 | 0.8341±0.0136 |
| 6 | 127.4±1.8166 | 142 | 0.8972±0.0128 | 0.8621±0.0140 | 0.8080±0.0179 | 0.9162±0.0127 | 0.8356±0.0180 |
| 7 | 103.0±1.5811 | 117 | 0.8803±0.0135 | 0.8303±0.0219 | 0.7478±0.0364 | 0.9128±0.0089 | 0.8176±0.0207 |
| 8 | 89.2±1.6432 | 100 | 0.8920±0.0164 | 0.8366±0.0318 | 0.7474±0.0577 | 0.9259±0.0087 | 0.8284±0.0277 |
| 9 | 67.4±1.6733 | 79 | 0.8532±0.0212 | 0.8054±0.0339 | 0.7250±0.0559 | 0.8857±0.0133 | 0.7862±0.0311 |
| 10 | 50.2±1.6432 | 62 | 0.8097±0.0265 | 0.7779±0.0416 | 0.7231±0.0688 | 0.8327±0.0171 | 0.7439±0.0364 |
| 11 | 45.8±1.6432 | 54 | 0.8481±0.0304 | 0.7952±0.0433 | 0.7000±0.0745 | 0.8905±0.0271 | 0.7866±0.0416 |
| 12 | 40.4±2.1909 | 50 | 0.8080±0.0438 | 0.7425±0.0409 | 0.6167±0.0456 | 0.8684±0.0492 | 0.7409±0.0485 |
| 13 | 34.8±0.8367 | 44 | 0.7909±0.0190 | 0.6583±0.0507 | 0.4500±0.1118 | 0.8667±0.0232 | 0.6529±0.0460 |
| 14 | 27.0±1.0000 | 37 | 0.7297±0.0270 | 0.6647±0.0482 | 0.5500±0.1118 | 0.7793±0.0393 | 0.6419±0.0384 |
| 15 | 23.2±0.4472 | 31 | 0.7484±0.0144 | 0.6756±0.0501 | 0.5429±0.1195 | 0.8083±0.0228 | 0.6609±0.0349 |
| 16 | 20.8±1.0954 | 28 | 0.7429±0.0391 | 0.7333±0.0543 | 0.7143±0.1010 | 0.7524±0.0398 | 0.6975±0.0455 |
| 17 | 18.6±0.5477 | 23 | 0.8087±0.0238 | 0.7304±0.0420 | 0.5667±0.0913 | 0.8941±0.0263 | 0.7392±0.0367 |
| 18 | 9.8±0.8367 | 16 | 0.6125±0.0523 | 0.6083±0.0632 | 0.6000±0.1369 | 0.6167±0.0745 | 0.5688±0.0493 |
| 19 | 8.0±0.7071 | 11 | 0.7273±0.0643 | 0.7214±0.0710 | 0.7000±0.1118 | 0.7429±0.0639 | 0.7129±0.0698 |
| 20 | 7.2±0.4472 | 10 | 0.7200±0.0447 | 0.7250±0.0373 | 0.7500±0.0000 | 0.7000±0.0745 | 0.7159±0.0423 |

Table 54: HM SVC with missing flags, results of CV, taking best model of grid using F1. Predictions from outcome.

| Day | Corrects | Totals | Accuracy | AUC | Sensitivity | Specificity | F1 |
|---|---|---|---|---|---|---|---|
| 1 | 161.2±1.9235 | 194 | 0.8309±0.0099 | 0.7464±0.0253 | 0.6182±0.0507 | 0.8745±0.0068 | 0.7248±0.0190 |
| 2 | 167.2±2.1679 | 194 | 0.8619±0.0112 | 0.7361±0.0179 | 0.5455±0.0303 | 0.9267±0.0092 | 0.7455±0.0191 |
| 3 | 165.2±1.3038 | 187 | 0.8834±0.0070 | 0.7634±0.0133 | 0.5867±0.0380 | 0.9401±0.0140 | 0.7743±0.0076 |
| 4 | 163.0±1.0000 | 180 | 0.9056±0.0056 | 0.8156±0.0085 | 0.6828±0.0154 | 0.9483±0.0055 | 0.8218±0.0094 |
| 5 | 147.0±1.2247 | 160 | 0.9187±0.0077 | 0.8301±0.0121 | 0.6963±0.0310 | 0.9639±0.0124 | 0.8475±0.0113 |
| 6 | 129.6±1.1402 | 142 | 0.9127±0.0080 | 0.8275±0.0126 | 0.6960±0.0219 | 0.9590±0.0072 | 0.8425±0.0136 |
| 7 | 105.8±1.3038 | 117 | 0.9043±0.0111 | 0.8288±0.0124 | 0.7043±0.0194 | 0.9532±0.0121 | 0.8423±0.0160 |
| 8 | 86.2±1.3038 | 100 | 0.8620±0.0130 | 0.7658±0.0204 | 0.6105±0.0471 | 0.9210±0.0187 | 0.7711±0.0181 |
| 9 | 69.4±0.8944 | 79 | 0.8785±0.0113 | 0.7839±0.0071 | 0.6250±0.0000 | 0.9429±0.0142 | 0.8007±0.0138 |
| 10 | 52.0±0.7071 | 62 | 0.8387±0.0114 | 0.7397±0.0209 | 0.5692±0.0421 | 0.9102±0.0112 | 0.7478±0.0188 |
| 11 | 48.2±0.4472 | 54 | 0.8926±0.0083 | 0.8119±0.0053 | 0.6667±0.0000 | 0.9571±0.0106 | 0.8334±0.0105 |
| 12 | 42.4±0.8944 | 50 | 0.8480±0.0179 | 0.7575±0.0303 | 0.5833±0.0589 | 0.9316±0.0144 | 0.7752±0.0289 |
| 13 | 36.8±0.8367 | 44 | 0.8364±0.0190 | 0.6764±0.0363 | 0.4250±0.0685 | 0.9278±0.0152 | 0.6937±0.0374 |
| 14 | 30.6±1.1402 | 37 | 0.8270±0.0308 | 0.6724±0.0366 | 0.4000±0.0559 | 0.9448±0.0308 | 0.6982±0.0444 |
| 15 | 24.8±0.8367 | 31 | 0.8000±0.0270 | 0.6583±0.0598 | 0.4000±0.1195 | 0.9167±0.0000 | 0.6726±0.0607 |
| 16 | 22.2±0.4472 | 28 | 0.7929±0.0160 | 0.7000±0.0319 | 0.5143±0.0782 | 0.8857±0.0261 | 0.7084±0.0282 |
| 17 | 21.2±1.7889 | 23 | 0.9217±0.0778 | 0.8716±0.1211 | 0.7667±0.2236 | 0.9765±0.0526 | 0.8879±0.1087 |
| 18 | 14.8±0.4472 | 16 | 0.9250±0.0280 | 0.8667±0.0186 | 0.7500±0.0000 | 0.9833±0.0373 | 0.8935±0.0336 |
| 19 | 9.0±1.0000 | 11 | 0.8182±0.0909 | 0.8250±0.0887 | 0.8500±0.1369 | 0.8000±0.1278 | 0.8108±0.0915 |
| 20 | 8.0±0.0000 | 10 | 0.8000±0.0000 | 0.7917±0.0000 | 0.7500±0.0000 | 0.8333±0.0000 | 0.7917±0.0000 |

Table 55: HM SVC applying laboratory reference values, results of CV, taking best model of grid using accuracy. Predictions from admission.

| Day | Corrects | Totals | Accuracy | AUC | Sensitivity | Specificity | F1 |
|---|---|---|---|---|---|---|---|
| 1 | 161.2±1.9235 | 194 | 0.8309±0.0099 | 0.7464±0.0253 | 0.6182±0.0507 | 0.8745±0.0068 | 0.7248±0.0190 |
| 2 | 167.2±2.1679 | 194 | 0.8619±0.0112 | 0.7361±0.0179 | 0.5455±0.0303 | 0.9267±0.0092 | 0.7455±0.0191 |
| 3 | 165.2±1.3038 | 187 | 0.8834±0.0070 | 0.7634±0.0133 | 0.5867±0.0380 | 0.9401±0.0140 | 0.7743±0.0076 |
| 4 | 163.0±1.0000 | 180 | 0.9056±0.0056 | 0.8156±0.0085 | 0.6828±0.0154 | 0.9483±0.0055 | 0.8218±0.0094 |
| 5 | 147.0±1.2247 | 160 | 0.9187±0.0077 | 0.8301±0.0121 | 0.6963±0.0310 | 0.9639±0.0124 | 0.8475±0.0113 |
| 6 | 129.6±1.1402 | 142 | 0.9127±0.0080 | 0.8275±0.0126 | 0.6960±0.0219 | 0.9590±0.0072 | 0.8425±0.0136 |
| 7 | 105.8±1.3038 | 117 | 0.9043±0.0111 | 0.8288±0.0124 | 0.7043±0.0194 | 0.9532±0.0121 | 0.8423±0.0160 |
| 8 | 86.2±1.3038 | 100 | 0.8620±0.0130 | 0.7658±0.0204 | 0.6105±0.0471 | 0.9210±0.0187 | 0.7711±0.0181 |
| 9 | 69.4±0.8944 | 79 | 0.8785±0.0113 | 0.7839±0.0071 | 0.6250±0.0000 | 0.9429±0.0142 | 0.8007±0.0138 |
| 10 | 52.0±0.7071 | 62 | 0.8387±0.0114 | 0.7397±0.0209 | 0.5692±0.0421 | 0.9102±0.0112 | 0.7478±0.0188 |
| 11 | 48.2±0.4472 | 54 | 0.8926±0.0083 | 0.8119±0.0053 | 0.6667±0.0000 | 0.9571±0.0106 | 0.8334±0.0105 |
| 12 | 42.4±0.8944 | 50 | 0.8480±0.0179 | 0.7575±0.0303 | 0.5833±0.0589 | 0.9316±0.0144 | 0.7752±0.0289 |
| 13 | 36.8±0.8367 | 44 | 0.8364±0.0190 | 0.6764±0.0363 | 0.4250±0.0685 | 0.9278±0.0152 | 0.6937±0.0374 |
| 14 | 30.6±1.1402 | 37 | 0.8270±0.0308 | 0.6724±0.0366 | 0.4000±0.0559 | 0.9448±0.0308 | 0.6982±0.0444 |
| 15 | 24.8±0.8367 | 31 | 0.8000±0.0270 | 0.6583±0.0598 | 0.4000±0.1195 | 0.9167±0.0000 | 0.6726±0.0607 |
| 16 | 22.2±0.4472 | 28 | 0.7929±0.0160 | 0.7000±0.0319 | 0.5143±0.0782 | 0.8857±0.0261 | 0.7084±0.0282 |
| 17 | 21.2±1.7889 | 23 | 0.9217±0.0778 | 0.8716±0.1211 | 0.7667±0.2236 | 0.9765±0.0526 | 0.8879±0.1087 |
| 18 | 14.8±0.4472 | 16 | 0.9250±0.0280 | 0.8667±0.0186 | 0.7500±0.0000 | 0.9833±0.0373 | 0.8935±0.0336 |
| 19 | 9.0±1.0000 | 11 | 0.8182±0.0909 | 0.8250±0.0887 | 0.8500±0.1369 | 0.8000±0.1278 | 0.8108±0.0915 |
| 20 | 8.0±0.0000 | 10 | 0.8000±0.0000 | 0.7917±0.0000 | 0.7500±0.0000 | 0.8333±0.0000 | 0.7917±0.0000 |

Table 56: HM SVC applying laboratory reference values, results of CV, taking best model of grid using accuracy. Predictions from outcome.

| Day | Corrects | Totals | Accuracy | AUC | Sensitivity | Specificity | F1 |
|---|---|---|---|---|---|---|---|
| 1 | 164.2±2.7749 | 194 | 0.8464±0.0143 | 0.7942±0.0267 | 0.7152±0.0507 | 0.8733±0.0129 | 0.7585±0.0221 |
| 2 | 167.8±1.6432 | 194 | 0.8649±0.0085 | 0.7524±0.0172 | 0.5818±0.0332 | 0.9230±0.0071 | 0.7566±0.0155 |
| 3 | 165.2±1.3038 | 187 | 0.8834±0.0070 | 0.7904±0.0077 | 0.6533±0.0183 | 0.9274±0.0097 | 0.7866±0.0090 |
| 4 | 163.0±1.4142 | 180 | 0.9056±0.0079 | 0.8323±0.0047 | 0.7241±0.0000 | 0.9404±0.0094 | 0.8278±0.0111 |
| 5 | 147.0±1.5811 | 160 | 0.9187±0.0099 | 0.8449±0.0116 | 0.7333±0.0166 | 0.9564±0.0098 | 0.8523±0.0160 |
| 6 | 130.0±1.0000 | 142 | 0.9155±0.0070 | 0.8418±0.0101 | 0.7280±0.0179 | 0.9556±0.0072 | 0.8506±0.0113 |
| 7 | 105.0±1.2247 | 117 | 0.8974±0.0105 | 0.8475±0.0089 | 0.7652±0.0238 | 0.9298±0.0161 | 0.8409±0.0122 |
| 8 | 85.8±1.4832 | 100 | 0.8580±0.0148 | 0.7875±0.0166 | 0.6737±0.0235 | 0.9012±0.0151 | 0.7775±0.0200 |
| 9 | 69.4±0.5477 | 79 | 0.8785±0.0069 | 0.7933±0.0104 | 0.6500±0.0342 | 0.9365±0.0159 | 0.8045±0.0064 |
| 10 | 51.8±0.4472 | 62 | 0.8355±0.0072 | 0.7546±0.0046 | 0.6154±0.0000 | 0.8939±0.0091 | 0.7533±0.0076 |
| 11 | 48.6±0.5477 | 54 | 0.9000±0.0101 | 0.8286±0.0212 | 0.7000±0.0456 | 0.9571±0.0106 | 0.8467±0.0169 |
| 12 | 43.2±0.8367 | 50 | 0.8640±0.0167 | 0.7908±0.0226 | 0.6500±0.0373 | 0.9316±0.0144 | 0.8044±0.0239 |
| 13 | 38.8±1.0954 | 44 | 0.8818±0.0249 | 0.7917±0.0545 | 0.6500±0.1046 | 0.9333±0.0152 | 0.7963±0.0477 |
| 14 | 31.6±1.1402 | 37 | 0.8541±0.0308 | 0.7349±0.0366 | 0.5250±0.0559 | 0.9448±0.0308 | 0.7601±0.0434 |
| 15 | 26.2±1.3038 | 31 | 0.8452±0.0421 | 0.7583±0.0931 | 0.6000±0.1863 | 0.9167±0.0000 | 0.7640±0.0782 |
| 16 | 21.6±0.8944 | 28 | 0.7714±0.0319 | 0.6857±0.0639 | 0.5143±0.1278 | 0.8571±0.0000 | 0.6870±0.0554 |
| 17 | 21.2±2.0494 | 23 | 0.9217±0.0891 | 0.8931±0.1348 | 0.8333±0.2357 | 0.9529±0.0492 | 0.8931±0.1244 |
| 18 | 13.8±0.4472 | 16 | 0.8625±0.0280 | 0.8250±0.0186 | 0.7500±0.0000 | 0.9000±0.0373 | 0.8203±0.0292 |
| 19 | 9.6±0.8944 | 11 | 0.8727±0.0813 | 0.9000±0.0639 | 1.0000±0.0000 | 0.8000±0.1278 | 0.8712±0.0817 |
| 20 | 8.0±0.0000 | 10 | 0.8000±0.0000 | 0.7917±0.0000 | 0.7500±0.0000 | 0.8333±0.0000 | 0.7917±0.0000 |

Table 57: HM SVC applying laboratory reference values, results of CV, taking best model of grid using F1. Predictions from admission.

| Day | Corrects | Totals | Accuracy | AUC | Sensitivity | Specificity | F1 |
|---|---|---|---|---|---|---|---|
| 1 | 175.0±1.7321 | 194 | 0.9021±0.0089 | 0.8735±0.0182 | 0.8303±0.0346 | 0.9168±0.0071 | 0.8410±0.0147 |
| 2 | 179.2±2.1679 | 194 | 0.9237±0.0112 | 0.8697±0.0212 | 0.7879±0.0371 | 0.9516±0.0068 | 0.8661±0.0200 |
| 3 | 171.2±1.3038 | 187 | 0.9155±0.0070 | 0.8445±0.0096 | 0.7400±0.0149 | 0.9490±0.0064 | 0.8437±0.0117 |
| 4 | 165.6±1.6733 | 180 | 0.9200±0.0093 | 0.8632±0.0201 | 0.7793±0.0393 | 0.9470±0.0081 | 0.8552±0.0171 |
| 5 | 149.0±1.4142 | 160 | 0.9313±0.0088 | 0.8701±0.0139 | 0.7778±0.0262 | 0.9624±0.0092 | 0.8757±0.0150 |
| 6 | 125.4±2.3022 | 142 | 0.8831±0.0162 | 0.8095±0.0274 | 0.6960±0.0537 | 0.9231±0.0171 | 0.8028±0.0252 |
| 7 | 105.4±0.8944 | 117 | 0.9009±0.0076 | 0.8168±0.0190 | 0.6783±0.0389 | 0.9553±0.0048 | 0.8339±0.0154 |
| 8 | 89.6±1.5166 | 100 | 0.8960±0.0152 | 0.8069±0.0306 | 0.6632±0.0600 | 0.9506±0.0123 | 0.8220±0.0276 |
| 9 | 67.4±0.8944 | 79 | 0.8532±0.0113 | 0.7727±0.0241 | 0.6375±0.0523 | 0.9079±0.0133 | 0.7724±0.0193 |
| 10 | 51.0±0.7071 | 62 | 0.8226±0.0114 | 0.7860±0.0209 | 0.7231±0.0421 | 0.8490±0.0112 | 0.7569±0.0162 |
| 11 | 45.6±1.6733 | 54 | 0.8444±0.0310 | 0.7571±0.0254 | 0.6000±0.0373 | 0.9143±0.0398 | 0.7674±0.0346 |
| 12 | 38.8±0.4472 | 50 | 0.7760±0.0089 | 0.6816±0.0059 | 0.5000±0.0000 | 0.8632±0.0118 | 0.6858±0.0082 |
| 13 | 36.2±1.0954 | 44 | 0.8227±0.0249 | 0.6875±0.0333 | 0.4750±0.0559 | 0.9000±0.0248 | 0.6933±0.0368 |
| 14 | 27.0±1.0000 | 37 | 0.7297±0.0270 | 0.6194±0.0361 | 0.4250±0.0685 | 0.8138±0.0308 | 0.6146±0.0338 |
| 15 | 23.4±0.8944 | 31 | 0.7548±0.0289 | 0.6292±0.0327 | 0.4000±0.0639 | 0.8583±0.0373 | 0.6339±0.0368 |
| 16 | 22.4±0.8944 | 28 | 0.8000±0.0319 | 0.7048±0.0621 | 0.5143±0.1278 | 0.8952±0.0213 | 0.7134±0.0630 |
| 17 | 16.4±0.5477 | 23 | 0.7130±0.0238 | 0.6010±0.0363 | 0.3667±0.0745 | 0.8353±0.0263 | 0.6048±0.0353 |
| 18 | 9.0±0.0000 | 16 | 0.5625±0.0000 | 0.4583±0.0000 | 0.2500±0.0000 | 0.6667±0.0000 | 0.4589±0.0000 |
| 19 | 7.0±0.7071 | 11 | 0.6364±0.0643 | 0.6071±0.0884 | 0.5000±0.1768 | 0.7143±0.0000 | 0.6031±0.0857 |
| 20 | 7.2±0.4472 | 10 | 0.7200±0.0447 | 0.6917±0.0559 | 0.5500±0.1118 | 0.8333±0.0000 | 0.6946±0.0543 |

Table 58: HM SVC applying laboratory reference values, results of CV, taking best model of grid using F1. Predictions from outcome.

##### 8.5.3 Ensemble

| Day | Corrects | Totals | Accuracy | AUC | Sensitivity | Specificity | F1 |
|---|---|---|---|---|---|---|---|
| 1 | 166.2±1.7889 | 194 | 0.8567±0.0092 | 0.7599±0.0245 | 0.4667±0.0460 | 0.9366±0.0052 | 0.7203±0.0221 |
| 2 | 167.2±0.4472 | 194 | 0.8619±0.0023 | 0.7790±0.0235 | 0.5576±0.0346 | 0.9242±0.0068 | 0.7478±0.0085 |
| 3 | 166.4±0.8944 | 187 | 0.8898±0.0048 | 0.8469±0.0035 | 0.6467±0.0298 | 0.9363±0.0064 | 0.7938±0.0097 |
| 4 | 165.4±1.3416 | 180 | 0.9189±0.0075 | 0.8799±0.0086 | 0.7172±0.0663 | 0.9576±0.0166 | 0.8458±0.0131 |
| 5 | 146.4±1.1402 | 160 | 0.9150±0.0071 | 0.9119±0.0064 | 0.7259±0.0422 | 0.9534±0.0124 | 0.8457±0.0114 |
| 6 | 130.2±1.7889 | 142 | 0.9169±0.0126 | 0.9354±0.0216 | 0.7520±0.0179 | 0.9521±0.0155 | 0.8558±0.0183 |
| 7 | 109.2±1.0954 | 117 | 0.9333±0.0094 | 0.9556±0.0128 | 0.8348±0.0194 | 0.9574±0.0130 | 0.8950±0.0131 |
| 8 | 92.4±0.8944 | 100 | 0.9240±0.0089 | 0.9511±0.0036 | 0.8526±0.0235 | 0.9407±0.0135 | 0.8814±0.0120 |
| 9 | 73.0±1.2247 | 79 | 0.9241±0.0155 | 0.9496±0.0041 | 0.8375±0.0713 | 0.9460±0.0142 | 0.8842±0.0250 |
| 10 | 56.8±2.0494 | 62 | 0.9161±0.0331 | 0.9666±0.0108 | 0.9385±0.0644 | 0.9102±0.0310 | 0.8852±0.0444 |
| 11 | 51.2±1.3038 | 54 | 0.9481±0.0241 | 0.9714±0.0239 | 0.9333±0.0697 | 0.9524±0.0168 | 0.9274±0.0343 |
| 12 | 46.8±0.4472 | 50 | 0.9360±0.0089 | 0.9741±0.0135 | 0.9000±0.0373 | 0.9474±0.0186 | 0.9143±0.0104 |
| 13 | 41.2±0.4472 | 44 | 0.9364±0.0102 | 0.9392±0.0273 | 0.8500±0.0559 | 0.9556±0.0248 | 0.8955±0.0109 |
| 14 | 34.4±0.5477 | 37 | 0.9297±0.0148 | 0.9789±0.0065 | 0.9000±0.0559 | 0.9379±0.0154 | 0.9006±0.0209 |
| 15 | 28.2±0.4472 | 31 | 0.9097±0.0144 | 0.8887±0.0142 | 0.8571±0.0000 | 0.9250±0.0186 | 0.8760±0.0177 |
| 16 | 25.2±0.4472 | 28 | 0.9000±0.0160 | 0.9497±0.0182 | 0.9143±0.0782 | 0.8952±0.0213 | 0.8752±0.0213 |
| 17 | 22.0±0.0000 | 23 | 0.9565±0.0000 | 0.9735±0.0132 | 1.0000±0.0000 | 0.9412±0.0000 | 0.9464±0.0000 |
| 18 | 15.0±0.0000 | 16 | 0.9375±0.0000 | 0.9479±0.0104 | 1.0000±0.0000 | 0.9167±0.0000 | 0.9227±0.0000 |
| 19 | 10.2±0.4472 | 11 | 0.9273±0.0407 | 0.9964±0.0080 | 1.0000±0.0000 | 0.8857±0.0639 | 0.9248±0.0420 |
| 20 | 9.0±0.0000 | 10 | 0.9000±0.0000 | 0.9333±0.0342 | 1.0000±0.0000 | 0.8333±0.0000 | 0.8990±0.0000 |

Table 59: HM RNN-Ensemble-F1, using accuracy to choose models. Predictions from admission.

| Day | Corrects | Totals | Accuracy | AUC | Sensitivity | Specificity | F1 |
|---|---|---|---|---|---|---|---|
| 1 | 185.6±1.5166 | 194 | 0.9567±0.0078 | 0.9761±0.0096 | 0.9152±0.0136 | 0.9652±0.0094 | 0.9259±0.0121 |
| 2 | 181.8±1.0954 | 194 | 0.9371±0.0056 | 0.9277±0.0094 | 0.8000±0.0166 | 0.9652±0.0071 | 0.8873±0.0089 |
| 3 | 174.8±1.4832 | 187 | 0.9348±0.0079 | 0.9172±0.0012 | 0.8067±0.0279 | 0.9592±0.0057 | 0.8799±0.0147 |
| 4 | 167.6±1.1402 | 180 | 0.9311±0.0063 | 0.9359±0.0121 | 0.8000±0.0289 | 0.9563±0.0111 | 0.8741±0.0088 |
| 5 | 146.6±0.5477 | 160 | 0.9163±0.0034 | 0.8833±0.0134 | 0.7259±0.0422 | 0.9549±0.0075 | 0.8474±0.0089 |
| 6 | 130.4±0.8944 | 142 | 0.9183±0.0063 | 0.8972±0.0133 | 0.7280±0.0522 | 0.9590±0.0072 | 0.8543±0.0142 |
| 7 | 107.6±0.8944 | 117 | 0.9197±0.0076 | 0.8989±0.0107 | 0.8000±0.0238 | 0.9489±0.0117 | 0.8733±0.0104 |
| 8 | 89.4±0.5477 | 100 | 0.8940±0.0055 | 0.8643±0.0046 | 0.7474±0.0440 | 0.9284±0.0135 | 0.8311±0.0077 |
| 9 | 70.2±1.9235 | 79 | 0.8886±0.0243 | 0.9116±0.0172 | 0.8000±0.0523 | 0.9111±0.0309 | 0.8369±0.0296 |
| 10 | 55.6±2.7019 | 62 | 0.8968±0.0436 | 0.9278±0.0099 | 0.8769±0.0688 | 0.9020±0.0465 | 0.8582±0.0579 |
| 11 | 48.6±0.8944 | 54 | 0.9000±0.0166 | 0.9177±0.0192 | 0.8333±0.0000 | 0.9190±0.0213 | 0.8614±0.0200 |
| 12 | 42.4±1.3416 | 50 | 0.8480±0.0268 | 0.8366±0.0198 | 0.6000±0.0373 | 0.9263±0.0343 | 0.7791±0.0303 |
| 13 | 37.0±1.0000 | 44 | 0.8409±0.0227 | 0.8413±0.0320 | 0.6000±0.1046 | 0.8944±0.0304 | 0.7391±0.0355 |
| 14 | 30.0±0.7071 | 37 | 0.8108±0.0191 | 0.8082±0.0258 | 0.7250±0.0559 | 0.8345±0.0154 | 0.7485±0.0264 |
| 15 | 24.0±0.7071 | 31 | 0.7742±0.0228 | 0.7810±0.0585 | 0.5714±0.0000 | 0.8333±0.0295 | 0.6926±0.0213 |
| 16 | 21.4±1.5166 | 28 | 0.7643±0.0542 | 0.7347±0.0606 | 0.3143±0.1195 | 0.9143±0.0398 | 0.6257±0.0869 |
| 17 | 17.4±1.6733 | 23 | 0.7565±0.0728 | 0.6892±0.0476 | 0.3000±0.2472 | 0.9176±0.0322 | 0.6038±0.1540 |
| 18 | 11.8±0.4472 | 16 | 0.7375±0.0280 | 0.6729±0.0587 | 0.3500±0.1369 | 0.8667±0.0456 | 0.6111±0.0542 |
| 19 | 7.0±0.0000 | 11 | 0.6364±0.0000 | 0.7071±0.0889 | 0.5000±0.0000 | 0.7143±0.0000 | 0.6071±0.0000 |
| 20 | 7.2±0.4472 | 10 | 0.7200±0.0447 | 0.7333±0.0559 | 0.5000±0.0000 | 0.8667±0.0745 | 0.6886±0.0410 |

Table 60: HM RNN-Ensemble-F1, using accuracy to choose models. Predictions from outcome.

| Day | Corrects | Totals | Accuracy | AUC | Sensitivity | Specificity | F1 |
|---|---|---|---|---|---|---|---|
| 1 | 185.6±1.5166 | 194 | 0.9567±0.0078 | 0.9761±0.0096 | 0.9152±0.0136 | 0.9652±0.0094 | 0.9259±0.0121 |
| 2 | 181.8±1.0954 | 194 | 0.9371±0.0056 | 0.9277±0.0094 | 0.8000±0.0166 | 0.9652±0.0071 | 0.8873±0.0089 |
| 3 | 174.8±1.4832 | 187 | 0.9348±0.0079 | 0.9172±0.0012 | 0.8067±0.0279 | 0.9592±0.0057 | 0.8799±0.0147 |
| 4 | 167.6±1.1402 | 180 | 0.9311±0.0063 | 0.9359±0.0121 | 0.8000±0.0289 | 0.9563±0.0111 | 0.8741±0.0088 |
| 5 | 146.6±0.5477 | 160 | 0.9163±0.0034 | 0.8833±0.0134 | 0.7259±0.0422 | 0.9549±0.0075 | 0.8474±0.0089 |
| 6 | 130.4±0.8944 | 142 | 0.9183±0.0063 | 0.8972±0.0133 | 0.7280±0.0522 | 0.9590±0.0072 | 0.8543±0.0142 |
| 7 | 107.6±0.8944 | 117 | 0.9197±0.0076 | 0.8989±0.0107 | 0.8000±0.0238 | 0.9489±0.0117 | 0.8733±0.0104 |
| 8 | 89.4±0.5477 | 100 | 0.8940±0.0055 | 0.8643±0.0046 | 0.7474±0.0440 | 0.9284±0.0135 | 0.8311±0.0077 |
| 9 | 70.2±1.9235 | 79 | 0.8886±0.0243 | 0.9116±0.0172 | 0.8000±0.0523 | 0.9111±0.0309 | 0.8369±0.0296 |
| 10 | 55.6±2.7019 | 62 | 0.8968±0.0436 | 0.9278±0.0099 | 0.8769±0.0688 | 0.9020±0.0465 | 0.8582±0.0579 |
| 11 | 48.6±0.8944 | 54 | 0.9000±0.0166 | 0.9177±0.0192 | 0.8333±0.0000 | 0.9190±0.0213 | 0.8614±0.0200 |
| 12 | 42.4±1.3416 | 50 | 0.8480±0.0268 | 0.8366±0.0198 | 0.6000±0.0373 | 0.9263±0.0343 | 0.7791±0.0303 |
| 13 | 37.0±1.0000 | 44 | 0.8409±0.0227 | 0.8413±0.0320 | 0.6000±0.1046 | 0.8944±0.0304 | 0.7391±0.0355 |
| 14 | 30.0±0.7071 | 37 | 0.8108±0.0191 | 0.8082±0.0258 | 0.7250±0.0559 | 0.8345±0.0154 | 0.7485±0.0264 |
| 15 | 24.0±0.7071 | 31 | 0.7742±0.0228 | 0.7810±0.0585 | 0.5714±0.0000 | 0.8333±0.0295 | 0.6926±0.0213 |
| 16 | 21.4±1.5166 | 28 | 0.7643±0.0542 | 0.7347±0.0606 | 0.3143±0.1195 | 0.9143±0.0398 | 0.6257±0.0869 |
| 17 | 17.4±1.6733 | 23 | 0.7565±0.0728 | 0.6892±0.0476 | 0.3000±0.2472 | 0.9176±0.0322 | 0.6038±0.1540 |
| 18 | 11.8±0.4472 | 16 | 0.7375±0.0280 | 0.6729±0.0587 | 0.3500±0.1369 | 0.8667±0.0456 | 0.6111±0.0542 |
| 19 | 7.0±0.0000 | 11 | 0.6364±0.0000 | 0.7071±0.0889 | 0.5000±0.0000 | 0.7143±0.0000 | 0.6071±0.0000 |
| 20 | 7.2±0.4472 | 10 | 0.7200±0.0447 | 0.7333±0.0559 | 0.5000±0.0000 | 0.8667±0.0745 | 0.6886±0.0410 |

Table 61: HM RNN-Ensemble-F1, using F1 to choose models. Predictions from admission.

| Day | Corrects | Totals | Accuracy | AUC | Sensitivity | Specificity | F1 |
|---|---|---|---|---|---|---|---|
| 1 | 185.6±1.1402 | 194 | 0.9567±0.0059 | 0.9690±0.0046 | 0.9333±0.0254 | 0.9615±0.0111 | 0.9269±0.0082 |
| 2 | 181.0±1.8708 | 194 | 0.9330±0.0096 | 0.9233±0.0074 | 0.7939±0.0395 | 0.9615±0.0052 | 0.8803±0.0180 |
| 3 | 174.2±1.6432 | 187 | 0.9316±0.0088 | 0.9164±0.0015 | 0.8133±0.0447 | 0.9541±0.0094 | 0.8755±0.0163 |
| 4 | 167.6±1.3416 | 180 | 0.9311±0.0075 | 0.9306±0.0145 | 0.8345±0.0154 | 0.9497±0.0076 | 0.8774±0.0120 |
| 5 | 145.6±1.5166 | 160 | 0.9100±0.0095 | 0.8749±0.0097 | 0.7333±0.0310 | 0.9459±0.0112 | 0.8397±0.0149 |
| 6 | 129.0±0.7071 | 142 | 0.9085±0.0050 | 0.8806±0.0248 | 0.7600±0.0283 | 0.9402±0.0105 | 0.8447±0.0059 |
| 7 | 106.4±1.5166 | 117 | 0.9094±0.0130 | 0.8976±0.0105 | 0.8087±0.0496 | 0.9340±0.0265 | 0.8609±0.0130 |
| 8 | 89.0±0.7071 | 100 | 0.8900±0.0071 | 0.8676±0.0106 | 0.7368±0.0372 | 0.9259±0.0087 | 0.8247±0.0119 |
| 9 | 69.2±1.3038 | 79 | 0.8759±0.0165 | 0.8966±0.0207 | 0.7875±0.0342 | 0.8984±0.0241 | 0.8204±0.0182 |
| 10 | 55.2±1.6432 | 62 | 0.8903±0.0265 | 0.9171±0.0126 | 0.8615±0.0344 | 0.8980±0.0323 | 0.8484±0.0317 |
| 11 | 47.4±1.1402 | 54 | 0.8778±0.0211 | 0.9042±0.0180 | 0.8333±0.0589 | 0.8905±0.0130 | 0.8353±0.0293 |
| 12 | 42.0±1.5811 | 50 | 0.8400±0.0316 | 0.8237±0.0266 | 0.6500±0.0373 | 0.9000±0.0343 | 0.7788±0.0383 |
| 13 | 37.2±1.0954 | 44 | 0.8455±0.0249 | 0.8326±0.0259 | 0.7000±0.0685 | 0.8778±0.0248 | 0.7627±0.0358 |
| 14 | 28.8±1.0954 | 37 | 0.7784±0.0296 | 0.8108±0.0490 | 0.7250±0.0559 | 0.7931±0.0244 | 0.7175±0.0354 |
| 15 | 23.6±1.6733 | 31 | 0.7613±0.0540 | 0.7923±0.0340 | 0.6000±0.0639 | 0.8083±0.0697 | 0.6872±0.0527 |
| 16 | 21.0±1.8708 | 28 | 0.7500±0.0668 | 0.6755±0.0577 | 0.4571±0.2119 | 0.8476±0.0522 | 0.6499±0.1107 |
| 17 | 17.2±0.8367 | 23 | 0.7478±0.0364 | 0.6902±0.0992 | 0.3333±0.1179 | 0.8941±0.0644 | 0.6190±0.0495 |
| 18 | 11.0±1.0000 | 16 | 0.6875±0.0625 | 0.5813±0.0677 | 0.3000±0.1118 | 0.8167±0.0697 | 0.5599±0.0703 |
| 19 | 7.2±0.4472 | 11 | 0.6545±0.0407 | 0.6893±0.0573 | 0.5500±0.1118 | 0.7143±0.0000 | 0.6293±0.0496 |
| 20 | 7.2±0.4472 | 10 | 0.7200±0.0447 | 0.7542±0.0632 | 0.5500±0.1118 | 0.8333±0.0000 | 0.6946±0.0543 |

Table 62: HM RNN-Ensemble-F1, using F1 to choose models. Predictions from outcome.

| Day | Corrects | Totals | Accuracy | AUC | Sensitivity | Specificity | F1 |
|---|---|---|---|---|---|---|---|
| 1 | 164.0±2.0000 | 194 | 0.8454±0.0103 | 0.7599±0.0245 | 0.6061±0.0525 | 0.8944±0.0176 | 0.7383±0.0139 |
| 2 | 163.8±2.1679 | 194 | 0.8443±0.0112 | 0.7790±0.0235 | 0.6364±0.0567 | 0.8870±0.0155 | 0.7428±0.0180 |
| 3 | 160.6±2.0736 | 187 | 0.8588±0.0111 | 0.8469±0.0035 | 0.7667±0.0000 | 0.8764±0.0132 | 0.7741±0.0126 |
| 4 | 160.4±0.5477 | 180 | 0.8911±0.0030 | 0.8799±0.0086 | 0.8000±0.0154 | 0.9086±0.0055 | 0.8182±0.0036 |
| 5 | 146.4±0.8944 | 160 | 0.9150±0.0056 | 0.9119±0.0064 | 0.8593±0.0166 | 0.9263±0.0063 | 0.8605±0.0082 |
| 6 | 129.2±1.6432 | 142 | 0.9099±0.0116 | 0.9354±0.0216 | 0.9040±0.0456 | 0.9111±0.0047 | 0.8612±0.0189 |
| 7 | 107.4±0.8944 | 117 | 0.9179±0.0076 | 0.9556±0.0128 | 0.9391±0.0238 | 0.9128±0.0089 | 0.8826±0.0105 |
| 8 | 91.0±1.8708 | 100 | 0.9100±0.0187 | 0.9511±0.0036 | 0.9474±0.0000 | 0.9012±0.0231 | 0.8714±0.0227 |
| 9 | 72.2±1.3038 | 79 | 0.9139±0.0165 | 0.9496±0.0041 | 0.9375±0.0000 | 0.9079±0.0207 | 0.8799±0.0203 |
| 10 | 55.4±1.8166 | 62 | 0.8935±0.0293 | 0.9666±0.0108 | 1.0000±0.0000 | 0.8653±0.0371 | 0.8635±0.0332 |
| 11 | 50.2±0.8367 | 54 | 0.9296±0.0155 | 0.9714±0.0239 | 0.9833±0.0373 | 0.9143±0.0130 | 0.9071±0.0204 |
| 12 | 46.4±0.5477 | 50 | 0.9280±0.0110 | 0.9741±0.0135 | 0.9833±0.0373 | 0.9105±0.0235 | 0.9093±0.0117 |
| 13 | 40.0±0.7071 | 44 | 0.9091±0.0161 | 0.9392±0.0273 | 0.9000±0.0559 | 0.9111±0.0232 | 0.8628±0.0211 |
| 14 | 34.2±0.8367 | 37 | 0.9243±0.0226 | 0.9789±0.0065 | 1.0000±0.0000 | 0.9034±0.0289 | 0.9007±0.0267 |
| 15 | 28.0±0.0000 | 31 | 0.9032±0.0000 | 0.8887±0.0142 | 0.8571±0.0000 | 0.9167±0.0000 | 0.8681±0.0000 |
| 16 | 24.8±0.8367 | 28 | 0.8857±0.0299 | 0.9497±0.0182 | 1.0000±0.0000 | 0.8476±0.0398 | 0.8663±0.0318 |
| 17 | 21.8±0.4472 | 23 | 0.9478±0.0194 | 0.9735±0.0132 | 1.0000±0.0000 | 0.9294±0.0263 | 0.9366±0.0219 |
| 18 | 14.6±0.5477 | 16 | 0.9125±0.0342 | 0.9479±0.0104 | 1.0000±0.0000 | 0.8833±0.0456 | 0.8954±0.0373 |
| 19 | 10.0±0.0000 | 11 | 0.9091±0.0000 | 0.9964±0.0080 | 1.0000±0.0000 | 0.8571±0.0000 | 0.9060±0.0000 |
| 20 | 8.8±0.4472 | 10 | 0.8800±0.0447 | 0.9333±0.0342 | 1.0000±0.0000 | 0.8000±0.0745 | 0.8792±0.0443 |

Table 63: HM RNN-Ensemble-SEN, using accuracy to choose models. Predictions from admission.

| Day | Corrects | Totals | Accuracy | AUC | Sensitivity | Specificity | F1 |
|---|---|---|---|---|---|---|---|
| 1 | 181.8±2.5884 | 194 | 0.9371±0.0133 | 0.9761±0.0096 | 0.9758±0.0254 | 0.9292±0.0209 | 0.9013±0.0176 |
| 2 | 180.6±1.8166 | 194 | 0.9309±0.0094 | 0.9277±0.0094 | 0.8788±0.0214 | 0.9416±0.0094 | 0.8851±0.0145 |
| 3 | 173.8±1.0954 | 187 | 0.9294±0.0059 | 0.9172±0.0012 | 0.8667±0.0000 | 0.9414±0.0070 | 0.8775±0.0086 |
| 4 | 166.2±1.7889 | 180 | 0.9233±0.0099 | 0.9359±0.0121 | 0.8966±0.0244 | 0.9285±0.0086 | 0.8718±0.0157 |
| 5 | 146.0±1.7321 | 160 | 0.9125±0.0108 | 0.8833±0.0134 | 0.8074±0.0310 | 0.9338±0.0112 | 0.8519±0.0166 |
| 6 | 129.4±0.8944 | 142 | 0.9113±0.0063 | 0.8972±0.0133 | 0.8400±0.0283 | 0.9265±0.0076 | 0.8571±0.0097 |
| 7 | 104.6±2.3022 | 117 | 0.8940±0.0197 | 0.8989±0.0107 | 0.8435±0.0238 | 0.9064±0.0254 | 0.8455±0.0239 |
| 8 | 88.8±0.8367 | 100 | 0.8880±0.0084 | 0.8643±0.0046 | 0.7895±0.0000 | 0.9111±0.0103 | 0.8289±0.0103 |
| 9 | 68.6±1.6733 | 79 | 0.8684±0.0212 | 0.9116±0.0172 | 0.9125±0.0559 | 0.8571±0.0317 | 0.8250±0.0233 |
| 10 | 52.6±1.3416 | 62 | 0.8484±0.0216 | 0.9278±0.0099 | 0.9231±0.0000 | 0.8286±0.0274 | 0.8078±0.0227 |
| 11 | 45.8±1.7889 | 54 | 0.8481±0.0331 | 0.9177±0.0192 | 0.9000±0.0373 | 0.8333±0.0376 | 0.8106±0.0367 |
| 12 | 40.4±1.3416 | 50 | 0.8080±0.0268 | 0.8366±0.0198 | 0.7833±0.0456 | 0.8158±0.0372 | 0.7641±0.0271 |
| 13 | 36.2±0.8367 | 44 | 0.8227±0.0190 | 0.8413±0.0320 | 0.8250±0.0685 | 0.8222±0.0317 | 0.7560±0.0185 |
| 14 | 27.8±1.0954 | 37 | 0.7514±0.0296 | 0.8082±0.0258 | 0.8000±0.0685 | 0.7379±0.0463 | 0.7023±0.0261 |
| 15 | 22.6±1.5166 | 31 | 0.7290±0.0489 | 0.7810±0.0585 | 0.7429±0.1195 | 0.7250±0.0475 | 0.6790±0.0557 |
| 16 | 21.0±0.7071 | 28 | 0.7500±0.0253 | 0.7347±0.0606 | 0.6571±0.1278 | 0.7810±0.0261 | 0.6943±0.0388 |
| 17 | 17.2±0.8367 | 23 | 0.7478±0.0364 | 0.6892±0.0476 | 0.5333±0.0745 | 0.8235±0.0416 | 0.6763±0.0412 |
| 18 | 11.0±0.7071 | 16 | 0.6875±0.0442 | 0.6729±0.0587 | 0.6000±0.1369 | 0.7167±0.0456 | 0.6307±0.0542 |
| 19 | 7.6±0.5477 | 11 | 0.6909±0.0498 | 0.7071±0.0889 | 0.6500±0.1369 | 0.7143±0.0000 | 0.6736±0.0607 |
| 20 | 7.2±0.4472 | 10 | 0.7200±0.0447 | 0.7333±0.0559 | 0.5500±0.1118 | 0.8333±0.0000 | 0.6946±0.0543 |

Table 64: HM RNN-Ensemble-SEN, using accuracy to choose models. Predictions from outcome.

| Day | Corrects | Totals | Accuracy | AUC | Sensitivity | Specificity | F1 |
|---|---|---|---|---|---|---|---|
| 1 | 162.4±4.2778 | 194 | 0.8371±0.0221 | 0.7692±0.0255 | 0.6364±0.0429 | 0.8783±0.0251 | 0.7355±0.0274 |
| 2 | 164.2±1.7889 | 194 | 0.8464±0.0092 | 0.7851±0.0154 | 0.6545±0.0346 | 0.8857±0.0094 | 0.7485±0.0146 |
| 3 | 159.0±0.7071 | 187 | 0.8503±0.0038 | 0.8345±0.0122 | 0.7467±0.0298 | 0.8701±0.0073 | 0.7611±0.0062 |
| 4 | 158.2±1.7889 | 180 | 0.8789±0.0099 | 0.8734±0.0212 | 0.7931±0.0422 | 0.8954±0.0109 | 0.8019±0.0158 |
| 5 | 145.4±1.5166 | 160 | 0.9088±0.0095 | 0.9040±0.0144 | 0.8444±0.0310 | 0.9218±0.0114 | 0.8507±0.0141 |
| 6 | 127.8±1.3038 | 142 | 0.9000±0.0092 | 0.9405±0.0131 | 0.9200±0.0283 | 0.8957±0.0094 | 0.8504±0.0130 |
| 7 | 106.2±0.8367 | 117 | 0.9077±0.0072 | 0.9492±0.0121 | 0.9304±0.0238 | 0.9021±0.0089 | 0.8693±0.0095 |
| 8 | 89.8±1.3038 | 100 | 0.8980±0.0130 | 0.9430±0.0079 | 0.9368±0.0235 | 0.8889±0.0195 | 0.8558±0.0145 |
| 9 | 71.0±1.4142 | 79 | 0.8987±0.0179 | 0.9340±0.0150 | 0.9125±0.0342 | 0.8952±0.0241 | 0.8597±0.0211 |
| 10 | 55.4±1.5166 | 62 | 0.8935±0.0245 | 0.9648±0.0100 | 1.0000±0.0000 | 0.8653±0.0310 | 0.8633±0.0283 |
| 11 | 49.6±1.1402 | 54 | 0.9185±0.0211 | 0.9591±0.0262 | 0.9667±0.0456 | 0.9048±0.0238 | 0.8931±0.0263 |
| 12 | 45.8±0.8367 | 50 | 0.9160±0.0167 | 0.9649±0.0241 | 0.9667±0.0456 | 0.9000±0.0343 | 0.8948±0.0173 |
| 13 | 40.2±0.4472 | 44 | 0.9136±0.0102 | 0.9497±0.0313 | 0.9250±0.0685 | 0.9111±0.0232 | 0.8705±0.0124 |
| 14 | 33.8±0.8367 | 37 | 0.9135±0.0226 | 0.9608±0.0265 | 0.9750±0.0559 | 0.8966±0.0345 | 0.8863±0.0263 |
| 15 | 28.0±0.0000 | 31 | 0.9032±0.0000 | 0.8875±0.0074 | 0.8571±0.0000 | 0.9167±0.0000 | 0.8681±0.0000 |
| 16 | 24.4±1.1402 | 28 | 0.8714±0.0407 | 0.9286±0.0457 | 0.9429±0.0782 | 0.8476±0.0398 | 0.8472±0.0473 |
| 17 | 21.8±0.4472 | 23 | 0.9478±0.0194 | 0.9637±0.0102 | 1.0000±0.0000 | 0.9294±0.0263 | 0.9366±0.0219 |
| 18 | 14.6±0.5477 | 16 | 0.9125±0.0342 | 0.9562±0.0186 | 1.0000±0.0000 | 0.8833±0.0456 | 0.8954±0.0373 |
| 19 | 10.0±0.0000 | 11 | 0.9091±0.0000 | 0.9964±0.0080 | 1.0000±0.0000 | 0.8571±0.0000 | 0.9060±0.0000 |
| 20 | 8.8±0.4472 | 10 | 0.8800±0.0447 | 0.9458±0.0280 | 1.0000±0.0000 | 0.8000±0.0745 | 0.8792±0.0443 |

Table 65: HM RNN-Ensemble-SEN, using F1 to choose models. Predictions from admission.

| Day | Corrects | Totals | Accuracy | AUC | Sensitivity | Specificity | F1 |
|---|---|---|---|---|---|---|---|
| 1 | 181.6±3.2863 | 194 | 0.9361±0.0169 | 0.9817±0.0065 | 0.9879±0.0166 | 0.9255±0.0232 | 0.9008±0.0233 |
| 2 | 180.6±1.5166 | 194 | 0.9309±0.0078 | 0.9319±0.0095 | 0.8909±0.0166 | 0.9391±0.0081 | 0.8861±0.0119 |
| 3 | 173.2±0.8367 | 187 | 0.9262±0.0045 | 0.9172±0.0013 | 0.8667±0.0000 | 0.9376±0.0053 | 0.8728±0.0064 |
| 4 | 165.2±2.0494 | 180 | 0.9178±0.0114 | 0.9302±0.0143 | 0.8897±0.0289 | 0.9232±0.0100 | 0.8634±0.0176 |
| 5 | 144.8±1.6432 | 160 | 0.9050±0.0103 | 0.8757±0.0100 | 0.7926±0.0203 | 0.9278±0.0114 | 0.8401±0.0146 |
| 6 | 128.2±2.1679 | 142 | 0.9028±0.0153 | 0.8813±0.0261 | 0.8080±0.0522 | 0.9231±0.0105 | 0.8426±0.0250 |
| 7 | 104.0±2.0000 | 117 | 0.8889±0.0171 | 0.8974±0.0096 | 0.8435±0.0238 | 0.9000±0.0256 | 0.8393±0.0192 |
| 8 | 88.6±0.5477 | 100 | 0.8860±0.0055 | 0.8682±0.0096 | 0.8000±0.0235 | 0.9062±0.0068 | 0.8276±0.0080 |
| 9 | 67.8±1.0954 | 79 | 0.8582±0.0139 | 0.9042±0.0224 | 0.9125±0.0559 | 0.8444±0.0235 | 0.8137±0.0152 |
| 10 | 50.4±1.1402 | 62 | 0.8129±0.0184 | 0.9146±0.0055 | 0.9231±0.0000 | 0.7837±0.0233 | 0.7717±0.0182 |
| 11 | 44.6±1.9494 | 54 | 0.8259±0.0361 | 0.9010±0.0230 | 0.8833±0.0456 | 0.8095±0.0337 | 0.7862±0.0405 |
| 12 | 39.2±1.3038 | 50 | 0.7840±0.0261 | 0.8169±0.0258 | 0.7667±0.0697 | 0.7895±0.0186 | 0.7386±0.0328 |
| 13 | 34.6±0.8944 | 44 | 0.7864±0.0203 | 0.8229±0.0303 | 0.8000±0.0685 | 0.7833±0.0304 | 0.7168±0.0208 |
| 14 | 28.0±1.2247 | 37 | 0.7568±0.0331 | 0.8194±0.0534 | 0.8250±0.1425 | 0.7379±0.0189 | 0.7092±0.0466 |
| 15 | 22.4±1.1402 | 31 | 0.7226±0.0368 | 0.7881±0.0393 | 0.7714±0.0782 | 0.7083±0.0295 | 0.6774±0.0414 |
| 16 | 19.6±1.6733 | 28 | 0.7000±0.0598 | 0.6830±0.0444 | 0.5714±0.1010 | 0.7429±0.0722 | 0.6376±0.0602 |
| 17 | 17.4±1.3416 | 23 | 0.7565±0.0583 | 0.6980±0.1153 | 0.5667±0.2528 | 0.8235±0.0416 | 0.6799±0.1099 |
| 18 | 10.8±1.4832 | 16 | 0.6750±0.0927 | 0.6292±0.1014 | 0.5000±0.1768 | 0.7333±0.0697 | 0.6035±0.1116 |
| 19 | 7.8±0.8367 | 11 | 0.7091±0.0761 | 0.7107±0.0719 | 0.7000±0.2092 | 0.7143±0.0000 | 0.6934±0.0884 |
| 20 | 7.6±0.5477 | 10 | 0.7600±0.0548 | 0.7750±0.0614 | 0.6500±0.1369 | 0.8333±0.0000 | 0.7431±0.0665 |

Table 66: HM RNN-Ensemble-SEN, using F1 to choose models. Predictions from outcome.

##### 8.6 Daily Results for H12O

###### 8.6.1 Random Forest

| Day | Corrects | Totals | Accuracy | AUC | Sensitivity | Specificity | F1 |
|---|---|---|---|---|---|---|---|
| 1 | 335.2±2.3875 | 374 | 0.8963±0.0064 | 0.6763±0.0070 | 0.3760±0.0089 | 0.9765±0.0064 | 0.7174±0.0119 |
| 2 | 335.6±1.8166 | 374 | 0.8973±0.0049 | 0.6735±0.0074 | 0.3680±0.0110 | 0.9790±0.0040 | 0.7162±0.0109 |
| 3 | 341.6±1.3416 | 374 | 0.9134±0.0036 | 0.7453±0.0127 | 0.5160±0.0261 | 0.9747±0.0026 | 0.7826±0.0111 |
| 4 | 338.0±1.2247 | 364 | 0.9286±0.0034 | 0.7705±0.0114 | 0.5574±0.0233 | 0.9836±0.0026 | 0.8141±0.0100 |
| 5 | 312.2±1.3038 | 332 | 0.9404±0.0039 | 0.7884±0.0087 | 0.5897±0.0181 | 0.9870±0.0044 | 0.8330±0.0093 |
| 6 | 274.2±0.4472 | 291 | 0.9423±0.0015 | 0.8040±0.0106 | 0.6235±0.0246 | 0.9844±0.0039 | 0.8420±0.0048 |
| 7 | 232.4±0.8944 | 250 | 0.9296±0.0036 | 0.7924±0.0085 | 0.6138±0.0154 | 0.9710±0.0025 | 0.8149±0.0092 |
| 8 | 188.8±1.4832 | 201 | 0.9393±0.0074 | 0.8215±0.0168 | 0.6667±0.0295 | 0.9763±0.0047 | 0.8450±0.0185 |
| 9 | 147.0±1.5811 | 159 | 0.9245±0.0099 | 0.7870±0.0286 | 0.6000±0.0543 | 0.9739±0.0040 | 0.8170±0.0270 |
| 10 | 124.6±0.8944 | 133 | 0.9368±0.0067 | 0.8022±0.0222 | 0.6250±0.0442 | 0.9795±0.0047 | 0.8342±0.0193 |
| 11 | 108.2±0.8367 | 117 | 0.9248±0.0072 | 0.7863±0.0041 | 0.6000±0.0000 | 0.9725±0.0082 | 0.8148±0.0125 |
| 12 | 91.2±1.0954 | 99 | 0.9212±0.0111 | 0.7784±0.0243 | 0.5846±0.0421 | 0.9721±0.0064 | 0.8081±0.0269 |
| 13 | 82.8±0.8367 | 91 | 0.9099±0.0092 | 0.8321±0.0219 | 0.7231±0.0421 | 0.9410±0.0070 | 0.8216±0.0186 |
| 14 | 71.0±1.0000 | 75 | 0.9467±0.0133 | 0.9345±0.0079 | 0.9167±0.0000 | 0.9524±0.0159 | 0.9074±0.0205 |
| 15 | 62.4±0.5477 | 67 | 0.9313±0.0082 | 0.8859±0.0049 | 0.8182±0.0000 | 0.9536±0.0098 | 0.8778±0.0123 |
| 16 | 54.8±0.4472 | 59 | 0.9288±0.0076 | 0.8696±0.0224 | 0.7800±0.0447 | 0.9592±0.0000 | 0.8723±0.0163 |
| 17 | 52.8±0.8367 | 54 | 0.9778±0.0155 | 0.9333±0.0465 | 0.8667±0.0930 | 1.0000±0.0000 | 0.9567±0.0309 |
| 18 | 41.4±0.8944 | 45 | 0.9200±0.0199 | 0.9024±0.0121 | 0.8750±0.0000 | 0.9297±0.0242 | 0.8736±0.0261 |
| 19 | 35.8±0.4472 | 38 | 0.9421±0.0118 | 0.8979±0.0070 | 0.8333±0.0000 | 0.9625±0.0140 | 0.8930±0.0180 |
| 20 | 30.8±0.4472 | 35 | 0.8800±0.0128 | 0.8483±0.0373 | 0.8000±0.0745 | 0.8966±0.0000 | 0.8099±0.0258 |

Table 67: H12O Random Forest baseline results of CV using base features, taking best model of grid using accuracy. Predictions from admission.

| Day | Corrects | Totals | Accuracy | AUC | Sensitivity | Specificity | F1 |
|---|---|---|---|---|---|---|---|
| 1 | 357.2±0.8367 | 374 | 0.9551±0.0022 | 0.8861±0.0044 | 0.7920±0.0110 | 0.9802±0.0035 | 0.8996±0.0040 |
| 2 | 361.2±1.4832 | 374 | 0.9658±0.0040 | 0.8991±0.0119 | 0.8080±0.0228 | 0.9901±0.0014 | 0.9218±0.0096 |
| 3 | 359.8±1.4832 | 374 | 0.9620±0.0040 | 0.8884±0.0095 | 0.7880±0.0179 | 0.9889±0.0028 | 0.9128±0.0091 |
| 4 | 350.6±1.1402 | 364 | 0.9632±0.0031 | 0.8701±0.0081 | 0.7447±0.0150 | 0.9956±0.0017 | 0.9093±0.0079 |
| 5 | 312.8±1.6432 | 332 | 0.9422±0.0049 | 0.7805±0.0189 | 0.5692±0.0380 | 0.9918±0.0031 | 0.8328±0.0169 |
| 6 | 271.4±1.5166 | 291 | 0.9326±0.0052 | 0.7475±0.0238 | 0.5059±0.0483 | 0.9891±0.0017 | 0.7995±0.0209 |
| 7 | 231.8±0.8367 | 250 | 0.9272±0.0033 | 0.7641±0.0108 | 0.5517±0.0244 | 0.9765±0.0050 | 0.7984±0.0080 |
| 8 | 181.8±0.8367 | 201 | 0.9045±0.0042 | 0.6828±0.0118 | 0.3917±0.0228 | 0.9740±0.0031 | 0.7209±0.0130 |
| 9 | 140.4±1.1402 | 159 | 0.8830±0.0072 | 0.6258±0.0106 | 0.2762±0.0213 | 0.9754±0.0083 | 0.6598±0.0146 |
| 10 | 118.6±0.5477 | 133 | 0.8917±0.0041 | 0.6417±0.0023 | 0.3125±0.0000 | 0.9709±0.0047 | 0.6752±0.0058 |
| 11 | 103.0±0.7071 | 117 | 0.8803±0.0060 | 0.6471±0.0236 | 0.3333±0.0471 | 0.9608±0.0000 | 0.6745±0.0249 |
| 12 | 85.2±1.6432 | 99 | 0.8606±0.0166 | 0.6521±0.0365 | 0.3692±0.0644 | 0.9349±0.0104 | 0.6656±0.0402 |
| 13 | 79.8±1.4832 | 91 | 0.8769±0.0163 | 0.6910±0.0258 | 0.4308±0.0421 | 0.9513±0.0140 | 0.7153±0.0324 |
| 14 | 65.0±0.0000 | 75 | 0.8667±0.0000 | 0.6306±0.0185 | 0.2833±0.0456 | 0.9778±0.0087 | 0.6638±0.0187 |
| 15 | 59.4±1.3416 | 67 | 0.8866±0.0200 | 0.7714±0.0260 | 0.6000±0.0498 | 0.9429±0.0233 | 0.7842±0.0307 |
| 16 | 48.8±1.3038 | 59 | 0.8271±0.0221 | 0.6810±0.0466 | 0.4600±0.0894 | 0.9020±0.0171 | 0.6845±0.0432 |
| 17 | 44.4±0.8944 | 54 | 0.8222±0.0166 | 0.6089±0.0298 | 0.2889±0.0609 | 0.9289±0.0186 | 0.6234±0.0331 |
| 18 | 34.4±0.8944 | 45 | 0.7644±0.0199 | 0.5922±0.0336 | 0.3250±0.0685 | 0.8595±0.0226 | 0.5924±0.0323 |
| 19 | 28.4±1.1402 | 38 | 0.7474±0.0300 | 0.5792±0.0178 | 0.3333±0.0000 | 0.8250±0.0356 | 0.5708±0.0232 |
| 20 | 26.8±0.8367 | 35 | 0.7657±0.0239 | 0.5282±0.0144 | 0.1667±0.0000 | 0.8897±0.0289 | 0.5299±0.0163 |

Table 68: H12O Random Forest baseline results of CV using base features, taking best model of grid using accuracy. Predictions from outcome.

| Day | Corrects | Totals | Accuracy | AUC | Sensitivity | Specificity | F1 |
|---|---|---|---|---|---|---|---|
| 1 | 332.6±1.3416 | 374 | 0.8893±0.0036 | 0.6824±0.0115 | 0.4000±0.0245 | 0.9648±0.0041 | 0.7145±0.0111 |
| 2 | 331.6±2.7019 | 374 | 0.8866±0.0072 | 0.6910±0.0105 | 0.4240±0.0167 | 0.9580±0.0068 | 0.7182±0.0138 |
| 3 | 336.0±1.0000 | 374 | 0.8984±0.0027 | 0.7485±0.0068 | 0.5440±0.0167 | 0.9531±0.0046 | 0.7654±0.0045 |
| 4 | 336.0±2.0000 | 364 | 0.9231±0.0055 | 0.7819±0.0149 | 0.5915±0.0277 | 0.9722±0.0026 | 0.8107±0.0144 |
| 5 | 309.2±2.2804 | 332 | 0.9313±0.0069 | 0.8077±0.0240 | 0.6462±0.0493 | 0.9693±0.0064 | 0.8248±0.0189 |
| 6 | 271.6±2.3022 | 291 | 0.9333±0.0079 | 0.8244±0.0123 | 0.6824±0.0246 | 0.9665±0.0090 | 0.8340±0.0155 |
| 7 | 231.0±0.7071 | 250 | 0.9240±0.0028 | 0.8222±0.0122 | 0.6897±0.0244 | 0.9548±0.0000 | 0.8174±0.0087 |
| 8 | 185.6±0.8944 | 201 | 0.9234±0.0044 | 0.8340±0.0172 | 0.7167±0.0349 | 0.9514±0.0031 | 0.8234±0.0121 |
| 9 | 146.6±0.8944 | 159 | 0.9220±0.0056 | 0.8097±0.0107 | 0.6571±0.0213 | 0.9623±0.0061 | 0.8227±0.0113 |
| 10 | 123.2±2.0494 | 133 | 0.9263±0.0154 | 0.8394±0.0234 | 0.7250±0.0342 | 0.9538±0.0130 | 0.8311±0.0310 |
| 11 | 103.4±0.8944 | 117 | 0.8838±0.0076 | 0.7627±0.0044 | 0.6000±0.0000 | 0.9255±0.0088 | 0.7514±0.0107 |
| 12 | 90.4±0.5477 | 99 | 0.9131±0.0055 | 0.7802±0.0298 | 0.6000±0.0644 | 0.9605±0.0064 | 0.7969±0.0196 |
| 13 | 81.6±0.8944 | 91 | 0.8967±0.0098 | 0.8115±0.0057 | 0.6923±0.0000 | 0.9308±0.0115 | 0.7983±0.0134 |
| 14 | 69.4±0.5477 | 75 | 0.9253±0.0073 | 0.9151±0.0173 | 0.9000±0.0373 | 0.9302±0.0087 | 0.8742±0.0122 |
| 15 | 61.6±0.5477 | 67 | 0.9194±0.0082 | 0.8787±0.0049 | 0.8182±0.0000 | 0.9393±0.0098 | 0.8604±0.0115 |
| 16 | 54.6±0.5477 | 59 | 0.9254±0.0093 | 0.8835±0.0207 | 0.8200±0.0447 | 0.9469±0.0112 | 0.8716±0.0154 |
| 17 | 50.8±0.4472 | 54 | 0.9407±0.0083 | 0.9378±0.0268 | 0.9333±0.0609 | 0.9422±0.0122 | 0.9017±0.0141 |
| 18 | 40.0±0.0000 | 45 | 0.8889±0.0000 | 0.8834±0.0000 | 0.8750±0.0000 | 0.8919±0.0000 | 0.8332±0.0000 |
| 19 | 36.0±0.7071 | 38 | 0.9474±0.0186 | 0.9146±0.0396 | 0.8667±0.0745 | 0.9625±0.0140 | 0.9035±0.0330 |
| 20 | 31.2±0.4472 | 35 | 0.8914±0.0128 | 0.8684±0.0077 | 0.8333±0.0000 | 0.9034±0.0154 | 0.8288±0.0165 |

Table 69: H12O Random Forest baseline results of CV using base features, taking best model of grid using F1. Predictions from admission.

| Day | Corrects | Totals | Accuracy | AUC | Sensitivity | Specificity | F1 |
|---|---|---|---|---|---|---|---|
| 1 | 353.6±0.8944 | 374 | 0.9455±0.0024 | 0.8856±0.0035 | 0.8040±0.0089 | 0.9673±0.0035 | 0.8831±0.0040 |
| 2 | 361.2±1.9235 | 374 | 0.9658±0.0051 | 0.9041±0.0130 | 0.8200±0.0245 | 0.9883±0.0034 | 0.9227±0.0117 |
| 3 | 360.2±1.3038 | 374 | 0.9631±0.0035 | 0.9026±0.0073 | 0.8200±0.0141 | 0.9852±0.0034 | 0.9174±0.0074 |
| 4 | 348.4±0.5477 | 364 | 0.9571±0.0015 | 0.8848±0.0065 | 0.7872±0.0150 | 0.9823±0.0028 | 0.9007±0.0033 |
| 5 | 312.6±1.6733 | 332 | 0.9416±0.0050 | 0.8002±0.0164 | 0.6154±0.0314 | 0.9850±0.0019 | 0.8397±0.0152 |
| 6 | 272.8±0.8367 | 291 | 0.9375±0.0029 | 0.7757±0.0120 | 0.5647±0.0246 | 0.9868±0.0021 | 0.8218±0.0100 |
| 7 | 229.6±0.5477 | 250 | 0.9184±0.0022 | 0.7771±0.0135 | 0.5931±0.0289 | 0.9611±0.0025 | 0.7908±0.0089 |
| 8 | 178.2±1.3038 | 201 | 0.8866±0.0065 | 0.6871±0.0218 | 0.4250±0.0456 | 0.9492±0.0069 | 0.7040±0.0190 |
| 9 | 138.6±0.8944 | 159 | 0.8717±0.0056 | 0.6314±0.0261 | 0.3048±0.0543 | 0.9580±0.0032 | 0.6561±0.0272 |
| 10 | 116.2±1.3038 | 133 | 0.8737±0.0098 | 0.6530±0.0174 | 0.3625±0.0280 | 0.9436±0.0076 | 0.6691±0.0210 |
| 11 | 99.4±0.8944 | 117 | 0.8496±0.0076 | 0.6522±0.0167 | 0.3867±0.0298 | 0.9176±0.0054 | 0.6557±0.0170 |
| 12 | 84.4±0.8944 | 99 | 0.8525±0.0090 | 0.6866±0.0052 | 0.4615±0.0000 | 0.9116±0.0104 | 0.6832±0.0102 |
| 13 | 76.4±0.8944 | 91 | 0.8396±0.0098 | 0.7013±0.0206 | 0.5077±0.0421 | 0.8949±0.0107 | 0.6899±0.0174 |
| 14 | 63.6±0.8944 | 75 | 0.8480±0.0119 | 0.6734±0.0071 | 0.4167±0.0000 | 0.9302±0.0142 | 0.6896±0.0139 |
| 15 | 54.8±1.0954 | 67 | 0.8179±0.0163 | 0.7377±0.0182 | 0.6182±0.0407 | 0.8571±0.0219 | 0.7073±0.0178 |
| 16 | 47.6±1.5166 | 59 | 0.8068±0.0257 | 0.7086±0.0275 | 0.5600±0.0548 | 0.8571±0.0323 | 0.6884±0.0297 |
| 17 | 42.0±0.7071 | 54 | 0.7778±0.0131 | 0.6178±0.0256 | 0.3778±0.0609 | 0.8578±0.0199 | 0.6129±0.0212 |
| 18 | 31.8±1.3038 | 45 | 0.7067±0.0290 | 0.5473±0.0427 | 0.3000±0.0685 | 0.7946±0.0242 | 0.5418±0.0402 |
| 19 | 25.0±1.2247 | 38 | 0.6579±0.0322 | 0.5260±0.0191 | 0.3333±0.0000 | 0.7188±0.0383 | 0.5077±0.0210 |
| 20 | 26.0±0.0000 | 35 | 0.7429±0.0000 | 0.5144±0.0000 | 0.1667±0.0000 | 0.8621±0.0000 | 0.5146±0.0000 |

Table 70: H12O Random Forest baseline results of CV using base features, taking best model of grid using F1. Predictions from outcome.

| Day | Corrects | Totals | Accuracy | AUC | Sensitivity | Specificity | F1 |
|---|---|---|---|---|---|---|---|
| 1 | 335.0±2.3452 | 374 | 0.8957±0.0063 | 0.6743±0.0197 | 0.3720±0.0390 | 0.9765±0.0035 | 0.7147±0.0216 |
| 2 | 340.0±1.0000 | 374 | 0.9091±0.0027 | 0.7124±0.0122 | 0.4440±0.0261 | 0.9809±0.0026 | 0.7576±0.0108 |
| 3 | 345.0±1.5811 | 374 | 0.9225±0.0042 | 0.7692±0.0150 | 0.5600±0.0316 | 0.9784±0.0044 | 0.8074±0.0122 |
| 4 | 336.8±1.7889 | 364 | 0.9253±0.0049 | 0.7831±0.0122 | 0.5915±0.0233 | 0.9748±0.0039 | 0.8146±0.0123 |
| 5 | 313.4±1.5166 | 332 | 0.9440±0.0046 | 0.8193±0.0146 | 0.6564±0.0292 | 0.9823±0.0037 | 0.8510±0.0127 |
| 6 | 274.0±1.0000 | 291 | 0.9416±0.0034 | 0.8087±0.0163 | 0.6353±0.0335 | 0.9821±0.0021 | 0.8423±0.0119 |
| 7 | 234.2±1.0954 | 250 | 0.9368±0.0044 | 0.7905±0.0189 | 0.6000±0.0393 | 0.9810±0.0038 | 0.8261±0.0144 |
| 8 | 190.4±1.3416 | 201 | 0.9473±0.0067 | 0.8440±0.0286 | 0.7083±0.0589 | 0.9797±0.0051 | 0.8659±0.0202 |
| 9 | 148.8±0.4472 | 159 | 0.9358±0.0028 | 0.8137±0.0121 | 0.6476±0.0261 | 0.9797±0.0032 | 0.8454±0.0082 |
| 10 | 124.8±0.4472 | 133 | 0.9383±0.0034 | 0.8247±0.0117 | 0.6750±0.0280 | 0.9744±0.0060 | 0.8450±0.0066 |
| 11 | 109.0±1.2247 | 117 | 0.9316±0.0105 | 0.8129±0.0184 | 0.6533±0.0298 | 0.9725±0.0082 | 0.8359±0.0230 |
| 12 | 91.0±0.7071 | 99 | 0.9192±0.0071 | 0.7968±0.0180 | 0.6308±0.0344 | 0.9628±0.0052 | 0.8130±0.0166 |
| 13 | 82.2±0.8367 | 91 | 0.9033±0.0092 | 0.7769±0.0189 | 0.6000±0.0344 | 0.9538±0.0070 | 0.7917±0.0196 |
| 14 | 71.2±0.8367 | 75 | 0.9493±0.0112 | 0.9024±0.0297 | 0.8333±0.0589 | 0.9714±0.0071 | 0.9048±0.0224 |
| 15 | 64.0±0.0000 | 67 | 0.9552±0.0000 | 0.9367±0.0000 | 0.9091±0.0000 | 0.9643±0.0000 | 0.9213±0.0000 |
| 16 | 57.4±0.8944 | 59 | 0.9729±0.0152 | 0.9598±0.0447 | 0.9400±0.0894 | 0.9796±0.0000 | 0.9518±0.0290 |
| 17 | 53.6±0.8944 | 54 | 0.9926±0.0166 | 0.9778±0.0497 | 0.9556±0.0994 | 1.0000±0.0000 | 0.9853±0.0328 |
| 18 | 44.6±0.5477 | 45 | 0.9911±0.0122 | 0.9750±0.0342 | 0.9500±0.0685 | 1.0000±0.0000 | 0.9840±0.0219 |
| 19 | 38.0±0.0000 | 38 | 1.0000±0.0000 | 1.0000±0.0000 | 1.0000±0.0000 | 1.0000±0.0000 | 1.0000±0.0000 |
| 20 | 35.0±0.0000 | 35 | 1.0000±0.0000 | 1.0000±0.0000 | 1.0000±0.0000 | 1.0000±0.0000 | 1.0000±0.0000 |

Table 71: H12O Random Forest with imputation, results of CV, taking best model of grid using accuracy. Predictions from admission.

| Day | Corrects | Totals | Accuracy | AUC | Sensitivity | Specificity | F1 |
|---|---|---|---|---|---|---|---|
| 1 | 368.2±0.8367 | 374 | 0.9845±0.0022 | 0.9488±0.0071 | 0.9000±0.0141 | 0.9975±0.0014 | 0.9653±0.0051 |
| 2 | 367.8±1.3038 | 374 | 0.9834±0.0035 | 0.9481±0.0077 | 0.9000±0.0141 | 0.9963±0.0026 | 0.9630±0.0077 |
| 3 | 363.2±1.3038 | 374 | 0.9711±0.0035 | 0.9089±0.0130 | 0.8240±0.0261 | 0.9938±0.0000 | 0.9337±0.0089 |
| 4 | 350.6±1.1402 | 364 | 0.9632±0.0031 | 0.8756±0.0121 | 0.7574±0.0243 | 0.9937±0.0000 | 0.9103±0.0087 |
| 5 | 315.0±1.5811 | 332 | 0.9488±0.0048 | 0.7998±0.0226 | 0.6051±0.0466 | 0.9945±0.0031 | 0.8530±0.0170 |
| 6 | 273.6±0.5477 | 291 | 0.9402±0.0019 | 0.7722±0.0064 | 0.5529±0.0132 | 0.9914±0.0017 | 0.8253±0.0059 |
| 7 | 235.4±0.8944 | 250 | 0.9416±0.0036 | 0.7932±0.0154 | 0.6000±0.0308 | 0.9864±0.0000 | 0.8359±0.0129 |
| 8 | 182.6±1.6733 | 201 | 0.9085±0.0083 | 0.6923±0.0124 | 0.4083±0.0186 | 0.9763±0.0074 | 0.7329±0.0187 |
| 9 | 138.6±0.5477 | 159 | 0.8717±0.0034 | 0.6193±0.0104 | 0.2762±0.0213 | 0.9623±0.0032 | 0.6455±0.0121 |
| 10 | 118.6±1.1402 | 133 | 0.8917±0.0086 | 0.6525±0.0152 | 0.3375±0.0342 | 0.9675±0.0111 | 0.6843±0.0162 |
| 11 | 102.8±1.3038 | 117 | 0.8786±0.0111 | 0.6518±0.0169 | 0.3467±0.0298 | 0.9569±0.0112 | 0.6777±0.0213 |
| 12 | 85.4±0.5477 | 99 | 0.8626±0.0055 | 0.6597±0.0032 | 0.3846±0.0000 | 0.9349±0.0064 | 0.6730±0.0067 |
| 13 | 80.8±0.8367 | 91 | 0.8879±0.0092 | 0.7103±0.0054 | 0.4615±0.0000 | 0.9590±0.0107 | 0.7387±0.0131 |
| 14 | 64.2±1.0954 | 75 | 0.8560±0.0146 | 0.6377±0.0361 | 0.3167±0.0697 | 0.9587±0.0087 | 0.6644±0.0415 |
| 15 | 57.4±0.5477 | 67 | 0.8567±0.0082 | 0.7170±0.0372 | 0.5091±0.0813 | 0.9250±0.0080 | 0.7256±0.0274 |
| 16 | 50.2±2.3875 | 59 | 0.8508±0.0405 | 0.7192±0.0537 | 0.5200±0.0837 | 0.9184±0.0382 | 0.7280±0.0624 |
| 17 | 43.4±0.5477 | 54 | 0.8037±0.0101 | 0.5800±0.0396 | 0.2444±0.0930 | 0.9156±0.0186 | 0.5864±0.0441 |
| 18 | 34.6±0.5477 | 45 | 0.7689±0.0122 | 0.5851±0.0322 | 0.3000±0.0685 | 0.8703±0.0121 | 0.5874±0.0305 |
| 19 | 29.6±0.8944 | 38 | 0.7789±0.0235 | 0.5979±0.0140 | 0.3333±0.0000 | 0.8625±0.0280 | 0.5958±0.0188 |
| 20 | 26.4±0.5477 | 35 | 0.7543±0.0156 | 0.5213±0.0094 | 0.1667±0.0000 | 0.8759±0.0189 | 0.5221±0.0102 |

Table 72: H12O Random Forest with imputation, results of CV, taking best model of grid using accuracy. Predictions from outcome.

| Day | Corrects | Totals | Accuracy | AUC | Sensitivity | Specificity | F1 |
|---|---|---|---|---|---|---|---|
| 1 | 334.6±2.0736 | 374 | 0.8947±0.0055 | 0.6753±0.0157 | 0.3760±0.0329 | 0.9747±0.0059 | 0.7146±0.0163 |
| 2 | 339.8±0.8367 | 374 | 0.9086±0.0022 | 0.7070±0.0037 | 0.4320±0.0110 | 0.9821±0.0040 | 0.7536±0.0024 |
| 3 | 343.6±1.8166 | 374 | 0.9187±0.0049 | 0.7653±0.0134 | 0.5560±0.0261 | 0.9747±0.0034 | 0.8002±0.0129 |
| 4 | 336.8±2.1679 | 364 | 0.9253±0.0060 | 0.7831±0.0105 | 0.5915±0.0178 | 0.9748±0.0050 | 0.8147±0.0133 |
| 5 | 314.4±2.0736 | 332 | 0.9470±0.0062 | 0.8322±0.0197 | 0.6821±0.0389 | 0.9823±0.0044 | 0.8607±0.0172 |
| 6 | 274.6±0.5477 | 291 | 0.9436±0.0019 | 0.8124±0.0064 | 0.6412±0.0132 | 0.9837±0.0017 | 0.8476±0.0054 |
| 7 | 234.4±1.6733 | 250 | 0.9376±0.0067 | 0.7910±0.0307 | 0.6000±0.0626 | 0.9819±0.0032 | 0.8272±0.0238 |
| 8 | 190.0±1.2247 | 201 | 0.9453±0.0061 | 0.8393±0.0268 | 0.7000±0.0543 | 0.9785±0.0025 | 0.8609±0.0191 |
| 9 | 149.2±0.8367 | 159 | 0.9384±0.0053 | 0.8272±0.0199 | 0.6762±0.0398 | 0.9783±0.0000 | 0.8540±0.0153 |
| 10 | 124.8±0.4472 | 133 | 0.9383±0.0034 | 0.8301±0.0192 | 0.6875±0.0442 | 0.9726±0.0072 | 0.8467±0.0087 |
| 11 | 108.8±1.3038 | 117 | 0.9299±0.0111 | 0.8006±0.0304 | 0.6267±0.0596 | 0.9745±0.0088 | 0.8281±0.0283 |
| 12 | 91.2±0.8367 | 99 | 0.9212±0.0085 | 0.7979±0.0177 | 0.6308±0.0344 | 0.9651±0.0082 | 0.8165±0.0177 |
| 13 | 82.4±0.5477 | 91 | 0.9055±0.0060 | 0.7846±0.0035 | 0.6154±0.0000 | 0.9538±0.0070 | 0.7980±0.0091 |
| 14 | 71.6±1.5166 | 75 | 0.9547±0.0202 | 0.9123±0.0391 | 0.8500±0.0697 | 0.9746±0.0142 | 0.9151±0.0386 |
| 15 | 64.0±0.7071 | 67 | 0.9552±0.0106 | 0.9294±0.0217 | 0.8909±0.0407 | 0.9679±0.0080 | 0.9201±0.0193 |
| 16 | 57.4±0.5477 | 59 | 0.9729±0.0093 | 0.9598±0.0274 | 0.9400±0.0548 | 0.9796±0.0000 | 0.9523±0.0171 |
| 17 | 53.6±0.8944 | 54 | 0.9926±0.0166 | 0.9778±0.0497 | 0.9556±0.0994 | 1.0000±0.0000 | 0.9853±0.0328 |
| 18 | 44.8±0.4472 | 45 | 0.9956±0.0099 | 0.9875±0.0280 | 0.9750±0.0559 | 1.0000±0.0000 | 0.9920±0.0179 |
| 19 | 38.0±0.0000 | 38 | 1.0000±0.0000 | 1.0000±0.0000 | 1.0000±0.0000 | 1.0000±0.0000 | 1.0000±0.0000 |
| 20 | 35.0±0.0000 | 35 | 1.0000±0.0000 | 1.0000±0.0000 | 1.0000±0.0000 | 1.0000±0.0000 | 1.0000±0.0000 |

Table 73: H12O Random Forest with imputation, results of CV, taking best model of grid using F1. Predictions from admission.

| Day | Corrects | Totals | Accuracy | AUC | Sensitivity | Specificity | F1 |
|---|---|---|---|---|---|---|---|
| 1 | 368.8±0.8367 | 374 | 0.9861±0.0022 | 0.9548±0.0058 | 0.9120±0.0110 | 0.9975±0.0014 | 0.9690±0.0050 |
| 2 | 367.8±1.3038 | 374 | 0.9834±0.0035 | 0.9481±0.0077 | 0.9000±0.0141 | 0.9963±0.0026 | 0.9630±0.0077 |
| 3 | 363.4±0.8944 | 374 | 0.9717±0.0024 | 0.9109±0.0089 | 0.8280±0.0179 | 0.9938±0.0000 | 0.9351±0.0060 |
| 4 | 350.4±0.5477 | 364 | 0.9626±0.0015 | 0.8734±0.0058 | 0.7532±0.0117 | 0.9937±0.0000 | 0.9088±0.0042 |
| 5 | 314.4±2.3022 | 332 | 0.9470±0.0069 | 0.7921±0.0297 | 0.5897±0.0601 | 0.9945±0.0031 | 0.8463±0.0244 |
| 6 | 274.4±1.3416 | 291 | 0.9430±0.0046 | 0.7789±0.0235 | 0.5647±0.0483 | 0.9930±0.0017 | 0.8328±0.0188 |
| 7 | 235.4±0.8944 | 250 | 0.9416±0.0036 | 0.7932±0.0154 | 0.6000±0.0308 | 0.9864±0.0000 | 0.8359±0.0129 |
| 8 | 182.6±1.3416 | 201 | 0.9085±0.0067 | 0.6959±0.0038 | 0.4167±0.0000 | 0.9751±0.0076 | 0.7354±0.0111 |
| 9 | 139.2±0.8367 | 159 | 0.8755±0.0053 | 0.6335±0.0124 | 0.3048±0.0261 | 0.9623±0.0061 | 0.6615±0.0136 |
| 10 | 119.8±1.3038 | 133 | 0.9008±0.0098 | 0.6792±0.0269 | 0.3875±0.0523 | 0.9709±0.0076 | 0.7144±0.0298 |
| 11 | 103.0±0.7071 | 117 | 0.8803±0.0060 | 0.6584±0.0180 | 0.3600±0.0365 | 0.9569±0.0054 | 0.6840±0.0184 |
| 12 | 85.2±0.4472 | 99 | 0.8606±0.0045 | 0.6586±0.0026 | 0.3846±0.0000 | 0.9326±0.0052 | 0.6705±0.0054 |
| 13 | 80.4±0.5477 | 91 | 0.8835±0.0060 | 0.7077±0.0035 | 0.4615±0.0000 | 0.9538±0.0070 | 0.7324±0.0083 |
| 14 | 64.2±1.0954 | 75 | 0.8560±0.0146 | 0.6377±0.0361 | 0.3167±0.0697 | 0.9587±0.0087 | 0.6644±0.0415 |
| 15 | 57.2±0.4472 | 67 | 0.8537±0.0067 | 0.7080±0.0227 | 0.4909±0.0498 | 0.9250±0.0080 | 0.7185±0.0180 |
| 16 | 50.0±2.4495 | 59 | 0.8475±0.0415 | 0.7171±0.0524 | 0.5200±0.0837 | 0.9143±0.0418 | 0.7242±0.0622 |
| 17 | 42.6±0.8944 | 54 | 0.7889±0.0166 | 0.5356±0.0298 | 0.1556±0.0609 | 0.9156±0.0186 | 0.5367±0.0366 |
| 18 | 34.4±1.1402 | 45 | 0.7644±0.0253 | 0.5824±0.0411 | 0.3000±0.0685 | 0.8649±0.0191 | 0.5848±0.0424 |
| 19 | 29.4±1.3416 | 38 | 0.7737±0.0353 | 0.5948±0.0210 | 0.3333±0.0000 | 0.8562±0.0419 | 0.5921±0.0290 |
| 20 | 26.4±0.5477 | 35 | 0.7543±0.0156 | 0.5213±0.0094 | 0.1667±0.0000 | 0.8759±0.0189 | 0.5221±0.0102 |

Table 74: H12O Random Forest with imputation, results of CV, taking best model of grid using F1. Predictions from outcome.

| Day | Corrects | Totals | Accuracy | AUC | Sensitivity | Specificity | F1 |
|---|---|---|---|---|---|---|---|
| 1 | 334.2±0.8367 | 374 | 0.8936±0.0022 | 0.6663±0.0123 | 0.3560±0.0261 | 0.9765±0.0017 | 0.7063±0.0123 |
| 2 | 339.0±2.2361 | 374 | 0.9064±0.0060 | 0.7041±0.0130 | 0.4280±0.0228 | 0.9802±0.0035 | 0.7489±0.0161 |
| 3 | 344.4±2.3022 | 374 | 0.9209±0.0062 | 0.7649±0.0154 | 0.5520±0.0335 | 0.9778±0.0077 | 0.8031±0.0142 |
| 4 | 339.4±1.5166 | 364 | 0.9324±0.0042 | 0.7963±0.0097 | 0.6128±0.0178 | 0.9798±0.0028 | 0.8313±0.0103 |
| 5 | 314.8±0.8367 | 332 | 0.9482±0.0025 | 0.8262±0.0091 | 0.6667±0.0181 | 0.9857±0.0015 | 0.8612±0.0075 |
| 6 | 276.2±1.3038 | 291 | 0.9491±0.0045 | 0.8283±0.0190 | 0.6706±0.0383 | 0.9860±0.0021 | 0.8631±0.0145 |
| 7 | 237.2±1.4832 | 250 | 0.9488±0.0059 | 0.8332±0.0227 | 0.6828±0.0450 | 0.9837±0.0025 | 0.8633±0.0183 |
| 8 | 190.6±0.5477 | 201 | 0.9483±0.0027 | 0.8410±0.0164 | 0.7000±0.0349 | 0.9819±0.0025 | 0.8671±0.0097 |
| 9 | 148.8±1.0954 | 159 | 0.9358±0.0069 | 0.8177±0.0261 | 0.6571±0.0522 | 0.9783±0.0000 | 0.8464±0.0207 |
| 10 | 125.6±0.8944 | 133 | 0.9444±0.0067 | 0.8227±0.0174 | 0.6625±0.0342 | 0.9829±0.0060 | 0.8550±0.0172 |
| 11 | 109.6±0.8944 | 117 | 0.9368±0.0076 | 0.8102±0.0186 | 0.6400±0.0365 | 0.9804±0.0069 | 0.8430±0.0187 |
| 12 | 90.8±0.4472 | 99 | 0.9172±0.0045 | 0.7760±0.0196 | 0.5846±0.0421 | 0.9674±0.0052 | 0.8010±0.0141 |
| 13 | 83.6±1.1402 | 91 | 0.9187±0.0125 | 0.8179±0.0206 | 0.6769±0.0344 | 0.9590±0.0107 | 0.8287±0.0243 |
| 14 | 71.8±0.8367 | 75 | 0.9573±0.0112 | 0.9071±0.0273 | 0.8333±0.0589 | 0.9810±0.0133 | 0.9184±0.0204 |
| 15 | 64.0±0.7071 | 67 | 0.9552±0.0106 | 0.9367±0.0321 | 0.9091±0.0643 | 0.9643±0.0000 | 0.9209±0.0205 |
| 16 | 57.8±0.4472 | 59 | 0.9797±0.0076 | 0.9798±0.0224 | 0.9800±0.0447 | 0.9796±0.0000 | 0.9648±0.0140 |
| 17 | 53.8±0.4472 | 54 | 0.9963±0.0083 | 0.9889±0.0248 | 0.9778±0.0497 | 1.0000±0.0000 | 0.9930±0.0156 |
| 18 | 44.6±0.5477 | 45 | 0.9911±0.0122 | 0.9750±0.0342 | 0.9500±0.0685 | 1.0000±0.0000 | 0.9840±0.0219 |
| 19 | 38.0±0.0000 | 38 | 1.0000±0.0000 | 1.0000±0.0000 | 1.0000±0.0000 | 1.0000±0.0000 | 1.0000±0.0000 |
| 20 | 35.0±0.0000 | 35 | 1.0000±0.0000 | 1.0000±0.0000 | 1.0000±0.0000 | 1.0000±0.0000 | 1.0000±0.0000 |

Table 75: H12O Random Forest with imputation, with missing flags, results of CV, taking best model of grid using accuracy. Predictions from admission.

| Day | Corrects | Totals | Accuracy | AUC | Sensitivity | Specificity | F1 |
|---|---|---|---|---|---|---|---|
| 1 | 369.8±0.4472 | 374 | 0.9888±0.0012 | 0.9665±0.0045 | 0.9360±0.0089 | 0.9969±0.0000 | 0.9753±0.0027 |
| 2 | 368.2±0.8367 | 374 | 0.9845±0.0022 | 0.9538±0.0082 | 0.9120±0.0179 | 0.9957±0.0028 | 0.9656±0.0050 |
| 3 | 364.2±1.3038 | 374 | 0.9738±0.0035 | 0.9172±0.0153 | 0.8400±0.0316 | 0.9944±0.0014 | 0.9402±0.0089 |
| 4 | 351.4±0.8944 | 364 | 0.9654±0.0025 | 0.8823±0.0088 | 0.7702±0.0178 | 0.9943±0.0014 | 0.9160±0.0065 |
| 5 | 314.4±1.9494 | 332 | 0.9470±0.0059 | 0.7899±0.0215 | 0.5846±0.0421 | 0.9952±0.0019 | 0.8458±0.0202 |
| 6 | 274.6±0.8944 | 291 | 0.9436±0.0031 | 0.7818±0.0130 | 0.5706±0.0263 | 0.9930±0.0017 | 0.8358±0.0108 |
| 7 | 235.2±0.4472 | 250 | 0.9408±0.0018 | 0.7868±0.0114 | 0.5862±0.0244 | 0.9873±0.0020 | 0.8318±0.0076 |
| 8 | 182.8±1.6432 | 201 | 0.9095±0.0082 | 0.6929±0.0273 | 0.4083±0.0543 | 0.9774±0.0056 | 0.7337±0.0289 |
| 9 | 139.8±0.8367 | 159 | 0.8792±0.0053 | 0.6236±0.0199 | 0.2762±0.0398 | 0.9710±0.0000 | 0.6544±0.0233 |
| 10 | 120.2±0.8367 | 133 | 0.9038±0.0063 | 0.6701±0.0130 | 0.3625±0.0280 | 0.9778±0.0076 | 0.7111±0.0151 |
| 11 | 104.6±1.1402 | 117 | 0.8940±0.0097 | 0.6663±0.0200 | 0.3600±0.0365 | 0.9725±0.0082 | 0.7034±0.0246 |
| 12 | 86.6±0.5477 | 99 | 0.8747±0.0055 | 0.6667±0.0032 | 0.3846±0.0000 | 0.9488±0.0064 | 0.6880±0.0071 |
| 13 | 81.2±0.8367 | 91 | 0.8923±0.0092 | 0.7128±0.0054 | 0.4615±0.0000 | 0.9641±0.0107 | 0.7450±0.0132 |
| 14 | 65.0±0.7071 | 75 | 0.8667±0.0094 | 0.6373±0.0348 | 0.3000±0.0745 | 0.9746±0.0087 | 0.6697±0.0376 |
| 15 | 58.4±0.8944 | 67 | 0.8716±0.0133 | 0.7187±0.0257 | 0.4909±0.0498 | 0.9464±0.0126 | 0.7407±0.0267 |
| 16 | 50.8±0.8367 | 59 | 0.8610±0.0142 | 0.7412±0.0259 | 0.5600±0.0548 | 0.9224±0.0171 | 0.7469±0.0238 |
| 17 | 42.6±0.5477 | 54 | 0.7889±0.0101 | 0.5622±0.0357 | 0.2222±0.0786 | 0.9022±0.0122 | 0.5664±0.0390 |
| 18 | 34.8±0.8367 | 45 | 0.7733±0.0186 | 0.5878±0.0313 | 0.3000±0.0685 | 0.8757±0.0242 | 0.5911±0.0310 |
| 19 | 28.4±0.5477 | 38 | 0.7474±0.0144 | 0.5792±0.0086 | 0.3333±0.0000 | 0.8250±0.0171 | 0.5703±0.0111 |
| 20 | 26.2±0.4472 | 35 | 0.7486±0.0128 | 0.5178±0.0077 | 0.1667±0.0000 | 0.8690±0.0154 | 0.5184±0.0084 |

Table 76: H12O Random Forest with imputation, with missing flags, results of CV, taking best model of grid using accuracy. Predictions from outcome.

| Day | Corrects | Totals | Accuracy | AUC | Sensitivity | Specificity | F1 |
|---|---|---|---|---|---|---|---|
| 1 | 332.2±2.1679 | 374 | 0.8882±0.0058 | 0.6987±0.0089 | 0.4400±0.0141 | 0.9574±0.0051 | 0.7249±0.0118 |
| 2 | 337.2±1.6432 | 374 | 0.9016±0.0044 | 0.7166±0.0090 | 0.4640±0.0167 | 0.9691±0.0038 | 0.7512±0.0103 |
| 3 | 342.0±0.7071 | 374 | 0.9144±0.0019 | 0.7730±0.0061 | 0.5800±0.0141 | 0.9660±0.0031 | 0.7979±0.0041 |
| 4 | 338.8±1.9235 | 364 | 0.9308±0.0053 | 0.8135±0.0145 | 0.6553±0.0277 | 0.9716±0.0032 | 0.8351±0.0134 |
| 5 | 311.6±3.2863 | 332 | 0.9386±0.0099 | 0.8163±0.0353 | 0.6564±0.0693 | 0.9761±0.0042 | 0.8397±0.0304 |
| 6 | 274.2±2.5884 | 291 | 0.9423±0.0089 | 0.8244±0.0271 | 0.6706±0.0526 | 0.9782±0.0059 | 0.8490±0.0247 |
| 7 | 237.2±1.3038 | 250 | 0.9488±0.0052 | 0.8422±0.0163 | 0.7034±0.0308 | 0.9810±0.0020 | 0.8662±0.0147 |
| 8 | 190.4±0.5477 | 201 | 0.9473±0.0027 | 0.8476±0.0091 | 0.7167±0.0186 | 0.9785±0.0025 | 0.8674±0.0070 |
| 9 | 147.6±1.1402 | 159 | 0.9283±0.0072 | 0.8093±0.0148 | 0.6476±0.0261 | 0.9710±0.0051 | 0.8320±0.0163 |
| 10 | 124.2±0.8367 | 133 | 0.9338±0.0063 | 0.8005±0.0208 | 0.6250±0.0442 | 0.9761±0.0072 | 0.8285±0.0168 |
| 11 | 109.2±0.8367 | 117 | 0.9333±0.0072 | 0.8082±0.0189 | 0.6400±0.0365 | 0.9765±0.0054 | 0.8366±0.0180 |
| 12 | 91.0±1.2247 | 99 | 0.9192±0.0124 | 0.7772±0.0345 | 0.5846±0.0688 | 0.9698±0.0104 | 0.8043±0.0303 |
| 13 | 84.4±0.8944 | 91 | 0.9275±0.0098 | 0.8359±0.0303 | 0.7077±0.0644 | 0.9641±0.0107 | 0.8466±0.0223 |
| 14 | 72.0±0.7071 | 75 | 0.9600±0.0094 | 0.9290±0.0225 | 0.8833±0.0456 | 0.9746±0.0087 | 0.9260±0.0177 |
| 15 | 64.2±0.4472 | 67 | 0.9582±0.0067 | 0.9458±0.0203 | 0.9273±0.0407 | 0.9643±0.0000 | 0.9269±0.0125 |
| 16 | 57.4±0.5477 | 59 | 0.9729±0.0093 | 0.9598±0.0274 | 0.9400±0.0548 | 0.9796±0.0000 | 0.9523±0.0171 |
| 17 | 53.0±0.7071 | 54 | 0.9815±0.0131 | 0.9444±0.0393 | 0.8889±0.0786 | 1.0000±0.0000 | 0.9644±0.0260 |
| 18 | 44.0±0.0000 | 45 | 0.9778±0.0000 | 0.9375±0.0000 | 0.8750±0.0000 | 1.0000±0.0000 | 0.9600±0.0000 |
| 19 | 37.4±0.5477 | 38 | 0.9842±0.0144 | 0.9500±0.0456 | 0.9000±0.0913 | 1.0000±0.0000 | 0.9681±0.0291 |
| 20 | 34.4±0.5477 | 35 | 0.9829±0.0156 | 0.9500±0.0456 | 0.9000±0.0913 | 1.0000±0.0000 | 0.9676±0.0295 |

Table 77: H12O Random Forest with imputation, with missing flags, results of CV, taking best model of grid using F1. Predictions from admission.

| Day | Corrects | Totals | Accuracy | AUC | Sensitivity | Specificity | F1 |
|---|---|---|---|---|---|---|---|
| 1 | 369.0±0.7071 | 374 | 0.9866±0.0019 | 0.9601±0.0047 | 0.9240±0.0089 | 0.9963±0.0014 | 0.9705±0.0042 |
| 2 | 367.8±0.4472 | 374 | 0.9834±0.0012 | 0.9431±0.0050 | 0.8880±0.0110 | 0.9981±0.0017 | 0.9626±0.0027 |
| 3 | 364.0±1.2247 | 374 | 0.9733±0.0033 | 0.9118±0.0146 | 0.8280±0.0303 | 0.9957±0.0017 | 0.9384±0.0086 |
| 4 | 351.0±1.5811 | 364 | 0.9643±0.0043 | 0.8744±0.0128 | 0.7532±0.0243 | 0.9956±0.0017 | 0.9123±0.0112 |
| 5 | 315.2±1.9235 | 332 | 0.9494±0.0058 | 0.8046±0.0223 | 0.6154±0.0444 | 0.9939±0.0029 | 0.8561±0.0186 |
| 6 | 275.6±1.5166 | 291 | 0.9471±0.0052 | 0.8195±0.0178 | 0.6529±0.0383 | 0.9860±0.0065 | 0.8564±0.0135 |
| 7 | 235.4±1.5166 | 250 | 0.9416±0.0061 | 0.8022±0.0216 | 0.6207±0.0422 | 0.9837±0.0025 | 0.8392±0.0188 |
| 8 | 183.6±2.0736 | 201 | 0.9134±0.0103 | 0.7167±0.0173 | 0.4583±0.0295 | 0.9751±0.0095 | 0.7555±0.0231 |
| 9 | 139.0±2.5495 | 159 | 0.8742±0.0160 | 0.6449±0.0376 | 0.3333±0.0673 | 0.9565±0.0089 | 0.6704±0.0436 |
| 10 | 119.4±0.5477 | 133 | 0.8977±0.0041 | 0.6883±0.0146 | 0.4125±0.0342 | 0.9641±0.0072 | 0.7176±0.0112 |
| 11 | 102.0±0.7071 | 117 | 0.8718±0.0060 | 0.6478±0.0102 | 0.3467±0.0298 | 0.9490±0.0107 | 0.6686±0.0061 |
| 12 | 83.2±0.8367 | 99 | 0.8404±0.0085 | 0.6470±0.0049 | 0.3846±0.0000 | 0.9093±0.0097 | 0.6481±0.0089 |
| 13 | 79.8±0.4472 | 91 | 0.8769±0.0049 | 0.7038±0.0029 | 0.4615±0.0000 | 0.9462±0.0057 | 0.7234±0.0064 |
| 14 | 64.8±1.3038 | 75 | 0.8640±0.0174 | 0.6694±0.0483 | 0.3833±0.0950 | 0.9556±0.0071 | 0.6962±0.0517 |
| 15 | 56.0±1.4142 | 67 | 0.8358±0.0211 | 0.6972±0.0350 | 0.4909±0.0813 | 0.9036±0.0299 | 0.6979±0.0320 |
| 16 | 48.4±1.6733 | 59 | 0.8203±0.0284 | 0.6929±0.0435 | 0.5000±0.0707 | 0.8857±0.0233 | 0.6886±0.0453 |
| 17 | 41.4±2.0736 | 54 | 0.7667±0.0384 | 0.6200±0.0616 | 0.4000±0.0994 | 0.8400±0.0290 | 0.6108±0.0580 |
| 18 | 33.0±0.7071 | 45 | 0.7333±0.0157 | 0.5831±0.0458 | 0.3500±0.1046 | 0.8162±0.0226 | 0.5743±0.0368 |
| 19 | 27.8±0.4472 | 38 | 0.7316±0.0118 | 0.5833±0.0327 | 0.3667±0.0745 | 0.8000±0.0171 | 0.5668±0.0225 |
| 20 | 26.0±0.0000 | 35 | 0.7429±0.0000 | 0.5144±0.0000 | 0.1667±0.0000 | 0.8621±0.0000 | 0.5146±0.0000 |

Table 78: H12O Random Forest with imputation, with missing flags, results of CV, taking best model of grid using F1. Predictions from outcome.

| Day | Corrects | Totals | Accuracy | AUC | Sensitivity | Specificity | F1 |
|---|---|---|---|---|---|---|---|
| 1 | 326.2±1.4832 | 374 | 0.8722±0.0040 | 0.6472±0.0106 | 0.3400±0.0245 | 0.9543±0.0059 | 0.6718±0.0097 |
| 2 | 333.6±2.3022 | 374 | 0.8920±0.0062 | 0.6823±0.0127 | 0.3960±0.0219 | 0.9685±0.0040 | 0.7173±0.0156 |
| 3 | 339.4±0.8944 | 374 | 0.9075±0.0024 | 0.7504±0.0098 | 0.5360±0.0219 | 0.9648±0.0035 | 0.7776±0.0072 |
| 4 | 337.2±0.8367 | 364 | 0.9264±0.0023 | 0.7946±0.0065 | 0.6170±0.0150 | 0.9722±0.0035 | 0.8211±0.0048 |
| 5 | 310.8±1.3038 | 332 | 0.9361±0.0039 | 0.8082±0.0254 | 0.6410±0.0544 | 0.9754±0.0044 | 0.8328±0.0157 |
| 6 | 275.0±2.1213 | 291 | 0.9450±0.0073 | 0.8540±0.0167 | 0.7353±0.0294 | 0.9728±0.0048 | 0.8633±0.0177 |
| 7 | 234.6±0.8944 | 250 | 0.9384±0.0036 | 0.8244±0.0086 | 0.6759±0.0189 | 0.9729±0.0045 | 0.8417±0.0078 |
| 8 | 190.2±0.4472 | 201 | 0.9463±0.0022 | 0.8470±0.0203 | 0.7167±0.0456 | 0.9774±0.0056 | 0.8652±0.0086 |
| 9 | 149.4±0.8944 | 159 | 0.9396±0.0056 | 0.8401±0.0250 | 0.7048±0.0522 | 0.9754±0.0040 | 0.8600±0.0169 |
| 10 | 122.8±0.4472 | 133 | 0.9233±0.0034 | 0.8161±0.0117 | 0.6750±0.0280 | 0.9573±0.0060 | 0.8178±0.0063 |
| 11 | 108.0±0.7071 | 117 | 0.9231±0.0060 | 0.8251±0.0180 | 0.6933±0.0365 | 0.9569±0.0054 | 0.8269±0.0141 |
| 12 | 92.6±0.5477 | 99 | 0.9354±0.0055 | 0.8322±0.0032 | 0.6923±0.0000 | 0.9721±0.0064 | 0.8506±0.0099 |
| 13 | 84.6±1.1402 | 91 | 0.9297±0.0125 | 0.8308±0.0386 | 0.6923±0.0769 | 0.9692±0.0070 | 0.8479±0.0307 |
| 14 | 71.8±0.4472 | 75 | 0.9573±0.0060 | 0.9139±0.0163 | 0.8500±0.0373 | 0.9778±0.0087 | 0.9195±0.0107 |
| 15 | 64.0±0.0000 | 67 | 0.9552±0.0000 | 0.9367±0.0000 | 0.9091±0.0000 | 0.9643±0.0000 | 0.9213±0.0000 |
| 16 | 57.0±0.0000 | 59 | 0.9661±0.0000 | 0.9398±0.0000 | 0.9000±0.0000 | 0.9796±0.0000 | 0.9398±0.0000 |
| 17 | 53.0±0.0000 | 54 | 0.9815±0.0000 | 0.9444±0.0000 | 0.8889±0.0000 | 1.0000±0.0000 | 0.9651±0.0000 |
| 18 | 43.4±0.8944 | 45 | 0.9644±0.0199 | 0.9294±0.0121 | 0.8750±0.0000 | 0.9838±0.0242 | 0.9390±0.0310 |
| 19 | 36.6±0.5477 | 38 | 0.9632±0.0144 | 0.9510±0.0456 | 0.9333±0.0913 | 0.9688±0.0000 | 0.9326±0.0288 |
| 20 | 33.8±0.4472 | 35 | 0.9657±0.0128 | 0.9661±0.0373 | 0.9667±0.0745 | 0.9655±0.0000 | 0.9421±0.0239 |

Table 79: H12O Random Forest with imputation, with missing flags, applying reference values, results of CV, taking best model of grid using accuracy. Predictions from admission.

| Day | Corrects | Totals | Accuracy | AUC | Sensitivity | Specificity | F1 |
|---|---|---|---|---|---|---|---|
| 1 | 364.0±0.7071 | 374 | 0.9733±0.0019 | 0.9355±0.0072 | 0.8840±0.0167 | 0.9870±0.0034 | 0.9415±0.0038 |
| 2 | 363.8±0.8367 | 374 | 0.9727±0.0022 | 0.9200±0.0057 | 0.8480±0.0110 | 0.9920±0.0017 | 0.9385±0.0051 |
| 3 | 361.8±1.3038 | 374 | 0.9674±0.0035 | 0.8966±0.0101 | 0.8000±0.0200 | 0.9932±0.0026 | 0.9245±0.0083 |
| 4 | 347.6±1.5166 | 364 | 0.9549±0.0042 | 0.8545±0.0139 | 0.7191±0.0277 | 0.9899±0.0026 | 0.8896±0.0112 |
| 5 | 314.0±1.5811 | 332 | 0.9458±0.0048 | 0.7937±0.0146 | 0.5949±0.0281 | 0.9925±0.0029 | 0.8452±0.0143 |
| 6 | 273.2±1.3038 | 291 | 0.9388±0.0045 | 0.7714±0.0127 | 0.5529±0.0246 | 0.9899±0.0035 | 0.8224±0.0131 |
| 7 | 232.8±0.8367 | 250 | 0.9312±0.0033 | 0.7634±0.0179 | 0.5448±0.0378 | 0.9819±0.0032 | 0.8044±0.0137 |
| 8 | 185.0±1.4142 | 201 | 0.9204±0.0070 | 0.7279±0.0243 | 0.4750±0.0475 | 0.9808±0.0031 | 0.7714±0.0242 |
| 9 | 139.8±0.4472 | 159 | 0.8792±0.0028 | 0.6559±0.0195 | 0.3524±0.0426 | 0.9594±0.0040 | 0.6833±0.0186 |
| 10 | 118.8±0.4472 | 133 | 0.8932±0.0034 | 0.6965±0.0019 | 0.4375±0.0000 | 0.9556±0.0038 | 0.7184±0.0049 |
| 11 | 101.4±1.5166 | 117 | 0.8667±0.0130 | 0.6676±0.0074 | 0.4000±0.0000 | 0.9353±0.0149 | 0.6801±0.0155 |
| 12 | 86.6±1.1402 | 99 | 0.8747±0.0115 | 0.6928±0.0148 | 0.4462±0.0344 | 0.9395±0.0152 | 0.7061±0.0164 |
| 13 | 81.4±0.5477 | 91 | 0.8945±0.0060 | 0.7333±0.0201 | 0.5077±0.0421 | 0.9590±0.0057 | 0.7590±0.0173 |
| 14 | 62.0±1.4142 | 75 | 0.8267±0.0189 | 0.6540±0.0249 | 0.4000±0.0373 | 0.9079±0.0174 | 0.6618±0.0291 |
| 15 | 55.8±0.8367 | 67 | 0.8328±0.0125 | 0.7174±0.0075 | 0.5455±0.0000 | 0.8893±0.0149 | 0.7083±0.0136 |
| 16 | 47.2±1.0954 | 59 | 0.8000±0.0186 | 0.7045±0.0261 | 0.5600±0.0548 | 0.8490±0.0233 | 0.6813±0.0237 |
| 17 | 43.2±1.3038 | 54 | 0.8000±0.0241 | 0.6489±0.0714 | 0.4222±0.1449 | 0.8756±0.0122 | 0.6431±0.0626 |
| 18 | 33.2±1.3038 | 45 | 0.7378±0.0290 | 0.6152±0.0727 | 0.4250±0.1425 | 0.8054±0.0121 | 0.5980±0.0586 |
| 19 | 27.2±0.4472 | 38 | 0.7158±0.0118 | 0.5604±0.0070 | 0.3333±0.0000 | 0.7875±0.0140 | 0.5470±0.0085 |
| 20 | 25.2±0.4472 | 35 | 0.7200±0.0128 | 0.5006±0.0077 | 0.1667±0.0000 | 0.8345±0.0154 | 0.5006±0.0078 |

Table 80: H12O Random Forest with imputation, with missing flags, applying reference values, results of CV, taking best model of grid using accuracy. Predictions from outcome.

| Day | Corrects | Totals | Accuracy | AUC | Sensitivity | Specificity | F1 |
|---|---|---|---|---|---|---|---|
| 1 | 327.0±0.7071 | 374 | 0.8743±0.0019 | 0.6670±0.0191 | 0.3840±0.0434 | 0.9500±0.0055 | 0.6888±0.0157 |
| 2 | 334.2±1.4832 | 374 | 0.8936±0.0040 | 0.6900±0.0131 | 0.4120±0.0303 | 0.9679±0.0064 | 0.7243±0.0108 |
| 3 | 339.4±1.1402 | 374 | 0.9075±0.0030 | 0.7504±0.0152 | 0.5360±0.0329 | 0.9648±0.0035 | 0.7774±0.0111 |
| 4 | 338.4±1.5166 | 364 | 0.9297±0.0042 | 0.7983±0.0157 | 0.6213±0.0316 | 0.9754±0.0014 | 0.8276±0.0127 |
| 5 | 310.6±2.0736 | 332 | 0.9355±0.0062 | 0.8190±0.0135 | 0.6667±0.0256 | 0.9713±0.0057 | 0.8362±0.0144 |
| 6 | 273.2±0.8367 | 291 | 0.9388±0.0029 | 0.8429±0.0113 | 0.7176±0.0263 | 0.9681±0.0051 | 0.8491±0.0060 |
| 7 | 234.6±1.6733 | 250 | 0.9384±0.0067 | 0.8274±0.0195 | 0.6828±0.0378 | 0.9719±0.0050 | 0.8426±0.0176 |
| 8 | 189.4±1.1402 | 201 | 0.9423±0.0057 | 0.8448±0.0151 | 0.7167±0.0349 | 0.9729±0.0084 | 0.8577±0.0115 |
| 9 | 150.8±0.4472 | 159 | 0.9484±0.0028 | 0.8653±0.0106 | 0.7524±0.0213 | 0.9783±0.0000 | 0.8822±0.0077 |
| 10 | 123.4±0.8944 | 133 | 0.9278±0.0067 | 0.8295±0.0140 | 0.7000±0.0280 | 0.9590±0.0072 | 0.8296±0.0136 |
| 11 | 108.6±1.1402 | 117 | 0.9282±0.0097 | 0.8280±0.0200 | 0.6933±0.0365 | 0.9627±0.0082 | 0.8357±0.0208 |
| 12 | 92.8±1.3038 | 99 | 0.9374±0.0132 | 0.8268±0.0210 | 0.6769±0.0344 | 0.9767±0.0116 | 0.8524±0.0281 |
| 13 | 84.8±1.0954 | 91 | 0.9319±0.0120 | 0.8385±0.0331 | 0.7077±0.0644 | 0.9692±0.0070 | 0.8539±0.0280 |
| 14 | 71.2±0.8367 | 75 | 0.9493±0.0112 | 0.8956±0.0337 | 0.8167±0.0697 | 0.9746±0.0087 | 0.9034±0.0229 |
| 15 | 64.0±0.0000 | 67 | 0.9552±0.0000 | 0.9367±0.0000 | 0.9091±0.0000 | 0.9643±0.0000 | 0.9213±0.0000 |
| 16 | 57.4±0.5477 | 59 | 0.9729±0.0093 | 0.9598±0.0274 | 0.9400±0.0548 | 0.9796±0.0000 | 0.9523±0.0171 |
| 17 | 53.0±0.0000 | 54 | 0.9815±0.0000 | 0.9444±0.0000 | 0.8889±0.0000 | 1.0000±0.0000 | 0.9651±0.0000 |
| 18 | 43.4±0.5477 | 45 | 0.9644±0.0122 | 0.9294±0.0074 | 0.8750±0.0000 | 0.9838±0.0148 | 0.9384±0.0197 |
| 19 | 36.4±0.5477 | 38 | 0.9579±0.0144 | 0.9344±0.0456 | 0.9000±0.0913 | 0.9688±0.0000 | 0.9221±0.0288 |
| 20 | 33.8±0.4472 | 35 | 0.9657±0.0128 | 0.9661±0.0373 | 0.9667±0.0745 | 0.9655±0.0000 | 0.9421±0.0239 |

Table 81: H12O Random Forest with imputation, with missing flags, applying reference values, results of CV, taking best model of grid using F1. Predictions from admission.

| Day | Corrects | Totals | Accuracy | AUC | Sensitivity | Specificity | F1 |
|---|---|---|---|---|---|---|---|
| 1 | 365.2±0.8367 | 374 | 0.9765±0.0022 | 0.9424±0.0049 | 0.8960±0.0089 | 0.9889±0.0017 | 0.9485±0.0049 |
| 2 | 363.2±1.3038 | 374 | 0.9711±0.0035 | 0.9191±0.0116 | 0.8480±0.0228 | 0.9901±0.0014 | 0.9352±0.0083 |
| 3 | 362.6±1.8166 | 374 | 0.9695±0.0049 | 0.9029±0.0152 | 0.8120±0.0303 | 0.9938±0.0031 | 0.9297±0.0116 |
| 4 | 348.8±1.6432 | 364 | 0.9582±0.0045 | 0.8619±0.0176 | 0.7319±0.0356 | 0.9918±0.0017 | 0.8976±0.0126 |
| 5 | 315.6±1.8166 | 332 | 0.9506±0.0055 | 0.8231±0.0260 | 0.6564±0.0532 | 0.9898±0.0024 | 0.8645±0.0189 |
| 6 | 274.6±1.6733 | 291 | 0.9436±0.0058 | 0.7946±0.0175 | 0.6000±0.0335 | 0.9891±0.0033 | 0.8409±0.0172 |
| 7 | 233.8±0.8367 | 250 | 0.9352±0.0033 | 0.7836±0.0122 | 0.5862±0.0244 | 0.9810±0.0020 | 0.8206±0.0107 |
| 8 | 184.2±0.8367 | 201 | 0.9164±0.0042 | 0.7220±0.0216 | 0.4667±0.0456 | 0.9774±0.0040 | 0.7621±0.0177 |
| 9 | 140.0±1.5811 | 159 | 0.8805±0.0099 | 0.6526±0.0319 | 0.3429±0.0621 | 0.9623±0.0032 | 0.6814±0.0345 |
| 10 | 118.6±1.1402 | 133 | 0.8917±0.0086 | 0.6795±0.0287 | 0.4000±0.0559 | 0.9590±0.0038 | 0.7046±0.0297 |
| 11 | 102.0±1.2247 | 117 | 0.8718±0.0105 | 0.6763±0.0164 | 0.4133±0.0298 | 0.9392±0.0107 | 0.6901±0.0185 |
| 12 | 85.0±0.7071 | 99 | 0.8586±0.0071 | 0.6705±0.0325 | 0.4154±0.0688 | 0.9256±0.0064 | 0.6764±0.0255 |
| 13 | 80.4±0.8944 | 91 | 0.8835±0.0098 | 0.7205±0.0215 | 0.4923±0.0421 | 0.9487±0.0091 | 0.7400±0.0211 |
| 14 | 62.6±1.3416 | 75 | 0.8347±0.0179 | 0.6520±0.0219 | 0.3833±0.0456 | 0.9206±0.0224 | 0.6647±0.0244 |
| 15 | 54.0±1.2247 | 67 | 0.8060±0.0183 | 0.6867±0.0429 | 0.5091±0.0813 | 0.8643±0.0098 | 0.6716±0.0355 |
| 16 | 47.6±1.5166 | 59 | 0.8068±0.0257 | 0.7165±0.0297 | 0.5800±0.0447 | 0.8531±0.0266 | 0.6927±0.0320 |
| 17 | 43.0±1.2247 | 54 | 0.7963±0.0227 | 0.6556±0.0437 | 0.4444±0.0786 | 0.8667±0.0157 | 0.6484±0.0410 |
| 18 | 32.8±0.8367 | 45 | 0.7289±0.0186 | 0.5902±0.0446 | 0.3750±0.0884 | 0.8054±0.0121 | 0.5789±0.0376 |
| 19 | 27.6±0.5477 | 38 | 0.7263±0.0144 | 0.5667±0.0086 | 0.3333±0.0000 | 0.8000±0.0171 | 0.5546±0.0104 |
| 20 | 24.8±0.4472 | 35 | 0.7086±0.0128 | 0.4937±0.0077 | 0.1667±0.0000 | 0.8207±0.0154 | 0.4938±0.0075 |

Table 82: H12O Random Forest with imputation, with missing flags, applying reference values, results of CV, taking best model of grid using F1. Predictions from outcome.

| Day | Corrects | Totals | Accuracy | AUC | Sensitivity | Specificity | F1 |
|---|---|---|---|---|---|---|---|
| 1 | 333.6±1.1402 | 374 | 0.8920±0.0030 | 0.6603±0.0140 | 0.3440±0.0297 | 0.9765±0.0028 | 0.6997±0.0145 |
| 2 | 335.2±1.6432 | 374 | 0.8963±0.0044 | 0.6627±0.0065 | 0.3440±0.0167 | 0.9815±0.0065 | 0.7062±0.0069 |
| 3 | 339.6±1.1402 | 374 | 0.9080±0.0030 | 0.7237±0.0144 | 0.4720±0.0303 | 0.9753±0.0022 | 0.7632±0.0124 |
| 4 | 338.0±1.2247 | 364 | 0.9286±0.0034 | 0.7760±0.0091 | 0.5702±0.0178 | 0.9817±0.0026 | 0.8166±0.0089 |
| 5 | 312.4±1.5166 | 332 | 0.9410±0.0046 | 0.7887±0.0094 | 0.5897±0.0181 | 0.9877±0.0046 | 0.8343±0.0113 |
| 6 | 275.0±1.5811 | 291 | 0.9450±0.0054 | 0.7979±0.0176 | 0.6059±0.0335 | 0.9899±0.0021 | 0.8448±0.0170 |
| 7 | 232.8±1.3038 | 250 | 0.9312±0.0052 | 0.7993±0.0086 | 0.6276±0.0154 | 0.9710±0.0052 | 0.8204±0.0113 |
| 8 | 188.8±1.3038 | 201 | 0.9393±0.0065 | 0.8215±0.0159 | 0.6667±0.0295 | 0.9763±0.0047 | 0.8449±0.0163 |
| 9 | 147.6±0.8944 | 159 | 0.9283±0.0056 | 0.7972±0.0032 | 0.6190±0.0000 | 0.9754±0.0065 | 0.8274±0.0101 |
| 10 | 126.2±0.8367 | 133 | 0.9489±0.0063 | 0.8145±0.0261 | 0.6375±0.0523 | 0.9915±0.0000 | 0.8603±0.0211 |
| 11 | 107.2±0.8367 | 117 | 0.9162±0.0072 | 0.7700±0.0189 | 0.5733±0.0365 | 0.9667±0.0054 | 0.7947±0.0182 |
| 12 | 91.8±0.4472 | 99 | 0.9273±0.0045 | 0.8014±0.0294 | 0.6308±0.0644 | 0.9721±0.0064 | 0.8261±0.0182 |
| 13 | 82.8±1.3038 | 91 | 0.9099±0.0143 | 0.8513±0.0084 | 0.7692±0.0000 | 0.9333±0.0167 | 0.8285±0.0207 |
| 14 | 70.4±0.5477 | 75 | 0.9387±0.0073 | 0.9298±0.0043 | 0.9167±0.0000 | 0.9429±0.0087 | 0.8950±0.0109 |
| 15 | 62.2±0.4472 | 67 | 0.9284±0.0067 | 0.8768±0.0178 | 0.8000±0.0407 | 0.9536±0.0098 | 0.8713±0.0120 |
| 16 | 54.8±0.8367 | 59 | 0.9288±0.0142 | 0.8696±0.0418 | 0.7800±0.0837 | 0.9592±0.0000 | 0.8717±0.0294 |
| 17 | 52.6±1.1402 | 54 | 0.9741±0.0211 | 0.9222±0.0633 | 0.8444±0.1267 | 1.0000±0.0000 | 0.9481±0.0443 |
| 18 | 41.0±0.7071 | 45 | 0.9111±0.0157 | 0.8970±0.0403 | 0.8750±0.0884 | 0.9189±0.0191 | 0.8607±0.0248 |
| 19 | 35.8±0.4472 | 38 | 0.9421±0.0118 | 0.9115±0.0327 | 0.8667±0.0745 | 0.9563±0.0171 | 0.8953±0.0199 |
| 20 | 31.0±1.0000 | 35 | 0.8857±0.0286 | 0.8649±0.0172 | 0.8333±0.0000 | 0.8966±0.0345 | 0.8228±0.0352 |

Table 83: H12O Random Forest with missing flags, results of CV, taking best model of grid using accuracy. Predictions from admission.

| Day | Corrects | Totals | Accuracy | AUC | Sensitivity | Specificity | F1 |
|---|---|---|---|---|---|---|---|
| 1 | 356.8±1.4832 | 374 | 0.9540±0.0040 | 0.8889±0.0023 | 0.8000±0.0000 | 0.9778±0.0046 | 0.8984±0.0075 |
| 2 | 361.4±1.3416 | 374 | 0.9663±0.0036 | 0.9044±0.0120 | 0.8200±0.0245 | 0.9889±0.0028 | 0.9237±0.0086 |
| 3 | 357.6±1.1402 | 374 | 0.9561±0.0030 | 0.8664±0.0126 | 0.7440±0.0261 | 0.9889±0.0017 | 0.8971±0.0085 |
| 4 | 349.8±2.1679 | 364 | 0.9610±0.0060 | 0.8634±0.0170 | 0.7319±0.0323 | 0.9950±0.0028 | 0.9033±0.0156 |
| 5 | 313.0±1.2247 | 332 | 0.9428±0.0037 | 0.7853±0.0119 | 0.5795±0.0229 | 0.9911±0.0019 | 0.8361±0.0118 |
| 6 | 269.8±0.8367 | 291 | 0.9271±0.0029 | 0.7240±0.0129 | 0.4588±0.0263 | 0.9891±0.0017 | 0.7775±0.0118 |
| 7 | 231.2±1.0954 | 250 | 0.9248±0.0044 | 0.7597±0.0135 | 0.5448±0.0289 | 0.9747±0.0052 | 0.7925±0.0117 |
| 8 | 180.6±1.6733 | 201 | 0.8985±0.0083 | 0.6758±0.0190 | 0.3833±0.0349 | 0.9684±0.0064 | 0.7090±0.0228 |
| 9 | 140.6±1.1402 | 159 | 0.8843±0.0072 | 0.6265±0.0180 | 0.2762±0.0398 | 0.9768±0.0094 | 0.6609±0.0205 |
| 10 | 118.8±0.8367 | 133 | 0.8932±0.0063 | 0.6426±0.0036 | 0.3125±0.0000 | 0.9726±0.0072 | 0.6774±0.0091 |
| 11 | 104.4±0.5477 | 117 | 0.8923±0.0047 | 0.6653±0.0183 | 0.3600±0.0365 | 0.9706±0.0000 | 0.7005±0.0191 |
| 12 | 85.0±1.0000 | 99 | 0.8586±0.0101 | 0.6378±0.0233 | 0.3385±0.0421 | 0.9372±0.0064 | 0.6529±0.0253 |
| 13 | 80.8±0.8367 | 91 | 0.8879±0.0092 | 0.7103±0.0054 | 0.4615±0.0000 | 0.9590±0.0107 | 0.7387±0.0131 |
| 14 | 66.4±0.5477 | 75 | 0.8853±0.0073 | 0.6619±0.0043 | 0.3333±0.0000 | 0.9905±0.0087 | 0.7089±0.0102 |
| 15 | 60.8±0.8367 | 67 | 0.9075±0.0125 | 0.7839±0.0268 | 0.6000±0.0498 | 0.9679±0.0080 | 0.8130±0.0269 |
| 16 | 48.6±0.5477 | 59 | 0.8237±0.0093 | 0.6710±0.0355 | 0.4400±0.0894 | 0.9020±0.0224 | 0.6746±0.0270 |
| 17 | 44.0±0.7071 | 54 | 0.8148±0.0131 | 0.6044±0.0300 | 0.2889±0.0609 | 0.9200±0.0122 | 0.6164±0.0328 |
| 18 | 34.4±0.8944 | 45 | 0.7644±0.0199 | 0.5628±0.0121 | 0.2500±0.0000 | 0.8757±0.0242 | 0.5670±0.0149 |
| 19 | 28.0±1.2247 | 38 | 0.7368±0.0322 | 0.5729±0.0191 | 0.3333±0.0000 | 0.8125±0.0383 | 0.5629±0.0234 |
| 20 | 26.8±0.8367 | 35 | 0.7657±0.0239 | 0.5282±0.0144 | 0.1667±0.0000 | 0.8897±0.0289 | 0.5299±0.0163 |

Table 84: H12O Random Forest with missing flags, results of CV, taking best model of grid using accuracy. Predictions from outcome.

| Day | Corrects | Totals | Accuracy | AUC | Sensitivity | Specificity | F1 |
|---|---|---|---|---|---|---|---|
| 1 | 330.8±1.4832 | 374 | 0.8845±0.0040 | 0.6796±0.0091 | 0.4000±0.0200 | 0.9593±0.0051 | 0.7078±0.0089 |
| 2 | 329.8±1.3038 | 374 | 0.8818±0.0035 | 0.6764±0.0114 | 0.3960±0.0261 | 0.9568±0.0053 | 0.7028±0.0100 |
| 3 | 339.2±1.3038 | 374 | 0.9070±0.0035 | 0.7670±0.0083 | 0.5760±0.0167 | 0.9580±0.0035 | 0.7851±0.0077 |
| 4 | 339.2±2.5884 | 364 | 0.9319±0.0071 | 0.8177±0.0187 | 0.6638±0.0350 | 0.9716±0.0039 | 0.8384±0.0175 |
| 5 | 313.0±0.7071 | 332 | 0.9428±0.0021 | 0.8298±0.0069 | 0.6821±0.0140 | 0.9775±0.0019 | 0.8524±0.0057 |
| 6 | 276.0±1.2247 | 291 | 0.9485±0.0042 | 0.8509±0.0089 | 0.7235±0.0161 | 0.9782±0.0035 | 0.8687±0.0100 |
| 7 | 233.0±0.7071 | 250 | 0.9320±0.0028 | 0.8327±0.0084 | 0.7034±0.0189 | 0.9620±0.0040 | 0.8337±0.0058 |
| 8 | 187.0±0.7071 | 201 | 0.9303±0.0035 | 0.8416±0.0154 | 0.7250±0.0373 | 0.9582±0.0076 | 0.8367±0.0065 |
| 9 | 148.0±1.2247 | 159 | 0.9308±0.0077 | 0.8350±0.0204 | 0.7048±0.0398 | 0.9652±0.0061 | 0.8446±0.0175 |
| 10 | 124.4±0.8944 | 133 | 0.9353±0.0067 | 0.8446±0.0275 | 0.7250±0.0559 | 0.9641±0.0038 | 0.8460±0.0198 |
| 11 | 104.2±1.3038 | 117 | 0.8906±0.0111 | 0.7724±0.0181 | 0.6133±0.0298 | 0.9314±0.0098 | 0.7635±0.0204 |
| 12 | 91.0±0.7071 | 99 | 0.9192±0.0071 | 0.8098±0.0208 | 0.6615±0.0421 | 0.9581±0.0064 | 0.8180±0.0170 |
| 13 | 81.0±1.0000 | 91 | 0.8901±0.0110 | 0.8397±0.0276 | 0.7692±0.0544 | 0.9103±0.0091 | 0.8003±0.0207 |
| 14 | 70.2±0.4472 | 75 | 0.9360±0.0060 | 0.9282±0.0035 | 0.9167±0.0000 | 0.9397±0.0071 | 0.8911±0.0089 |
| 15 | 62.0±0.0000 | 67 | 0.9254±0.0000 | 0.8823±0.0000 | 0.8182±0.0000 | 0.9464±0.0000 | 0.8688±0.0000 |
| 16 | 55.6±0.5477 | 59 | 0.9424±0.0093 | 0.9096±0.0274 | 0.8600±0.0548 | 0.9592±0.0000 | 0.8997±0.0184 |
| 17 | 51.6±0.8944 | 54 | 0.9556±0.0166 | 0.9378±0.0290 | 0.9111±0.0497 | 0.9644±0.0122 | 0.9228±0.0283 |
| 18 | 39.6±0.5477 | 45 | 0.8800±0.0122 | 0.8584±0.0342 | 0.8250±0.0685 | 0.8919±0.0000 | 0.8166±0.0228 |
| 19 | 36.0±0.7071 | 38 | 0.9474±0.0186 | 0.9146±0.0396 | 0.8667±0.0745 | 0.9625±0.0140 | 0.9035±0.0330 |
| 20 | 30.8±0.4472 | 35 | 0.8800±0.0128 | 0.8615±0.0077 | 0.8333±0.0000 | 0.8897±0.0154 | 0.8147±0.0150 |

Table 85: H12O Random Forest with missing flags, results of CV, taking best model of grid using F1. Predictions from admission.

| Day | Corrects | Totals | Accuracy | AUC | Sensitivity | Specificity | F1 |
|---|---|---|---|---|---|---|---|
| 1 | 354.8±0.4472 | 374 | 0.9487±0.0012 | 0.8976±0.0082 | 0.8280±0.0179 | 0.9673±0.0017 | 0.8910±0.0037 |
| 2 | 361.8±1.6432 | 374 | 0.9674±0.0044 | 0.9101±0.0164 | 0.8320±0.0335 | 0.9883±0.0026 | 0.9266±0.0107 |
| 3 | 359.0±0.7071 | 374 | 0.9599±0.0019 | 0.8974±0.0054 | 0.8120±0.0110 | 0.9827±0.0017 | 0.9105±0.0043 |
| 4 | 351.0±1.5811 | 364 | 0.9643±0.0043 | 0.8998±0.0204 | 0.8128±0.0436 | 0.9868±0.0047 | 0.9169±0.0113 |
| 5 | 315.6±1.1402 | 332 | 0.9506±0.0034 | 0.8298±0.0065 | 0.6718±0.0115 | 0.9877±0.0031 | 0.8671±0.0084 |
| 6 | 274.4±1.3416 | 291 | 0.9430±0.0046 | 0.7993±0.0191 | 0.6118±0.0383 | 0.9868±0.0021 | 0.8413±0.0157 |
| 7 | 229.2±0.8367 | 250 | 0.9168±0.0033 | 0.7762±0.0144 | 0.5931±0.0289 | 0.9593±0.0000 | 0.7881±0.0114 |
| 8 | 180.0±1.0000 | 201 | 0.8955±0.0050 | 0.7138±0.0225 | 0.4750±0.0475 | 0.9525±0.0051 | 0.7305±0.0180 |
| 9 | 139.8±0.8367 | 159 | 0.8792±0.0053 | 0.6559±0.0250 | 0.3524±0.0543 | 0.9594±0.0065 | 0.6830±0.0236 |
| 10 | 117.6±1.3416 | 133 | 0.8842±0.0101 | 0.6698±0.0161 | 0.3875±0.0280 | 0.9521±0.0097 | 0.6909±0.0200 |
| 11 | 100.8±1.7889 | 117 | 0.8615±0.0153 | 0.6590±0.0208 | 0.3867±0.0298 | 0.9314±0.0139 | 0.6699±0.0261 |
| 12 | 85.6±1.1402 | 99 | 0.8646±0.0115 | 0.7001±0.0197 | 0.4769±0.0344 | 0.9233±0.0104 | 0.7015±0.0209 |
| 13 | 78.6±0.5477 | 91 | 0.8637±0.0060 | 0.7154±0.0154 | 0.5077±0.0421 | 0.9231±0.0128 | 0.7179±0.0077 |
| 14 | 65.4±1.5166 | 75 | 0.8720±0.0202 | 0.6877±0.0353 | 0.4167±0.0589 | 0.9587±0.0142 | 0.7184±0.0419 |
| 15 | 57.0±0.7071 | 67 | 0.8507±0.0106 | 0.7573±0.0217 | 0.6182±0.0407 | 0.8964±0.0080 | 0.7428±0.0192 |
| 16 | 47.8±1.3038 | 59 | 0.8102±0.0221 | 0.7106±0.0278 | 0.5600±0.0548 | 0.8612±0.0266 | 0.6915±0.0268 |
| 17 | 42.8±1.0954 | 54 | 0.7926±0.0203 | 0.6356±0.0512 | 0.4000±0.0994 | 0.8711±0.0099 | 0.6320±0.0448 |
| 18 | 32.2±0.8367 | 45 | 0.7156±0.0186 | 0.5821±0.0446 | 0.3750±0.0884 | 0.7892±0.0121 | 0.5688±0.0364 |
| 19 | 24.4±0.8944 | 38 | 0.6421±0.0235 | 0.5167±0.0140 | 0.3333±0.0000 | 0.7000±0.0280 | 0.4973±0.0151 |
| 20 | 24.6±0.5477 | 35 | 0.7029±0.0156 | 0.4902±0.0094 | 0.1667±0.0000 | 0.8138±0.0189 | 0.4904±0.0091 |

Table 86: H12O Random Forest with missing flags, results of CV, taking best model of grid using F1. Predictions from outcome.

| Day | Corrects | Totals | Accuracy | AUC | Sensitivity | Specificity | F1 |
|---|---|---|---|---|---|---|---|
| 1 | 326.8±1.6432 | 374 | 0.8738±0.0044 | 0.6176±0.0096 | 0.2680±0.0179 | 0.9673±0.0035 | 0.6460±0.0120 |
| 2 | 331.4±1.5166 | 374 | 0.8861±0.0041 | 0.6417±0.0098 | 0.3080±0.0179 | 0.9753±0.0022 | 0.6782±0.0123 |
| 3 | 339.4±1.3416 | 374 | 0.9075±0.0036 | 0.7132±0.0132 | 0.4480±0.0268 | 0.9784±0.0022 | 0.7561±0.0127 |
| 4 | 335.0±0.7071 | 364 | 0.9203±0.0019 | 0.7477±0.0085 | 0.5149±0.0178 | 0.9804±0.0014 | 0.7903±0.0072 |
| 5 | 307.0±1.0000 | 332 | 0.9247±0.0030 | 0.7395±0.0142 | 0.4974±0.0292 | 0.9816±0.0019 | 0.7831±0.0121 |
| 6 | 271.2±1.9235 | 291 | 0.9320±0.0066 | 0.8007±0.0133 | 0.6294±0.0263 | 0.9720±0.0070 | 0.8229±0.0149 |
| 7 | 233.0±2.3452 | 250 | 0.9320±0.0094 | 0.8058±0.0201 | 0.6414±0.0393 | 0.9701±0.0094 | 0.8243±0.0218 |
| 8 | 190.0±1.5811 | 201 | 0.9453±0.0079 | 0.8321±0.0244 | 0.6833±0.0475 | 0.9808±0.0051 | 0.8589±0.0214 |
| 9 | 146.6±0.5477 | 159 | 0.9220±0.0034 | 0.7815±0.0125 | 0.5905±0.0261 | 0.9725±0.0032 | 0.8112±0.0098 |
| 10 | 121.2±1.3038 | 133 | 0.9113±0.0098 | 0.7230±0.0294 | 0.4750±0.0559 | 0.9709±0.0047 | 0.7564±0.0298 |
| 11 | 108.0±0.7071 | 117 | 0.9231±0.0060 | 0.7853±0.0035 | 0.6000±0.0000 | 0.9706±0.0069 | 0.8118±0.0105 |
| 12 | 91.8±1.4832 | 99 | 0.9273±0.0150 | 0.8080±0.0367 | 0.6462±0.0688 | 0.9698±0.0104 | 0.8291±0.0368 |
| 13 | 83.8±0.4472 | 91 | 0.9209±0.0049 | 0.8321±0.0153 | 0.7077±0.0344 | 0.9564±0.0070 | 0.8363±0.0098 |
| 14 | 70.2±1.3038 | 75 | 0.9360±0.0174 | 0.9079±0.0404 | 0.8667±0.0745 | 0.9492±0.0071 | 0.8866±0.0327 |
| 15 | 62.0±0.7071 | 67 | 0.9254±0.0106 | 0.8677±0.0246 | 0.7818±0.0498 | 0.9536±0.0098 | 0.8649±0.0197 |
| 16 | 54.4±0.8944 | 59 | 0.9220±0.0152 | 0.8814±0.0269 | 0.8200±0.0447 | 0.9429±0.0091 | 0.8668±0.0259 |
| 17 | 50.4±0.8944 | 54 | 0.9333±0.0166 | 0.8800±0.0461 | 0.8000±0.0930 | 0.9600±0.0099 | 0.8792±0.0337 |
| 18 | 41.6±0.5477 | 45 | 0.9244±0.0122 | 0.9051±0.0074 | 0.8750±0.0000 | 0.9351±0.0148 | 0.8792±0.0165 |
| 19 | 35.8±0.4472 | 38 | 0.9421±0.0118 | 0.9115±0.0327 | 0.8667±0.0745 | 0.9563±0.0171 | 0.8953±0.0199 |
| 20 | 31.4±0.5477 | 35 | 0.8971±0.0156 | 0.8718±0.0094 | 0.8333±0.0000 | 0.9103±0.0189 | 0.8362±0.0202 |

Table 87: H12O Random Forest applying laboratory reference values, results of CV, taking best model of grid using accuracy. Predictions from admission.

| Day | Corrects | Totals | Accuracy | AUC | Sensitivity | Specificity | F1 |
|---|---|---|---|---|---|---|---|
| 1 | 357.2±1.0954 | 374 | 0.9551±0.0029 | 0.8810±0.0111 | 0.7800±0.0245 | 0.9821±0.0040 | 0.8985±0.0068 |
| 2 | 356.8±1.3038 | 374 | 0.9540±0.0035 | 0.8720±0.0117 | 0.7600±0.0245 | 0.9840±0.0034 | 0.8945±0.0084 |
| 3 | 356.2±1.3038 | 374 | 0.9524±0.0035 | 0.8491±0.0148 | 0.7080±0.0303 | 0.9901±0.0014 | 0.8859±0.0102 |
| 4 | 345.0±2.4495 | 364 | 0.9478±0.0067 | 0.8251±0.0230 | 0.6596±0.0451 | 0.9905±0.0022 | 0.8677±0.0195 |
| 5 | 309.2±1.0954 | 332 | 0.9313±0.0033 | 0.7521±0.0137 | 0.5179±0.0281 | 0.9863±0.0024 | 0.8005±0.0118 |
| 6 | 268.4±1.1402 | 291 | 0.9223±0.0039 | 0.6957±0.0135 | 0.4000±0.0263 | 0.9914±0.0017 | 0.7517±0.0150 |
| 7 | 229.4±1.3416 | 250 | 0.9176±0.0054 | 0.7107±0.0150 | 0.4414±0.0289 | 0.9801±0.0040 | 0.7543±0.0165 |
| 8 | 183.4±0.8944 | 201 | 0.9124±0.0044 | 0.6802±0.0025 | 0.3750±0.0000 | 0.9853±0.0051 | 0.7289±0.0075 |
| 9 | 138.0±1.2247 | 159 | 0.8679±0.0077 | 0.6211±0.0183 | 0.2857±0.0337 | 0.9565±0.0051 | 0.6449±0.0220 |
| 10 | 118.8±0.8367 | 133 | 0.8932±0.0063 | 0.6426±0.0036 | 0.3125±0.0000 | 0.9726±0.0072 | 0.6774±0.0091 |
| 11 | 101.6±1.5166 | 117 | 0.8684±0.0130 | 0.6402±0.0259 | 0.3333±0.0471 | 0.9471±0.0112 | 0.6599±0.0304 |
| 12 | 86.8±1.3038 | 99 | 0.8768±0.0132 | 0.6679±0.0400 | 0.3846±0.0769 | 0.9512±0.0052 | 0.6896±0.0410 |
| 13 | 80.2±0.8367 | 91 | 0.8813±0.0092 | 0.7128±0.0146 | 0.4769±0.0344 | 0.9487±0.0128 | 0.7332±0.0139 |
| 14 | 62.6±0.8944 | 75 | 0.8347±0.0119 | 0.6587±0.0282 | 0.4000±0.0697 | 0.9175±0.0207 | 0.6686±0.0222 |
| 15 | 56.0±1.5811 | 67 | 0.8358±0.0236 | 0.6972±0.0438 | 0.4909±0.0813 | 0.9036±0.0204 | 0.6983±0.0432 |
| 16 | 48.4±1.1402 | 59 | 0.8203±0.0193 | 0.6610±0.0526 | 0.4200±0.1095 | 0.9020±0.0171 | 0.6654±0.0486 |
| 17 | 43.4±0.8944 | 54 | 0.8037±0.0166 | 0.5978±0.0337 | 0.2889±0.0609 | 0.9067±0.0099 | 0.6067±0.0366 |
| 18 | 34.8±0.8367 | 45 | 0.7733±0.0186 | 0.6074±0.0295 | 0.3500±0.0559 | 0.8649±0.0191 | 0.6082±0.0302 |
| 19 | 28.6±0.8944 | 38 | 0.7526±0.0235 | 0.5958±0.0430 | 0.3667±0.0745 | 0.8250±0.0171 | 0.5838±0.0379 |
| 20 | 26.2±0.4472 | 35 | 0.7486±0.0128 | 0.5310±0.0373 | 0.2000±0.0745 | 0.8621±0.0000 | 0.5313±0.0371 |

Table 88: H12O Random Forest applying laboratory reference values, results of CV, taking best model of grid using accuracy. Predictions from outcome.

| Day | Corrects | Totals | Accuracy | AUC | Sensitivity | Specificity | F1 |
|---|---|---|---|---|---|---|---|
| 1 | 327.2±1.7889 | 374 | 0.8749±0.0048 | 0.6791±0.0155 | 0.4120±0.0303 | 0.9463±0.0017 | 0.6985±0.0151 |
| 2 | 330.0±1.0000 | 374 | 0.8824±0.0027 | 0.7038±0.0131 | 0.4600±0.0283 | 0.9475±0.0031 | 0.7220±0.0105 |
| 3 | 331.6±1.9494 | 374 | 0.8866±0.0052 | 0.7350±0.0060 | 0.5280±0.0110 | 0.9420±0.0059 | 0.7449±0.0081 |
| 4 | 332.0±1.8708 | 364 | 0.9121±0.0051 | 0.7810±0.0129 | 0.6043±0.0243 | 0.9577±0.0036 | 0.7948±0.0122 |
| 5 | 305.0±1.5811 | 332 | 0.9187±0.0048 | 0.8028±0.0150 | 0.6513±0.0292 | 0.9543±0.0031 | 0.8034±0.0125 |
| 6 | 266.6±1.1402 | 291 | 0.9162±0.0039 | 0.8198±0.0066 | 0.6941±0.0161 | 0.9455±0.0055 | 0.8057±0.0061 |
| 7 | 226.4±1.8166 | 250 | 0.9056±0.0073 | 0.8118±0.0155 | 0.6897±0.0345 | 0.9339±0.0094 | 0.7875±0.0125 |
| 8 | 183.2±1.9235 | 201 | 0.9114±0.0096 | 0.8273±0.0094 | 0.7167±0.0186 | 0.9379±0.0113 | 0.8044±0.0153 |
| 9 | 143.4±1.1402 | 159 | 0.9019±0.0072 | 0.7941±0.0077 | 0.6476±0.0261 | 0.9406±0.0119 | 0.7896±0.0070 |
| 10 | 120.6±0.5477 | 133 | 0.9068±0.0041 | 0.8067±0.0113 | 0.6750±0.0280 | 0.9385±0.0072 | 0.7909±0.0062 |
| 11 | 105.0±0.7071 | 117 | 0.8974±0.0060 | 0.8104±0.0160 | 0.6933±0.0365 | 0.9275±0.0088 | 0.7872±0.0105 |
| 12 | 90.2±0.8367 | 99 | 0.9111±0.0085 | 0.8378±0.0332 | 0.7385±0.0688 | 0.9372±0.0064 | 0.8165±0.0210 |
| 13 | 82.8±0.8367 | 91 | 0.9099±0.0092 | 0.8449±0.0178 | 0.7538±0.0344 | 0.9359±0.0091 | 0.8260±0.0167 |
| 14 | 70.6±0.5477 | 75 | 0.9413±0.0073 | 0.9516±0.0228 | 0.9667±0.0456 | 0.9365±0.0000 | 0.9021±0.0137 |
| 15 | 62.0±0.7071 | 67 | 0.9254±0.0106 | 0.8896±0.0217 | 0.8364±0.0407 | 0.9429±0.0080 | 0.8705±0.0181 |
| 16 | 55.0±1.0000 | 59 | 0.9322±0.0169 | 0.9035±0.0314 | 0.8600±0.0548 | 0.9469±0.0112 | 0.8850±0.0290 |
| 17 | 49.2±0.4472 | 54 | 0.9111±0.0083 | 0.9022±0.0050 | 0.8889±0.0000 | 0.9156±0.0099 | 0.8572±0.0112 |
| 18 | 40.2±0.8367 | 45 | 0.8933±0.0186 | 0.8861±0.0113 | 0.8750±0.0000 | 0.8973±0.0226 | 0.8392±0.0227 |
| 19 | 35.6±0.5477 | 38 | 0.9368±0.0144 | 0.9219±0.0433 | 0.9000±0.0913 | 0.9437±0.0140 | 0.8895±0.0266 |
| 20 | 31.0±0.7071 | 35 | 0.8857±0.0202 | 0.8649±0.0122 | 0.8333±0.0000 | 0.8966±0.0244 | 0.8221±0.0249 |

Table 89: H12O Random Forest applying laboratory reference values, results of CV, taking best model of grid using F1. Predictions from admission.

| Day | Corrects | Totals | Accuracy | AUC | Sensitivity | Specificity | F1 |
|---|---|---|---|---|---|---|---|
| 1 | 354.2±0.8367 | 374 | 0.9471±0.0022 | 0.8883±0.0058 | 0.8080±0.0110 | 0.9685±0.0014 | 0.8863±0.0049 |
| 2 | 357.4±0.8944 | 374 | 0.9556±0.0024 | 0.8881±0.0078 | 0.7960±0.0167 | 0.9802±0.0028 | 0.9010±0.0054 |
| 3 | 359.4±1.1402 | 374 | 0.9610±0.0030 | 0.9081±0.0045 | 0.8360±0.0089 | 0.9802±0.0035 | 0.9145±0.0060 |
| 4 | 343.2±0.4472 | 364 | 0.9429±0.0012 | 0.8621±0.0142 | 0.7532±0.0323 | 0.9710±0.0041 | 0.8700±0.0053 |
| 5 | 314.8±1.4832 | 332 | 0.9482±0.0045 | 0.8351±0.0118 | 0.6872±0.0215 | 0.9829±0.0024 | 0.8640±0.0120 |
| 6 | 271.6±1.9494 | 291 | 0.9333±0.0067 | 0.7785±0.0284 | 0.5765±0.0573 | 0.9805±0.0028 | 0.8154±0.0237 |
| 7 | 230.6±1.3416 | 250 | 0.9224±0.0054 | 0.7674±0.0095 | 0.5655±0.0189 | 0.9692±0.0059 | 0.7926±0.0113 |
| 8 | 177.8±2.2804 | 201 | 0.8846±0.0113 | 0.6859±0.0127 | 0.4250±0.0186 | 0.9469±0.0117 | 0.7020±0.0189 |
| 9 | 138.2±0.8367 | 159 | 0.8692±0.0053 | 0.6663±0.0110 | 0.3905±0.0213 | 0.9420±0.0051 | 0.6834±0.0113 |
| 10 | 115.6±0.8944 | 133 | 0.8692±0.0067 | 0.6667±0.0167 | 0.4000±0.0342 | 0.9333±0.0072 | 0.6749±0.0157 |
| 11 | 97.0±1.2247 | 117 | 0.8291±0.0105 | 0.6631±0.0233 | 0.4400±0.0596 | 0.8863±0.0178 | 0.6483±0.0156 |
| 12 | 80.0±1.7321 | 99 | 0.8081±0.0175 | 0.7002±0.0148 | 0.5538±0.0344 | 0.8465±0.0224 | 0.6581±0.0168 |
| 13 | 72.6±1.6733 | 91 | 0.7978±0.0184 | 0.6897±0.0107 | 0.5385±0.0000 | 0.8410±0.0215 | 0.6550±0.0176 |
| 14 | 58.0±1.0000 | 75 | 0.7733±0.0133 | 0.6492±0.0201 | 0.4667±0.0456 | 0.8317±0.0181 | 0.6286±0.0162 |
| 15 | 52.8±0.8367 | 67 | 0.7881±0.0125 | 0.7198±0.0213 | 0.6182±0.0407 | 0.8214±0.0126 | 0.6777±0.0175 |
| 16 | 43.4±0.5477 | 59 | 0.7356±0.0093 | 0.6418±0.0056 | 0.5000±0.0000 | 0.7837±0.0112 | 0.6110±0.0077 |
| 17 | 39.2±1.3038 | 54 | 0.7259±0.0241 | 0.5778±0.0503 | 0.3556±0.0930 | 0.8000±0.0157 | 0.5651±0.0433 |
| 18 | 31.0±1.2247 | 45 | 0.6889±0.0272 | 0.5757±0.0275 | 0.4000±0.0559 | 0.7514±0.0352 | 0.5561±0.0233 |
| 19 | 25.0±0.7071 | 38 | 0.6579±0.0186 | 0.5667±0.0450 | 0.4333±0.0913 | 0.7000±0.0171 | 0.5298±0.0298 |
| 20 | 25.8±0.4472 | 35 | 0.7371±0.0128 | 0.5374±0.0400 | 0.2333±0.0913 | 0.8414±0.0189 | 0.5352±0.0369 |

Table 90: HM Random Forest applying laboratory reference values, results of CV, taking best model of grid using F1. Predictions from admission.

##### 8.6.2 SVC

| Day | Corrects | Totals | Accuracy | AUC | Sensitivity | Specificity | F1 |
|---|---|---|---|---|---|---|---|
| 1 | 321.8±2.7857 | 374 | 0.8604±0.0074 | 0.8668±0.0054 | 0.4640±0.0294 | 0.9216±0.0046 | 0.6951±0.0164 |
| 2 | 325.8±3.5440 | 374 | 0.8711±0.0095 | 0.8820±0.0110 | 0.6280±0.0240 | 0.9086±0.0075 | 0.7452±0.0165 |
| 3 | 331.8±1.7205 | 374 | 0.8872±0.0046 | 0.8766±0.0093 | 0.6320±0.0240 | 0.9265±0.0023 | 0.7669±0.0105 |
| 4 | 325.8±2.4000 | 364 | 0.8951±0.0066 | 0.8955±0.0081 | 0.7106±0.0217 | 0.9224±0.0081 | 0.7875±0.0100 |
| 5 | 305.0±1.0954 | 332 | 0.9187±0.0033 | 0.9410±0.0050 | 0.7538±0.0126 | 0.9406±0.0051 | 0.8194±0.0042 |
| 6 | 270.8±2.0396 | 291 | 0.9306±0.0070 | 0.9416±0.0052 | 0.8059±0.0478 | 0.9471±0.0053 | 0.8452±0.0168 |
| 7 | 224.8±1.1662 | 250 | 0.8992±0.0047 | 0.9416±0.0097 | 0.7379±0.0352 | 0.9204±0.0084 | 0.7855±0.0067 |
| 8 | 177.6±1.8547 | 201 | 0.8836±0.0092 | 0.9078±0.0106 | 0.7333±0.0204 | 0.9040±0.0095 | 0.7665±0.0142 |
| 9 | 134.2±1.7205 | 159 | 0.8440±0.0108 | 0.8738±0.0102 | 0.6286±0.0467 | 0.8768±0.0079 | 0.7113±0.0203 |
| 10 | 109.4±1.0198 | 133 | 0.8226±0.0077 | 0.8614±0.0168 | 0.6250±0.0000 | 0.8496±0.0087 | 0.6764±0.0079 |
| 11 | 97.0±2.0976 | 117 | 0.8291±0.0179 | 0.8642±0.0115 | 0.5733±0.0533 | 0.8667±0.0147 | 0.6807±0.0294 |
| 12 | 85.0±1.0954 | 99 | 0.8586±0.0111 | 0.8986±0.0195 | 0.6769±0.0576 | 0.8860±0.0114 | 0.7362±0.0201 |
| 13 | 75.6±2.6533 | 91 | 0.8308±0.0292 | 0.8966±0.0183 | 0.7077±0.0576 | 0.8513±0.0385 | 0.7214±0.0304 |
| 14 | 67.0±1.4142 | 75 | 0.8933±0.0189 | 0.9659±0.0104 | 0.9000±0.0333 | 0.8921±0.0254 | 0.8324±0.0227 |
| 15 | 58.8±1.1662 | 67 | 0.8776±0.0174 | 0.9643±0.0118 | 0.8909±0.0364 | 0.8750±0.0160 | 0.8142±0.0240 |
| 16 | 53.4±1.0198 | 59 | 0.9051±0.0173 | 0.9780±0.0121 | 0.9800±0.0400 | 0.8898±0.0208 | 0.8591±0.0226 |
| 17 | 49.4±1.3565 | 54 | 0.9148±0.0251 | 0.9664±0.0112 | 0.9333±0.0544 | 0.9111±0.0281 | 0.8670±0.0355 |
| 18 | 42.0±0.0000 | 45 | 0.9333±0.0000 | 0.9804±0.0072 | 0.9250±0.0612 | 0.9351±0.0132 | 0.8947±0.0043 |
| 19 | 34.6±1.3565 | 38 | 0.9105±0.0357 | 0.9823±0.0071 | 0.9000±0.0816 | 0.9125±0.0306 | 0.8542±0.0563 |
| 20 | 33.6±0.4899 | 35 | 0.9600±0.0140 | 0.9943±0.0051 | 1.0000±0.0000 | 0.9517±0.0169 | 0.9359±0.0206 |

Table 91: H12O SVC baseline results of CV using base features, taking best model of grid using accuracy. Predictions from admission.

| Day | Corrects | Totals | Accuracy | AUC | Sensitivity | Specificity | F1 |
|---|---|---|---|---|---|---|---|
| 1 | 365.2±1.1662 | 374 | 0.9765±0.0031 | 0.9893±0.0056 | 0.9240±0.0150 | 0.9846±0.0052 | 0.9498±0.0059 |
| 2 | 360.4±1.6248 | 374 | 0.9636±0.0043 | 0.9912±0.0029 | 0.8880±0.0204 | 0.9753±0.0034 | 0.9231±0.0091 |
| 3 | 358.2±2.4000 | 374 | 0.9578±0.0064 | 0.9716±0.0024 | 0.8600±0.0335 | 0.9728±0.0030 | 0.9101±0.0142 |
| 4 | 346.8±2.6382 | 364 | 0.9527±0.0072 | 0.9744±0.0042 | 0.9149±0.0356 | 0.9584±0.0046 | 0.9028±0.0153 |
| 5 | 308.4±2.0591 | 332 | 0.9289±0.0062 | 0.9597±0.0047 | 0.7487±0.0377 | 0.9529±0.0073 | 0.8358±0.0129 |
| 6 | 262.4±3.7202 | 291 | 0.9017±0.0128 | 0.9447±0.0094 | 0.7118±0.0343 | 0.9268±0.0133 | 0.7864±0.0222 |
| 7 | 226.6±2.5768 | 250 | 0.9064±0.0103 | 0.9200±0.0036 | 0.7241±0.0378 | 0.9303±0.0074 | 0.7943±0.0213 |
| 8 | 173.4±2.5768 | 201 | 0.8627±0.0128 | 0.8464±0.0213 | 0.6417±0.0333 | 0.8927±0.0129 | 0.7238±0.0201 |
| 9 | 131.6±1.3565 | 159 | 0.8277±0.0085 | 0.8093±0.0075 | 0.5429±0.0381 | 0.8710±0.0054 | 0.6759±0.0170 |
| 10 | 109.6±1.3565 | 133 | 0.8241±0.0102 | 0.7903±0.0270 | 0.4500±0.0829 | 0.8752±0.0087 | 0.6381±0.0297 |
| 11 | 94.0±2.7568 | 117 | 0.8034±0.0236 | 0.6901±0.0506 | 0.4000±0.0943 | 0.8627±0.0164 | 0.6134±0.0445 |
| 12 | 73.0±1.8974 | 99 | 0.7374±0.0192 | 0.6739±0.0136 | 0.2923±0.0576 | 0.8047±0.0186 | 0.5340±0.0257 |
| 13 | 71.6±0.4899 | 91 | 0.7868±0.0054 | 0.7199±0.0354 | 0.5077±0.0377 | 0.8333±0.0081 | 0.6373±0.0106 |
| 14 | 59.2±2.0396 | 75 | 0.7893±0.0272 | 0.7011±0.0429 | 0.4167±0.1394 | 0.8603±0.0156 | 0.6275±0.0587 |
| 15 | 48.0±1.0954 | 67 | 0.7164±0.0163 | 0.5558±0.0216 | 0.2182±0.0445 | 0.8143±0.0182 | 0.5143±0.0214 |
| 16 | 41.6±1.0198 | 59 | 0.7051±0.0173 | 0.4461±0.0469 | 0.1200±0.0400 | 0.8245±0.0208 | 0.4716±0.0200 |
| 17 | 39.0±1.2649 | 54 | 0.7222±0.0234 | 0.5348±0.0461 | 0.2000±0.0444 | 0.8267±0.0259 | 0.5129±0.0272 |
| 18 | 32.4±1.9596 | 45 | 0.7200±0.0435 | 0.7108±0.0392 | 0.2250±0.0500 | 0.8270±0.0530 | 0.5264±0.0391 |
| 19 | 26.6±1.7436 | 38 | 0.7000±0.0459 | 0.5000±0.0401 | 0.1000±0.0816 | 0.8125±0.0395 | 0.4599±0.0556 |
| 20 | 26.6±1.8547 | 35 | 0.7600±0.0530 | 0.5966±0.0431 | 0.2000±0.0667 | 0.8759±0.0560 | 0.5429±0.0595 |

Table 92: H12O SVC baseline results of CV using base features, taking best model of grid using accuracy. Predictions from outcome.

| Day | Corrects | Totals | Accuracy | AUC | Sensitivity | Specificity | F1 |
|---|---|---|---|---|---|---|---|
| 1 | 321.8±2.7857 | 374 | 0.8604±0.0074 | 0.8668±0.0054 | 0.4640±0.0294 | 0.9216±0.0046 | 0.6951±0.0164 |
| 2 | 325.8±3.5440 | 374 | 0.8711±0.0095 | 0.8820±0.0110 | 0.6280±0.0240 | 0.9086±0.0075 | 0.7452±0.0165 |
| 3 | 331.8±1.7205 | 374 | 0.8872±0.0046 | 0.8766±0.0093 | 0.6320±0.0240 | 0.9265±0.0023 | 0.7669±0.0105 |
| 4 | 325.8±2.4000 | 364 | 0.8951±0.0066 | 0.8955±0.0081 | 0.7106±0.0217 | 0.9224±0.0081 | 0.7875±0.0100 |
| 5 | 305.0±1.0954 | 332 | 0.9187±0.0033 | 0.9410±0.0050 | 0.7538±0.0126 | 0.9406±0.0051 | 0.8194±0.0042 |
| 6 | 270.8±2.0396 | 291 | 0.9306±0.0070 | 0.9416±0.0052 | 0.8059±0.0478 | 0.9471±0.0053 | 0.8452±0.0168 |
| 7 | 224.8±1.1662 | 250 | 0.8992±0.0047 | 0.9416±0.0097 | 0.7379±0.0352 | 0.9204±0.0084 | 0.7855±0.0067 |
| 8 | 177.6±1.8547 | 201 | 0.8836±0.0092 | 0.9078±0.0106 | 0.7333±0.0204 | 0.9040±0.0095 | 0.7665±0.0142 |
| 9 | 134.2±1.7205 | 159 | 0.8440±0.0108 | 0.8738±0.0102 | 0.6286±0.0467 | 0.8768±0.0079 | 0.7113±0.0203 |
| 10 | 109.4±1.0198 | 133 | 0.8226±0.0077 | 0.8614±0.0168 | 0.6250±0.0000 | 0.8496±0.0087 | 0.6764±0.0079 |
| 11 | 97.0±2.0976 | 117 | 0.8291±0.0179 | 0.8642±0.0115 | 0.5733±0.0533 | 0.8667±0.0147 | 0.6807±0.0294 |
| 12 | 85.0±1.0954 | 99 | 0.8586±0.0111 | 0.8986±0.0195 | 0.6769±0.0576 | 0.8860±0.0114 | 0.7362±0.0201 |
| 13 | 75.6±2.6533 | 91 | 0.8308±0.0292 | 0.8966±0.0183 | 0.7077±0.0576 | 0.8513±0.0385 | 0.7214±0.0304 |
| 14 | 67.0±1.4142 | 75 | 0.8933±0.0189 | 0.9659±0.0104 | 0.9000±0.0333 | 0.8921±0.0254 | 0.8324±0.0227 |
| 15 | 58.8±1.1662 | 67 | 0.8776±0.0174 | 0.9643±0.0118 | 0.8909±0.0364 | 0.8750±0.0160 | 0.8142±0.0240 |
| 16 | 53.4±1.0198 | 59 | 0.9051±0.0173 | 0.9780±0.0121 | 0.9800±0.0400 | 0.8898±0.0208 | 0.8591±0.0226 |
| 17 | 49.4±1.3565 | 54 | 0.9148±0.0251 | 0.9664±0.0112 | 0.9333±0.0544 | 0.9111±0.0281 | 0.8670±0.0355 |
| 18 | 42.0±0.0000 | 45 | 0.9333±0.0000 | 0.9804±0.0072 | 0.9250±0.0612 | 0.9351±0.0132 | 0.8947±0.0043 |
| 19 | 34.6±1.3565 | 38 | 0.9105±0.0357 | 0.9823±0.0071 | 0.9000±0.0816 | 0.9125±0.0306 | 0.8542±0.0563 |
| 20 | 33.6±0.4899 | 35 | 0.9600±0.0140 | 0.9943±0.0051 | 1.0000±0.0000 | 0.9517±0.0169 | 0.9359±0.0206 |

Table 93: H12O SVC baseline results of CV using base features, taking best model of grid using F1. Predictions from admission.

| Day | Corrects | Totals | Accuracy | AUC | Sensitivity | Specificity | F1 |
|---|---|---|---|---|---|---|---|
| 1 | 365.2±1.1662 | 374 | 0.9765±0.0031 | 0.9893±0.0056 | 0.9240±0.0150 | 0.9846±0.0052 | 0.9498±0.0059 |
| 2 | 360.4±1.6248 | 374 | 0.9636±0.0043 | 0.9912±0.0029 | 0.8880±0.0204 | 0.9753±0.0034 | 0.9231±0.0091 |
| 3 | 358.2±2.4000 | 374 | 0.9578±0.0064 | 0.9716±0.0024 | 0.8600±0.0335 | 0.9728±0.0030 | 0.9101±0.0142 |
| 4 | 346.8±2.6382 | 364 | 0.9527±0.0072 | 0.9744±0.0042 | 0.9149±0.0356 | 0.9584±0.0046 | 0.9028±0.0153 |
| 5 | 308.4±2.0591 | 332 | 0.9289±0.0062 | 0.9597±0.0047 | 0.7487±0.0377 | 0.9529±0.0073 | 0.8358±0.0129 |
| 6 | 262.4±3.7202 | 291 | 0.9017±0.0128 | 0.9447±0.0094 | 0.7118±0.0343 | 0.9268±0.0133 | 0.7864±0.0222 |
| 7 | 226.6±2.5768 | 250 | 0.9064±0.0103 | 0.9200±0.0036 | 0.7241±0.0378 | 0.9303±0.0074 | 0.7943±0.0213 |
| 8 | 173.4±2.5768 | 201 | 0.8627±0.0128 | 0.8464±0.0213 | 0.6417±0.0333 | 0.8927±0.0129 | 0.7238±0.0201 |
| 9 | 131.6±1.3565 | 159 | 0.8277±0.0085 | 0.8093±0.0075 | 0.5429±0.0381 | 0.8710±0.0054 | 0.6759±0.0170 |
| 10 | 109.6±1.3565 | 133 | 0.8241±0.0102 | 0.7903±0.0270 | 0.4500±0.0829 | 0.8752±0.0087 | 0.6381±0.0297 |
| 11 | 94.0±2.7568 | 117 | 0.8034±0.0236 | 0.6901±0.0506 | 0.4000±0.0943 | 0.8627±0.0164 | 0.6134±0.0445 |
| 12 | 73.0±1.8974 | 99 | 0.7374±0.0192 | 0.6739±0.0136 | 0.2923±0.0576 | 0.8047±0.0186 | 0.5340±0.0257 |
| 13 | 71.6±0.4899 | 91 | 0.7868±0.0054 | 0.7199±0.0354 | 0.5077±0.0377 | 0.8333±0.0081 | 0.6373±0.0106 |
| 14 | 59.2±2.0396 | 75 | 0.7893±0.0272 | 0.7011±0.0429 | 0.4167±0.1394 | 0.8603±0.0156 | 0.6275±0.0587 |
| 15 | 48.0±1.0954 | 67 | 0.7164±0.0163 | 0.5558±0.0216 | 0.2182±0.0445 | 0.8143±0.0182 | 0.5143±0.0214 |
| 16 | 41.6±1.0198 | 59 | 0.7051±0.0173 | 0.4461±0.0469 | 0.1200±0.0400 | 0.8245±0.0208 | 0.4716±0.0200 |
| 17 | 39.0±1.2649 | 54 | 0.7222±0.0234 | 0.5348±0.0461 | 0.2000±0.0444 | 0.8267±0.0259 | 0.5129±0.0272 |
| 18 | 32.4±1.9596 | 45 | 0.7200±0.0435 | 0.7108±0.0392 | 0.2250±0.0500 | 0.8270±0.0530 | 0.5264±0.0391 |
| 19 | 26.6±1.7436 | 38 | 0.7000±0.0459 | 0.5000±0.0401 | 0.1000±0.0816 | 0.8125±0.0395 | 0.4599±0.0556 |
| 20 | 26.6±1.8547 | 35 | 0.7600±0.0530 | 0.5966±0.0431 | 0.2000±0.0667 | 0.8759±0.0560 | 0.5429±0.0595 |

Table 94: H12O SVC baseline results of CV using base features, taking best model of grid using F1. Predictions from outcome.

| Day | Corrects | Totals | Accuracy | AUC | Sensitivity | Specificity | F1 |
|---|---|---|---|---|---|---|---|
| 1 | 327.6±2.0591 | 374 | 0.8759±0.0055 | 0.8567±0.0060 | 0.4040±0.0150 | 0.9488±0.0050 | 0.6977±0.0108 |
| 2 | 332.4±1.7436 | 374 | 0.8888±0.0047 | 0.8729±0.0070 | 0.5960±0.0080 | 0.9340±0.0057 | 0.7624±0.0064 |
| 3 | 337.4±1.3565 | 374 | 0.9021±0.0036 | 0.9011±0.0065 | 0.5800±0.0179 | 0.9519±0.0031 | 0.7785±0.0083 |
| 4 | 330.0±1.6733 | 364 | 0.9066±0.0046 | 0.9187±0.0012 | 0.6255±0.0289 | 0.9483±0.0032 | 0.7899±0.0116 |
| 5 | 305.0±2.0976 | 332 | 0.9187±0.0063 | 0.9415±0.0057 | 0.6718±0.0377 | 0.9515±0.0050 | 0.8068±0.0154 |
| 6 | 272.4±1.2000 | 291 | 0.9361±0.0041 | 0.9465±0.0048 | 0.7412±0.0288 | 0.9619±0.0045 | 0.8470±0.0098 |
| 7 | 231.4±0.8000 | 250 | 0.9256±0.0032 | 0.9528±0.0057 | 0.7448±0.0169 | 0.9493±0.0053 | 0.8283±0.0046 |
| 8 | 183.6±1.3565 | 201 | 0.9134±0.0067 | 0.9379±0.0055 | 0.6833±0.0425 | 0.9446±0.0075 | 0.8018±0.0150 |
| 9 | 143.0±1.0954 | 159 | 0.8994±0.0069 | 0.9102±0.0070 | 0.6857±0.0381 | 0.9319±0.0118 | 0.7921±0.0089 |
| 10 | 115.0±1.2649 | 133 | 0.8647±0.0095 | 0.8838±0.0075 | 0.5875±0.0306 | 0.9026±0.0103 | 0.7163±0.0152 |
| 11 | 104.0±1.6733 | 117 | 0.8889±0.0143 | 0.9084±0.0061 | 0.6000±0.0843 | 0.9314±0.0062 | 0.7574±0.0369 |
| 12 | 89.6±0.8000 | 99 | 0.9051±0.0081 | 0.9150±0.0079 | 0.7231±0.0377 | 0.9326±0.0087 | 0.8056±0.0148 |
| 13 | 80.6±1.8547 | 91 | 0.8857±0.0204 | 0.9316±0.0123 | 0.7231±0.0615 | 0.9128±0.0274 | 0.7886±0.0268 |
| 14 | 68.8±1.3266 | 75 | 0.9173±0.0177 | 0.9741±0.0081 | 0.9000±0.0333 | 0.9206±0.0224 | 0.8639±0.0245 |
| 15 | 61.0±0.6325 | 67 | 0.9104±0.0094 | 0.9831±0.0081 | 0.9818±0.0364 | 0.8964±0.0134 | 0.8632±0.0125 |
| 16 | 53.4±0.8000 | 59 | 0.9051±0.0136 | 0.9857±0.0095 | 1.0000±0.0000 | 0.8857±0.0163 | 0.8607±0.0165 |
| 17 | 50.6±1.3565 | 54 | 0.9370±0.0251 | 0.9837±0.0037 | 0.9333±0.0544 | 0.9378±0.0259 | 0.8975±0.0390 |
| 18 | 41.8±0.7483 | 45 | 0.9289±0.0166 | 0.9872±0.0039 | 0.9000±0.0500 | 0.9351±0.0216 | 0.8875±0.0241 |
| 19 | 35.0±0.6325 | 38 | 0.9211±0.0166 | 0.9885±0.0061 | 0.9667±0.0667 | 0.9125±0.0125 | 0.8727±0.0279 |
| 20 | 33.6±0.4899 | 35 | 0.9600±0.0140 | 1.0000±0.0000 | 1.0000±0.0000 | 0.9517±0.0169 | 0.9359±0.0206 |

Table 95: H12O SVC with imputation, results of CV, taking best model of grid using accuracy. Predictions from admission.

| Day | Corrects | Totals | Accuracy | AUC | Sensitivity | Specificity | F1 |
|---|---|---|---|---|---|---|---|
| 1 | 368.0±1.0954 | 374 | 0.9840±0.0029 | 0.9940±0.0025 | 0.9400±0.0000 | 0.9907±0.0034 | 0.9654±0.0060 |
| 2 | 366.8±0.9798 | 374 | 0.9807±0.0026 | 0.9940±0.0017 | 0.9320±0.0098 | 0.9883±0.0023 | 0.9586±0.0055 |
| 3 | 360.8±0.7483 | 374 | 0.9647±0.0020 | 0.9799±0.0036 | 0.8720±0.0204 | 0.9790±0.0012 | 0.9240±0.0051 |
| 4 | 344.8±1.1662 | 364 | 0.9473±0.0032 | 0.9748±0.0022 | 0.8213±0.0217 | 0.9659±0.0024 | 0.8852±0.0075 |
| 5 | 314.0±1.8974 | 332 | 0.9458±0.0057 | 0.9643±0.0021 | 0.7385±0.0192 | 0.9734±0.0040 | 0.8657±0.0135 |
| 6 | 269.4±1.8547 | 291 | 0.9258±0.0064 | 0.9610±0.0022 | 0.7353±0.0263 | 0.9510±0.0063 | 0.8281±0.0132 |
| 7 | 230.6±1.4967 | 250 | 0.9224±0.0060 | 0.9340±0.0041 | 0.7103±0.0352 | 0.9502±0.0029 | 0.8177±0.0153 |
| 8 | 181.2±1.4697 | 201 | 0.9015±0.0073 | 0.8794±0.0175 | 0.5750±0.0486 | 0.9458±0.0045 | 0.7629±0.0206 |
| 9 | 135.8±1.1662 | 159 | 0.8541±0.0073 | 0.8214±0.0179 | 0.4571±0.0883 | 0.9145±0.0096 | 0.6824±0.0305 |
| 10 | 116.2±1.1662 | 133 | 0.8737±0.0088 | 0.8528±0.0102 | 0.4500±0.0612 | 0.9316±0.0054 | 0.6944±0.0262 |
| 11 | 96.4±2.4166 | 117 | 0.8239±0.0207 | 0.7311±0.0207 | 0.1733±0.0680 | 0.9196±0.0144 | 0.5518±0.0461 |
| 12 | 77.2±1.3266 | 99 | 0.7798±0.0134 | 0.7456±0.0146 | 0.2615±0.0615 | 0.8581±0.0186 | 0.5536±0.0205 |
| 13 | 74.6±1.7436 | 91 | 0.8198±0.0192 | 0.7400±0.0104 | 0.4308±0.0923 | 0.8846±0.0162 | 0.6487±0.0401 |
| 14 | 61.6±1.6248 | 75 | 0.8213±0.0217 | 0.7452±0.0524 | 0.2833±0.0667 | 0.9238±0.0156 | 0.6167±0.0454 |
| 15 | 51.6±1.4967 | 67 | 0.7701±0.0223 | 0.6110±0.0390 | 0.2364±0.0727 | 0.8750±0.0160 | 0.5579±0.0428 |
| 16 | 44.0±1.4142 | 59 | 0.7458±0.0240 | 0.5224±0.0281 | 0.0600±0.0490 | 0.8857±0.0208 | 0.4639±0.0374 |
| 17 | 41.8±1.1662 | 54 | 0.7741±0.0216 | 0.6119±0.0194 | 0.2222±0.0703 | 0.8844±0.0259 | 0.5555±0.0371 |
| 18 | 35.0±1.0954 | 45 | 0.7778±0.0243 | 0.6959±0.0386 | 0.2500±0.0000 | 0.8919±0.0296 | 0.5778±0.0188 |
| 19 | 27.8±1.1662 | 38 | 0.7316±0.0307 | 0.5760±0.0299 | 0.1667±0.0000 | 0.8375±0.0364 | 0.5026±0.0184 |
| 20 | 27.4±0.8000 | 35 | 0.7829±0.0229 | 0.6655±0.0516 | 0.1667±0.0000 | 0.9103±0.0276 | 0.5418±0.0157 |

Table 96: H12O SVC with imputation, results of CV, taking best model of grid using accuracy. Predictions from outcome.

| Day | Corrects | Totals | Accuracy | AUC | Sensitivity | Specificity | F1 |
|---|---|---|---|---|---|---|---|
| 1 | 327.6±2.0591 | 374 | 0.8759±0.0055 | 0.8567±0.0060 | 0.4040±0.0150 | 0.9488±0.0050 | 0.6977±0.0108 |
| 2 | 332.4±1.7436 | 374 | 0.8888±0.0047 | 0.8729±0.0070 | 0.5960±0.0080 | 0.9340±0.0057 | 0.7624±0.0064 |
| 3 | 337.4±1.3565 | 374 | 0.9021±0.0036 | 0.9011±0.0065 | 0.5800±0.0179 | 0.9519±0.0031 | 0.7785±0.0083 |
| 4 | 330.0±1.6733 | 364 | 0.9066±0.0046 | 0.9187±0.0012 | 0.6255±0.0289 | 0.9483±0.0032 | 0.7899±0.0116 |
| 5 | 305.0±2.0976 | 332 | 0.9187±0.0063 | 0.9415±0.0057 | 0.6718±0.0377 | 0.9515±0.0050 | 0.8068±0.0154 |
| 6 | 272.4±1.2000 | 291 | 0.9361±0.0041 | 0.9465±0.0048 | 0.7412±0.0288 | 0.9619±0.0045 | 0.8470±0.0098 |
| 7 | 231.4±0.8000 | 250 | 0.9256±0.0032 | 0.9528±0.0057 | 0.7448±0.0169 | 0.9493±0.0053 | 0.8283±0.0046 |
| 8 | 183.6±1.3565 | 201 | 0.9134±0.0067 | 0.9379±0.0055 | 0.6833±0.0425 | 0.9446±0.0075 | 0.8018±0.0150 |
| 9 | 143.0±1.0954 | 159 | 0.8994±0.0069 | 0.9102±0.0070 | 0.6857±0.0381 | 0.9319±0.0118 | 0.7921±0.0089 |
| 10 | 115.0±1.2649 | 133 | 0.8647±0.0095 | 0.8838±0.0075 | 0.5875±0.0306 | 0.9026±0.0103 | 0.7163±0.0152 |
| 11 | 104.0±1.6733 | 117 | 0.8889±0.0143 | 0.9084±0.0061 | 0.6000±0.0843 | 0.9314±0.0062 | 0.7574±0.0369 |
| 12 | 89.6±0.8000 | 99 | 0.9051±0.0081 | 0.9150±0.0079 | 0.7231±0.0377 | 0.9326±0.0087 | 0.8056±0.0148 |
| 13 | 80.6±1.8547 | 91 | 0.8857±0.0204 | 0.9316±0.0123 | 0.7231±0.0615 | 0.9128±0.0274 | 0.7886±0.0268 |
| 14 | 68.8±1.3266 | 75 | 0.9173±0.0177 | 0.9741±0.0081 | 0.9000±0.0333 | 0.9206±0.0224 | 0.8639±0.0245 |
| 15 | 61.0±0.6325 | 67 | 0.9104±0.0094 | 0.9831±0.0081 | 0.9818±0.0364 | 0.8964±0.0134 | 0.8632±0.0125 |
| 16 | 53.4±0.8000 | 59 | 0.9051±0.0136 | 0.9857±0.0095 | 1.0000±0.0000 | 0.8857±0.0163 | 0.8607±0.0165 |
| 17 | 50.6±1.3565 | 54 | 0.9370±0.0251 | 0.9837±0.0037 | 0.9333±0.0544 | 0.9378±0.0259 | 0.8975±0.0390 |
| 18 | 41.8±0.7483 | 45 | 0.9289±0.0166 | 0.9872±0.0039 | 0.9000±0.0500 | 0.9351±0.0216 | 0.8875±0.0241 |
| 19 | 35.0±0.6325 | 38 | 0.9211±0.0166 | 0.9885±0.0061 | 0.9667±0.0667 | 0.9125±0.0125 | 0.8727±0.0279 |
| 20 | 33.6±0.4899 | 35 | 0.9600±0.0140 | 1.0000±0.0000 | 1.0000±0.0000 | 0.9517±0.0169 | 0.9359±0.0206 |

Table 97: H12O SVC with imputation, results of CV, taking best model of grid using F1. Predictions from admission.

| Day | Corrects | Totals | Accuracy | AUC | Sensitivity | Specificity | F1 |
|---|---|---|---|---|---|---|---|
| 1 | 368.0±1.0954 | 374 | 0.9840±0.0029 | 0.9940±0.0025 | 0.9400±0.0000 | 0.9907±0.0034 | 0.9654±0.0060 |
| 2 | 366.8±0.9798 | 374 | 0.9807±0.0026 | 0.9940±0.0017 | 0.9320±0.0098 | 0.9883±0.0023 | 0.9586±0.0055 |
| 3 | 360.8±0.7483 | 374 | 0.9647±0.0020 | 0.9799±0.0036 | 0.8720±0.0204 | 0.9790±0.0012 | 0.9240±0.0051 |
| 4 | 344.8±1.1662 | 364 | 0.9473±0.0032 | 0.9748±0.0022 | 0.8213±0.0217 | 0.9659±0.0024 | 0.8852±0.0075 |
| 5 | 314.0±1.8974 | 332 | 0.9458±0.0057 | 0.9643±0.0021 | 0.7385±0.0192 | 0.9734±0.0040 | 0.8657±0.0135 |
| 6 | 269.4±1.8547 | 291 | 0.9258±0.0064 | 0.9610±0.0022 | 0.7353±0.0263 | 0.9510±0.0063 | 0.8281±0.0132 |
| 7 | 230.6±1.4967 | 250 | 0.9224±0.0060 | 0.9340±0.0041 | 0.7103±0.0352 | 0.9502±0.0029 | 0.8177±0.0153 |
| 8 | 181.2±1.4697 | 201 | 0.9015±0.0073 | 0.8794±0.0175 | 0.5750±0.0486 | 0.9458±0.0045 | 0.7629±0.0206 |
| 9 | 135.8±1.1662 | 159 | 0.8541±0.0073 | 0.8214±0.0179 | 0.4571±0.0883 | 0.9145±0.0096 | 0.6824±0.0305 |
| 10 | 116.2±1.1662 | 133 | 0.8737±0.0088 | 0.8528±0.0102 | 0.4500±0.0612 | 0.9316±0.0054 | 0.6944±0.0262 |
| 11 | 96.4±2.4166 | 117 | 0.8239±0.0207 | 0.7311±0.0207 | 0.1733±0.0680 | 0.9196±0.0144 | 0.5518±0.0461 |
| 12 | 77.2±1.3266 | 99 | 0.7798±0.0134 | 0.7456±0.0146 | 0.2615±0.0615 | 0.8581±0.0186 | 0.5536±0.0205 |
| 13 | 74.6±1.7436 | 91 | 0.8198±0.0192 | 0.7400±0.0104 | 0.4308±0.0923 | 0.8846±0.0162 | 0.6487±0.0401 |
| 14 | 61.6±1.6248 | 75 | 0.8213±0.0217 | 0.7452±0.0524 | 0.2833±0.0667 | 0.9238±0.0156 | 0.6167±0.0454 |
| 15 | 51.6±1.4967 | 67 | 0.7701±0.0223 | 0.6110±0.0390 | 0.2364±0.0727 | 0.8750±0.0160 | 0.5579±0.0428 |
| 16 | 44.0±1.4142 | 59 | 0.7458±0.0240 | 0.5224±0.0281 | 0.0600±0.0490 | 0.8857±0.0208 | 0.4639±0.0374 |
| 17 | 41.8±1.1662 | 54 | 0.7741±0.0216 | 0.6119±0.0194 | 0.2222±0.0703 | 0.8844±0.0259 | 0.5555±0.0371 |
| 18 | 35.0±1.0954 | 45 | 0.7778±0.0243 | 0.6959±0.0386 | 0.2500±0.0000 | 0.8919±0.0296 | 0.5778±0.0188 |
| 19 | 27.8±1.1662 | 38 | 0.7316±0.0307 | 0.5760±0.0299 | 0.1667±0.0000 | 0.8375±0.0364 | 0.5026±0.0184 |
| 20 | 27.4±0.8000 | 35 | 0.7829±0.0229 | 0.6655±0.0516 | 0.1667±0.0000 | 0.9103±0.0276 | 0.5418±0.0157 |

Table 98: H12O SVC with imputation, results of CV, taking best model of grid using F1. Predictions from outcome.

| Day | Corrects | Totals | Accuracy | AUC | Sensitivity | Specificity | F1 |
|---|---|---|---|---|---|---|---|
| 1 | 331.4±1.3565 | 374 | 0.8861±0.0036 | 0.8589±0.0040 | 0.2960±0.0150 | 0.9772±0.0042 | 0.6734±0.0087 |
| 2 | 331.8±2.8566 | 374 | 0.8872±0.0076 | 0.8899±0.0042 | 0.3600±0.0310 | 0.9685±0.0045 | 0.6986±0.0212 |
| 3 | 336.2±1.1662 | 374 | 0.8989±0.0031 | 0.9169±0.0026 | 0.4280±0.0240 | 0.9716±0.0012 | 0.7370±0.0117 |
| 4 | 332.0±1.0954 | 364 | 0.9121±0.0030 | 0.9379±0.0027 | 0.5106±0.0000 | 0.9716±0.0035 | 0.7754±0.0050 |
| 5 | 310.4±1.3565 | 332 | 0.9349±0.0041 | 0.9683±0.0016 | 0.6051±0.0308 | 0.9788±0.0014 | 0.8247±0.0132 |
| 6 | 273.0±0.6325 | 291 | 0.9381±0.0022 | 0.9532±0.0045 | 0.6294±0.0235 | 0.9790±0.0031 | 0.8346±0.0069 |
| 7 | 233.6±1.8547 | 250 | 0.9344±0.0074 | 0.9469±0.0059 | 0.6552±0.0378 | 0.9710±0.0046 | 0.8308±0.0193 |
| 8 | 189.2±1.3266 | 201 | 0.9413±0.0066 | 0.9210±0.0065 | 0.6750±0.0312 | 0.9774±0.0051 | 0.8500±0.0166 |
| 9 | 146.8±0.7483 | 159 | 0.9233±0.0047 | 0.9104±0.0051 | 0.6000±0.0233 | 0.9725±0.0054 | 0.8151±0.0104 |
| 10 | 122.6±0.4899 | 133 | 0.9218±0.0037 | 0.9156±0.0058 | 0.5750±0.0250 | 0.9692±0.0042 | 0.7975±0.0094 |
| 11 | 109.0±0.0000 | 117 | 0.9316±0.0000 | 0.9378±0.0026 | 0.6800±0.0267 | 0.9686±0.0039 | 0.8396±0.0037 |
| 12 | 93.0±1.0954 | 99 | 0.9394±0.0111 | 0.9435±0.0021 | 0.8462±0.0000 | 0.9535±0.0127 | 0.8758±0.0182 |
| 13 | 86.2±0.7483 | 91 | 0.9473±0.0082 | 0.9535±0.0051 | 0.7692±0.0000 | 0.9769±0.0096 | 0.8883±0.0145 |
| 14 | 71.4±0.4899 | 75 | 0.9520±0.0065 | 0.9817±0.0029 | 0.7333±0.0333 | 0.9937±0.0078 | 0.9011±0.0133 |
| 15 | 64.6±0.4899 | 67 | 0.9642±0.0073 | 0.9929±0.0022 | 0.8182±0.0000 | 0.9929±0.0087 | 0.9309±0.0127 |
| 16 | 56.2±1.1662 | 59 | 0.9525±0.0198 | 0.9837±0.0092 | 0.8000±0.0632 | 0.9837±0.0153 | 0.9116±0.0362 |
| 17 | 50.2±0.4000 | 54 | 0.9296±0.0074 | 0.9812±0.0051 | 0.8000±0.0444 | 0.9556±0.0000 | 0.8742±0.0150 |
| 18 | 43.8±0.7483 | 45 | 0.9733±0.0166 | 1.0000±0.0000 | 1.0000±0.0000 | 0.9676±0.0202 | 0.9577±0.0258 |
| 19 | 36.8±0.4000 | 38 | 0.9684±0.0105 | 0.9917±0.0026 | 1.0000±0.0000 | 0.9625±0.0125 | 0.9454±0.0165 |
| 20 | 34.0±0.0000 | 35 | 0.9714±0.0000 | 0.9954±0.0023 | 1.0000±0.0000 | 0.9655±0.0000 | 0.9528±0.0000 |

Table 99: H12O SVC with imputation, with missing flags, results of CV, taking best model of grid using accuracy. Predictions from admission.

| Day | Corrects | Totals | Accuracy | AUC | Sensitivity | Specificity | F1 |
|---|---|---|---|---|---|---|---|
| 1 | 367.0±0.6325 | 374 | 0.9813±0.0017 | 0.9955±0.0004 | 0.9120±0.0098 | 0.9920±0.0015 | 0.9590±0.0037 |
| 2 | 365.2±0.7483 | 374 | 0.9765±0.0020 | 0.9961±0.0004 | 0.9000±0.0126 | 0.9883±0.0012 | 0.9487±0.0045 |
| 3 | 366.4±1.0198 | 374 | 0.9797±0.0027 | 0.9949±0.0006 | 0.9280±0.0204 | 0.9877±0.0000 | 0.9562±0.0062 |
| 4 | 346.4±1.2000 | 364 | 0.9516±0.0033 | 0.9783±0.0020 | 0.7702±0.0248 | 0.9785±0.0013 | 0.8883±0.0087 |
| 5 | 310.8±1.1662 | 332 | 0.9361±0.0035 | 0.9637±0.0024 | 0.6615±0.0192 | 0.9727±0.0031 | 0.8365±0.0089 |
| 6 | 273.6±2.3324 | 291 | 0.9402±0.0080 | 0.9547±0.0040 | 0.6353±0.0353 | 0.9805±0.0049 | 0.8397±0.0212 |
| 7 | 228.8±1.3266 | 250 | 0.9152±0.0053 | 0.9332±0.0041 | 0.5517±0.0308 | 0.9629±0.0044 | 0.7769±0.0144 |
| 8 | 183.8±0.9798 | 201 | 0.9144±0.0049 | 0.8912±0.0085 | 0.4167±0.0000 | 0.9819±0.0055 | 0.7454±0.0083 |
| 9 | 143.6±0.4899 | 159 | 0.9031±0.0031 | 0.8349±0.0144 | 0.3333±0.0000 | 0.9899±0.0035 | 0.7115±0.0048 |
| 10 | 120.2±0.7483 | 133 | 0.9038±0.0056 | 0.8397±0.0095 | 0.3500±0.0306 | 0.9795±0.0087 | 0.7066±0.0117 |
| 11 | 102.0±0.8944 | 117 | 0.8718±0.0076 | 0.7788±0.0127 | 0.1467±0.0267 | 0.9784±0.0114 | 0.5778±0.0120 |
| 12 | 85.6±0.4899 | 99 | 0.8646±0.0049 | 0.7934±0.0146 | 0.0615±0.0576 | 0.9860±0.0087 | 0.5127±0.0449 |
| 13 | 77.6±0.4899 | 91 | 0.8527±0.0054 | 0.7710±0.0076 | 0.0462±0.0377 | 0.9872±0.0081 | 0.4991±0.0321 |
| 14 | 63.8±0.7483 | 75 | 0.8507±0.0100 | 0.8153±0.0109 | 0.0667±0.0624 | 1.0000±0.0000 | 0.5185±0.0566 |
| 15 | 57.0±0.0000 | 67 | 0.8507±0.0000 | 0.7130±0.0218 | 0.0909±0.0000 | 1.0000±0.0000 | 0.5423±0.0000 |
| 16 | 49.0±0.0000 | 59 | 0.8305±0.0000 | 0.6600±0.0140 | 0.0000±0.0000 | 1.0000±0.0000 | 0.4537±0.0000 |
| 17 | 45.0±0.0000 | 54 | 0.8333±0.0000 | 0.6770±0.0088 | 0.0000±0.0000 | 1.0000±0.0000 | 0.4545±0.0000 |
| 18 | 37.2±0.4000 | 45 | 0.8267±0.0089 | 0.7527±0.0174 | 0.0250±0.0500 | 1.0000±0.0000 | 0.4746±0.0467 |
| 19 | 32.0±0.0000 | 38 | 0.8421±0.0000 | 0.7115±0.0488 | 0.0000±0.0000 | 1.0000±0.0000 | 0.4571±0.0000 |
| 20 | 29.0±0.0000 | 35 | 0.8286±0.0000 | 0.7218±0.0414 | 0.0000±0.0000 | 1.0000±0.0000 | 0.4531±0.0000 |

Table 100: H12O SVC with imputation, with missing flags, results of CV, taking best model of grid using accuracy. Predictions from outcome.

| Day | Corrects | Totals | Accuracy | AUC | Sensitivity | Specificity | F1 |
|---|---|---|---|---|---|---|---|
| 1 | 325.0±3.8471 | 374 | 0.8690±0.0103 | 0.8673±0.0096 | 0.4880±0.0098 | 0.9278±0.0120 | 0.7121±0.0130 |
| 2 | 328.2±1.9391 | 374 | 0.8775±0.0052 | 0.8777±0.0057 | 0.6360±0.0233 | 0.9148±0.0064 | 0.7548±0.0086 |
| 3 | 334.0±1.5492 | 374 | 0.8930±0.0041 | 0.8734±0.0087 | 0.6360±0.0265 | 0.9327±0.0030 | 0.7758±0.0100 |
| 4 | 331.2±2.4819 | 364 | 0.9099±0.0068 | 0.9087±0.0111 | 0.6936±0.0255 | 0.9420±0.0047 | 0.8066±0.0142 |
| 5 | 304.2±2.4819 | 332 | 0.9163±0.0075 | 0.9433±0.0102 | 0.7282±0.0308 | 0.9413±0.0050 | 0.8117±0.0164 |
| 6 | 271.0±0.8944 | 291 | 0.9313±0.0031 | 0.9466±0.0086 | 0.7647±0.0186 | 0.9533±0.0025 | 0.8415±0.0073 |
| 7 | 227.8±2.7129 | 250 | 0.9112±0.0109 | 0.9210±0.0082 | 0.7379±0.0352 | 0.9339±0.0079 | 0.8039±0.0221 |
| 8 | 180.8±2.3152 | 201 | 0.8995±0.0115 | 0.9284±0.0097 | 0.7750±0.0425 | 0.9164±0.0121 | 0.7950±0.0203 |
| 9 | 141.2±1.9391 | 159 | 0.8881±0.0122 | 0.9018±0.0065 | 0.6667±0.0522 | 0.9217±0.0116 | 0.7730±0.0225 |
| 10 | 112.8±0.4000 | 133 | 0.8481±0.0030 | 0.8715±0.0038 | 0.6375±0.0468 | 0.8769±0.0068 | 0.7061±0.0100 |
| 11 | 101.6±1.7436 | 117 | 0.8684±0.0149 | 0.8970±0.0143 | 0.6533±0.0653 | 0.9000±0.0200 | 0.7413±0.0233 |
| 12 | 88.4±1.4967 | 99 | 0.8929±0.0151 | 0.8984±0.0258 | 0.6923±0.0000 | 0.9233±0.0174 | 0.7843±0.0219 |
| 13 | 79.6±1.3565 | 91 | 0.8747±0.0149 | 0.9211±0.0209 | 0.7846±0.0576 | 0.8897±0.0103 | 0.7827±0.0257 |
| 14 | 67.6±1.0198 | 75 | 0.9013±0.0136 | 0.9709±0.0090 | 0.9000±0.0333 | 0.9016±0.0119 | 0.8420±0.0204 |
| 15 | 62.0±1.0954 | 67 | 0.9254±0.0163 | 0.9864±0.0044 | 0.9818±0.0364 | 0.9143±0.0175 | 0.8831±0.0241 |
| 16 | 53.4±1.0198 | 59 | 0.9051±0.0173 | 0.9820±0.0080 | 1.0000±0.0000 | 0.8857±0.0208 | 0.8609±0.0216 |
| 17 | 50.2±1.3266 | 54 | 0.9296±0.0246 | 0.9783±0.0048 | 0.9111±0.1089 | 0.9333±0.0199 | 0.8837±0.0416 |
| 18 | 43.4±0.8000 | 45 | 0.9644±0.0178 | 0.9980±0.0017 | 1.0000±0.0000 | 0.9568±0.0216 | 0.9443±0.0257 |
| 19 | 36.2±0.7483 | 38 | 0.9526±0.0197 | 0.9958±0.0021 | 1.0000±0.0000 | 0.9437±0.0234 | 0.9215±0.0295 |
| 20 | 33.6±0.4899 | 35 | 0.9600±0.0140 | 1.0000±0.0000 | 1.0000±0.0000 | 0.9517±0.0169 | 0.9359±0.0206 |

Table 101: H12O SVC with imputation, with missing flags, results of CV, taking best model of grid using F1. Predictions from admission.

| Day | Corrects | Totals | Accuracy | AUC | Sensitivity | Specificity | F1 |
|---|---|---|---|---|---|---|---|
| 1 | 368.4±1.8547 | 374 | 0.9850±0.0050 | 0.9940±0.0028 | 0.9360±0.0080 | 0.9926±0.0046 | 0.9676±0.0104 |
| 2 | 364.6±1.6248 | 374 | 0.9749±0.0043 | 0.9943±0.0012 | 0.9120±0.0204 | 0.9846±0.0044 | 0.9461±0.0090 |
| 3 | 363.2±1.1662 | 374 | 0.9711±0.0031 | 0.9738±0.0031 | 0.9080±0.0204 | 0.9809±0.0023 | 0.9385±0.0069 |
| 4 | 347.6±1.4967 | 364 | 0.9549±0.0041 | 0.9724±0.0029 | 0.8851±0.0289 | 0.9653±0.0020 | 0.9045±0.0097 |
| 5 | 305.8±1.6000 | 332 | 0.9211±0.0048 | 0.9600±0.0065 | 0.7077±0.0126 | 0.9495±0.0050 | 0.8167±0.0090 |
| 6 | 267.6±1.8547 | 291 | 0.9196±0.0064 | 0.9606±0.0038 | 0.7412±0.0390 | 0.9432±0.0068 | 0.8184±0.0137 |
| 7 | 223.8±1.3266 | 250 | 0.8952±0.0053 | 0.9182±0.0061 | 0.6828±0.0402 | 0.9231±0.0050 | 0.7706±0.0133 |
| 8 | 176.6±2.3324 | 201 | 0.8786±0.0116 | 0.8809±0.0074 | 0.5333±0.0612 | 0.9254±0.0115 | 0.7211±0.0256 |
| 9 | 132.0±2.4495 | 159 | 0.8302±0.0154 | 0.8167±0.0260 | 0.6000±0.0233 | 0.8652±0.0175 | 0.6910±0.0175 |
| 10 | 112.6±2.1541 | 133 | 0.8466±0.0162 | 0.8330±0.0240 | 0.4125±0.0306 | 0.9060±0.0153 | 0.6532±0.0254 |
| 11 | 92.8±0.7483 | 117 | 0.7932±0.0064 | 0.6987±0.0411 | 0.2933±0.0327 | 0.8667±0.0078 | 0.5729±0.0127 |
| 12 | 77.0±1.0954 | 99 | 0.7778±0.0111 | 0.6834±0.0274 | 0.3538±0.0377 | 0.8419±0.0119 | 0.5814±0.0166 |
| 13 | 71.8±2.5612 | 91 | 0.7890±0.0281 | 0.7026±0.0193 | 0.5231±0.1231 | 0.8333±0.0281 | 0.6414±0.0483 |
| 14 | 58.8±3.1875 | 75 | 0.7840±0.0425 | 0.6947±0.0259 | 0.4000±0.0972 | 0.8571±0.0348 | 0.6225±0.0647 |
| 15 | 48.2±2.3152 | 67 | 0.7194±0.0346 | 0.6036±0.0535 | 0.2364±0.0927 | 0.8143±0.0331 | 0.5226±0.0493 |
| 16 | 42.4±1.4967 | 59 | 0.7186±0.0254 | 0.5045±0.0344 | 0.1000±0.0000 | 0.8449±0.0305 | 0.4705±0.0131 |
| 17 | 39.2±1.7205 | 54 | 0.7259±0.0319 | 0.5610±0.0400 | 0.2889±0.0544 | 0.8133±0.0435 | 0.5453±0.0224 |
| 18 | 33.4±2.1541 | 45 | 0.7422±0.0479 | 0.7209±0.0507 | 0.3250±0.1000 | 0.8324±0.0524 | 0.5753±0.0561 |
| 19 | 26.0±2.6077 | 38 | 0.6842±0.0686 | 0.4646±0.0628 | 0.1667±0.1054 | 0.7812±0.0625 | 0.4790±0.0759 |
| 20 | 28.4±1.3565 | 35 | 0.8114±0.0388 | 0.7540±0.0533 | 0.4000±0.0816 | 0.8966±0.0436 | 0.6549±0.0497 |

Table 102: H12O SVC with imputation, with missing flags, results of CV, taking best model of grid using F1. Predictions from outcome.

| Day | Corrects | Totals | Accuracy | AUC | Sensitivity | Specificity | F1 |
|---|---|---|---|---|---|---|---|
| 1 | 328.0±0.8944 | 374 | 0.8770±0.0024 | 0.8436±0.0047 | 0.2520±0.0160 | 0.9735±0.0031 | 0.6429±0.0084 |
| 2 | 331.8±0.7483 | 374 | 0.8872±0.0020 | 0.8949±0.0050 | 0.3800±0.0253 | 0.9654±0.0023 | 0.7051±0.0105 |
| 3 | 338.2±1.1662 | 374 | 0.9043±0.0031 | 0.9254±0.0048 | 0.5000±0.0126 | 0.9667±0.0030 | 0.7643±0.0070 |
| 4 | 332.8±0.7483 | 364 | 0.9143±0.0021 | 0.9374±0.0028 | 0.5574±0.0159 | 0.9672±0.0032 | 0.7892±0.0050 |
| 5 | 309.8±2.0396 | 332 | 0.9331±0.0061 | 0.9682±0.0015 | 0.6462±0.0299 | 0.9713±0.0035 | 0.8283±0.0161 |
| 6 | 275.2±0.9798 | 291 | 0.9457±0.0034 | 0.9563±0.0022 | 0.6529±0.0220 | 0.9844±0.0025 | 0.8536±0.0097 |
| 7 | 234.2±1.1662 | 250 | 0.9368±0.0047 | 0.9580±0.0012 | 0.6759±0.0169 | 0.9710±0.0046 | 0.8387±0.0104 |
| 8 | 189.8±1.1662 | 201 | 0.9443±0.0058 | 0.9498±0.0052 | 0.7250±0.0333 | 0.9740±0.0045 | 0.8625±0.0148 |
| 9 | 148.8±1.1662 | 159 | 0.9358±0.0073 | 0.9273±0.0059 | 0.7524±0.0356 | 0.9638±0.0065 | 0.8595±0.0157 |
| 10 | 122.8±0.7483 | 133 | 0.9233±0.0056 | 0.9329±0.0038 | 0.6500±0.0306 | 0.9607±0.0068 | 0.8138±0.0120 |
| 11 | 109.2±0.9798 | 117 | 0.9333±0.0084 | 0.9454±0.0012 | 0.7333±0.0422 | 0.9627±0.0039 | 0.8499±0.0200 |
| 12 | 95.2±0.9798 | 99 | 0.9616±0.0099 | 0.9367±0.0033 | 0.8462±0.0000 | 0.9791±0.0114 | 0.9159±0.0189 |
| 13 | 85.2±0.7483 | 91 | 0.9363±0.0082 | 0.9525±0.0050 | 0.7692±0.0000 | 0.9641±0.0096 | 0.8693±0.0136 |
| 14 | 71.8±0.4000 | 75 | 0.9573±0.0053 | 0.9767±0.0032 | 0.7500±0.0000 | 0.9968±0.0063 | 0.9122±0.0094 |
| 15 | 65.0±0.8944 | 67 | 0.9701±0.0133 | 0.9919±0.0023 | 0.8727±0.0445 | 0.9893±0.0087 | 0.9439±0.0253 |
| 16 | 56.2±0.7483 | 59 | 0.9525±0.0127 | 0.9853±0.0035 | 0.8600±0.0490 | 0.9714±0.0100 | 0.9156±0.0227 |
| 17 | 51.0±0.8944 | 54 | 0.9444±0.0166 | 0.9916±0.0030 | 0.8889±0.0994 | 0.9556±0.0000 | 0.9030±0.0322 |
| 18 | 45.0±0.0000 | 45 | 1.0000±0.0000 | 1.0000±0.0000 | 1.0000±0.0000 | 1.0000±0.0000 | 1.0000±0.0000 |
| 19 | 36.8±0.4000 | 38 | 0.9684±0.0105 | 0.9990±0.0021 | 1.0000±0.0000 | 0.9625±0.0125 | 0.9454±0.0165 |
| 20 | 34.2±0.4000 | 35 | 0.9771±0.0114 | 1.0000±0.0000 | 1.0000±0.0000 | 0.9724±0.0138 | 0.9622±0.0189 |

Table 103: H12O SVC with imputation, with missing flags, applying reference values, results of CV, taking best model of grid using accuracy. Predictions from admission.

| Day | Corrects | Totals | Accuracy | AUC | Sensitivity | Specificity | F1 |
|---|---|---|---|---|---|---|---|
| 1 | 367.4±0.4899 | 374 | 0.9824±0.0013 | 0.9965±0.0003 | 0.9200±0.0000 | 0.9920±0.0015 | 0.9615±0.0027 |
| 2 | 366.6±0.8000 | 374 | 0.9802±0.0021 | 0.9970±0.0001 | 0.9080±0.0098 | 0.9914±0.0012 | 0.9566±0.0047 |
| 3 | 366.0±0.8944 | 374 | 0.9786±0.0024 | 0.9956±0.0005 | 0.9160±0.0150 | 0.9883±0.0012 | 0.9536±0.0054 |
| 4 | 350.2±0.7483 | 364 | 0.9621±0.0021 | 0.9795±0.0014 | 0.8426±0.0104 | 0.9798±0.0015 | 0.9149±0.0046 |
| 5 | 314.4±1.4967 | 332 | 0.9470±0.0045 | 0.9703±0.0013 | 0.7231±0.0299 | 0.9768±0.0014 | 0.8660±0.0126 |
| 6 | 273.0±1.7889 | 291 | 0.9381±0.0061 | 0.9577±0.0033 | 0.6647±0.0399 | 0.9743±0.0053 | 0.8401±0.0162 |
| 7 | 231.6±1.8547 | 250 | 0.9264±0.0074 | 0.9438±0.0055 | 0.6000±0.0276 | 0.9692±0.0078 | 0.8067±0.0166 |
| 8 | 185.0±0.6325 | 201 | 0.9204±0.0031 | 0.8895±0.0107 | 0.4583±0.0264 | 0.9831±0.0000 | 0.7673±0.0127 |
| 9 | 142.0±0.6325 | 159 | 0.8931±0.0040 | 0.8531±0.0078 | 0.3048±0.0233 | 0.9826±0.0035 | 0.6851±0.0136 |
| 10 | 119.0±1.0954 | 133 | 0.8947±0.0082 | 0.8632±0.0077 | 0.3625±0.0612 | 0.9675±0.0034 | 0.6964±0.0333 |
| 11 | 102.6±1.0198 | 117 | 0.8769±0.0087 | 0.8301±0.0174 | 0.1600±0.0327 | 0.9824±0.0096 | 0.5911±0.0234 |
| 12 | 85.8±0.7483 | 99 | 0.8667±0.0076 | 0.8397±0.0086 | 0.1231±0.0923 | 0.9791±0.0114 | 0.5536±0.0642 |
| 13 | 78.6±0.8000 | 91 | 0.8637±0.0088 | 0.7842±0.0225 | 0.1538±0.1088 | 0.9821±0.0131 | 0.5746±0.0742 |
| 14 | 63.6±0.8000 | 75 | 0.8480±0.0107 | 0.8106±0.0076 | 0.0833±0.0913 | 0.9937±0.0078 | 0.5255±0.0710 |
| 15 | 56.0±0.0000 | 67 | 0.8358±0.0000 | 0.7049±0.0141 | 0.0909±0.0000 | 0.9821±0.0000 | 0.5315±0.0000 |
| 16 | 49.0±0.0000 | 59 | 0.8305±0.0000 | 0.6257±0.0181 | 0.0000±0.0000 | 1.0000±0.0000 | 0.4537±0.0000 |
| 17 | 45.0±0.0000 | 54 | 0.8333±0.0000 | 0.6425±0.0222 | 0.0000±0.0000 | 1.0000±0.0000 | 0.4545±0.0000 |
| 18 | 37.0±0.0000 | 45 | 0.8222±0.0000 | 0.7264±0.0192 | 0.0250±0.0500 | 0.9946±0.0108 | 0.4710±0.0395 |
| 19 | 32.0±0.0000 | 38 | 0.8421±0.0000 | 0.6115±0.0437 | 0.0000±0.0000 | 1.0000±0.0000 | 0.4571±0.0000 |
| 20 | 29.0±0.6325 | 35 | 0.8286±0.0181 | 0.6966±0.0305 | 0.0333±0.0667 | 0.9931±0.0138 | 0.4814±0.0610 |

Table 104: H12O SVC with imputation, with missing flags, applying reference values, results of CV, taking best model of grid using accuracy. Predictions from outcome.

| Day | Corrects | Totals | Accuracy | AUC | Sensitivity | Specificity | F1 |
|---|---|---|---|---|---|---|---|
| 1 | 326.6±3.0067 | 374 | 0.8733±0.0080 | 0.8531±0.0091 | 0.4680±0.0204 | 0.9358±0.0063 | 0.7123±0.0156 |
| 2 | 328.6±1.4967 | 374 | 0.8786±0.0040 | 0.8816±0.0078 | 0.5600±0.0179 | 0.9278±0.0042 | 0.7410±0.0078 |
| 3 | 333.2±1.9391 | 374 | 0.8909±0.0052 | 0.9047±0.0053 | 0.6040±0.0388 | 0.9352±0.0044 | 0.7667±0.0134 |
| 4 | 329.2±1.7205 | 364 | 0.9044±0.0047 | 0.9127±0.0062 | 0.6894±0.0170 | 0.9363±0.0037 | 0.7976±0.0094 |
| 5 | 308.0±3.0332 | 332 | 0.9277±0.0091 | 0.9549±0.0062 | 0.7897±0.0497 | 0.9461±0.0045 | 0.8389±0.0213 |
| 6 | 268.4±1.4967 | 291 | 0.9223±0.0051 | 0.9467±0.0056 | 0.7235±0.0235 | 0.9486±0.0052 | 0.8205±0.0106 |
| 7 | 227.4±2.6533 | 250 | 0.9096±0.0106 | 0.9346±0.0052 | 0.7103±0.0352 | 0.9357±0.0078 | 0.7972±0.0222 |
| 8 | 184.8±1.7205 | 201 | 0.9194±0.0086 | 0.9430±0.0072 | 0.7750±0.0204 | 0.9390±0.0110 | 0.8254±0.0130 |
| 9 | 141.8±1.1662 | 159 | 0.8918±0.0073 | 0.9054±0.0106 | 0.6667±0.0301 | 0.9261±0.0054 | 0.7782±0.0148 |
| 10 | 117.6±0.8000 | 133 | 0.8842±0.0060 | 0.8683±0.0080 | 0.6250±0.0000 | 0.9197±0.0068 | 0.7492±0.0080 |
| 11 | 104.2±0.4000 | 117 | 0.8906±0.0034 | 0.8959±0.0131 | 0.6667±0.0000 | 0.9235±0.0039 | 0.7731±0.0048 |
| 12 | 87.6±1.4967 | 99 | 0.8848±0.0151 | 0.9059±0.0208 | 0.7231±0.0377 | 0.9093±0.0154 | 0.7777±0.0232 |
| 13 | 80.2±1.7205 | 91 | 0.8813±0.0189 | 0.9221±0.0126 | 0.7538±0.0576 | 0.9026±0.0174 | 0.7871±0.0309 |
| 14 | 68.0±0.6325 | 75 | 0.9067±0.0084 | 0.9616±0.0063 | 0.8000±0.0408 | 0.9270±0.0078 | 0.8380±0.0152 |
| 15 | 62.0±1.4142 | 67 | 0.9254±0.0211 | 0.9708±0.0062 | 0.8909±0.0364 | 0.9321±0.0237 | 0.8764±0.0311 |
| 16 | 52.2±1.1662 | 59 | 0.8847±0.0198 | 0.9755±0.0043 | 0.9000±0.0000 | 0.8816±0.0238 | 0.8272±0.0241 |
| 17 | 51.4±0.8000 | 54 | 0.9519±0.0148 | 0.9802±0.0027 | 0.9778±0.0444 | 0.9467±0.0178 | 0.9212±0.0221 |
| 18 | 43.4±0.8000 | 45 | 0.9644±0.0178 | 1.0000±0.0000 | 1.0000±0.0000 | 0.9568±0.0216 | 0.9443±0.0257 |
| 19 | 35.8±0.7483 | 38 | 0.9421±0.0197 | 1.0000±0.0000 | 1.0000±0.0000 | 0.9313±0.0234 | 0.9059±0.0291 |
| 20 | 35.0±0.0000 | 35 | 1.0000±0.0000 | 1.0000±0.0000 | 1.0000±0.0000 | 1.0000±0.0000 | 1.0000±0.0000 |

Table 105: H12O SVC with imputation, with missing flags, applying reference values, results of CV, taking best model of grid using F1. Predictions from admission.

| Day | Corrects | Totals | Accuracy | AUC | Sensitivity | Specificity | F1 |
|---|---|---|---|---|---|---|---|
| 1 | 368.6±1.3565 | 374 | 0.9856±0.0036 | 0.9939±0.0022 | 0.9360±0.0150 | 0.9932±0.0036 | 0.9686±0.0077 |
| 2 | 365.6±1.3565 | 374 | 0.9775±0.0036 | 0.9944±0.0012 | 0.9160±0.0233 | 0.9870±0.0030 | 0.9515±0.0080 |
| 3 | 360.8±1.1662 | 374 | 0.9647±0.0031 | 0.9790±0.0045 | 0.8880±0.0160 | 0.9765±0.0031 | 0.9251±0.0064 |
| 4 | 346.6±2.7276 | 364 | 0.9522±0.0075 | 0.9754±0.0025 | 0.9277±0.0104 | 0.9558±0.0082 | 0.9031±0.0135 |
| 5 | 306.6±2.2450 | 332 | 0.9235±0.0068 | 0.9596±0.0077 | 0.7179±0.0162 | 0.9509±0.0060 | 0.8223±0.0135 |
| 6 | 268.4±3.2619 | 291 | 0.9223±0.0112 | 0.9514±0.0059 | 0.7412±0.0471 | 0.9463±0.0067 | 0.8230±0.0253 |
| 7 | 227.0±0.8944 | 250 | 0.9080±0.0036 | 0.9220±0.0128 | 0.6897±0.0218 | 0.9367±0.0064 | 0.7912±0.0040 |
| 8 | 181.0±2.6077 | 201 | 0.9005±0.0130 | 0.8809±0.0115 | 0.5917±0.0312 | 0.9424±0.0131 | 0.7657±0.0234 |
| 9 | 136.0±2.0000 | 159 | 0.8553±0.0126 | 0.8466±0.0128 | 0.5810±0.0190 | 0.8971±0.0141 | 0.7153±0.0163 |
| 10 | 113.8±1.6000 | 133 | 0.8556±0.0120 | 0.8091±0.0249 | 0.5000±0.0685 | 0.9043±0.0126 | 0.6852±0.0252 |
| 11 | 94.6±1.6248 | 117 | 0.8085±0.0139 | 0.6946±0.0324 | 0.3067±0.0327 | 0.8824±0.0139 | 0.5904±0.0200 |
| 12 | 78.0±1.5492 | 99 | 0.7879±0.0156 | 0.7603±0.0168 | 0.4462±0.0897 | 0.8395±0.0200 | 0.6131±0.0286 |
| 13 | 72.2±1.3266 | 91 | 0.7934±0.0146 | 0.7272±0.0277 | 0.4769±0.0576 | 0.8462±0.0081 | 0.6363±0.0270 |
| 14 | 57.8±1.9391 | 75 | 0.7707±0.0259 | 0.6812±0.0255 | 0.4500±0.1130 | 0.8317±0.0162 | 0.6210±0.0505 |
| 15 | 47.4±2.4166 | 67 | 0.7075±0.0361 | 0.5877±0.0262 | 0.3273±0.0927 | 0.7821±0.0484 | 0.5410±0.0357 |
| 16 | 42.2±1.1662 | 59 | 0.7153±0.0198 | 0.5106±0.0513 | 0.2200±0.0980 | 0.8163±0.0183 | 0.5149±0.0433 |
| 17 | 38.4±1.0198 | 54 | 0.7111±0.0189 | 0.4780±0.0404 | 0.1778±0.0544 | 0.8178±0.0166 | 0.4974±0.0302 |
| 18 | 31.6±1.8547 | 45 | 0.7022±0.0412 | 0.6182±0.0649 | 0.4250±0.1000 | 0.7622±0.0524 | 0.5715±0.0390 |
| 19 | 24.8±2.0396 | 38 | 0.6526±0.0537 | 0.4365±0.0603 | 0.2667±0.0816 | 0.7250±0.0500 | 0.4886±0.0540 |
| 20 | 27.2±1.7205 | 35 | 0.7771±0.0492 | 0.6989±0.0745 | 0.3000±0.1247 | 0.8759±0.0468 | 0.5911±0.0723 |

Table 106: H12O SVC with imputation, with missing flags, applying reference values, results of CV, taking best model of grid using F1. Predictions from outcome.

| Day | Corrects | Totals | Accuracy | AUC | Sensitivity | Specificity | F1 |
|---|---|---|---|---|---|---|---|
| 1 | 331.6±1.2000 | 374 | 0.8866±0.0032 | 0.8615±0.0048 | 0.3240±0.0150 | 0.9735±0.0050 | 0.6850±0.0059 |
| 2 | 333.4±1.2000 | 374 | 0.8914±0.0032 | 0.8984±0.0048 | 0.3960±0.0233 | 0.9679±0.0042 | 0.7163±0.0098 |
| 3 | 335.4±1.0198 | 374 | 0.8968±0.0027 | 0.9165±0.0029 | 0.4320±0.0271 | 0.9685±0.0023 | 0.7349±0.0112 |
| 4 | 330.2±0.7483 | 364 | 0.9071±0.0021 | 0.9370±0.0021 | 0.5149±0.0085 | 0.9653±0.0020 | 0.7682±0.0046 |
| 5 | 309.0±0.8944 | 332 | 0.9307±0.0027 | 0.9682±0.0025 | 0.5692±0.0192 | 0.9788±0.0014 | 0.8100±0.0087 |
| 6 | 271.6±0.8000 | 291 | 0.9333±0.0027 | 0.9494±0.0027 | 0.6059±0.0235 | 0.9767±0.0000 | 0.8212±0.0094 |
| 7 | 231.2±0.9798 | 250 | 0.9248±0.0039 | 0.9456±0.0069 | 0.6483±0.0338 | 0.9611±0.0022 | 0.8119±0.0124 |
| 8 | 187.6±0.8000 | 201 | 0.9333±0.0040 | 0.9162±0.0072 | 0.6500±0.0204 | 0.9718±0.0036 | 0.8310±0.0097 |
| 9 | 144.8±0.7483 | 159 | 0.9107±0.0047 | 0.9010±0.0060 | 0.5714±0.0301 | 0.9623±0.0071 | 0.7887±0.0097 |
| 10 | 121.2±1.1662 | 133 | 0.9113±0.0088 | 0.9139±0.0090 | 0.5375±0.0500 | 0.9624±0.0042 | 0.7713±0.0248 |
| 11 | 109.2±0.9798 | 117 | 0.9333±0.0084 | 0.9290±0.0032 | 0.6800±0.0267 | 0.9706±0.0088 | 0.8430±0.0167 |
| 12 | 92.6±0.4899 | 99 | 0.9354±0.0049 | 0.9410±0.0040 | 0.8462±0.0000 | 0.9488±0.0057 | 0.8686±0.0081 |
| 13 | 84.6±1.4967 | 91 | 0.9297±0.0164 | 0.9404±0.0072 | 0.7692±0.0000 | 0.9564±0.0192 | 0.8593±0.0258 |
| 14 | 70.8±0.7483 | 75 | 0.9440±0.0100 | 0.9820±0.0023 | 0.8167±0.0624 | 0.9683±0.0000 | 0.8946±0.0212 |
| 15 | 64.6±0.4899 | 67 | 0.9642±0.0073 | 0.9906±0.0016 | 0.8182±0.0000 | 0.9929±0.0087 | 0.9309±0.0127 |
| 16 | 55.4±0.8000 | 59 | 0.9390±0.0136 | 0.9812±0.0073 | 0.7800±0.0400 | 0.9714±0.0100 | 0.8881±0.0247 |
| 17 | 51.0±0.0000 | 54 | 0.9444±0.0000 | 0.9699±0.0098 | 0.8889±0.0000 | 0.9556±0.0000 | 0.9042±0.0000 |
| 18 | 43.8±0.4000 | 45 | 0.9733±0.0089 | 0.9980±0.0017 | 1.0000±0.0000 | 0.9676±0.0108 | 0.9571±0.0133 |
| 19 | 36.2±0.4000 | 38 | 0.9526±0.0105 | 0.9885±0.0039 | 0.9667±0.0667 | 0.9500±0.0153 | 0.9184±0.0181 |
| 20 | 34.0±0.0000 | 35 | 0.9714±0.0000 | 0.9897±0.0023 | 1.0000±0.0000 | 0.9655±0.0000 | 0.9528±0.0000 |

Table 107: H12O SVC with missing flags, results of CV, taking best model of grid using accuracy. Predictions from admission.

| Day | Corrects | Totals | Accuracy | AUC | Sensitivity | Specificity | F1 |
|---|---|---|---|---|---|---|---|
| 1 | 362.4±0.8000 | 374 | 0.9690±0.0021 | 0.9899±0.0004 | 0.8440±0.0080 | 0.9883±0.0023 | 0.9307±0.0045 |
| 2 | 364.2±0.7483 | 374 | 0.9738±0.0020 | 0.9946±0.0005 | 0.8800±0.0126 | 0.9883±0.0012 | 0.9423±0.0046 |
| 3 | 364.4±1.0198 | 374 | 0.9743±0.0027 | 0.9937±0.0006 | 0.8920±0.0160 | 0.9870±0.0012 | 0.9440±0.0061 |
| 4 | 348.4±0.8000 | 364 | 0.9571±0.0022 | 0.9776±0.0013 | 0.8043±0.0208 | 0.9798±0.0015 | 0.9022±0.0060 |
| 5 | 311.6±0.8000 | 332 | 0.9386±0.0024 | 0.9647±0.0018 | 0.6462±0.0192 | 0.9775±0.0017 | 0.8387±0.0074 |
| 6 | 273.8±0.4000 | 291 | 0.9409±0.0014 | 0.9526±0.0047 | 0.6588±0.0144 | 0.9782±0.0031 | 0.8447±0.0022 |
| 7 | 229.6±1.8547 | 250 | 0.9184±0.0074 | 0.9361±0.0040 | 0.5793±0.0258 | 0.9629±0.0053 | 0.7883±0.0178 |
| 8 | 183.4±1.4967 | 201 | 0.9124±0.0074 | 0.9052±0.0088 | 0.4083±0.0167 | 0.9808±0.0085 | 0.7396±0.0148 |
| 9 | 142.6±1.8547 | 159 | 0.8969±0.0117 | 0.8562±0.0145 | 0.3333±0.0301 | 0.9826±0.0098 | 0.7022±0.0266 |
| 10 | 120.0±0.8944 | 133 | 0.9023±0.0067 | 0.8498±0.0095 | 0.3500±0.0306 | 0.9778±0.0087 | 0.7044±0.0151 |
| 11 | 101.8±0.4000 | 117 | 0.8701±0.0034 | 0.7898±0.0121 | 0.1733±0.0327 | 0.9725±0.0039 | 0.5912±0.0203 |
| 12 | 85.6±0.8000 | 99 | 0.8646±0.0081 | 0.8016±0.0119 | 0.1077±0.0615 | 0.9791±0.0087 | 0.5464±0.0482 |
| 13 | 77.4±0.8000 | 91 | 0.8505±0.0088 | 0.7692±0.0055 | 0.0923±0.0576 | 0.9769±0.0126 | 0.5303±0.0441 |
| 14 | 64.8±0.4000 | 75 | 0.8640±0.0053 | 0.8241±0.0161 | 0.1500±0.0333 | 1.0000±0.0000 | 0.5922±0.0277 |
| 15 | 56.6±0.4899 | 67 | 0.8448±0.0073 | 0.7182±0.0221 | 0.1091±0.0364 | 0.9893±0.0087 | 0.5498±0.0262 |
| 16 | 48.8±0.4000 | 59 | 0.8271±0.0068 | 0.6739±0.0150 | 0.0000±0.0000 | 0.9959±0.0082 | 0.4527±0.0020 |
| 17 | 45.0±0.0000 | 54 | 0.8333±0.0000 | 0.6840±0.0245 | 0.0000±0.0000 | 1.0000±0.0000 | 0.4545±0.0000 |
| 18 | 37.0±0.0000 | 45 | 0.8222±0.0000 | 0.7973±0.0200 | 0.0000±0.0000 | 1.0000±0.0000 | 0.4512±0.0000 |
| 19 | 32.0±0.0000 | 38 | 0.8421±0.0000 | 0.7375±0.0392 | 0.0000±0.0000 | 1.0000±0.0000 | 0.4571±0.0000 |
| 20 | 28.8±0.7483 | 35 | 0.8229±0.0214 | 0.7483±0.0516 | 0.0333±0.0667 | 0.9862±0.0169 | 0.4797±0.0619 |

Table 108: H12O SVC with missing flags, results of CV, taking best model of grid using accuracy. Predictions from outcome.

| Day | Corrects | Totals | Accuracy | AUC | Sensitivity | Specificity | F1 |
|---|---|---|---|---|---|---|---|
| 1 | 325.2±2.9257 | 374 | 0.8695±0.0078 | 0.8797±0.0070 | 0.5520±0.0371 | 0.9185±0.0060 | 0.7274±0.0170 |
| 2 | 326.6±4.3635 | 374 | 0.8733±0.0117 | 0.8911±0.0072 | 0.6480±0.0431 | 0.9080±0.0109 | 0.7516±0.0209 |
| 3 | 332.0±1.2649 | 374 | 0.8877±0.0034 | 0.8607±0.0049 | 0.6440±0.0265 | 0.9253±0.0036 | 0.7698±0.0085 |
| 4 | 326.6±1.0198 | 364 | 0.8973±0.0028 | 0.9016±0.0061 | 0.7234±0.0381 | 0.9230±0.0047 | 0.7923±0.0086 |
| 5 | 303.6±1.3565 | 332 | 0.9145±0.0041 | 0.9457±0.0044 | 0.7795±0.0261 | 0.9324±0.0073 | 0.8161±0.0051 |
| 6 | 272.0±1.2649 | 291 | 0.9347±0.0043 | 0.9431±0.0061 | 0.7882±0.0471 | 0.9541±0.0038 | 0.8502±0.0128 |
| 7 | 224.8±0.7483 | 250 | 0.8992±0.0030 | 0.9262±0.0109 | 0.7655±0.0552 | 0.9167±0.0093 | 0.7894±0.0072 |
| 8 | 176.4±2.3324 | 201 | 0.8776±0.0116 | 0.9137±0.0089 | 0.7583±0.0553 | 0.8938±0.0097 | 0.7622±0.0218 |
| 9 | 134.8±2.2271 | 159 | 0.8478±0.0140 | 0.8794±0.0092 | 0.6190±0.0426 | 0.8826±0.0116 | 0.7140±0.0237 |
| 10 | 113.8±0.7483 | 133 | 0.8556±0.0056 | 0.8636±0.0158 | 0.6500±0.0306 | 0.8838±0.0068 | 0.7175±0.0097 |
| 11 | 96.2±1.7205 | 117 | 0.8222±0.0147 | 0.8518±0.0114 | 0.5467±0.0499 | 0.8627±0.0196 | 0.6675±0.0162 |
| 12 | 89.2±0.9798 | 99 | 0.9010±0.0099 | 0.8934±0.0285 | 0.6923±0.0487 | 0.9326±0.0087 | 0.7948±0.0204 |
| 13 | 77.0±1.2649 | 91 | 0.8462±0.0139 | 0.9138±0.0196 | 0.7692±0.0487 | 0.8590±0.0140 | 0.7469±0.0204 |
| 14 | 68.4±1.9596 | 75 | 0.9120±0.0261 | 0.9772±0.0118 | 0.9667±0.0408 | 0.9016±0.0273 | 0.8629±0.0358 |
| 15 | 60.4±0.8000 | 67 | 0.9015±0.0119 | 0.9766±0.0090 | 0.9091±0.0000 | 0.9000±0.0143 | 0.8455±0.0150 |
| 16 | 51.6±2.2450 | 59 | 0.8746±0.0381 | 0.9751±0.0115 | 0.8800±0.0748 | 0.8735±0.0327 | 0.8134±0.0520 |
| 17 | 49.4±1.3565 | 54 | 0.9148±0.0251 | 0.9630±0.0167 | 0.8667±0.0444 | 0.9244±0.0227 | 0.8608±0.0376 |
| 18 | 42.6±0.4899 | 45 | 0.9467±0.0109 | 0.9959±0.0025 | 1.0000±0.0000 | 0.9351±0.0132 | 0.9183±0.0150 |
| 19 | 35.8±0.9798 | 38 | 0.9421±0.0258 | 0.9906±0.0069 | 1.0000±0.0000 | 0.9313±0.0306 | 0.9067±0.0383 |
| 20 | 33.6±0.4899 | 35 | 0.9600±0.0140 | 0.9943±0.0051 | 1.0000±0.0000 | 0.9517±0.0169 | 0.9359±0.0206 |

Table 109: H12O SVC with missing flags, results of CV, taking best model of grid using F1. Predictions from admission.

| Day | Corrects | Totals | Accuracy | AUC | Sensitivity | Specificity | F1 |
|---|---|---|---|---|---|---|---|
| 1 | 365.4±1.3565 | 374 | 0.9770±0.0036 | 0.9893±0.0055 | 0.9280±0.0204 | 0.9846±0.0052 | 0.9510±0.0072 |
| 2 | 362.8±0.7483 | 374 | 0.9701±0.0020 | 0.9920±0.0023 | 0.8960±0.0320 | 0.9815±0.0044 | 0.9357±0.0050 |
| 3 | 359.8±1.3266 | 374 | 0.9620±0.0035 | 0.9722±0.0040 | 0.8760±0.0150 | 0.9753±0.0039 | 0.9193±0.0071 |
| 4 | 347.0±2.1909 | 364 | 0.9533±0.0060 | 0.9719±0.0017 | 0.8936±0.0301 | 0.9621±0.0066 | 0.9023±0.0118 |
| 5 | 307.4±4.8415 | 332 | 0.9259±0.0146 | 0.9551±0.0058 | 0.7282±0.0417 | 0.9522±0.0110 | 0.8283±0.0310 |
| 6 | 264.0±1.6733 | 291 | 0.9072±0.0058 | 0.9393±0.0102 | 0.7588±0.0288 | 0.9268±0.0029 | 0.8014±0.0129 |
| 7 | 222.6±3.3226 | 250 | 0.8904±0.0133 | 0.9175±0.0072 | 0.6621±0.0402 | 0.9204±0.0106 | 0.7606±0.0261 |
| 8 | 172.4±0.8000 | 201 | 0.8577±0.0040 | 0.8794±0.0141 | 0.6333±0.0612 | 0.8881±0.0083 | 0.7154±0.0131 |
| 9 | 132.2±2.7857 | 159 | 0.8314±0.0175 | 0.8216±0.0206 | 0.6190±0.0426 | 0.8638±0.0180 | 0.6961±0.0241 |
| 10 | 108.8±1.1662 | 133 | 0.8180±0.0088 | 0.7728±0.0249 | 0.3875±0.0250 | 0.8769±0.0087 | 0.6167±0.0131 |
| 11 | 92.0±0.0000 | 117 | 0.7863±0.0000 | 0.6961±0.0431 | 0.4000±0.0000 | 0.8431±0.0000 | 0.5987±0.0000 |
| 12 | 73.8±1.8330 | 99 | 0.7455±0.0185 | 0.6773±0.0317 | 0.4000±0.0576 | 0.7977±0.0158 | 0.5686±0.0268 |
| 13 | 71.8±1.9391 | 91 | 0.7890±0.0213 | 0.7193±0.0108 | 0.4769±0.0576 | 0.8410±0.0174 | 0.6327±0.0322 |
| 14 | 58.0±2.2804 | 75 | 0.7733±0.0304 | 0.6910±0.0470 | 0.3500±0.0624 | 0.8540±0.0273 | 0.5980±0.0435 |
| 15 | 46.4±2.1541 | 67 | 0.6925±0.0322 | 0.5948±0.0310 | 0.2182±0.0727 | 0.7857±0.0319 | 0.4996±0.0406 |
| 16 | 42.8±1.1662 | 59 | 0.7254±0.0198 | 0.5294±0.0301 | 0.1000±0.0000 | 0.8531±0.0238 | 0.4739±0.0103 |
| 17 | 39.0±1.6733 | 54 | 0.7222±0.0310 | 0.5610±0.0424 | 0.1556±0.0544 | 0.8356±0.0301 | 0.4960±0.0375 |
| 18 | 33.4±1.6248 | 45 | 0.7422±0.0361 | 0.7149±0.0476 | 0.3000±0.1275 | 0.8378±0.0296 | 0.5655±0.0631 |
| 19 | 26.2±1.1662 | 38 | 0.6895±0.0307 | 0.5323±0.0559 | 0.2000±0.0667 | 0.7812±0.0342 | 0.4887±0.0318 |
| 20 | 28.4±1.0198 | 35 | 0.8114±0.0291 | 0.7517±0.0750 | 0.3333±0.1054 | 0.9103±0.0276 | 0.6309±0.0590 |

Table 110: H12O SVC with missing flags, results of CV, taking best model of grid using F1. Predictions from outcome.

| Day | Corrects | Totals | Accuracy | AUC | Sensitivity | Specificity | F1 |
|---|---|---|---|---|---|---|---|
| 1 | 330.0±1.0954 | 374 | 0.8824±0.0029 | 0.8421±0.0024 | 0.2920±0.0325 | 0.9735±0.0025 | 0.6664±0.0166 |
| 2 | 333.6±2.5768 | 374 | 0.8920±0.0069 | 0.9005±0.0022 | 0.4320±0.0325 | 0.9630±0.0034 | 0.7278±0.0190 |
| 3 | 338.6±0.8000 | 374 | 0.9053±0.0021 | 0.9157±0.0058 | 0.5240±0.0196 | 0.9642±0.0025 | 0.7715±0.0071 |
| 4 | 332.4±1.3565 | 364 | 0.9132±0.0037 | 0.9396±0.0013 | 0.5872±0.0104 | 0.9615±0.0037 | 0.7934±0.0075 |
| 5 | 311.8±0.7483 | 332 | 0.9392±0.0023 | 0.9584±0.0020 | 0.7231±0.0192 | 0.9679±0.0017 | 0.8509±0.0065 |
| 6 | 272.2±1.4697 | 291 | 0.9354±0.0051 | 0.9392±0.0038 | 0.6471±0.0186 | 0.9735±0.0038 | 0.8323±0.0124 |
| 7 | 234.8±1.3266 | 250 | 0.9392±0.0053 | 0.9502±0.0023 | 0.6966±0.0258 | 0.9710±0.0054 | 0.8462±0.0122 |
| 8 | 188.6±0.4899 | 201 | 0.9383±0.0024 | 0.9418±0.0046 | 0.7167±0.0167 | 0.9684±0.0028 | 0.8501±0.0058 |
| 9 | 144.8±0.7483 | 159 | 0.9107±0.0047 | 0.9239±0.0031 | 0.6476±0.0381 | 0.9507±0.0029 | 0.8026±0.0132 |
| 10 | 121.8±0.7483 | 133 | 0.9158±0.0056 | 0.9165±0.0032 | 0.6875±0.0000 | 0.9470±0.0064 | 0.8074±0.0092 |
| 11 | 108.0±1.0954 | 117 | 0.9231±0.0094 | 0.9461±0.0034 | 0.6933±0.0327 | 0.9569±0.0078 | 0.8271±0.0191 |
| 12 | 91.6±1.0198 | 99 | 0.9253±0.0103 | 0.9462±0.0027 | 0.8154±0.0615 | 0.9419±0.0074 | 0.8485±0.0231 |
| 13 | 83.8±0.7483 | 91 | 0.9209±0.0082 | 0.9357±0.0046 | 0.7538±0.0308 | 0.9487±0.0081 | 0.8425±0.0154 |
| 14 | 71.2±0.7483 | 75 | 0.9493±0.0100 | 0.9812±0.0053 | 0.8833±0.0408 | 0.9619±0.0078 | 0.9087±0.0181 |
| 15 | 63.6±0.8000 | 67 | 0.9493±0.0119 | 0.9802±0.0044 | 0.8545±0.0445 | 0.9679±0.0134 | 0.9084±0.0200 |
| 16 | 55.2±0.7483 | 59 | 0.9356±0.0127 | 0.9694±0.0034 | 0.8000±0.0000 | 0.9633±0.0153 | 0.8852±0.0191 |
| 17 | 50.4±0.4899 | 54 | 0.9333±0.0091 | 0.9783±0.0077 | 0.8889±0.0000 | 0.9422±0.0109 | 0.8880±0.0132 |
| 18 | 43.4±0.8000 | 45 | 0.9644±0.0178 | 0.9939±0.0025 | 0.9500±0.0612 | 0.9676±0.0108 | 0.9413±0.0294 |
| 19 | 37.0±0.0000 | 38 | 0.9737±0.0000 | 0.9896±0.0000 | 1.0000±0.0000 | 0.9688±0.0000 | 0.9536±0.0000 |
| 20 | 34.0±0.0000 | 35 | 0.9714±0.0000 | 0.9943±0.0000 | 1.0000±0.0000 | 0.9655±0.0000 | 0.9528±0.0000 |

Table 111: H12O SVC applying laboratory reference values, results of CV, taking best model of grid using accuracy. Predictions from admission.

| Day | Corrects | Totals | Accuracy | AUC | Sensitivity | Specificity | F1 |
|---|---|---|---|---|---|---|---|
| 1 | 364.4±0.4899 | 374 | 0.9743±0.0013 | 0.9907±0.0009 | 0.8880±0.0098 | 0.9877±0.0000 | 0.9438±0.0031 |
| 2 | 363.4±0.4899 | 374 | 0.9717±0.0013 | 0.9919±0.0006 | 0.9040±0.0150 | 0.9821±0.0012 | 0.9393±0.0032 |
| 3 | 361.4±0.4899 | 374 | 0.9663±0.0013 | 0.9904±0.0006 | 0.8600±0.0000 | 0.9827±0.0015 | 0.9264±0.0026 |
| 4 | 349.8±1.3266 | 364 | 0.9610±0.0036 | 0.9763±0.0020 | 0.8596±0.0104 | 0.9760±0.0032 | 0.9141±0.0076 |
| 5 | 313.6±0.8000 | 332 | 0.9446±0.0024 | 0.9667±0.0025 | 0.6923±0.0162 | 0.9782±0.0017 | 0.8573±0.0067 |
| 6 | 274.0±1.6733 | 291 | 0.9416±0.0058 | 0.9545±0.0023 | 0.6824±0.0390 | 0.9759±0.0029 | 0.8493±0.0166 |
| 7 | 233.0±1.0954 | 250 | 0.9320±0.0044 | 0.9342±0.0069 | 0.5793±0.0258 | 0.9783±0.0053 | 0.8131±0.0107 |
| 8 | 184.2±0.9798 | 201 | 0.9164±0.0049 | 0.8910±0.0105 | 0.4833±0.0333 | 0.9751±0.0085 | 0.7667±0.0091 |
| 9 | 140.2±1.1662 | 159 | 0.8818±0.0073 | 0.8355±0.0077 | 0.3714±0.0356 | 0.9594±0.0074 | 0.6934±0.0187 |
| 10 | 117.6±0.8000 | 133 | 0.8842±0.0060 | 0.8550±0.0055 | 0.2125±0.0306 | 0.9761±0.0064 | 0.6212±0.0189 |
| 11 | 100.8±1.1662 | 117 | 0.8615±0.0100 | 0.8082±0.0118 | 0.1867±0.0267 | 0.9608±0.0107 | 0.5903±0.0201 |
| 12 | 87.2±1.6000 | 99 | 0.8808±0.0162 | 0.8174±0.0114 | 0.1846±0.1043 | 0.9860±0.0047 | 0.6072±0.0750 |
| 13 | 78.8±0.7483 | 91 | 0.8659±0.0082 | 0.8002±0.0165 | 0.1385±0.0897 | 0.9872±0.0081 | 0.5703±0.0676 |
| 14 | 63.8±0.4000 | 75 | 0.8507±0.0053 | 0.7963±0.0137 | 0.1000±0.0333 | 0.9937±0.0078 | 0.5460±0.0236 |
| 15 | 54.8±0.9798 | 67 | 0.8179±0.0146 | 0.7474±0.0248 | 0.0364±0.0445 | 0.9714±0.0087 | 0.4804±0.0417 |
| 16 | 48.4±0.4899 | 59 | 0.8203±0.0083 | 0.6388±0.0124 | 0.0400±0.0490 | 0.9796±0.0129 | 0.4823±0.0394 |
| 17 | 44.8±0.4000 | 54 | 0.8296±0.0074 | 0.6894±0.0128 | 0.0000±0.0000 | 0.9956±0.0089 | 0.4534±0.0022 |
| 18 | 36.2±0.7483 | 45 | 0.8044±0.0166 | 0.7872±0.0315 | 0.0000±0.0000 | 0.9784±0.0202 | 0.4458±0.0051 |
| 19 | 31.6±0.4899 | 38 | 0.8316±0.0129 | 0.6573±0.0306 | 0.0000±0.0000 | 0.9875±0.0153 | 0.4540±0.0039 |
| 20 | 30.0±0.0000 | 35 | 0.8571±0.0000 | 0.7494±0.0340 | 0.1667±0.0000 | 1.0000±0.0000 | 0.6032±0.0000 |

Table 112: H12O SVC applying laboratory reference values, results of CV, taking best model of grid using accuracy. Predictions from outcome.

| Day | Corrects | Totals | Accuracy | AUC | Sensitivity | Specificity | F1 |
|---|---|---|---|---|---|---|---|
| 1 | 327.2±1.9391 | 374 | 0.8749±0.0052 | 0.8507±0.0039 | 0.4080±0.0299 | 0.9469±0.0036 | 0.6973±0.0150 |
| 2 | 331.0±3.0332 | 374 | 0.8850±0.0081 | 0.9043±0.0094 | 0.5640±0.0294 | 0.9346±0.0074 | 0.7506±0.0158 |
| 3 | 335.0±0.6325 | 374 | 0.8957±0.0017 | 0.8973±0.0036 | 0.5840±0.0196 | 0.9438±0.0023 | 0.7697±0.0059 |
| 4 | 330.4±1.3565 | 364 | 0.9077±0.0037 | 0.9253±0.0041 | 0.6809±0.0135 | 0.9413±0.0047 | 0.8013±0.0060 |
| 5 | 308.8±1.4697 | 332 | 0.9301±0.0044 | 0.9561±0.0045 | 0.7795±0.0476 | 0.9502±0.0063 | 0.8417±0.0115 |
| 6 | 271.0±2.6077 | 291 | 0.9313±0.0090 | 0.9330±0.0061 | 0.8118±0.0300 | 0.9471±0.0087 | 0.8475±0.0175 |
| 7 | 229.6±1.0198 | 250 | 0.9184±0.0041 | 0.9337±0.0104 | 0.7724±0.0169 | 0.9376±0.0034 | 0.8201±0.0084 |
| 8 | 182.2±0.9798 | 201 | 0.9065±0.0049 | 0.9433±0.0094 | 0.7417±0.0312 | 0.9288±0.0077 | 0.8001±0.0080 |
| 9 | 138.0±1.2649 | 159 | 0.8679±0.0080 | 0.8973±0.0092 | 0.6381±0.0233 | 0.9029±0.0074 | 0.7416±0.0132 |
| 10 | 114.8±1.6000 | 133 | 0.8632±0.0120 | 0.8734±0.0187 | 0.6750±0.0612 | 0.8889±0.0076 | 0.7309±0.0246 |
| 11 | 102.4±0.8000 | 117 | 0.8752±0.0068 | 0.8868±0.0069 | 0.6400±0.0327 | 0.9098±0.0073 | 0.7475±0.0126 |
| 12 | 87.4±0.4899 | 99 | 0.8828±0.0049 | 0.9111±0.0138 | 0.7385±0.0377 | 0.9047±0.0087 | 0.7769±0.0080 |
| 13 | 78.6±1.9596 | 91 | 0.8637±0.0215 | 0.9039±0.0082 | 0.7385±0.0615 | 0.8846±0.0199 | 0.7630±0.0342 |
| 14 | 66.2±0.9798 | 75 | 0.8827±0.0131 | 0.9569±0.0058 | 0.8500±0.0624 | 0.8889±0.0266 | 0.8131±0.0103 |
| 15 | 60.4±0.4899 | 67 | 0.9015±0.0073 | 0.9571±0.0064 | 0.9091±0.0000 | 0.9000±0.0087 | 0.8453±0.0094 |
| 16 | 52.8±0.4000 | 59 | 0.8949±0.0068 | 0.9641±0.0082 | 0.9000±0.0000 | 0.8939±0.0082 | 0.8389±0.0083 |
| 17 | 49.6±1.0198 | 54 | 0.9185±0.0189 | 0.9704±0.0077 | 0.9333±0.0544 | 0.9156±0.0166 | 0.8711±0.0284 |
| 18 | 41.8±0.7483 | 45 | 0.9289±0.0166 | 0.9912±0.0017 | 0.9000±0.0500 | 0.9351±0.0132 | 0.8870±0.0256 |
| 19 | 36.2±0.7483 | 38 | 0.9526±0.0197 | 0.9938±0.0021 | 1.0000±0.0000 | 0.9437±0.0234 | 0.9215±0.0295 |
| 20 | 34.0±0.0000 | 35 | 0.9714±0.0000 | 1.0000±0.0000 | 1.0000±0.0000 | 0.9655±0.0000 | 0.9528±0.0000 |

Table 113: H12O SVC applying laboratory reference values, results of CV, taking best model of grid using F1. Predictions from admission.

| Day | Corrects | Totals | Accuracy | AUC | Sensitivity | Specificity | F1 |
|---|---|---|---|---|---|---|---|
| 1 | 363.8±1.3266 | 374 | 0.9727±0.0035 | 0.9884±0.0037 | 0.8720±0.0098 | 0.9883±0.0036 | 0.9398±0.0074 |
| 2 | 364.0±1.2649 | 374 | 0.9733±0.0034 | 0.9924±0.0016 | 0.9200±0.0000 | 0.9815±0.0039 | 0.9433±0.0065 |
| 3 | 360.4±1.9596 | 374 | 0.9636±0.0052 | 0.9876±0.0016 | 0.8640±0.0233 | 0.9790±0.0036 | 0.9215±0.0114 |
| 4 | 348.2±1.3266 | 364 | 0.9566±0.0036 | 0.9763±0.0027 | 0.9064±0.0104 | 0.9640±0.0047 | 0.9093±0.0065 |
| 5 | 314.6±3.7202 | 332 | 0.9476±0.0112 | 0.9576±0.0035 | 0.7590±0.0528 | 0.9727±0.0057 | 0.8715±0.0281 |
| 6 | 269.6±1.3565 | 291 | 0.9265±0.0047 | 0.9546±0.0025 | 0.7471±0.0235 | 0.9502±0.0045 | 0.8308±0.0102 |
| 7 | 231.6±0.4899 | 250 | 0.9264±0.0020 | 0.9156±0.0033 | 0.6621±0.0138 | 0.9611±0.0036 | 0.8173±0.0028 |
| 8 | 179.0±2.2804 | 201 | 0.8905±0.0113 | 0.8780±0.0177 | 0.6083±0.0333 | 0.9288±0.0092 | 0.7541±0.0220 |
| 9 | 135.8±3.7630 | 159 | 0.8541±0.0237 | 0.8100±0.0143 | 0.5048±0.0381 | 0.9072±0.0273 | 0.6978±0.0305 |
| 10 | 110.6±1.0198 | 133 | 0.8316±0.0077 | 0.8234±0.0180 | 0.4250±0.0729 | 0.8872±0.0064 | 0.6393±0.0250 |
| 11 | 96.6±1.3565 | 117 | 0.8256±0.0116 | 0.7275±0.0329 | 0.4400±0.0533 | 0.8824±0.0062 | 0.6454±0.0253 |
| 12 | 78.0±1.7889 | 99 | 0.7879±0.0181 | 0.7776±0.0089 | 0.3692±0.1323 | 0.8512±0.0200 | 0.5910±0.0482 |
| 13 | 71.2±2.3152 | 91 | 0.7824±0.0254 | 0.7385±0.0214 | 0.4615±0.1088 | 0.8359±0.0150 | 0.6220±0.0483 |
| 14 | 58.2±2.2271 | 75 | 0.7760±0.0297 | 0.7627±0.0261 | 0.3667±0.0408 | 0.8540±0.0339 | 0.6052±0.0323 |
| 15 | 53.2±0.9798 | 67 | 0.7940±0.0146 | 0.6380±0.0153 | 0.3818±0.1060 | 0.8750±0.0113 | 0.6250±0.0424 |
| 16 | 44.8±0.7483 | 59 | 0.7593±0.0127 | 0.5412±0.0353 | 0.2000±0.0632 | 0.8735±0.0153 | 0.5374±0.0310 |
| 17 | 39.2±1.1662 | 54 | 0.7259±0.0216 | 0.5620±0.0242 | 0.0444±0.0544 | 0.8622±0.0166 | 0.4457±0.0377 |
| 18 | 33.6±2.2450 | 45 | 0.7467±0.0499 | 0.7196±0.0468 | 0.3000±0.1000 | 0.8432±0.0397 | 0.5730±0.0738 |
| 19 | 27.4±1.0198 | 38 | 0.7211±0.0268 | 0.5427±0.0506 | 0.2000±0.0667 | 0.8187±0.0364 | 0.5069±0.0240 |
| 20 | 27.6±0.8000 | 35 | 0.7886±0.0229 | 0.6379±0.0416 | 0.3333±0.1054 | 0.8828±0.0169 | 0.6099±0.0532 |

Table 114: H12O SVC applying laboratory reference values, results of CV, taking best model of grid using F1. Predictions from outcome.

##### 8.6.3 Ensemble

| Day | Corrects | Totals | Accuracy | AUC | Sensitivity | Specificity | F1 |
|---|---|---|---|---|---|---|---|
| 1 | 329.8±4.0249 | 374 | 0.8818±0.0108 | 0.7826±0.0089 | 0.5600±0.0510 | 0.9315±0.0179 | 0.7453±0.0131 |
| 2 | 331.0±1.5811 | 374 | 0.8850±0.0042 | 0.8280±0.0264 | 0.6040±0.0434 | 0.9284±0.0055 | 0.7585±0.0123 |
| 3 | 338.6±1.6733 | 374 | 0.9053±0.0045 | 0.8546±0.0201 | 0.6600±0.0400 | 0.9432±0.0104 | 0.7980±0.0056 |
| 4 | 337.8±1.4832 | 364 | 0.9280±0.0041 | 0.8932±0.0143 | 0.7532±0.0415 | 0.9539±0.0073 | 0.8441±0.0092 |
| 5 | 313.6±1.8166 | 332 | 0.9446±0.0055 | 0.9060±0.0196 | 0.7641±0.0215 | 0.9686±0.0074 | 0.8665±0.0103 |
| 6 | 273.4±2.5100 | 291 | 0.9395±0.0086 | 0.8912±0.0151 | 0.7529±0.0335 | 0.9642±0.0075 | 0.8551±0.0193 |
| 7 | 234.2±2.1679 | 250 | 0.9368±0.0087 | 0.9059±0.0083 | 0.7793±0.0523 | 0.9575±0.0082 | 0.8524±0.0196 |
| 8 | 185.6±1.5166 | 201 | 0.9234±0.0075 | 0.8819±0.0087 | 0.7667±0.0228 | 0.9446±0.0084 | 0.8306±0.0137 |
| 9 | 145.6±1.1402 | 159 | 0.9157±0.0072 | 0.8789±0.0293 | 0.7238±0.0522 | 0.9449±0.0083 | 0.8224±0.0161 |
| 10 | 121.6±2.0736 | 133 | 0.9143±0.0156 | 0.8850±0.0203 | 0.7125±0.0559 | 0.9419±0.0213 | 0.8093±0.0227 |
| 11 | 108.0±1.7321 | 117 | 0.9231±0.0148 | 0.8915±0.0165 | 0.7600±0.0365 | 0.9471±0.0203 | 0.8370±0.0207 |
| 12 | 91.6±2.3022 | 99 | 0.9253±0.0233 | 0.9212±0.0197 | 0.8462±0.0000 | 0.9372±0.0268 | 0.8539±0.0354 |
| 13 | 84.6±0.8944 | 91 | 0.9297±0.0098 | 0.9395±0.0034 | 0.9231±0.0000 | 0.9308±0.0115 | 0.8739±0.0143 |
| 14 | 70.8±1.3038 | 75 | 0.9440±0.0174 | 0.9845±0.0010 | 0.9667±0.0456 | 0.9397±0.0207 | 0.9067±0.0260 |
| 15 | 63.4±0.8944 | 67 | 0.9463±0.0133 | 0.9849±0.0019 | 0.9818±0.0407 | 0.9393±0.0098 | 0.9121±0.0217 |
| 16 | 55.6±0.8944 | 59 | 0.9424±0.0152 | 0.9867±0.0062 | 1.0000±0.0000 | 0.9306±0.0183 | 0.9098±0.0205 |
| 17 | 52.2±0.8367 | 54 | 0.9667±0.0155 | 0.9993±0.0011 | 1.0000±0.0000 | 0.9600±0.0186 | 0.9449±0.0239 |
| 18 | 43.2±0.8367 | 45 | 0.9600±0.0186 | 0.9966±0.0000 | 0.9750±0.0559 | 0.9568±0.0242 | 0.9364±0.0274 |
| 19 | 36.2±0.4472 | 38 | 0.9526±0.0118 | 0.9938±0.0023 | 0.9667±0.0745 | 0.9500±0.0171 | 0.9184±0.0203 |
| 20 | 33.0±0.0000 | 35 | 0.9429±0.0000 | 0.9943±0.0000 | 0.8667±0.0745 | 0.9586±0.0154 | 0.9017±0.0050 |

Table 115: H12O RNN-Ensemble-F1, using accuracy to choose models. Predictions from admission.

| Day | Corrects | Totals | Accuracy | AUC | Sensitivity | Specificity | F1 |
|---|---|---|---|---|---|---|---|
| 1 | 365.0±2.5495 | 374 | 0.9759±0.0068 | 0.9784±0.0066 | 0.9240±0.0089 | 0.9840±0.0091 | 0.9489±0.0129 |
| 2 | 368.4±1.8166 | 374 | 0.9850±0.0049 | 0.9876±0.0006 | 0.9680±0.0179 | 0.9877±0.0058 | 0.9684±0.0099 |
| 3 | 367.6±1.1402 | 374 | 0.9829±0.0030 | 0.9843±0.0042 | 0.9560±0.0261 | 0.9870±0.0040 | 0.9637±0.0066 |
| 4 | 353.2±1.0954 | 364 | 0.9703±0.0030 | 0.9728±0.0108 | 0.9106±0.0350 | 0.9792±0.0042 | 0.9354±0.0072 |
| 5 | 315.4±1.8166 | 332 | 0.9500±0.0055 | 0.9090±0.0107 | 0.7282±0.0532 | 0.9795±0.0024 | 0.8725±0.0172 |
| 6 | 275.0±1.5811 | 291 | 0.9450±0.0054 | 0.9141±0.0266 | 0.7412±0.0483 | 0.9720±0.0075 | 0.8639±0.0138 |
| 7 | 230.2±0.8367 | 250 | 0.9208±0.0033 | 0.8596±0.0246 | 0.6414±0.0523 | 0.9575±0.0052 | 0.8036±0.0138 |
| 8 | 181.8±0.8367 | 201 | 0.9045±0.0042 | 0.8113±0.0196 | 0.5833±0.0659 | 0.9480±0.0074 | 0.7688±0.0180 |
| 9 | 137.0±0.7071 | 159 | 0.8616±0.0044 | 0.7599±0.0438 | 0.5048±0.0722 | 0.9159±0.0110 | 0.7044±0.0188 |
| 10 | 118.8±0.4472 | 133 | 0.8932±0.0034 | 0.8118±0.0283 | 0.6000±0.0559 | 0.9333±0.0111 | 0.7565±0.0071 |
| 11 | 99.8±1.0954 | 117 | 0.8530±0.0094 | 0.7375±0.0284 | 0.4800±0.0298 | 0.9078±0.0149 | 0.6854±0.0051 |
| 12 | 83.8±1.3038 | 99 | 0.8465±0.0132 | 0.7465±0.0171 | 0.5538±0.0644 | 0.8907±0.0195 | 0.6978±0.0189 |
| 13 | 73.4±1.8166 | 91 | 0.8066±0.0200 | 0.7104±0.0188 | 0.5538±0.0344 | 0.8487±0.0278 | 0.6666±0.0155 |
| 14 | 58.6±2.0736 | 75 | 0.7813±0.0276 | 0.7116±0.0148 | 0.4667±0.1264 | 0.8413±0.0355 | 0.6341±0.0427 |
| 15 | 50.4±1.3416 | 67 | 0.7522±0.0200 | 0.7195±0.0299 | 0.5455±0.0000 | 0.7929±0.0240 | 0.6313±0.0168 |
| 16 | 45.6±2.3022 | 59 | 0.7729±0.0390 | 0.6908±0.0083 | 0.5000±0.0000 | 0.8286±0.0470 | 0.6443±0.0354 |
| 17 | 41.4±1.3416 | 54 | 0.7667±0.0248 | 0.7279±0.0054 | 0.6222±0.0609 | 0.7956±0.0365 | 0.6604±0.0201 |
| 18 | 34.2±1.9235 | 45 | 0.7600±0.0427 | 0.7473±0.0102 | 0.7500±0.0000 | 0.7622±0.0520 | 0.6838±0.0379 |
| 19 | 24.8±1.0954 | 38 | 0.6526±0.0288 | 0.5948±0.0412 | 0.2333±0.0913 | 0.7312±0.0356 | 0.4763±0.0333 |
| 20 | 25.4±1.5166 | 35 | 0.7257±0.0433 | 0.4948±0.0317 | 0.2333±0.0913 | 0.8276±0.0597 | 0.5276±0.0399 |

Table 116: H12O RNN-Ensemble-F1, using accuracy to choose models. Predictions from outcome.

| Day | Corrects | Totals | Accuracy | AUC | Sensitivity | Specificity | F1 |
|---|---|---|---|---|---|---|---|
| 1 | 329.6±3.1305 | 374 | 0.8813±0.0084 | 0.7685±0.0083 | 0.5360±0.0261 | 0.9346±0.0128 | 0.7394±0.0084 |
| 2 | 331.4±1.9494 | 374 | 0.8861±0.0052 | 0.8267±0.0281 | 0.6040±0.0555 | 0.9296±0.0116 | 0.7598±0.0111 |
| 3 | 337.8±2.9496 | 374 | 0.9032±0.0079 | 0.8561±0.0135 | 0.6400±0.0374 | 0.9438±0.0126 | 0.7915±0.0110 |
| 4 | 337.6±3.6469 | 364 | 0.9275±0.0100 | 0.8856±0.0140 | 0.7277±0.0409 | 0.9571±0.0133 | 0.8401±0.0174 |
| 5 | 313.6±1.6733 | 332 | 0.9446±0.0050 | 0.9080±0.0185 | 0.7590±0.0389 | 0.9693±0.0087 | 0.8658±0.0096 |
| 6 | 274.2±0.4472 | 291 | 0.9423±0.0015 | 0.8876±0.0319 | 0.7529±0.0492 | 0.9673±0.0065 | 0.8599±0.0070 |
| 7 | 234.2±1.6432 | 250 | 0.9368±0.0066 | 0.9024±0.0238 | 0.7586±0.0545 | 0.9602±0.0038 | 0.8496±0.0184 |
| 8 | 186.8±1.7889 | 201 | 0.9294±0.0089 | 0.8812±0.0077 | 0.8000±0.0186 | 0.9469±0.0124 | 0.8451±0.0129 |
| 9 | 146.4±1.1402 | 159 | 0.9208±0.0072 | 0.8827±0.0249 | 0.7429±0.0722 | 0.9478±0.0119 | 0.8328±0.0170 |
| 10 | 121.6±1.5166 | 133 | 0.9143±0.0114 | 0.8800±0.0268 | 0.7125±0.0342 | 0.9419±0.0164 | 0.8092±0.0146 |
| 11 | 108.4±0.8944 | 117 | 0.9265±0.0076 | 0.8919±0.0173 | 0.8133±0.0298 | 0.9431±0.0128 | 0.8485±0.0093 |
| 12 | 91.0±1.2247 | 99 | 0.9192±0.0124 | 0.9102±0.0143 | 0.8308±0.0344 | 0.9326±0.0152 | 0.8415±0.0189 |
| 13 | 83.6±1.3416 | 91 | 0.9187±0.0147 | 0.9285±0.0194 | 0.8923±0.0421 | 0.9231±0.0157 | 0.8549±0.0233 |
| 14 | 70.2±1.3038 | 75 | 0.9360±0.0174 | 0.9774±0.0170 | 0.9333±0.0697 | 0.9365±0.0224 | 0.8925±0.0266 |
| 15 | 62.8±0.8367 | 67 | 0.9373±0.0125 | 0.9765±0.0204 | 0.9455±0.0498 | 0.9357±0.0160 | 0.8969±0.0189 |
| 16 | 55.2±0.8367 | 59 | 0.9356±0.0142 | 0.9863±0.0044 | 1.0000±0.0000 | 0.9224±0.0171 | 0.9004±0.0193 |
| 17 | 51.8±1.0954 | 54 | 0.9593±0.0203 | 0.9973±0.0020 | 1.0000±0.0000 | 0.9511±0.0243 | 0.9340±0.0298 |
| 18 | 42.4±0.8944 | 45 | 0.9422±0.0199 | 0.9943±0.0028 | 0.9500±0.0685 | 0.9405±0.0226 | 0.9092±0.0297 |
| 19 | 36.6±0.5477 | 38 | 0.9632±0.0144 | 0.9953±0.0022 | 0.9333±0.0913 | 0.9688±0.0221 | 0.9335±0.0249 |
| 20 | 33.4±0.5477 | 35 | 0.9543±0.0156 | 0.9776±0.0389 | 0.9333±0.0913 | 0.9586±0.0154 | 0.9230±0.0275 |

Table 117: H12O RNN-Ensemble-F1, using F1 to choose models. Predictions from admission.

| Day | Corrects | Totals | Accuracy | AUC | Sensitivity | Specificity | F1 |
|---|---|---|---|---|---|---|---|
| 1 | 366.0±3.0822 | 374 | 0.9786±0.0082 | 0.9762±0.0106 | 0.9320±0.0228 | 0.9858±0.0094 | 0.9545±0.0162 |
| 2 | 367.6±2.5100 | 374 | 0.9829±0.0067 | 0.9875±0.0008 | 0.9560±0.0261 | 0.9870±0.0088 | 0.9639±0.0133 |
| 3 | 365.6±2.4083 | 374 | 0.9775±0.0064 | 0.9834±0.0048 | 0.9320±0.0228 | 0.9846±0.0062 | 0.9522±0.0132 |
| 4 | 352.8±1.6432 | 364 | 0.9692±0.0045 | 0.9693±0.0139 | 0.8936±0.0261 | 0.9804±0.0088 | 0.9325±0.0074 |
| 5 | 316.6±1.1402 | 332 | 0.9536±0.0034 | 0.9038±0.0128 | 0.7590±0.0344 | 0.9795±0.0042 | 0.8836±0.0095 |
| 6 | 275.2±2.3875 | 291 | 0.9457±0.0082 | 0.9108±0.0379 | 0.7353±0.0857 | 0.9735±0.0058 | 0.8637±0.0262 |
| 7 | 229.6±1.1402 | 250 | 0.9184±0.0046 | 0.8612±0.0319 | 0.6276±0.0617 | 0.9566±0.0052 | 0.7969±0.0174 |
| 8 | 181.8±1.6432 | 201 | 0.9045±0.0082 | 0.7988±0.0413 | 0.5917±0.0456 | 0.9469±0.0103 | 0.7711±0.0169 |
| 9 | 136.6±0.8944 | 159 | 0.8591±0.0056 | 0.7525±0.0383 | 0.4952±0.1043 | 0.9145±0.0119 | 0.6981±0.0289 |
| 10 | 118.6±0.8944 | 133 | 0.8917±0.0067 | 0.8163±0.0550 | 0.6125±0.1118 | 0.9299±0.0111 | 0.7554±0.0298 |
| 11 | 98.4±1.8166 | 117 | 0.8410±0.0155 | 0.7295±0.0401 | 0.4533±0.0558 | 0.8980±0.0226 | 0.6649±0.0178 |
| 12 | 83.4±1.5166 | 99 | 0.8424±0.0153 | 0.7566±0.0222 | 0.5846±0.0688 | 0.8814±0.0208 | 0.6999±0.0210 |
| 13 | 73.0±2.9155 | 91 | 0.8022±0.0320 | 0.7000±0.0147 | 0.5385±0.0000 | 0.8462±0.0374 | 0.6599±0.0294 |
| 14 | 58.6±2.3022 | 75 | 0.7813±0.0307 | 0.6939±0.0206 | 0.4167±0.0589 | 0.8508±0.0429 | 0.6231±0.0253 |
| 15 | 51.0±1.4142 | 67 | 0.7612±0.0211 | 0.7011±0.0253 | 0.5636±0.0407 | 0.8000±0.0319 | 0.6426±0.0136 |
| 16 | 46.6±1.6733 | 59 | 0.7898±0.0284 | 0.6980±0.0154 | 0.5200±0.0447 | 0.8449±0.0398 | 0.6634±0.0235 |
| 17 | 41.0±1.8708 | 54 | 0.7593±0.0346 | 0.7128±0.0243 | 0.4889±0.0609 | 0.8133±0.0404 | 0.6270±0.0376 |
| 18 | 34.4±1.9494 | 45 | 0.7644±0.0433 | 0.7503±0.0203 | 0.6750±0.1118 | 0.7838±0.0573 | 0.6747±0.0420 |
| 19 | 26.2±1.0954 | 38 | 0.6895±0.0288 | 0.5630±0.0603 | 0.2667±0.1491 | 0.7688±0.0523 | 0.5053±0.0369 |
| 20 | 25.6±1.3416 | 35 | 0.7314±0.0383 | 0.5161±0.0465 | 0.2000±0.0745 | 0.8414±0.0393 | 0.5211±0.0470 |

Table 118: H12O RNN-Ensemble-F1, using F1 to choose models. Predictions from outcome.

| Day | Corrects | Totals | Accuracy | AUC | Sensitivity | Specificity | F1 |
|---|---|---|---|---|---|---|---|
| 1 | 322.2±7.3621 | 374 | 0.8615±0.0197 | 0.7683±0.0203 | 0.6080±0.0502 | 0.9006±0.0276 | 0.7297±0.0212 |
| 2 | 328.4±3.9115 | 374 | 0.8781±0.0105 | 0.8175±0.0241 | 0.7000±0.0529 | 0.9056±0.0146 | 0.7666±0.0165 |
| 3 | 336.0±4.7958 | 374 | 0.8984±0.0128 | 0.8355±0.0220 | 0.7200±0.0490 | 0.9259±0.0181 | 0.7977±0.0180 |
| 4 | 336.0±3.8079 | 364 | 0.9231±0.0105 | 0.8908±0.0162 | 0.8170±0.0323 | 0.9388±0.0108 | 0.8441±0.0185 |
| 5 | 309.6±2.7928 | 332 | 0.9325±0.0084 | 0.8878±0.0123 | 0.8000±0.0281 | 0.9502±0.0129 | 0.8489±0.0115 |
| 6 | 270.6±3.0496 | 291 | 0.9299±0.0105 | 0.8752±0.0227 | 0.7765±0.0446 | 0.9502±0.0093 | 0.8407±0.0220 |
| 7 | 231.2±2.7749 | 250 | 0.9248±0.0111 | 0.8929±0.0164 | 0.8207±0.0378 | 0.9385±0.0152 | 0.8372±0.0167 |
| 8 | 185.0±0.7071 | 201 | 0.9204±0.0035 | 0.8819±0.0080 | 0.8000±0.0186 | 0.9367±0.0062 | 0.8300±0.0041 |
| 9 | 144.8±1.0954 | 159 | 0.9107±0.0069 | 0.8528±0.0190 | 0.7429±0.0426 | 0.9362±0.0139 | 0.8176±0.0060 |
| 10 | 120.8±2.7749 | 133 | 0.9083±0.0209 | 0.8638±0.0147 | 0.7625±0.0280 | 0.9282±0.0223 | 0.8079±0.0318 |
| 11 | 105.8±1.3038 | 117 | 0.9043±0.0111 | 0.8748±0.0333 | 0.7867±0.0730 | 0.9216±0.0183 | 0.8109±0.0168 |
| 12 | 90.2±1.9235 | 99 | 0.9111±0.0194 | 0.9153±0.0176 | 0.8615±0.0344 | 0.9186±0.0201 | 0.8333±0.0293 |
| 13 | 83.6±1.1402 | 91 | 0.9187±0.0125 | 0.9403±0.0034 | 0.9231±0.0000 | 0.9179±0.0146 | 0.8580±0.0178 |
| 14 | 69.2±2.1679 | 75 | 0.9227±0.0289 | 0.9860±0.0019 | 1.0000±0.0000 | 0.9079±0.0344 | 0.8801±0.0379 |
| 15 | 62.4±1.3416 | 67 | 0.9313±0.0200 | 0.9860±0.0016 | 1.0000±0.0000 | 0.9179±0.0240 | 0.8929±0.0273 |
| 16 | 54.4±0.8944 | 59 | 0.9220±0.0152 | 0.9882±0.0051 | 1.0000±0.0000 | 0.9061±0.0183 | 0.8823±0.0195 |
| 17 | 51.0±1.5811 | 54 | 0.9444±0.0293 | 1.0000±0.0000 | 1.0000±0.0000 | 0.9333±0.0351 | 0.9132±0.0420 |
| 18 | 42.4±1.1402 | 45 | 0.9422±0.0253 | 0.9966±0.0012 | 1.0000±0.0000 | 0.9297±0.0308 | 0.9131±0.0352 |
| 19 | 36.0±0.7071 | 38 | 0.9474±0.0186 | 0.9776±0.0370 | 0.9667±0.0745 | 0.9437±0.0140 | 0.9103±0.0329 |
| 20 | 33.6±0.5477 | 35 | 0.9600±0.0156 | 0.9776±0.0357 | 0.9667±0.0745 | 0.9586±0.0154 | 0.9337±0.0264 |

Table 119: H12O RNN-Ensemble-SEN, using accuracy to choose models. Predictions from admission.

| Day | Corrects | Totals | Accuracy | AUC | Sensitivity | Specificity | F1 |
|---|---|---|---|---|---|---|---|
| 1 | 364.0±2.5495 | 374 | 0.9733±0.0068 | 0.9726±0.0108 | 0.9480±0.0228 | 0.9772±0.0099 | 0.9448±0.0123 |
| 2 | 366.8±2.2804 | 374 | 0.9807±0.0061 | 0.9839±0.0056 | 0.9720±0.0110 | 0.9821±0.0074 | 0.9601±0.0119 |
| 3 | 365.6±2.4083 | 374 | 0.9775±0.0064 | 0.9843±0.0043 | 0.9760±0.0089 | 0.9778±0.0080 | 0.9540±0.0122 |
| 4 | 352.0±1.5811 | 364 | 0.9670±0.0043 | 0.9650±0.0139 | 0.9404±0.0277 | 0.9710±0.0068 | 0.9307±0.0082 |
| 5 | 316.6±1.5166 | 332 | 0.9536±0.0046 | 0.8894±0.0228 | 0.7949±0.0480 | 0.9747±0.0075 | 0.8873±0.0109 |
| 6 | 274.6±2.8810 | 291 | 0.9436±0.0099 | 0.9038±0.0390 | 0.8294±0.0816 | 0.9588±0.0115 | 0.8709±0.0236 |
| 7 | 229.8±1.6432 | 250 | 0.9192±0.0066 | 0.8409±0.0065 | 0.7172±0.0154 | 0.9457±0.0078 | 0.8137±0.0108 |
| 8 | 179.8±2.6833 | 201 | 0.8945±0.0133 | 0.7944±0.0148 | 0.6333±0.0349 | 0.9299±0.0181 | 0.7648±0.0166 |
| 9 | 135.0±2.0000 | 159 | 0.8491±0.0126 | 0.7367±0.0367 | 0.5524±0.0797 | 0.8942±0.0110 | 0.7008±0.0299 |
| 10 | 116.0±3.0822 | 133 | 0.8722±0.0232 | 0.7956±0.0177 | 0.6625±0.0342 | 0.9009±0.0268 | 0.7413±0.0295 |
| 11 | 97.6±4.2778 | 117 | 0.8342±0.0366 | 0.7130±0.0192 | 0.5200±0.0558 | 0.8804±0.0487 | 0.6754±0.0278 |
| 12 | 80.0±3.8079 | 99 | 0.8081±0.0385 | 0.7368±0.0337 | 0.6000±0.0843 | 0.8395±0.0516 | 0.6679±0.0340 |
| 13 | 71.0±1.8708 | 91 | 0.7802±0.0206 | 0.7104±0.0096 | 0.5538±0.0344 | 0.8179±0.0292 | 0.6417±0.0127 |
| 14 | 55.4±2.4083 | 75 | 0.7387±0.0321 | 0.6771±0.0312 | 0.5333±0.0950 | 0.7778±0.0526 | 0.6132±0.0195 |
| 15 | 48.2±2.6833 | 67 | 0.7194±0.0400 | 0.7192±0.0175 | 0.6364±0.0643 | 0.7357±0.0584 | 0.6205±0.0258 |
| 16 | 43.0±1.0000 | 59 | 0.7288±0.0169 | 0.6839±0.0182 | 0.5400±0.0548 | 0.7673±0.0310 | 0.6134±0.0060 |
| 17 | 40.2±1.7889 | 54 | 0.7444±0.0331 | 0.7323±0.0085 | 0.6667±0.0000 | 0.7600±0.0398 | 0.6493±0.0294 |
| 18 | 32.6±2.3022 | 45 | 0.7244±0.0512 | 0.7500±0.0130 | 0.7500±0.0000 | 0.7189±0.0622 | 0.6525±0.0436 |
| 19 | 23.6±1.1402 | 38 | 0.6211±0.0300 | 0.5792±0.0500 | 0.5333±0.1394 | 0.6375±0.0280 | 0.5224±0.0414 |
| 20 | 21.4±1.1402 | 35 | 0.6114±0.0326 | 0.4908±0.0380 | 0.2667±0.0913 | 0.6828±0.0450 | 0.4660±0.0311 |

Table 120: H12O RNN-Ensemble-SEN, using accuracy to choose models. Predictions from outcome.

| Day | Corrects | Totals | Accuracy | AUC | Sensitivity | Specificity | F1 |
|---|---|---|---|---|---|---|---|
| 1 | 318.6±8.4735 | 374 | 0.8519±0.0227 | 0.7685±0.0083 | 0.6160±0.0297 | 0.8883±0.0305 | 0.7202±0.0199 |
| 2 | 328.6±1.9494 | 374 | 0.8786±0.0052 | 0.8267±0.0281 | 0.7200±0.0632 | 0.9031±0.0134 | 0.7703±0.0098 |
| 3 | 335.4±2.9665 | 374 | 0.8968±0.0079 | 0.8561±0.0135 | 0.7680±0.0335 | 0.9167±0.0133 | 0.8024±0.0087 |
| 4 | 330.8±2.7749 | 364 | 0.9088±0.0076 | 0.8856±0.0140 | 0.8128±0.0277 | 0.9230±0.0091 | 0.8218±0.0122 |
| 5 | 308.0±2.3452 | 332 | 0.9277±0.0071 | 0.9080±0.0185 | 0.8462±0.0405 | 0.9386±0.0128 | 0.8459±0.0081 |
| 6 | 267.4±3.4351 | 291 | 0.9189±0.0118 | 0.8876±0.0319 | 0.8059±0.0677 | 0.9339±0.0180 | 0.8262±0.0186 |
| 7 | 230.4±1.8166 | 250 | 0.9216±0.0073 | 0.9024±0.0238 | 0.8414±0.0523 | 0.9321±0.0147 | 0.8342±0.0067 |
| 8 | 182.4±1.8166 | 201 | 0.9075±0.0090 | 0.8812±0.0077 | 0.8000±0.0186 | 0.9220±0.0122 | 0.8102±0.0119 |
| 9 | 144.4±3.0496 | 159 | 0.9082±0.0192 | 0.8827±0.0249 | 0.8095±0.0583 | 0.9232±0.0283 | 0.8237±0.0243 |
| 10 | 118.8±2.5884 | 133 | 0.8932±0.0195 | 0.8800±0.0268 | 0.8125±0.0625 | 0.9043±0.0252 | 0.7926±0.0266 |
| 11 | 104.8±1.7889 | 117 | 0.8957±0.0153 | 0.8919±0.0173 | 0.8267±0.0365 | 0.9059±0.0203 | 0.8047±0.0191 |
| 12 | 89.2±2.1679 | 99 | 0.9010±0.0219 | 0.9102±0.0143 | 0.8615±0.0344 | 0.9070±0.0233 | 0.8192±0.0317 |
| 13 | 81.8±2.5884 | 91 | 0.8989±0.0284 | 0.9285±0.0194 | 0.9077±0.0344 | 0.8974±0.0339 | 0.8305±0.0373 |
| 14 | 67.6±2.0736 | 75 | 0.9013±0.0276 | 0.9774±0.0170 | 0.9833±0.0373 | 0.8857±0.0305 | 0.8506±0.0378 |
| 15 | 61.6±1.3416 | 67 | 0.9194±0.0200 | 0.9765±0.0204 | 0.9818±0.0407 | 0.9071±0.0233 | 0.8753±0.0275 |
| 16 | 54.0±0.7071 | 59 | 0.9153±0.0120 | 0.9863±0.0044 | 1.0000±0.0000 | 0.8980±0.0144 | 0.8734±0.0153 |
| 17 | 49.4±0.8944 | 54 | 0.9148±0.0166 | 0.9973±0.0020 | 1.0000±0.0000 | 0.8978±0.0199 | 0.8717±0.0209 |
| 18 | 41.6±0.5477 | 45 | 0.9244±0.0122 | 0.9943±0.0028 | 1.0000±0.0000 | 0.9081±0.0148 | 0.8885±0.0156 |
| 19 | 35.8±1.0954 | 38 | 0.9421±0.0288 | 0.9953±0.0022 | 1.0000±0.0000 | 0.9313±0.0342 | 0.9065±0.0404 |
| 20 | 32.2±0.8367 | 35 | 0.9200±0.0239 | 0.9776±0.0389 | 0.9667±0.0745 | 0.9103±0.0189 | 0.8777±0.0367 |

Table 121: H12O RNN-Ensemble-SEN, using F1 to choose models. Predictions from admission.

| Day | Corrects | Totals | Accuracy | AUC | Sensitivity | Specificity | F1 |
|---|---|---|---|---|---|---|---|
| 1 | 360.6±3.5071 | 374 | 0.9642±0.0094 | 0.9762±0.0106 | 0.9560±0.0219 | 0.9654±0.0122 | 0.9284±0.0162 |
| 2 | 364.6±4.3932 | 374 | 0.9749±0.0117 | 0.9875±0.0008 | 0.9800±0.0000 | 0.9741±0.0136 | 0.9495±0.0214 |
| 3 | 363.4±2.6077 | 374 | 0.9717±0.0070 | 0.9834±0.0048 | 0.9760±0.0089 | 0.9710±0.0077 | 0.9429±0.0131 |
| 4 | 350.0±1.8708 | 364 | 0.9615±0.0051 | 0.9693±0.0139 | 0.9532±0.0277 | 0.9628±0.0096 | 0.9214±0.0079 |
| 5 | 315.4±2.3022 | 332 | 0.9500±0.0069 | 0.9038±0.0128 | 0.8256±0.0281 | 0.9666±0.0106 | 0.8836±0.0117 |
| 6 | 272.4±3.9749 | 291 | 0.9361±0.0137 | 0.9108±0.0379 | 0.8471±0.0761 | 0.9479±0.0115 | 0.8594±0.0301 |
| 7 | 228.4±1.6733 | 250 | 0.9136±0.0067 | 0.8612±0.0319 | 0.7655±0.0663 | 0.9330±0.0087 | 0.8111±0.0171 |
| 8 | 177.4±1.8166 | 201 | 0.8826±0.0090 | 0.7988±0.0413 | 0.6500±0.0913 | 0.9141±0.0209 | 0.7499±0.0119 |
| 9 | 134.8±1.0954 | 159 | 0.8478±0.0069 | 0.7525±0.0383 | 0.5905±0.0865 | 0.8870±0.0150 | 0.7071±0.0201 |
| 10 | 114.8±2.1679 | 133 | 0.8632±0.0163 | 0.8163±0.0550 | 0.7125±0.1218 | 0.8838±0.0323 | 0.7366±0.0170 |
| 11 | 95.4±3.2094 | 117 | 0.8154±0.0274 | 0.7295±0.0401 | 0.5867±0.0869 | 0.8490±0.0315 | 0.6691±0.0350 |
| 12 | 78.0±4.0000 | 99 | 0.7879±0.0404 | 0.7566±0.0222 | 0.6615±0.0421 | 0.8070±0.0462 | 0.6609±0.0397 |
| 13 | 68.4±1.1402 | 91 | 0.7516±0.0125 | 0.7000±0.0147 | 0.5538±0.0344 | 0.7846±0.0167 | 0.6166±0.0113 |
| 14 | 54.2±2.4900 | 75 | 0.7227±0.0332 | 0.6939±0.0206 | 0.6000±0.0697 | 0.7460±0.0502 | 0.6137±0.0193 |
| 15 | 45.2±2.7749 | 67 | 0.6746±0.0414 | 0.7011±0.0253 | 0.6182±0.0761 | 0.6857±0.0530 | 0.5814±0.0331 |
| 16 | 42.4±1.9494 | 59 | 0.7186±0.0330 | 0.6980±0.0154 | 0.5800±0.0447 | 0.7469±0.0423 | 0.6135±0.0274 |
| 17 | 39.0±1.2247 | 54 | 0.7222±0.0227 | 0.7128±0.0243 | 0.6444±0.0497 | 0.7378±0.0290 | 0.6259±0.0209 |
| 18 | 33.0±2.0000 | 45 | 0.7333±0.0444 | 0.7503±0.0203 | 0.7500±0.0000 | 0.7297±0.0541 | 0.6600±0.0383 |
| 19 | 23.4±1.5166 | 38 | 0.6158±0.0399 | 0.5630±0.0603 | 0.5000±0.1667 | 0.6375±0.0474 | 0.5119±0.0487 |
| 20 | 22.0±2.3452 | 35 | 0.6286±0.0670 | 0.5161±0.0465 | 0.3333±0.1179 | 0.6897±0.0879 | 0.4925±0.0476 |

Table 122: H12O RNN-Ensemble-SEN, using F1 to choose models. Predictions from outcome.
